# A Wastewater-Based Epidemic Model for SARS-CoV-2 with Application to Three Canadian Cities

**DOI:** 10.1101/2021.07.19.21260773

**Authors:** Shokoofeh Nourbakhsh, Aamir Fazil, Michael Li, Chand S. Mangat, Shelley W. Peterson, Jade Daigle, Stacie Langner, Jayson Shurgold, Patrick D’Aoust, Robert Delatolla, Elizabeth Mercier, Xiaoli Pang, Bonita E. Lee, Rebecca Stuart, Shinthuja Wijayasri, David Champredon

## Abstract

The COVID-19 pandemic has stimulated wastewater-based surveillance, allowing public health to track the epidemic by monitoring the concentration of the genetic fingerprints of SARS-CoV-2 shed in wastewater by infected individuals. Wastewater-based surveillance for COVID-19 is still in its infancy. In particular, the quantitative link between clinical cases observed through traditional surveillance and the signals from viral concentrations in wastewater is still developing and hampers interpretation of the data and actionable public-health decisions.

We present a modelling framework that includes both SARS-CoV-2 transmission at the population level and the fate of SARS-CoV-2 RNA particles in the sewage system after faecal shedding by infected persons in the population.

Using our mechanistic representation of the combined clinical/wastewater system, we perform exploratory simulations to quantify the effect of surveillance effectiveness, public-health interventions and vaccination on the discordance between clinical and wastewater signals. We also apply our model to surveillance data from three Canadian cities to provide wastewater-informed estimates for the actual prevalence, the effective reproduction number and incidence forecasts. We find that wastewater-based surveillance, paired with this model, can complement clinical surveillance by supporting the estimation of key epidemiological metrics and hence better triangulate the state of an epidemic using this alternative data source.

## 2 Introduction

Wastewater has been used previously for monitoring of a wide range of behavioural, socio-economic and biological markers including: medical and illicit drugs [1, 2, 3]; antibiotic and antimicrobial resistance [4, 5, 6]; and industrial pollutant chemicals [7, 8]. Spatial and temporal screening of the wastewater collection system or “sewershed” can provide qualitative and quantitative information on the marker of interest within the population in a given sewer catchment contributing to the wastewater. The wastewater data when used as an index of disease burden can be incorporated into a clinical surveillance program that is purposeful, economical and action-oriented for public health [9]. Wastewater-based surveillance (WBS) has also proven to be a low-cost and non-invasive tool for the management of infectious disease pathogens such as norovirus [10, 11] and poliovirus [12, 13, 14] where viral concentration in wastewater served to supplement clinical surveillance. Since the start of the COVID-19 pandemic, SARS-CoV-2 RNA has been detected and quantified in sewage in many locations worldwide (as of June 2021, 55 countries have pilot programs for a wastewater surveillance system with 2,287 sampling sites [15]) and was employed successfully in correlating the concentration of SARS-CoV-2 in wastewater to clinical cases reported in the sewershed [16, 17, 18, 19, 20, 21, 22, 23, 24]. In some instances of targeted surveillance, the leading wastewater signal (measured as SARS-CoV-2 RNA concentration in wastewater) compared to the clinical reports provided an early sign for the introduction or resurgence of COVID-19 into a community [25, 26, 27, 28] enabling rapid deployment of public health response and mitigation efforts.

Despite numerous successes with wastewater-based surveillance during the pandemic, utilizing wastewater surveillance data as a public-health tool for quick response remains challenging for some jurisdictions, especially at the municipal level [29, 30]. A major hurdle is the lack of a quantitative framework to assess and interpret the wastewater data generated and to translate that into public health action [31, 32]. The common practice is to use the detection of SARS-CoV-2 in wastewater as a signal for COVID-19 (re)introduction in a community and/or perform trend analysis in parallel with clinical surveillance of COVID-19. At the time of this manuscript, it is generally not recommended to use SARS-CoV-2 WBS for direct inference of key epidemiological indicators such as prevalence of active infections [31, 32, 16, 33].

Public health response guided by SARS-CoV-2 levels in wastewater is currently hindered by a lack of structured interpretive criteria, which is at present obscured by the inherent complexity and variation imparted by diverse sewersheds and their contributing populations [34, 35, 36]. Sources of data variability includes individual’s shedding dynamics, sampling frequency of wastewater, non-standardized laboratory methods, sewershed-specific viral degradation and signal attenuation during its journey from the site of faecal shedding (and potentially from urinary or sputum deposit [37, 38]) to the sampling point. Attenuation of RNA signal in wastewater involves several factors, such as dilution in municipal wastewater constituents (e.g., storm water effects in combined sewers and infiltration effects in both combined and separated sewers), RNA degradation (*e*.*g*., due to household detergents and industrial wastewaters) and viral degeneration in the harsh wastewater environment due to temperature, bioactive chemicals, pH, etc. Solids sedimentation and resuspension may also play a key role in the transportation and decay of SARS-CoV-2 RNA because of the hydrophobic characteristics of the viral envelope and its strong associations to solids [39, 33]. In addition, concentration methods for detection enhancement and minimization of inhibitory substances of molecular tests can result in some loss of the viral target [40, 41].

Here, we present a modelling framework that attempts to link quantified SARS-CoV-2 levels in wastewater with estimates of infections in the population within the sewershed, and to support policy decisions. The model incorporates both the viral transmission within the population via a standard epidemiological SEIR-like model (“Susceptible - Exposed - Infectious - Recovered”) [42] and the fate of SARS-CoV-2 in wastewater using a simplified hydrological transport framework. To illustrate potential applications, we fit our model to WBS data and traditional clinical reports gathered from six wastewater treatment plants (WWTPs) located in three Canadian cities (Edmonton, Ottawa and Toronto) and provide wastewater-informed estimates of key epidemiological metrics. We also perform exploratory simulations to investigate how the wastewater signal can be mechanistically associated with clinical surveillance of COVID-19.

## 3 Methods

We develop a mathematical model that mechanistically describes both the transmission at the population level (“above ground”) and the concentration of SARS-CoV-2 in wastewater as a result of faecal shedding from the infected individuals (“below ground”).

### 3.1 Transmission between individuals

To model SARS-CoV-2 transmission in the population, we use a SEIR-type epidemiological model. The disease progression of individuals is captured through several compartments that reflect their epidemiological states and disease outcomes (Table 1). Individuals can be susceptible (*S*); exposed (infected but not yet infectious, *E*); symptomatically infected who will later become hospitalized (*J*) or recovered without hospitalization during active COVID-19 (*I*); asymptomatically infected (*A*); hospitalized (*H*); those recovered and no longer infectious but still shedding virus in faeces (*Z*); fully recovered and permanently immune but not shedding anymore (*R*) and deceased (*D*). We ignore any migration movements, so at any given time the total population is constant and equal to *N* = *S* + *E* + *J* + *I* + *A* + *H* + *Z* + *R* + *D*. Infection occurs at a time-dependent transmission rate *β*_*t*_ between infectious (states *I, J* or *A*) and susceptible individuals (*S*). Once infected, susceptible individuals enter the latent (non-infectious) state (*E*) for an average duration of 1*/ ϵ* days, where no faecal shedding occurs. A proportion *α* of all infections are asymptomatic. A fraction *h* of symptomatic individuals are hospitalized (*H*) for an average duration of 1*/ℓ* days and for those, the COVID-19-associatied mortality is *δ*. After their infectious period ends, patients enter the post-infection shedding state *Z* where SARS-CoV-2 faecal shedding still occurs for 1*/η* days on average. The exposed (*E*), infectious (*A, I* and *J*) and post-infection shedding (*Z*) states are modelled with a series of sub-compartments in order to have their respective sojourn time gamma-distributed [43, 44, 45]. Note that in our model, we make the simplifying assumption that hospitalized patients–assumed mostly bedridden and a small fraction of the shedding population–do not contribute significantly to faecal shedding.

**Table 1:** Description of the model’s compartments and parameters for the SARS-CoV-2 RNA transmission and disease outcome.

| Symbole | Definition |
| --- | --- |
| $S$ | susceptibles |
| $E$ | exposed susceptibles but not infectious |
| $A_k$ | asymptomatic infectious cases in $k^{th}$ subcompartment |
| $I_k$ | symptomatic infectious cases in $k^{th}$ subcompartment |
| $J_k$ | symptomatic infectious cases in $k^{th}$ subcompartment who later admits to hospital |
| $Z_k$ | non-infectious cases but fecal shedding SARS-CoV-2 RNA in $k^{th}$ subcompartment |
| $H$ | hospitalized patients |
| $R$ | recovered cases |
| $D$ | deceased cases |
| $\beta_{I,k}$ | transmission rate in $k^{th}$ subcompartment among symptomatics (per contact) |
| $\beta_{A,k}$ | transmission rate in $k^{th}$ subcompartment among asymptomatics (per contact) |
| $1/\varepsilon$ | ave. latency time (days) |
| $1/\nu_k$ | ave. duration in $k^{th}$ subcompartment among symptomatics (days) |
| $1/\mu_k$ | ave. duration in $k^{th}$ subcompartment among symptomatics goes to hospital (days) |
| $1/\theta_k$ | ave. duration in $k^{th}$ subcompartment among asymptomatics (days) |
| $1/\eta_k$ | ave. duration in $k^{th}$ subcompartment of shedding after infectiousness (days) |
| $1/\ell$ | ave. length of stay in a hospital (days) |
| $n_I$ | total number of subcompartments in $I$ state |
| $n_J$ | total number of subcompartments in $J$ state |
| $n_A$ | total number of subcompartments in $A$ state |
| $n_Z$ | total number of subcompartments in $Z$ state |
| $\alpha$ | proportion of exposed cases that are asymptomatic |
| $h$ | proportion of symptomatic cases that need hospital admission |
| $\delta$ | proportion of deceased individuals among hospitalized patients |

The transmission dynamics are represented by the system of differential equations 1a-1n and illustrated in Figure 1.

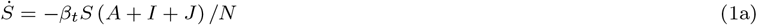

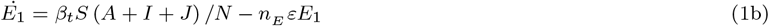

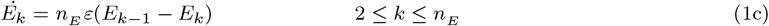

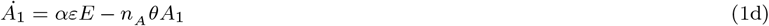

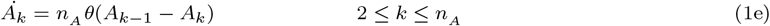

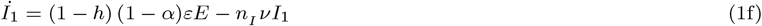

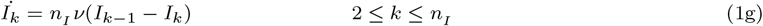

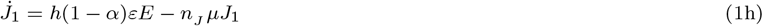

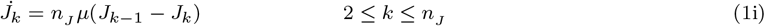

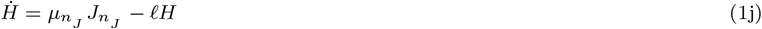

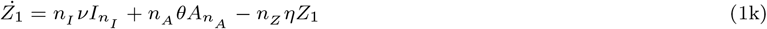

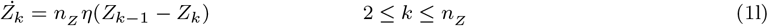

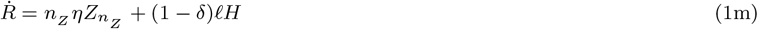

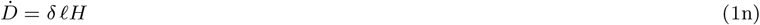

where 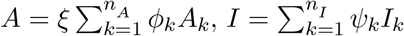 and 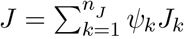. We use the dot notation to symbolize derivation with respect to time (*e*.*g*., 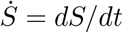).

**Figure 1:**
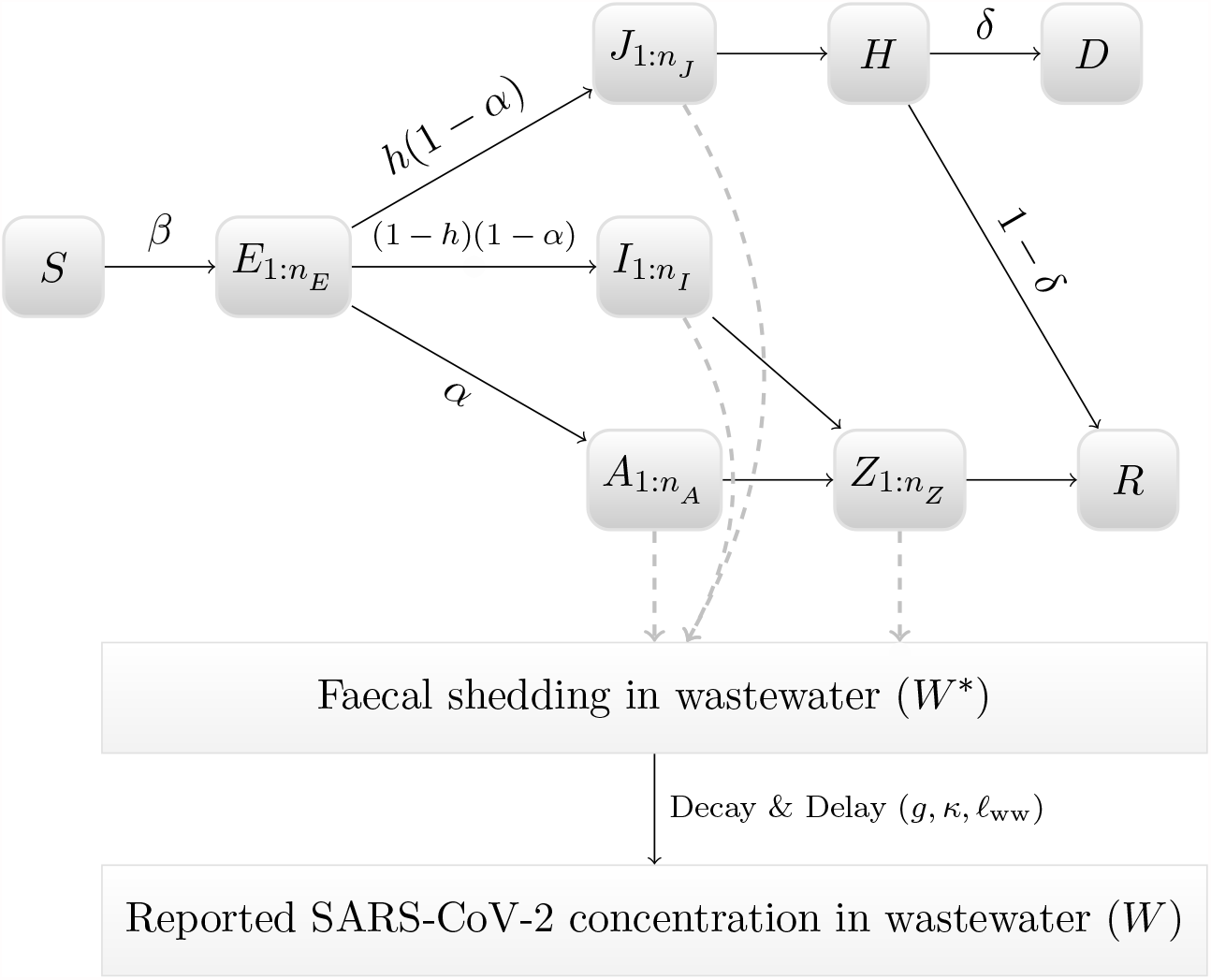
Diagram of compartmental model. See main text for a description of the epidemiological states. The notation 1 : n_•_ indicates a modelling using n_•_ sub-compartments to obtain a gamma-distributed sojourn time in the associated epidemiological state.

The parameters *ϕ* and *ψ* are multiplicative adjustments to the baseline transmission rate *β*_*t*_ to represent the infectious profile during the course of infection. The values for *ϕ*_*k*_ and *ψ*_*k*_ were chosen to represent the best estimate of the temporal infectiousness profile given the different results published (see Appendix, Figure S2). The parameter *ξ* models the relative infectiousness of asymptomatic cases compared to symptomatic ones. The effective reproduction number of this model is (see Appendix for details on its calculation):

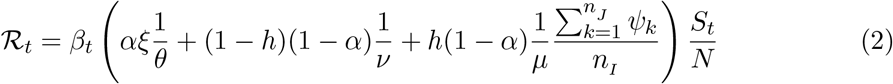

### 3.2 SARS-CoV-2 viral concentration in wastewater

#### 3.2.1 Deposited Viral Concentration

The daily concentration of SARS-CoV-2 in wastewater is directly calculated from the total number of individuals that are actively shedding into the sewage system. SARS-CoV-2 faecal shedding varies according to the infected individual’s clinical state and disease outcomes. Depending on the disease progression, infected individuals shed a variable amount of SARS-CoV-2 while they are in the shedding states (*A, I, J* and *Z*). The total concentration of SARS-CoV-2 RNA entering the wastewater at time *t* is given by

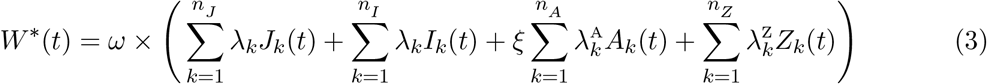

The parameters λ_*k*_, 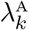 and 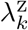 represent SARS-CoV-2 faecal shedding dynamics per capita when the infected individual is in the epidemiological states *I, J, A* and *Z* respectively. Given the current lack of observational data, we used the same parameters λ_*k*_ for all epidemiological states (*i*.*e*., 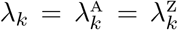). Values for the parameters λ_*k*_ were chosen to represent mid-range values of the different results published (see Appendix Figure S2). Note that we assume the same reduction in faecal shedding as in respiratory shedding for asymptomatic cases (parameter *ξ*). The parameter *ω* implies that our model can only determine up to a constant the concentration of SARS-CoV-2 in wastewater [14], even if the limit of detection of the assay is known. This reflects our current inability to quantify the various complex processes that affect the concentration, from patients’ shedding to the concentration measured in laboratories (*e*.*g*., frequency and timing of sampling, RNA degradation in the sewer system, recovery efficiency of assays).

#### 3.2.2 RNA transport and sampled viral concentration

We use a simple advection-dispersion-decay model to simulate the fate of SARS-CoV-2 along its journey in wastewater from the shedding points to the sampling site. This model is a combination of an exponential viral decay [46] and a *τ*-day dispersed plug-flow function, *g*(*τ*), representing all possible hydrodynamic processes (*e*.*g*., dilution, sedimentation and resuspension) that leads to RNA degradation as well as decrease and delay of signal at the time of sampling. The dispersed plug-flow *g*(*τ*) acts as a transformation function, which reshapes the initial deposited concentration, *W* ^∗^, into a delayed viral distribution over *τ* days as a result of the transit of SARS-CoV-2 in the sewer system. Hence, we defined the sampled viral concentration at time *t* as:

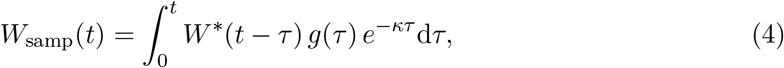

where *κ* is the daily decay rate of SARS-CoV-2 due to the harsh, complex and bioactive environment of wastewater [46]. The SARS-CoV-2 RNA concentration entering the sewage system daily is modelled as a single hydrodynamic pulse per day and the plug-flow function, *g*, is obtained by the analytical solution of the axial dispersed plug flow differential equation [47]. We then re-parametrize the analytical solution with the mean delay time 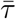 and its standard deviation *σ* into a Gaussian distribution

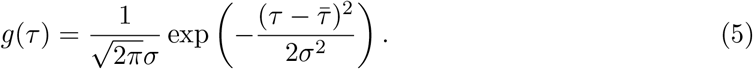

See the Appendix for more details on our advection-dispersion-decay model of the RNA transport.

#### 3.2.3 Wastewater reported sample

The sample transportation, laboratory processing time and reporting lags, introduce reporting delays of RNA concentration in wastewater. Hence, we define the reported wastewater concentration as

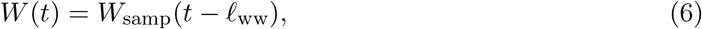

where *ℓ*_ww_ is the reporting lag between wastewater sampling and concentration report after laboratory analysis. (Note that the reporting delay of the wastewater measurement is independent from the delay caused by the transport of RNA particles in the sewer system as defined in Equation 4).

### 3.3 Clinical reported cases

We also model surveillance data derived from laboratory confirmed and clinically diagnosed COVID-19 cases, acknowledging that instantaneous identification and complete reporting after initial infection is not possible. We assume that a fraction *ρ* of symptomatic incidence is reported with a lag of a *ℓ*_clinical_ days from the time of infection. If *i*(*t*) is the total incidence at time *t*, we define the number of clinical cases reported at time *t* as:

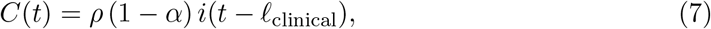

### 3.4 Wastewater and clinical surveillance data

We apply our modelling framework to data sets from six wastewater sampling sites located in three Canadian cities: Edmonton (Alberta), Ottawa (Ontario) and Toronto (Ontario). Sampling sites are the following municipal WWTPs (abbreviation / approximate population served): Gold Bar in Edmonton (EGB / 900,000); Robert O. Pickard Environmental Centre in Ottawa (OTW / 1,000,000 [48]); Toronto Ashbridges Bay (TAB / 1,603,700 [49]); Toronto Humber (THU / 685,000 [50]); Toronto Highland Creek (THC / 533,000 [51]); and Toronto North Toronto (TNT / 252,530 [52]).

#### 3.4.1 Data collection

Wastewater samples were collected approximately two (Edmonton and Toronto) to seven (Ottawa) times a week. The sampling location was at the influent of the wastewater treatment plant of each city. Wastewater samples were collected before de-gritting in Toronto, and after for Edmonton and Ottawa.

Wastewater samples from Edmonton and Toronto were shipped to the National Microbiology Laboratory in Winnipeg, Manitoba, where SARS-CoV-2 RNA concentration was measured. RNA from wastewater samples was purified using two methods. Prior to February 12th 2021, 15 mL of clarified supernatant (after 4000 × *g* centrifugation for 20 min at 4°C), was concentrated using an ultracentrifugal filter device (4000 × *g* for 35 min at 4°C) (Amicon Ultra-15, 10 kDa MWCO, Millipore-Sigma, St. Louis, MO, U.S.A). Total RNA was extracted from the resultant concentrate (∼200 *µ*L) using the MagNA Pure 96 DNA and Viral NA Large Volume Kit (Roche Diagnostics, Laval, QC) using the Plasma External Lysis 4.0 protocol as per manufacturer instructions. After February 12th 2021, the pellet resultant from clarifying (4000g for 20 minutes at 4°C) 30 mL of wastewater was resuspended in 700 *µ*L Qiagen Buffer RLT (Qiagen, Germantown, MD) containing 1% 2-mercaptoethanol. To this, 200 *µ*L of 0.5 mm zirconia-silica beads (Biospec, Bartlesville, OK) were added and the sample was processed with a Bead Mill 24 Homogenizer (Fisher Scientific, Ottawa, ON) using 4 × 30s pulses at 6 m/s, then clarified by centrifugation (12000 × *g*, 3 min) and the resultant lysate used for RNA extraction using the MagNA Pure 96 instrument as described above. Viral RNA was quantified using RTq-PCR with the US-CDC N1 and N2 primers.

For Ottawa, daily 24-hour composite primary sludge samples, consisting of four discrete samples collected at 6 hour intervals and subsequently mixed, were collected and transported on ice to the University of Ottawa, where samples were analyzed within 24 hours of reception. Samples were concentrated by centrifugation at 10,000g for 45 minutes and RNA was extracted from a 250 mg portion of the resulting pellet using a modified version of the Qiagen RNeasy PowerMicrobiome kit [28]. Quantification was performed using singleplex probe-based RTq-PCR for the N1 and N2 gene regions of the virus.

For Edmonton and Toronto, the SARS-CoV-2 concentrations in wastewater were normalized by the total solid suspension (TSS) measured on the day the sample was collected at the treatment plant. For Ottawa, they were normalized with the concentration of the Pepper mild mottle virus measured in the sample. For all cities, the reported viral concentration used in the model (*W*) was the average normalized concentration across all technical replicates for both the N1 and N2 genes.

We obtained clinical cases and hospital admissions (except for Toronto) for the catchment area of each of the six wastewater treatment plants. Hence, we were able to link clinical and wastewater surveillances. The data sets for the three cities are plotted in Figure 2 Seroprevalence values at the city and province level were obtained from Canadian Blood Services (CBS) [53]. The wastewater and clinical surveillance data used in this study are available in Supplementary File S1.

**Figure 2:**
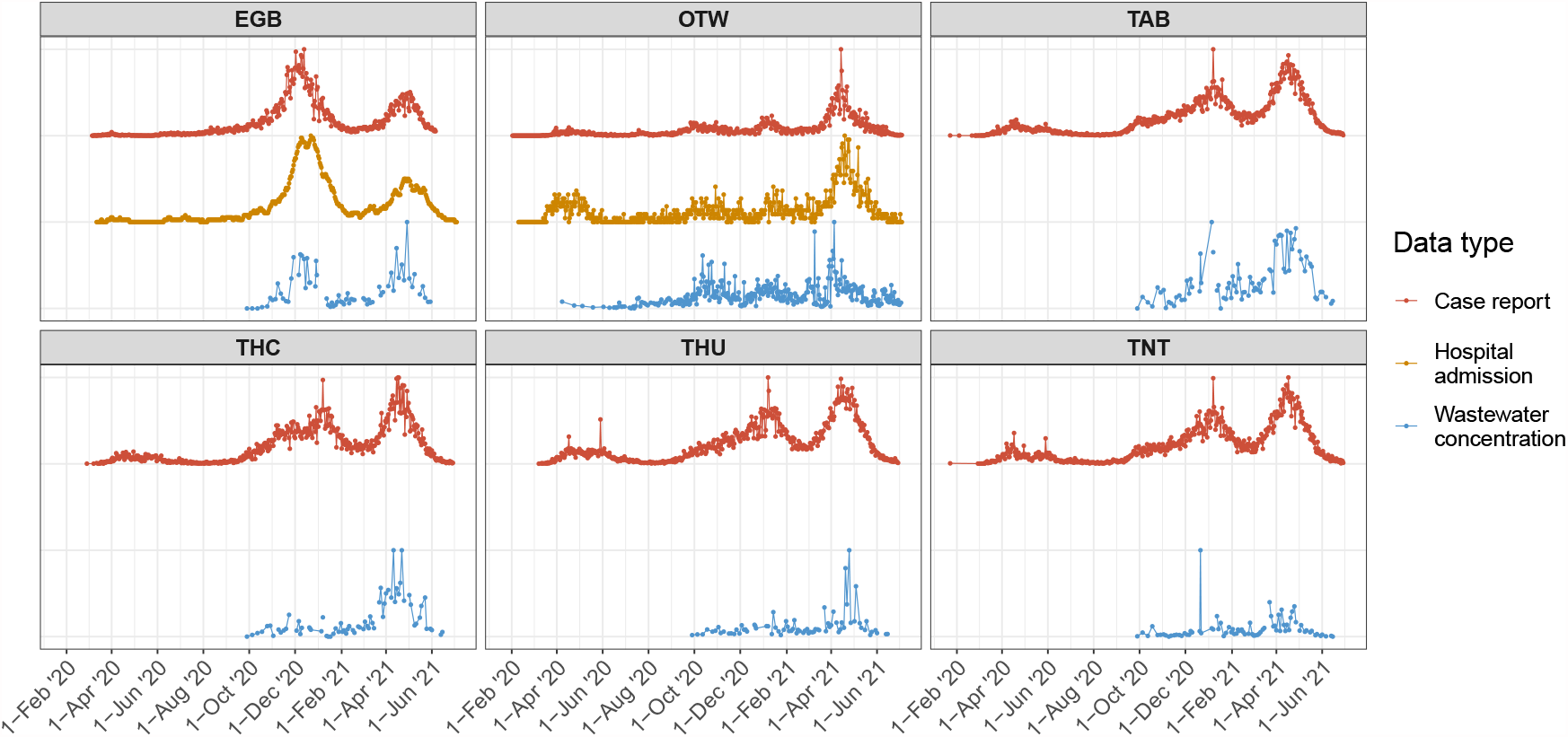
Data sets used in this study for Edmonton, Ottawa and Toronto. Each horizontal panel is a city and colors represent the type of data (reported cases, hospital admissions and SARS-CoV-2 RNA concentration in wastewater). All curves were normalized to 1 (dividing by their respective maximum value) to plot them in one single panel to facilitate visual comparison. All data sets used in this study are available in Supplementary File S1.

#### 3.4.2 Fit to data

We use an Approximate Bayesian Computation (ABC) algorithm [54] to fit the unknown or unobserved model parameters to the available data. For each ABC prior iteration, the error function is defined as a weighted trajectory matching

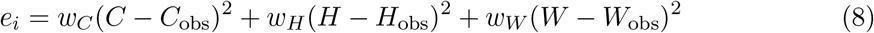

We use 50, 000 prior ABC iterations and retain the 100 smallest errors to generate posterior distributions (acceptance ratio 2 × 10^−3^). The parameters fitted are the time-dependent transmission rate *β*_*t*_, *ω*, the hospitalization rate *h* and the mean transit time 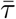. More details about the fitting procedure is given in Appendix.

We define three types of fitting-to-data procedures. “Clinical” when *w*_*C*_ = *w*_*H*_ = 1 and *w*_*W*_ = 0, to use data from clinical sources only ; “WW” when *w*_*W*_ = 1 and *w*_*H*_ = *w*_*C*_ = 0, to use wastewater data only; and finally “Combined” by choosing the weights *w*_*C*_, *w*_*H*_ and *w*_*W*_ such that the contribution of each error term (in Equation 8) are, on average, approximately equal. The “Combined” fitting procedure aims to have approximately the same contribution from clinical and wastewater data sources (despite the different observation frequencies).

#### 3.4.3 Inference of unobserved epidemiological quantities

For a given location, once the model is fitted data, we can infer unobserved quantities of epidemiological importance by generating epidemic trajectories from the posterior samples. The posterior prevalence distribution (at each time point) is defined by simply adding the populations from the compartments representing active infection, that is

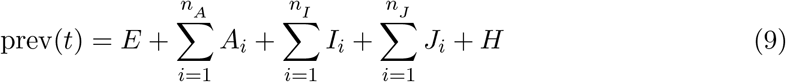

The posterior cumulative incidence is obtained by summing Equation 1a until time *t*

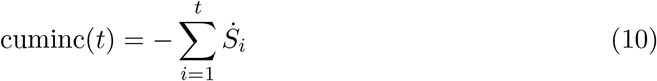

The fitted model can also provide an estimate of the effective reproduction number from the different data sources (e.g., clinical and/or wastewater) using Equation 2.

### 3.5 Simulations

#### 3.5.1 Detection timing differential

In order to explore wastewater-based surveillance as a leading indicator of infection in the community, the time when SARS-CoV-2 is first reported from wastewater is noted *d*_ww_ and defined as *W*(*t* = *d*_ww_) = LOD where LOD is the limit of detection of the laboratory method. In addition, the time *d*_clinical_ when COVID-19 is first reported is defined as *C*(*t* = *d*_clinical_) = 1. Finally, we define the reported detection differential Δ = *d*_ww_ − *d*_clinical_. As a result, the wastewater signal can be classified as a leading indicator over traditional clinical surveillance when Δ *<* 0. We assess how the reported detection differential Δ can be impacted by varying model parameters that would typically differ from one community-sewer system to another. We select only three combinations of parameters (among many) to illustrate how Δ can be affected, and most importantly how its sign can change indicating its transition between a leading and lagging indicator. We consider two levels of COVID-19 reporting, with *ρ* = 30% to reflect a relatively inefficient clinical surveillance system in the population, and *ρ* = 70% that represents a more efficient one.

#### 3.5.2 Impact of vaccination

Although the model presented here does not explicitly have a vaccination process, a mechanism using the existing framework can be implemented to mimic the main effects of an infection-permissive vaccine (as this is the case for the COVID-19 vaccines currently available). We model a simple scenario that rolls out an infection permissive vaccine by gradually decreasing the transmission rate (*β*) by 70% over 50 days and increasing the proportion of asymptomatic infection (*α*) from 30% to 90%. This reflects the growing protection of the population from severe outcomes of COVID-19 as well as decrease in transmissions as the vaccine is administered. To assess the differential impact of vaccination on clinical and wastewater observations, we consider the ratio of the level of SARS-CoV-2 in wastewater over the reported clinical cases, *W*(*t*)*/C*(*t*).

## 4 Results

We present our results in two sections. First, we apply our modelling framework to wastewater and clinical surveillance data from six sampling sites located in three Canadian cities (Edmonton, Ottawa and Toronto) and infer epidemiological parameters such as prevalence, effective reproduction number and incidence forecast. The second section is based on exploratory simulations (not fitted to data of a specific location) that highlight important mechanistic aspects between clinical and wastewater surveillances.

### 4.1 Application to Canadian surveillance data

In this section, we compare the inferences made on key epidemiological variables by fitting the model to different data sources. Our goal is to assess the added-value of the wastewater-based data stream. Hence, in the following, we present inferences by fitting the model to different data sources available (“Clinical”, “WW” and “Combined”) and comparing the outcomes.

#### 4.1.1 Prevalence estimates

Figure 3 shows, for selected locations, the SARS-CoV-2 prevalence estimated by sampling the posterior distributions fitted to the various data sources “Clinical”, “WW” and “Combined” and the evaluation of Equation 9. The right-most panel also displays estimates of cumulative incidence (Equation 10) compared to available SARS-CoV-2 seroprevalence levels estimated from surveys by the Canadian Blood Services performed on banks of blood donors in Edmonton, Ottawa and Toronto [53]. Note that our model was not fitted to seroprevalence data and this comparison acts as a crude check that prevalence estimates from the model are approximately consistent with other independent data sources. For all locations, as expected, wastewater-only prevalence estimates are close to the clinical-only ones when the levels of SARS-CoV-2 in wastewater mimic the COVID-19 trends in the population (Figure 2). For example, the prevalence estimated from wastewater-only and clinical-only are comparable for the December 2020 wave in Edmonton and April 2021 wave in Ottawa. However, when the clinical and wastewater signals are discordant, prevalence estimates can be significantly different. For example, wastewater-based prevalence estimates in January 2021 for Toronto Highland Creek (THC) do not show the peak seen from clinical observations. On the other hand, this January peak was captured in Toronto Ashbridges Bay (TAB)–another part of the city–and the subsequent March-May 2021 wave in Toronto Highland Creek (THC) was identified by both wastewater and clinical surveillance. Finally, we note that, because of the larger variability of SARS-CoV-2 WBS and/or their lower sampling frequency as compared to daily clinical surveillance, credible intervals of our wastewater-only inferences are generally larger than the clinical-only ones (Figure 3).

**Figure 3:**
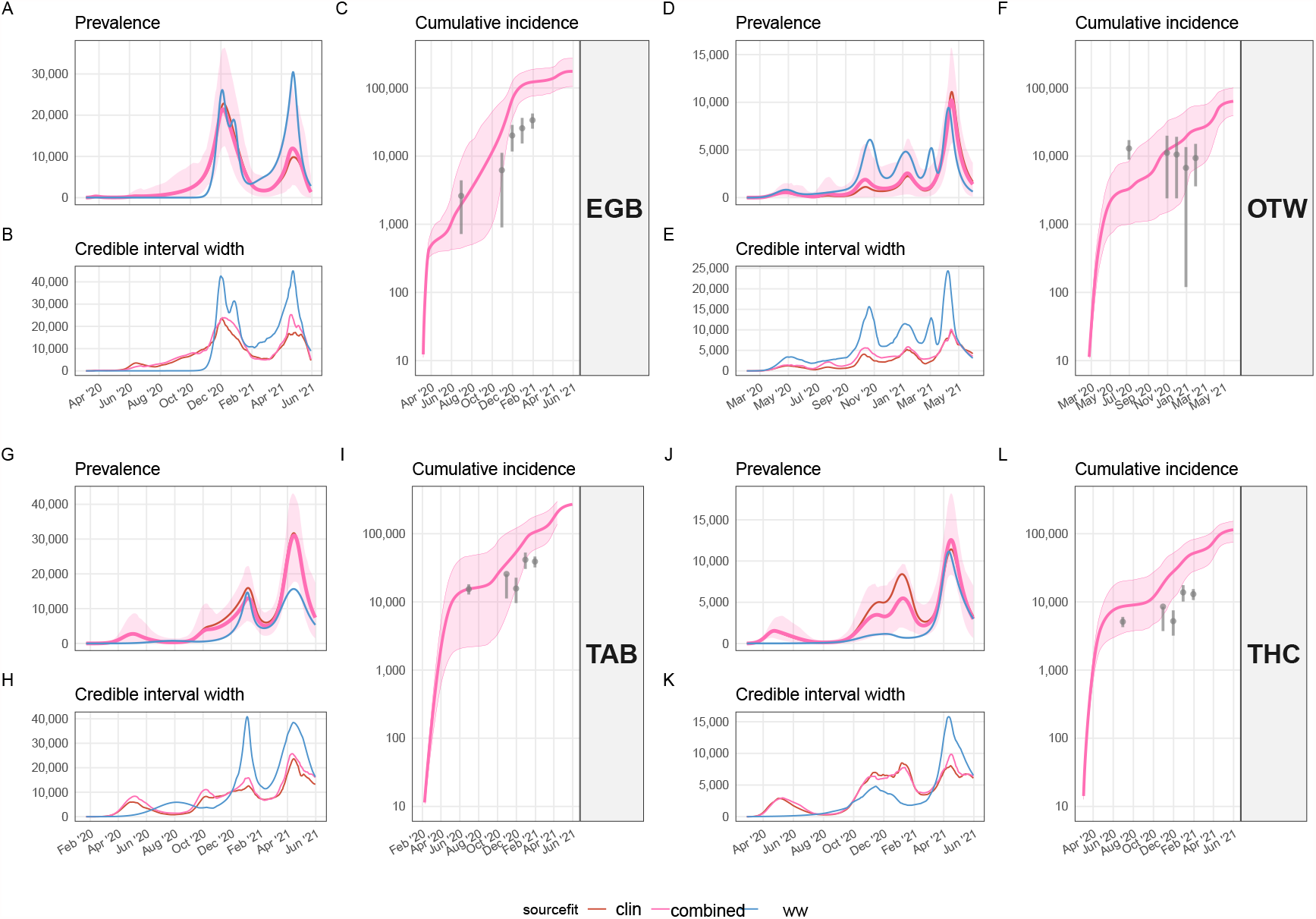
SARS-CoV-2 prevalence estimates. Each quadrant block represents a location. The top-left panel of each quadrant block (panels A,D,G,J) shows the estimates of SARS-CoV-2 prevalence time series. Each colour represents the different data sources used to fit the model (dark red: “Clinical”, pink: “Combined”, blue: “WW”). The lines show the mean estimate of prevalence. The shaded ribbon indicates the 95% CrI for the estimate fitted on the “Combined” data set (CrIs for other data sources are omitted for clarity). The bottom-left panel of each quadrant block (panels B,E,H,K) represents the width of the 95% CrI for the estimates fitted on the different data sets (using the same colour code as the panel for prevalence). The right-most panel of each quadrant block (panels C,F,I,L) compares the cumulative incidence estimated by the model fitted on the “Combined” data set to seroprevalence levels reported by the Canadian Blood Services for each city (grey point indicates the mean, the vertical grey bars show the 95% confidence intervals).

#### 4.1.2 Effective reproduction number

The effective reproduction number (ℛ_*t*_) is a key epidemiological parameter that has gained recognition and application during the COVID-19 pandemic [55, 56]. Using the same approach as for prevalence estimates, we inferred ℛ_*t*_ from epidemic trajectories generated from posterior distributions fitted to the three different data sources (*i*.*e*., “Clinical”, “WW” and “Combined”) and Equation 2. For comparison, we also calculate ℛ_*t*_ using the popular R package EpiEstim (version 2.2) [57] from the reported clinical cases as a separate approach. Results shown in Figure 4 exhibit the same behaviour as for the prevalence estimates, that is, mean estimates of ℛ_*t*_ are similar when trends of clinical and wastewater surveillance are comparable. Despite being based on a different modelling framework, estimates from EpiEstim are consistent with clinical-only estimates from our model. Finally, like for prevalence inferences, ℛ_*t*_ estimates from wastewater-only data (Figure 4, blue solid line) tend to have broader uncertainty interval compared to ℛ_*t*_ from clinical-only data.

**Figure 4:**
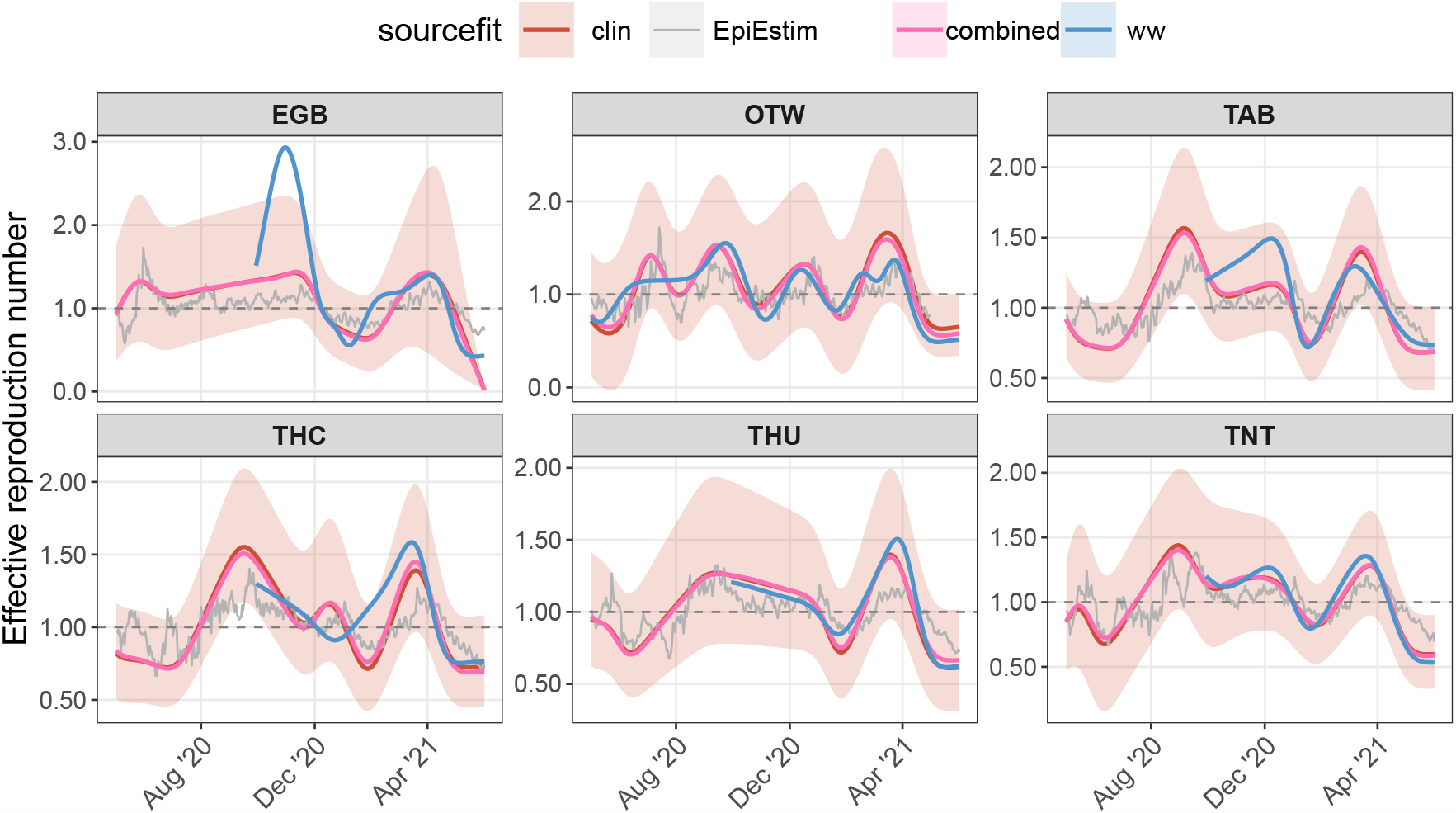
Effective reproduction number. Each panel represents a wastewater treatment plant. Solid lines represent the mean of effective reproduction number estimated with different methods (model presented here and, in grey, EpiEstim) and data sources (colour-coded). The ribbon indicates the 95% credible interval for the “Clinical” data source. Note that wastewater data are available from early October 2020 for Edmonton and Toronto. The ℛ_t_ curves were spline-smoothed, see Appendix for details.

#### 4.1.3 Forecasts

Another key advancement in our modelling framework is to generate forecasts based on clinical, hospital (if available) and wastewater data.

In Figure 5 we show three 1-month-ahead forecasting examples for Edmonton, Toronto Highland Creek and Ottawa using either wastewater data only, or clinical data only. The Edmonton example (Figure 5, left panel) shows forecasts made as of April 1st, 2021. In this case, the forecasts are relatively similar because both the clinical reports and the wastewater signals are comparable and the model fits were similar using either wastewater or clinical data. The Toronto Highland Creek example forecasts as of November 30th, 2020 (Figure 5, middle panel). For this location, the wastewater signal and clinical reports are discordant from December 2020 to February 2021. During this period the wastewater concentration is low and approximately flat whereas the clinical reports indicate a new wave of infections. As a result, the model fitted on these two data sources interprets the epidemic differently for this period and hence provides contrasting forecasts. The forecast for Ottawa as of December 15th, 2020 (Figure 5, right panel) illustrates the case when the wastewater forecast is more accurate than the one based on clinical surveillance only. At that time, the wastewater signal in Ottawa has picked up a resurgence earlier than clinical surveillance. This resurgence is then captured by the model fit and hence the wastewater-based forecast correctly projects the resurgence.

**Figure 5:**
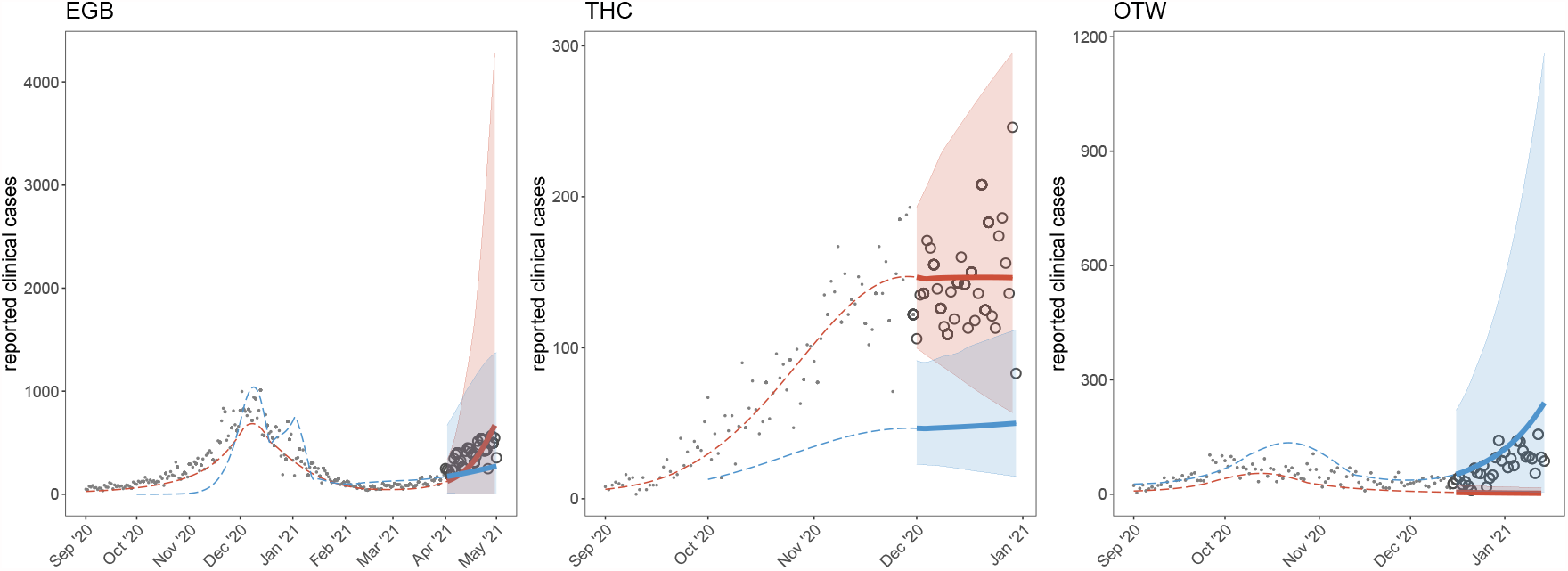
Forecast examples for Edmonton (left panel), Toronto/Highland Creek (middle panel) and Ottawa (right panel). Filled points represent past data of reported clinical cases. Circles represent reported clinical cases not yet observed at the time of forecast. Colour represents the type of data the model was fitted to (blue, wastewater only; red, clinical data only). Dashed coloured lines indicate the fitted mean for reported cases. The thick solid line shows the 1-month-ahead mean forecast, and the shaded areas their respective 95%CrI.

### 4.2 Simulations

In this section, we report results from simulations that provide general insights in interpreting WBS.

#### 4.2.1 Leading signal and reported detection differential

We vary the LOD of the wastewater assay across a broad, but realistic, range [58] and calculate Δ, the detection time difference, for each simulation for a given value of LOD and the clinical reporting rate *ρ*. Because SARS-CoV-2 RNA concentration in wastewater can only be determined up to a constant in our model, the LOD values chosen here are rescaled to the parameters used to run our simulations and cannot be directly interpreted as RNA copies per ml, the traditional unit for LOD. Panel A in Figure 6 shows that, depending on the LOD of the laboratory assay, the wastewater concentration of SARS-CoV-2 RNA can either be a leading (Δ *<* 0 for assays with low LODs) or a trailing indicator of cases (re)introduction when compared to reported clinical cases. This is the case whether the clinical surveillance system in the population is efficient or not (coloured curves, Figure 6A).

**Figure 6:**
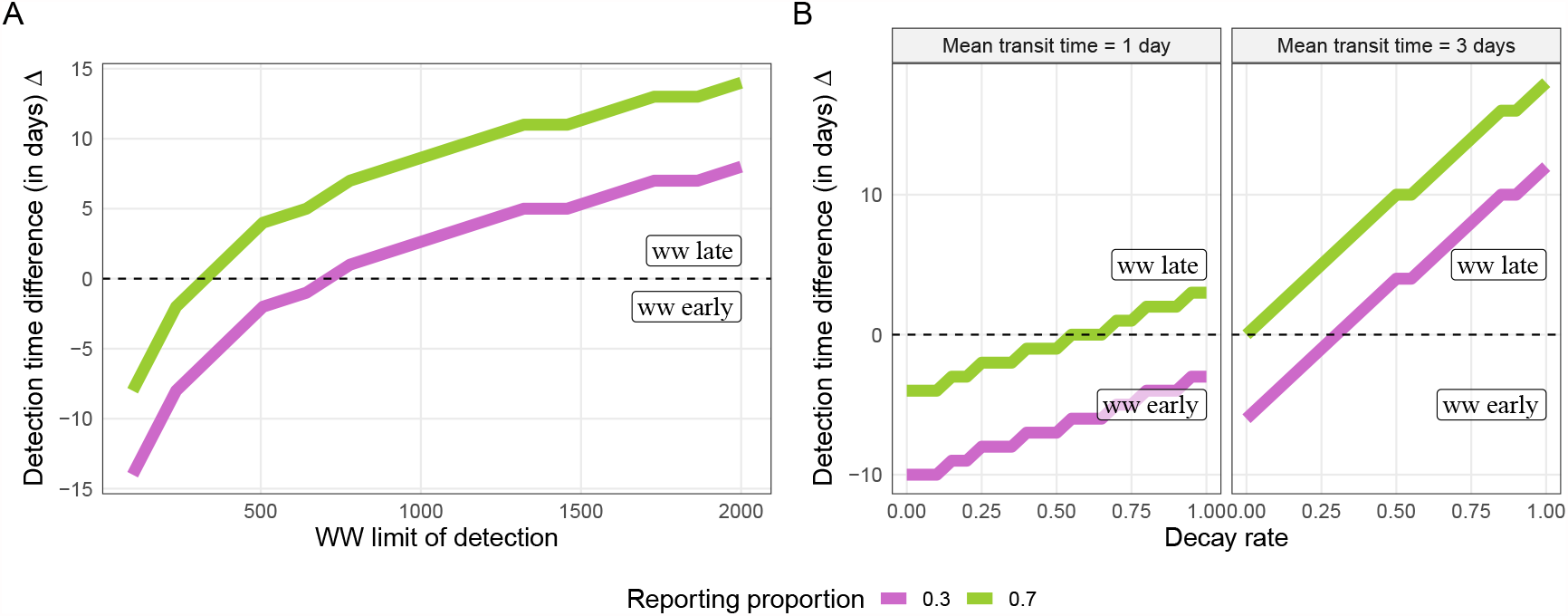
Simulations were run varying selected parameters to show their impact on the reported detection time differential (Δ). Panel A: effect of the limit of detection of the quantification assay performed on wastewater. Values below the 0-intercept horizontal dashed line indicate a leading signal from wastewater concentrations than from clinical reports. Panel B: effect of the SARS-CoV-2 RNA decay rate in wastewater for different transit times between the shedding and sampling site. The colour of the curves represents the proportion of clinical cases reported (ρ) out of the total symptomatic incidence.

We also vary the decay rate of RNA SARS-CoV-2 in wastewater within a broad realistic range [46, 59, 60], as well as the transit time of SARS-CoV-2 between the shedding and sampling sites. Figure 6B shows that, here again, the relative timing of (re)introduction detection by WBS compared to clinical surveillance can be affected by both the harshness of the wastewater (represented by the decay rate) and the transit time of SARS-CoV-2 in the sewer system. We note, as expected, that with a fast transit time (illustrated by a 1-day travel time in the left-most panel of Figure 6) the decay rate will not have a significant impact on Δ clinical surveillance (efficient, *ρ* = 70%, or not, *ρ* = 30%), but as the transit time increases to 3 days (an upper bound considering strong sediment and recirculation effects) the effect of RNA decay becomes more important (increasing slope for the 1-day and 3-day transit times, Figure 6B).

#### 4.2.2 Assessing public health intervention effectiveness

The ability to use wastewater signal to detect intervention effectiveness can also be explored using model simulations in order to explore if a public health intervention is more clearly observable in wastewater concentration measurements or in clinical reports. To do this, we choose to model an intervention such that the transmission rate *β*, constant until the intervention time, decreases linearly to a lower value *β/*3 during a period of time of *T*_interv_ and remains constant afterwards (this aims to crudely simulate a lockdown). We consider the relative difference between the observed peak and 7 days later for COVID-19 by clinical surveillance, *s*_cl_(*t*) = *C*(*t* + 7)*/C*(*t*) − 1 and the level of SARS-CoV-2 in wastewater, *s*_ww_ = *W*(*t* + 7)*/W*(*t*) − 1. The more negative the slope, the clearer the signal.

Using our baseline parameters (Table 2), we simulate an intervention that reduces the contact rate to a third of its pre-intervention value at different time of the simulations, ranging from 20 to 90 days after the introduction of the index case. Figure 7 shows that, overall, the effect of an intervention that significantly reduces transmission yields a larger relative decrease in number of COVID-19 cases than the level of SARS-CoV-2 in wastewater because the post-peak slope from clinical surveillance (*s*_cl_) is consistently more negative than the one from WBS (*s*_ww_). The difference is more pronounced as the change in transmission is more sudden (*T*_interv_ small). Hence, given the observation noise typically encountered, we expect that the effect of a sudden change in transmission rate would be more clearly observable from clinical surveillance than from WBS.

**Table 2:**
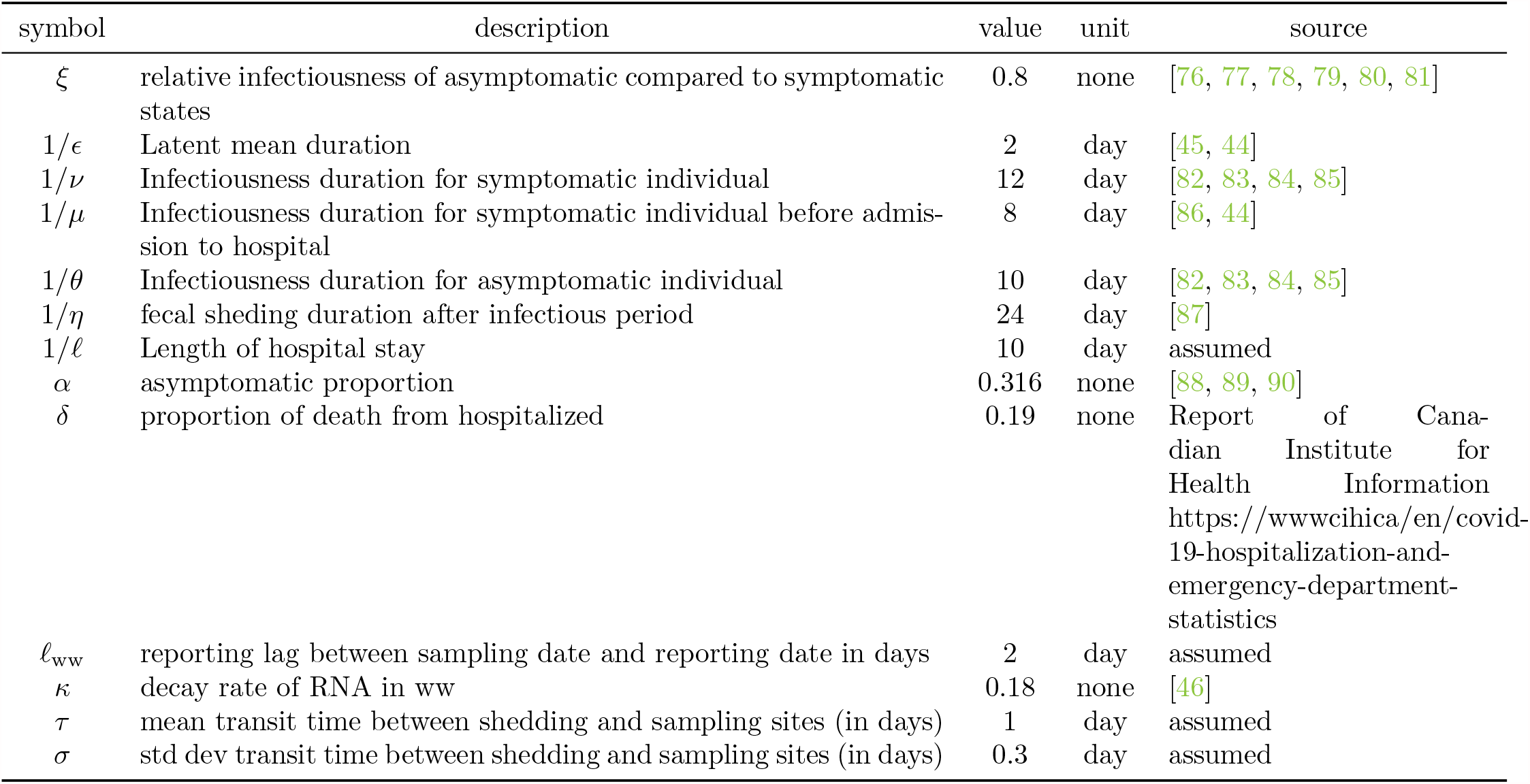
Description of fixed parameters used in this model and their sources

**Figure 7:**
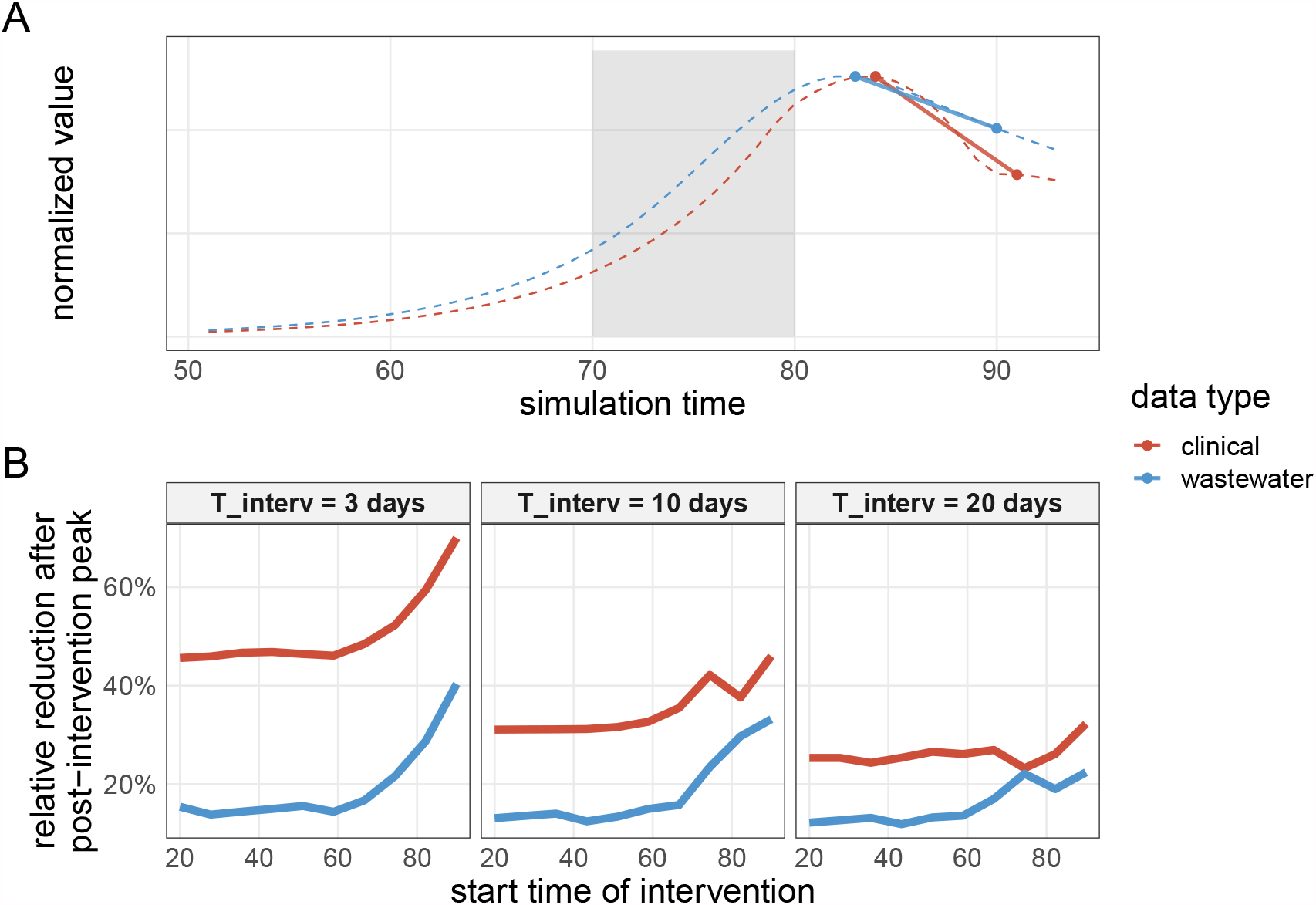
Detectability of a sharp transmission reduction. Panel A: example of how the post-peak relative changes are calculated. The colour-coded dashed lines represent the time series of reported clinical cases and SARS-CoV-2 RNA concentration in wastewater. The shaded area indicates when the transmission rate linearly decreases to a third of its baseline value (here, T_interv_ = 10 days). The segment illustrates the relative change between the peak value and 7 days later (s_ww_ and s_cl_). Panel B: the horizontal axis represents the time (since the start of the epidemic) when starts the intervention that reduces transmission to a third of its value. The vertical axis represents the post-peak relative changes from clinical reports (s_cl_, red lines) or wastewater (s_ww_, blue lines). Each panel indicates a different value (3, 10 and 20 days) for T_interv_, the time it takes to reduces the transmission rate to a third of its initial value.

#### 4.2.3 Differential impact of vaccination

In Figure 8 shows how *W*(*t*)*/C*(*t*), the ratio of reported wastewater concentration over reported cases, increases following vaccination with a infection-permissive vaccine. Indeed, while an infection-permissive vaccine does reduce transmission, it still allows for infections to occur (mostly asymptomatic) and in particular, faecal shedding. Hence, a smaller proportion of infections are reported (because most of them are asymptomatic) but faecal shedding is less affected by this reporting bias and level of SARS-CoV-2 in wastewater decreases less steadily than COVID-19 surveillance. In this simulation, the approximately constant ratio before the start of vaccination (Figure 8B) indicates that reports from clinical or wastewater data sources provide a similar picture of the epidemic for that period. However, once vaccination is implemented, the increasing ratio highlights a discordance between the two data sources.

**Figure 8:**
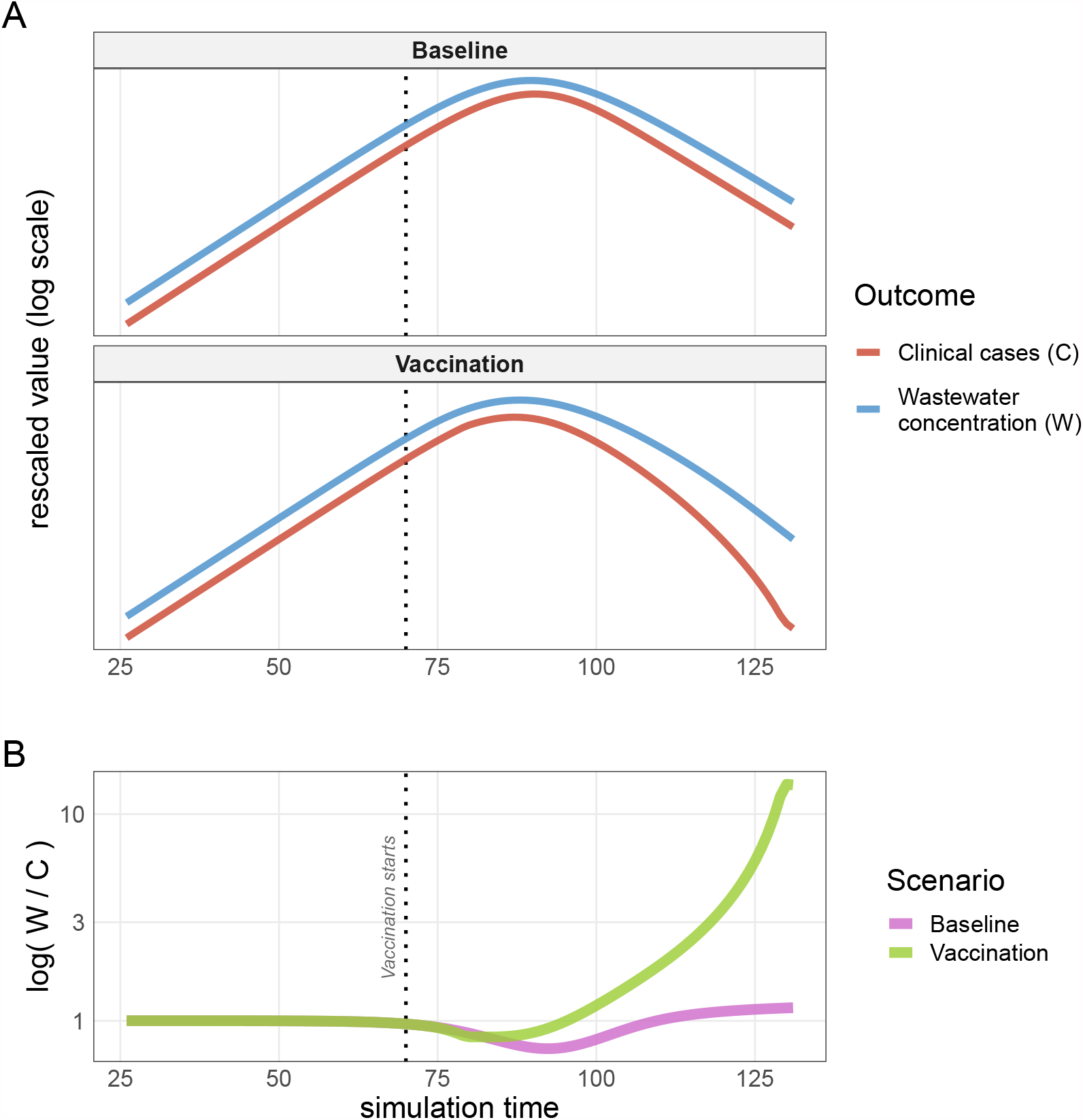
Infection-permissive vaccination. Panel A shows the trajectories of reported clinical cases and SARS-CoV-2 concentration in wastewater under a scenario using an infection-permissive vaccine (“Vaccination”), or not (“Baseline”). In the vaccination scenario, the reported clinical cases decrease more rapidly than the lavel of SARS-CoV-2 in wastewater because sub-clinical infections tend to be less reported whereas faecal shedding continues. Panel B highlights this difference showing W(t)/C(t), the ratio of reported wastewater concentration over reported cases, for the baseline / no-vaccination (pink) and the vaccination (green) scenarios. The ratio is normalized to have a starting value at 1 to make it easier to quantify the increase visually. The vertical dotted line indicates when vaccination starts (at time 70).

## 5 Discussion

Surveillance through detection and quantification of targeted pathogens in wastewater has been a noteworthy tool for public health across the world [61, 16, 18, 10, 11, 12, 13, 14]. While pathogen surveillance in wastewater is not new, the scale and urgency of scientific development for WBS are witnessed during the unprecedented COVID-19 pandemic. Because of the novelty of SARS-CoV-2-related WBS and the lack of quantitative tools for analysis, the interpretation of levels SARS-CoV-2 in wastewater and their translation into actionable public health measures is still challenging [31, 32, 16, 33].

Here, we have provided a modelling framework to improve the understanding of the mechanisms at play between the viral transmission in the population and viral concentration shed in wastewater. This model can also provide estimates of key unobserved epidemiological parameters. We demonstrated the applicability of our model by fitting it to data from three Canadian cities and made wastewater-informed inferences of important epidemiological metrics (prevalence, effective reproduction number and forecasted incidence). Our estimates for cumulative incidence were approximately in line with seroprevalence levels from cohorts of blood donors independently observed in the three cities.

Importantly, we observed that estimates based on wastewater-only data usually provide a similar picture of the epidemic trajectory (Figure 3) but discordant signals can occur and lead to drastically different interpretations. This was the case, for example, in January 2021 in Toronto Highland Creek where the wastewater signal did not indicate a resurgence of infections, despite the wave observed from the reported clinical cases. We believe this muted peak in wastewater signal was not caused by a laboratory issue, but rather from undetermined events in this particular sewershed at that specific time that need to be further investigated.

Similarly, the effective reproduction numbers inferred from wastewater data only are consistent with more traditional methods, such as using clinical reports with the popular software EpiEstim (Figure 4). The fact that wastewater data can potentially act as a substitute for clinical surveillance (albeit with more uncertainty) to provide critical epidemiological metrics is encouraging, although more realistically, it will likely act as a complementary data source. The ability to estimate epidemiological metrics using wastewater surveillance represents a step forward in demonstrating the use of wastewater data for actionable public health metrics, if they are available to public health in a timely manner. In addition, the ability to triangulate the state of an epidemic using alternative data sources helps ensure additional confidence in the estimation of relevant parameters or forecasting. Indeed, the COVID-19 pandemic has consumed public-health resources at levels that are probably not sustainable for long-term surveillance of this pathogen. However, the current wastewater surveillance performed in many communities can probably be continued as long as necessary given its relative low cost [9].

Our modelling framework provides a more principled alternative to simpler smoothing techniques (*e*.*g*., moving averages, polynomial interpolations) that have been used to support the interpretation of WBS [28]. However, we note that less complex modelling options are possible if the focus is on specific epidemiological metrics ([62, 63]). We also note recent efforts to use machine learning techniques and artificial neural network that incorporate WBS [64]. While those methods are promising, they cannot–by design–explain the epidemiological mechanisms at play.

Our model enables *in silico* experiments on the epidemic/wastewater system to identify key parameters and processes that can play an important role for the epidemiological interpretation. We showed, using simulations, that the relative timing of the wastewater signal (whether it is leading or not) compared to traditional clinical surveillance is actually influenced by the characteristics of *both* systems (Figure 6). On the one hand, the laboratory analysis of wastewater samples may not detect the presence of SARS-CoV-2 because, for example, its limit of detection is too high, or prevalence of infection in the community is very small, or the viral RNA has degraded before reaching the sampling site. Shipment time of wastewater samples can also be significant (*e*.*g*., several days) for remote sampling locations without any laboratory capacity. On the other hand, the delay in clinical cases reports is usually caused by the incubation period and the reporting time of an infection by the health system (turnaround time for contact tracing and/or laboratory results of individuals’ swab) or availability of testing. Some communities would typically have a longer lag for clinical reporting than for wastewater surveillance [20, 22, 28, 24], while others may have the opposite (for example when a very effective contact tracing system is in place [65] or rapid testing is implemented). Moreover, a community may experience both situations, that is a period when clinical surveillance is extremely efficient at detecting cases so rapidly that it leads wastewater surveillance while, at other times, it can lag (for example when incidence is high, overwhelming contact-tracing and clinical testing capacities). The model presented here allows to quantify how various factors can impacts the relative timing between clinical and wastewater surveillances.

It can be tempting to monitor the effect of public health interventions using changes in the levels of SARS-CoV-2 in wastewater given its non-invasive nature. Indeed, WBS should be less affected by sampling bias than clinical surveillance (for example the latter may miss most of the subclinical infections). However, our simulations showed that wastewater surveillance may be inferior to clinical surveillance to identify sharp declines in transmission, as typically seen after a lockdown is implemented (Figure 7). The long period of faecal shedding creates a lag in comparison to the sudden drop of incidence caused by the public health intervention, inhibiting a prompt signal in wastewater. This effect is visible on the Canadian data sets presented here (Figure 2).

We also highlight the potential for an infection-permissive vaccine to generate discordant signals between wastewater and clinical surveillances (Figure 8). Indeed, vaccination implies a larger proportion of asymptomatic infections which are less likely to be detected by clinical surveillance, but still picked up in wastewater because of continued faecal shedding. Note that we use our model to highlight this potential effect, whereas detecting it in real data is probably challenging without studies purposely designed to detect this.

Here, we used a mixed approach regarding the normalization of the levels of SARS-CoV-2 in wastewater, with Ottawa using PMMV-normalization versus TSS-normalization for Toronto and Edmonton. It is likely that some normalization is necessary to discount the variations of the total faecal mass shedded and viral degradation, but it is still not clear which normalization is the most appropriate for a given sewershed. We note that we considered separately each WWTP in Toronto to provide examples of application at the sub-municipal level.

Our modelling approach has several weaknesses. We did not precisely model the transport and fate of SARS-CoV-2 in municipal sewer systems. The lack of data about flow dynamics and particles binding of SARS-CoV-2 in wastewater hampered a more detailed approach. Hence, we took a simple approach to model the below-ground component and assumed the flow dynamic followed a low-dispersion plug flow model with a plausible fixed decay rate [46] and varied the mean transit time (from shedding to sampling sites) within a range of possible values. As more research focuses on the fate of SARS-CoV-2 in wastewater, the transport module of our model can be enhanced.

The comparison with observed seroprevalence level must be done with caution, because we do not model seroreversion (patients who were infected but subsequently test seronegative because of loss of immunity or antibodies falling to undetectable levels). Our model does not model vaccination explicitly. We made this choice to keep the first version of our model relatively simple. However, we believe that we can appropriately approximate the effects of infection-permissive vaccination by reducing the transmission rate and increasing the proportion of asymptomatic infections. As the proportion of vaccinated individuals increases, modelling an explicit vaccination process is necessary. We note that for the Canadian cities studied here, the vaccination coverage was either null or low during the study period.

We model SARS-CoV-2 as a single-strain pathogen which is an oversimplification of reality, given the numerous variants circulating in Canada since late 2020 [66]. However, it is not clear how (or if) multi-variants modelling would affect our results, given that the difference of viral shedding (respiratory and faecal) between variants is still not fully understood [67, 68].

Because of ordinary differential equations, this model is not well adapted to either small communities or very low prevalence settings. While its epidemiological structure (Figure 1) would still be valid for such environments, a more advanced statistical modelling would be preferable to handle low incidence counts and observation uncertainty [69, 70].

A further limitation of our model is the use of a scaling coefficient for the amount of SARS-CoV-2 shedded in the wastewater by the infected population (parameter *ω* in Equation 3). This scaling coefficient embeds all the uncertainties associated with sampling strategy and laboratory analysis, such as assay recovery efficiency, limit of detection, and total faecal mass normalization. Most of those processes are currently poorly known for SARS-CoV-2 and, as long as more observational data is not available, will constrain modelling (note that this limitation has already been identified for polio models [14]). An ultimate goal of wastewater surveillance may be to measure all the components of the scaling coefficient (here, *ω*) in order to estimate infection prevalence in a community directly from viral concentration readings.

To conclude, the model presented here–built upon previous similar approach for other pathogens [71, 14, 72]–is a first step to better understand the mechanistic relationships between the COVID-19 epidemic spreading in a community and the SARS-CoV-2 RNA concentration in wastewater caused by faecal shedding of infected individuals (and potentially from urinary or sputum shedding). Future developments should explicitly incorporate vaccination and multiple variants/strains given the ability of new assays to detect variants from wastewater samples [73, 74, 75]. This model can be the basis of quantitative tools to support public health decision making that embraces wastewater-based epidemiology. Beyond the SARS-CoV-2/COVID-19 pandemic, WBS coupled with the type of model presented here could be leveraged and applied to other transmissible pathogens where urinary or faecal shedding occurs, such as other respiratory diseases (*e*.*g*., influenza, respiratory syncytial virus, adenovirus) and some enteric diseases (*e*.*g*., norovirus, rotavirus, shigellosis).

## Supporting information

data availability statement

fit outputs

## Data Availability

All data to reproduce this work will be available as a supplementary file upon publication.

## 6 Acknowledgments

- Wastewater samples from Edmonton and Toronto were provided to the NML through a collaboration with Statistics Canada’s Canadian Wastewater Survey
- Dave Spreitzer, Ravinder Lidder, Codey Dueck, Quinn Wonitowy, Umar Mohammed and Graham Cox for their contribution in wastewater sample processing and technical assistance.
- Dana Al-Bargash provided data and local expertise for the City of Toronto.
- The wastewater treatment plant Gold Bar, EPCOR Water Services Inc., Edmonton, Alberta, Canada for providing sewage samples in this study for the city of Edmonton.
- Peter Vanrolleghen’s group at the University of Laval (QC, Canada) for insightful discussions on viral transport and fate in wastewater

## Appendix

**Figure S1:**
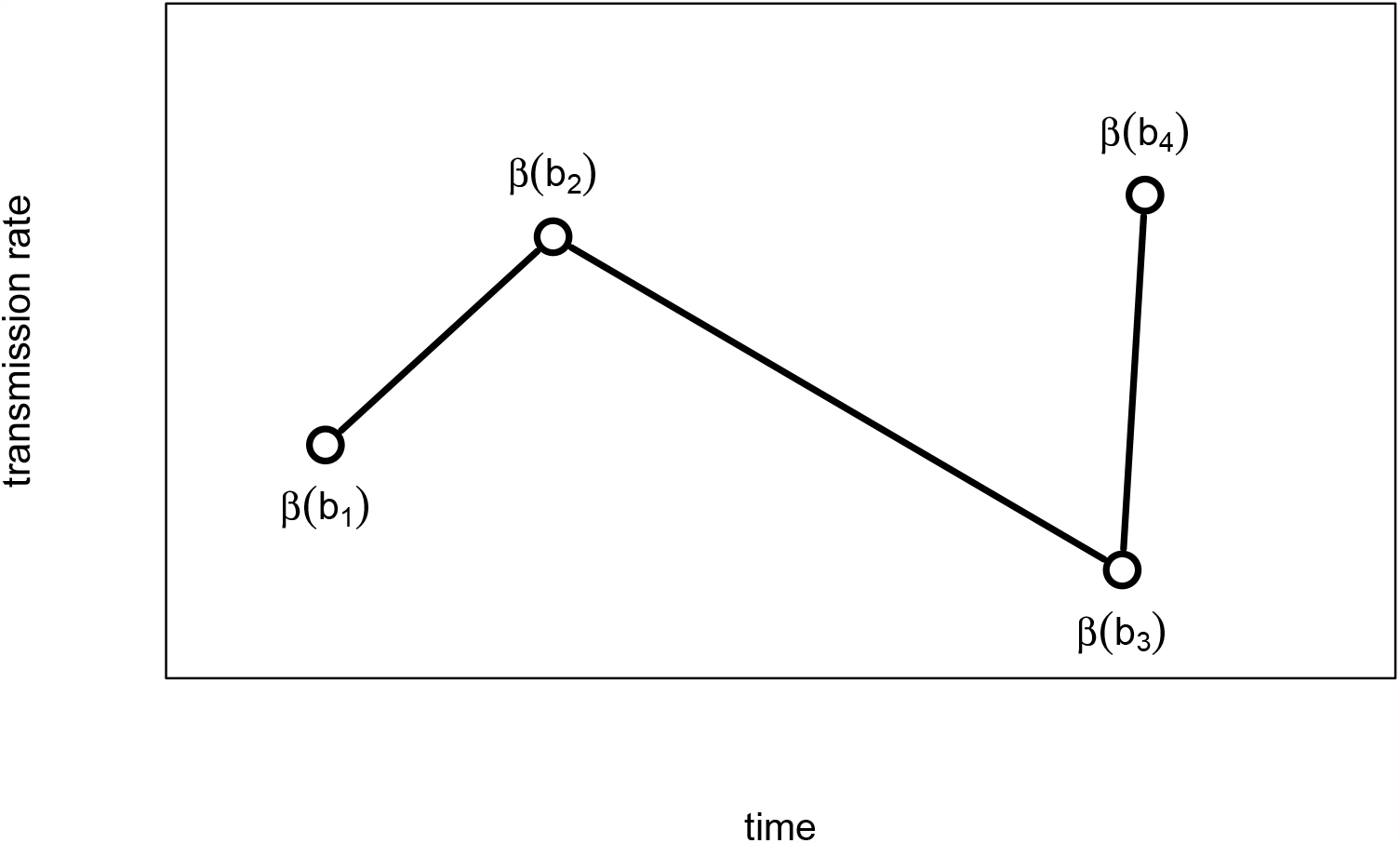
Illustration of the piecewise linear function to model the transmission rate β_t_. The break dates b_1_, b_2_, … are chosen manually by visual inspection of the time series of interest, and the values β(b_i_) are fitted with an ABC algorithm.

### A-1 Fitting procedure

To fit the model to the clinical and/or wastewater surveillance data, we model the transmission rate, *β*_*t*_ as a piecewise linear function. We parsimoniously choose the times *b*_1_, *b*_2_, … (“break times”) defining each segment by visual inspection of the time series (*i*.*e*., incidence of new COVID-19 cases and/or hospitalizations for clinical surveillance; SARS-CoV-2 concentration for wastewater surveillance). Those times should correspond to changes in the transmission dynamics. The value of the transmission rate at a break date, *β*(*b*_*i*_) is fitted using an Approximate Bayesian Computation (ABC). We chose not to fit the break times *b*_*i*_ because the fitting algorithm would require a computation time that would not be practical. See Figure S1.

The mean transit time 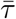 and the scaling factor *ω* are also fitted to data. For those parameters, the goal is primarily to allow for uncertainty rather than infer a posterior distribution as they are essentially not identifiable. Finally, the hospitalization rate is also fitted to the hospitalization data, when available.

Over the study period (from early March 2020 to June 1st, 2021), we have about a dozen break times *b*_*i*_ for each city to reflect the various interventions (*e*.*g*., lockdowns) or behaviour (*e*.*g*., change in contact rate when schools re-opened in September 2020). Hence, the parameter space to explore by the ABC algorithm is relatively large. To avoid unpractical computational times, we defined relative strong priors on all parameters, that is normal distributions with a mean close to the expected value (explored manually) and a relatively broad standard deviation (corresponding approximately to a coefficient of variation of 0.5). The normal distribution was censored to positive values. The outputs of the fitting procedure for all locations are shown in supplementary file File S2.

### A-2 Relative infectiousness

In Equation 1a, the force of infection from symptomatic cases is 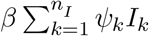. For convenience, the *ψ*_*k*_ are chosen such that

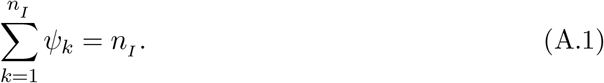

Hence, when the infectiousness profile is constant (λ_*k*_ = 1, for *k* = 1, …, *n*_*I*_), the force of infection is 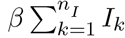. Using this normalization allows to keep a single baseline parameter *β*. Only the *relative* values of the *ψ*_*k*_ affect the infectiousness profile. Similarly for asymptomatic infections, the parameters *ϕ*_*k*_ are chosen such that 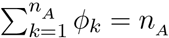.

We assume the infectious period for symptomatic infections is 12 days on average [91, 92, 85, 82] and divide this period into *n*_*I*_ = 6 sub-compartments *I*_1_, …, *I*_6_ where infected individuals will stay, on average, 2 days in each of them. Note that with this representation, the duration of infectiousness has an Erlang distribution [93] with shape *n*_*I*_ (and mean 12 days). The parameter *ψ*_*k*_ represents the relative infectiousness of sub-compartment *I*_*k*_. We assume infectiousness, that is the probability of transmission given contact, is proportional to the log viral load measured from respiratory samples in clinical studies [83, 84] and choose 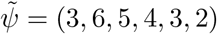 and then normalize according to Equation A.1 with 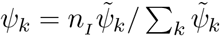.

Similarly, we assume a shorter infectious period for asymptomatic infections of 10 days on average [94, 91], divided into *n*_*A*_ = 5 sub-compartments with an average stay of 2 days each.

### A-3 Respiratory and faecal Viral kinetics

Our model explicitly accounts for the temporal profile of respiratory shedding via the multiple sub-compartments for the infectious states (*A, I* and *J*) combined with the parameters *ϕ* and *ψ*. Similarly, it explicitly accounts for the faecal shedding kinetics via the shedding states (*A, I, J* and *Z*) and the parameters λ.

**Figure S2:**
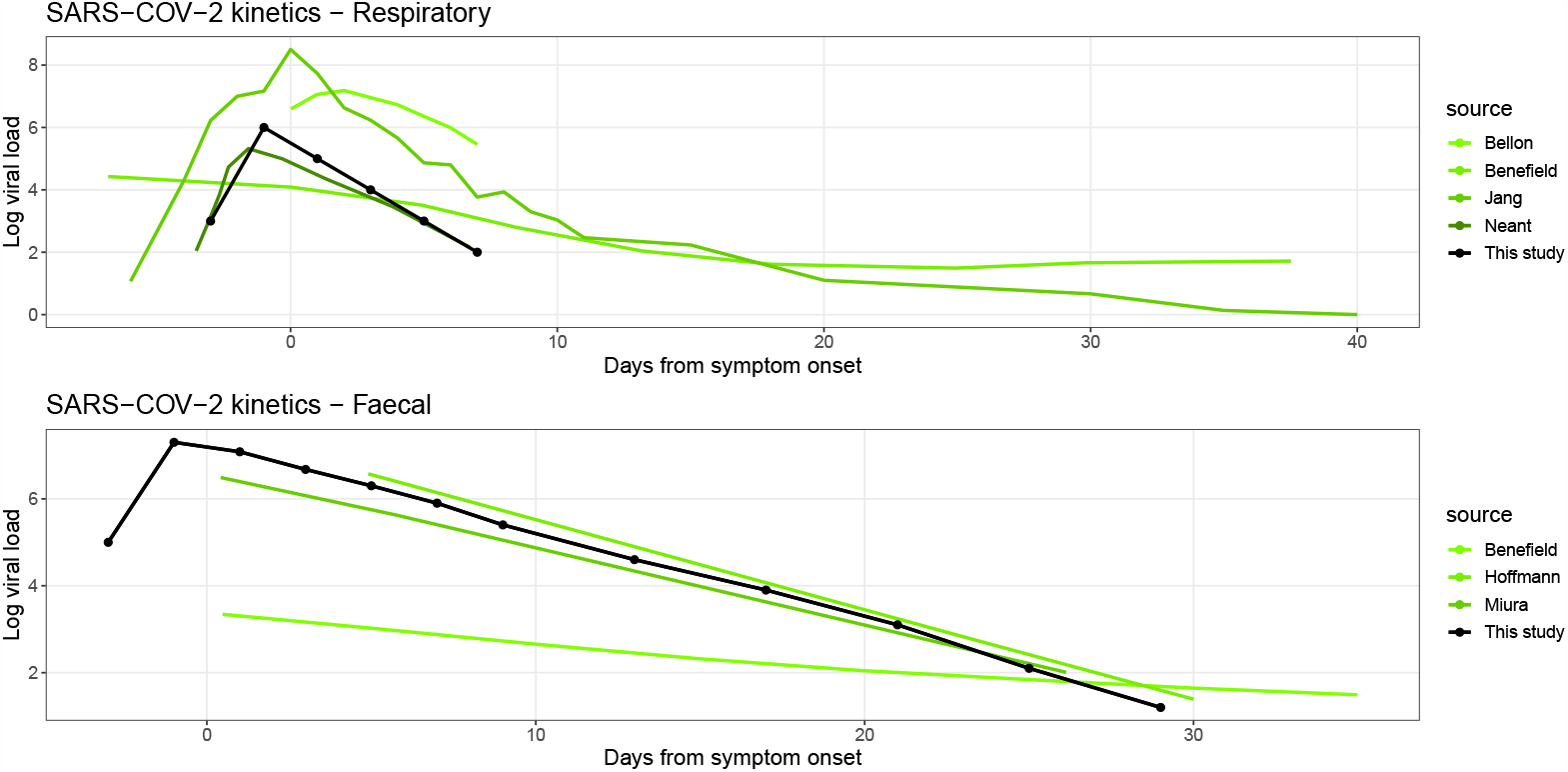
SARS-CoV-2 viral kinetics. The values used in our model are represented by the black curve and the green curves show estimates from the literature. The full reference of each study can be found in the bibliography: Benefield [95], Bellon [96], Jang [83], Neant [84], Hoffmann [87], Miura [97].

We parameterize our model such that the SARS-CoV-2 viral kinetics reflect with levels reported in the literature. The black line with points in Figure S2 shows the values used for respiratory (top panel) and faecal (bottom panel) shedding, and how they compare to observational studies.

### A-4 Effective reproduction number

As a first step, we establish the *basic* reproduction number, ℛ_0_, for the model defined by equations 1a-1n. To derive ℛ_0_, we follow the methodology presented in [42].

Asymptomatic individuals are infectious for an average duration of 1*/θ*, and the ratio of their infectiousness compared to all the other infectious states (*I* and *J*) is *ξ*. Asymptomatic incidence is a proportion *α* of the overall incidence. Hence, the contribution to the basic reproduction number from the asymptomatically infected individuals is

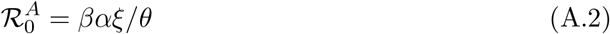

Infectious individuals that are symptomatic and that will recover without hospitalization (*I*) are infectious for an average duration of 1*/ν*. Their proportion of the overall incidence is calculated by simply stating they are not hospitalized (1 − *h*) and not asymptomatic (1 − *α*), hence the proportion is (1 − *h*)(1 − *α*). The contribution to the basic reproduction number from the symptomatically infected individuals that will not require hospitalization is

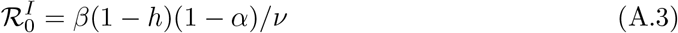

For the symptomatically infected persons that will require hospitalization (*J*), the same logic applies to the *I* sub-group, except that we need to take into account their relative infectiousness compared to *I*, which is 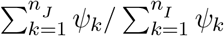. Note that the denominator is simply *n*_*I*_ thanks to the normalization defined in Equation A.1. The subgroup in state *J* are hospitalized (*h*) and not asymptomatic (1 − *α*) so their contribution to the basic reproduction number is

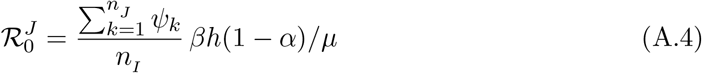

We obtain the (overall) basic reproduction number by summing the respective contributions from all infectious epidemiological states

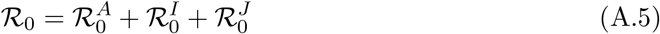

Finally, the *effective* reproduction number ℛ_*t*_ is simply defined as 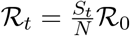 which gives Equation 2 in the main text.

### A-5 Fate and RNA transport in wastewater

Upon fecal deposition into the wastewater, viral RNA undergoes various hydrodynamic processes and degrades during its journey from the shedding point to the sampling site [16, 33]. This degradation mainly comes from RNA dilution in municipal wastewater constitutes (*e*.*g*., hygiene products, household detergents, industrial wastewater and storm waters) and RNA decay resulting from harsh wastewater environment (*e*.*g*., temperature, bioactive chemicals, solids, pH, etc.). As a common practice, complex hydraulic models simulate the in-fluid transportation of the water contaminants (endemic viruses and fecal microorganisms) via solving a set of physics-based differential equations describing the flow and transport mechanisms [98, 99, 100, 101, 102, 103, 104].

These hydrodynamical models are advection and dispersion mechanisms, which describe the microorganisms’ transportation by the flow velocity along the longitudinal axis and its diluting process into the surrounding fluid [103]. Once a mass concentration enters the stream, it gradually disperses due to many physical factors such as dissolving process, velocity profile, turbulent mixing, molecular diffusion, etc. [105]. Moreover, sediment association plays a significant role in the transport models. Several studies indicated attachments of microorganisms to sediments and the impact of this associations on their delayed transportation [104, 102, 100]. Due to solid mass and subsequent gravitational pull, sediment’s velocity differs from the flow velocity and results in solid settlement at the bottom of the stream. These microorganism-attached sediments, later, re-suspend during overflow periods, induced by heavy rainfall and industrial discharges, and act as a reservoir and increase in-stream microorganism’s concentration irrespective of new input from the shedding source [106, 107]. Enveloped viruses, such as SARS-CoV-2, dissolve less in water and tends to attach to solids as a result of their hydrophobic envelope [39, 33], and therefore, settlement/resuspension may play a key role in transportation of the SARS-CoV-2 genetic materials in wastewater.

**Figure S3:**
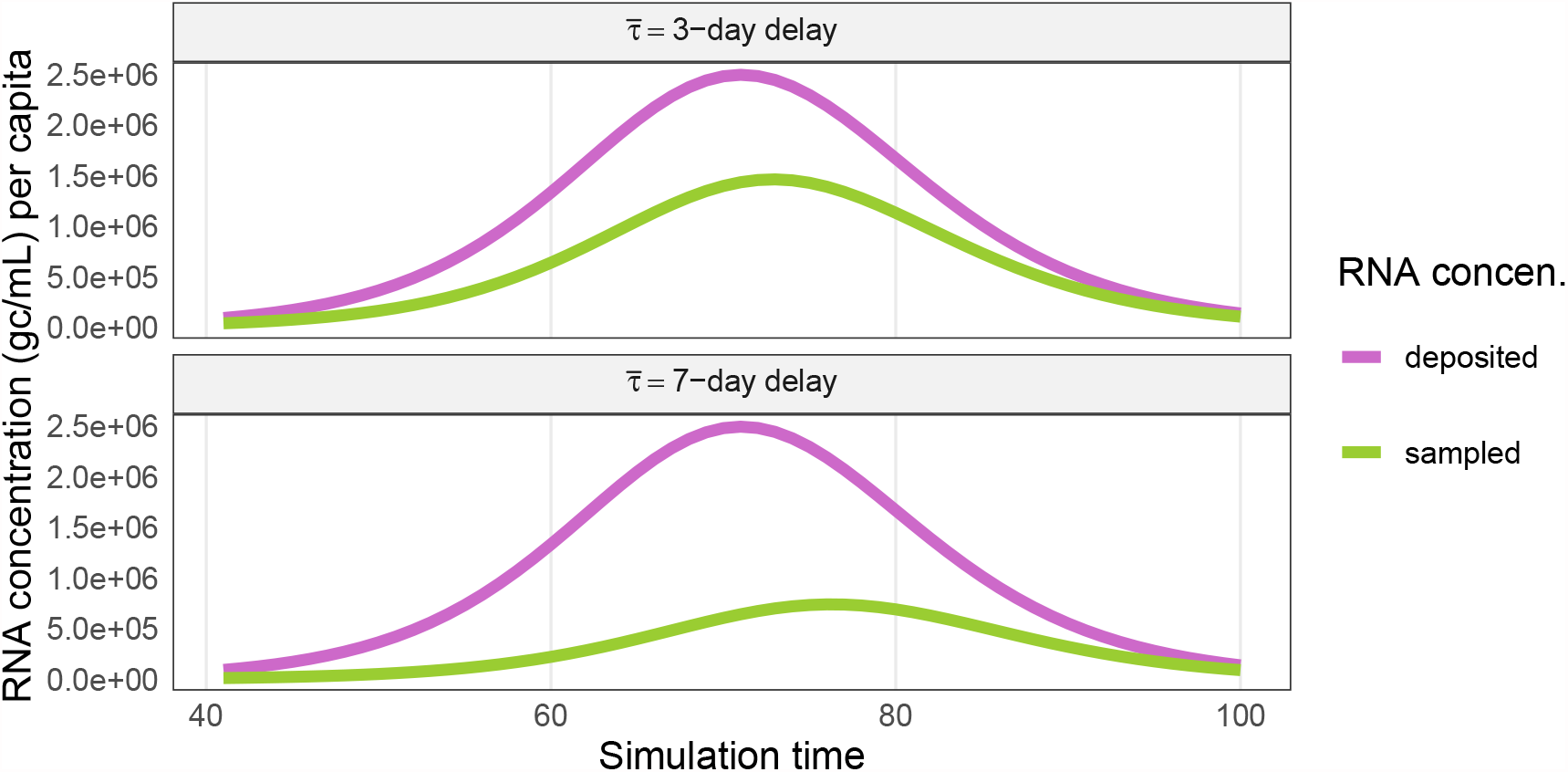
Impact of advection-dispersion-decay model (Eq. A.8). sampled RNA concentration (W_samp_) may have different viral distribution compared to deposited density (W^∗^) due to delayed viral genetic materials resulting from hydrodynamical phenomenon in sewer system.

Here, we used a simple advection-dispersion-decay model to simulate the fate of SARS-CoV-2 RNA along their journey from shedding points to the sampling site. We assumed the deposited RNA concentration in the sewage system is a one-time pulse input per day and described the transfer process by a dispersed plug-flow model. As the plug concentration of viral RNA enters into the flow with velocity *u*, it gradually disperses along the longitudinal axis (flow direction) with dispersion coefficient 𝒟 (m^2^/s) within the sewage pathway. The length from the shedding location to the sampling site is *L*. The total RNA mass arrives at the sampling point gradually over time, such that the daily pulse input concentration is delayed. Assuming a small deviation from plug flow (𝒟 */uL* ≤ 0.01), we can use the analytical solution of the 1-dimensional axial dispersed plug flow differential equation [47, 105] as a transfer function for viral RNA in wastewater. For low diffusion limit, the transfer function is approximately symmetrical and defined by a Gaussian distribution *g*(*τ*), which represents the fraction of the deposited concentration at the sampling site *τ* days after its introduction to the sewage system

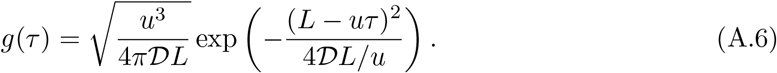

Defining the mean transit time 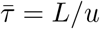 and its standard deviation 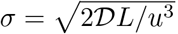, we can re-parametrize Equation A.6 based on the first two moments of the transit time:

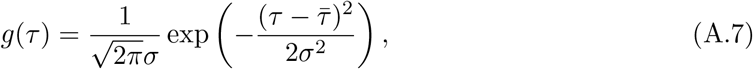

In addition to delay, the genetic materials of SARS-CoV-2 degrades exponentially at a daily rate *κ* due to the complex environment of wastewater [46]. As a result, *W*(*t*), the sampled concentration at time *t*, is a combination of both delayed viral materials and degradation

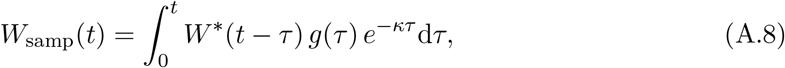

where *W* ^∗^ is the initial daily deposited concentration.

### A-6 ℛ_*t*_ posterior estimates

The effective reproduction number ℛ_*t*_ is estimated by fitting the model to different data sources (see main text). We also estimate ℛ_*t*_ using the R library EpiEstim [57] on reported cases.

The function ℛ_*t*_ is modelled as piecewise-linear. As a result, ℛ_*t*_ from Eq. 2 is also a piecewise linear function. We interpolated ℛ_*t*_ with a polynomial curve (by smooth spline function in R) for Figure 4. For validity purpose, we compared the interpolated and the piecewise linear function ℛ_*t*_ and checked the smoothed and actual curves are similar. See Figure S5.

**Figure S4:**
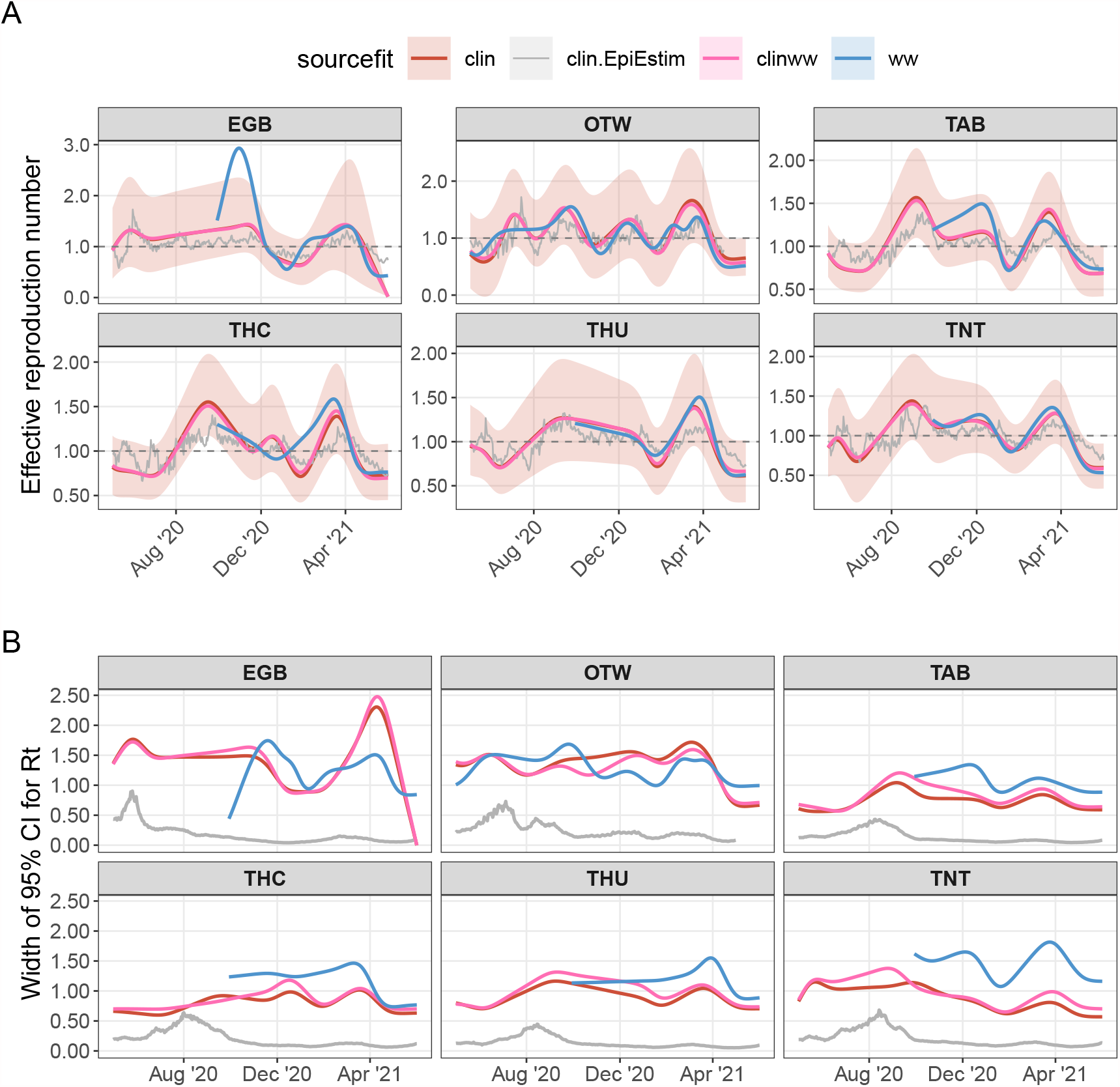
Panel A: Effective reproduction number inferred from model, utilizing different data sources, for all WWTP’s locations. ℛ_t_ inferred by fitted model to clinical reported cases (clin), RNA wastewater concentration (ww), and both clinical reported cases and viral load in wastewater (combined clinww). Panel B: Associated 95% credible interval widths.

**Figure S5:**
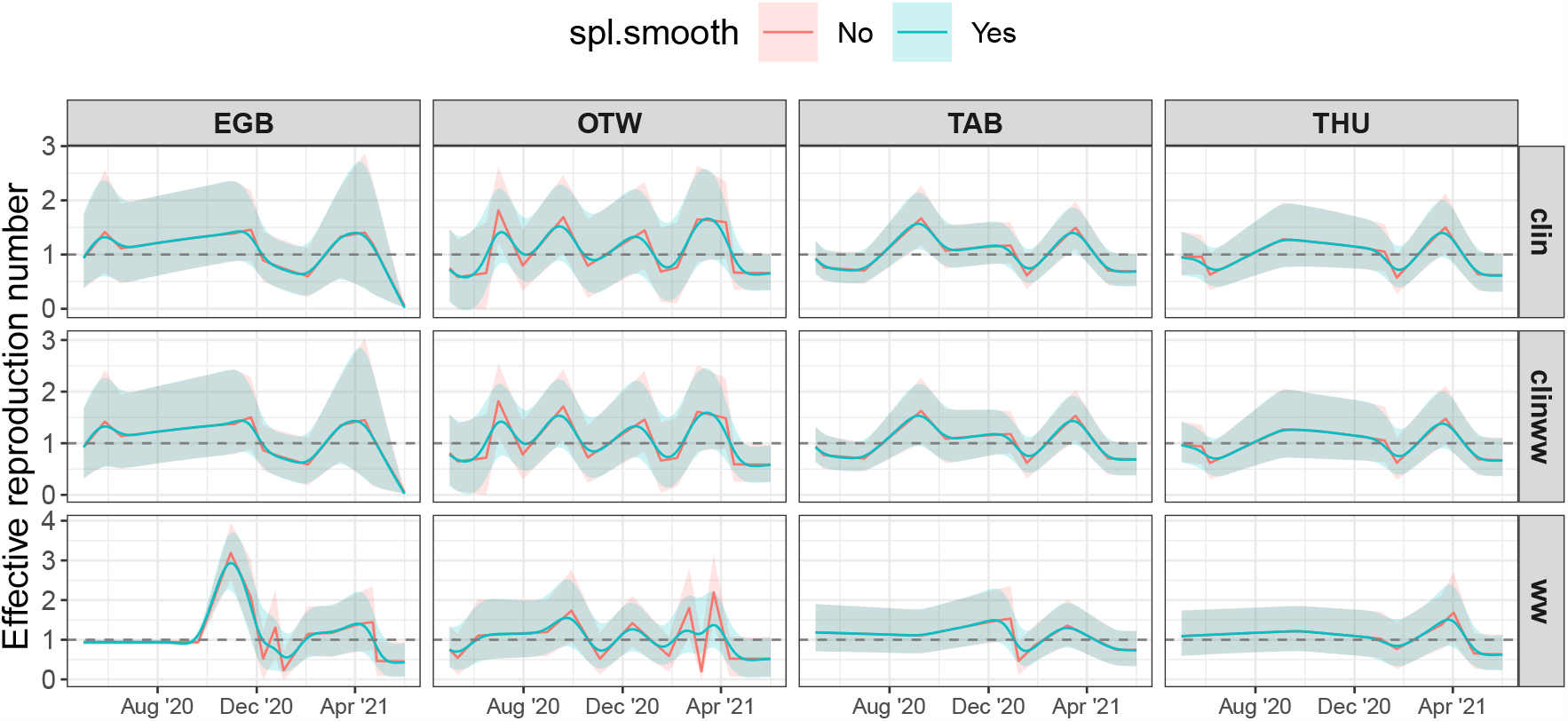
The effective reproduction number and its 95% credible interval plotted for different sources (clin, ww and combined clinww) of fitting in wastewater sampling locations; Edmonton (EGB), Ottawa (OTW), Toronto Ashbridges Bay (TAB) and Toronto Humber (THU). Colours represent ℛ_t_ obtained from model and Eq. 2 (pink) and its interpolated smoothed spline (green).

