## Supplementary material for "A Wastewater-Based Epidemic Model for SARS-CoV-2 with Application to Three Canadian Cities": fit outputs: _README.txt

The files in this directory provide the output of ABC fitting procedure (see main text and appendix for details).


=== File naming convention ===

plot-fit-[LOCATION_CODE]-[ID]-[DATA_TYPE_SOURCE].pdf

LOCATION_CODE: Location code for the wastewater treatment plant. EGB for Edmonton, OTW for Ottawa, TAB for Toronto Ashbridges, THC for Toronto Highland Creek, THU for Toronto Humber, TNT for North Toronto

ID: Identification number.

DATA_TYPE_SOURCE: Type of data used for the model fit. ww: wastewater data only; clin: clinical reports and, when available, hospital admissions; clinww: both clinical and wastewater data.


=== Page description ===

All the files have the same content.

Page 1: Data for this location

Page 2: Fixed break dates for the function beta_t. The values (y coordinates) are fitted to data (see page 4 and 5)

Page 3: Ranked errors from the ABC fit. 

Page 4: prior (grey) and posterior (red) distributions for all fitted parameters. the vertical line represent tehe mean of the posterior distribution.

Page 5: Mean, 50% and 95% creditble intervals for the fitted parameters.

Page 6: Model simulation from the posterior distribution (line: mean, ribbon: 95% credible interval). Points: observed data. This figure is a way to visually check if the fitted model reproduce correctly the observed data.
