## Supplementary material for "A Wastewater-Based Epidemic Model for SARS-CoV-2 with Application to Three Canadian Cities": fit outputs: plot-fit-EGB-07-06-1201-ww.pdf

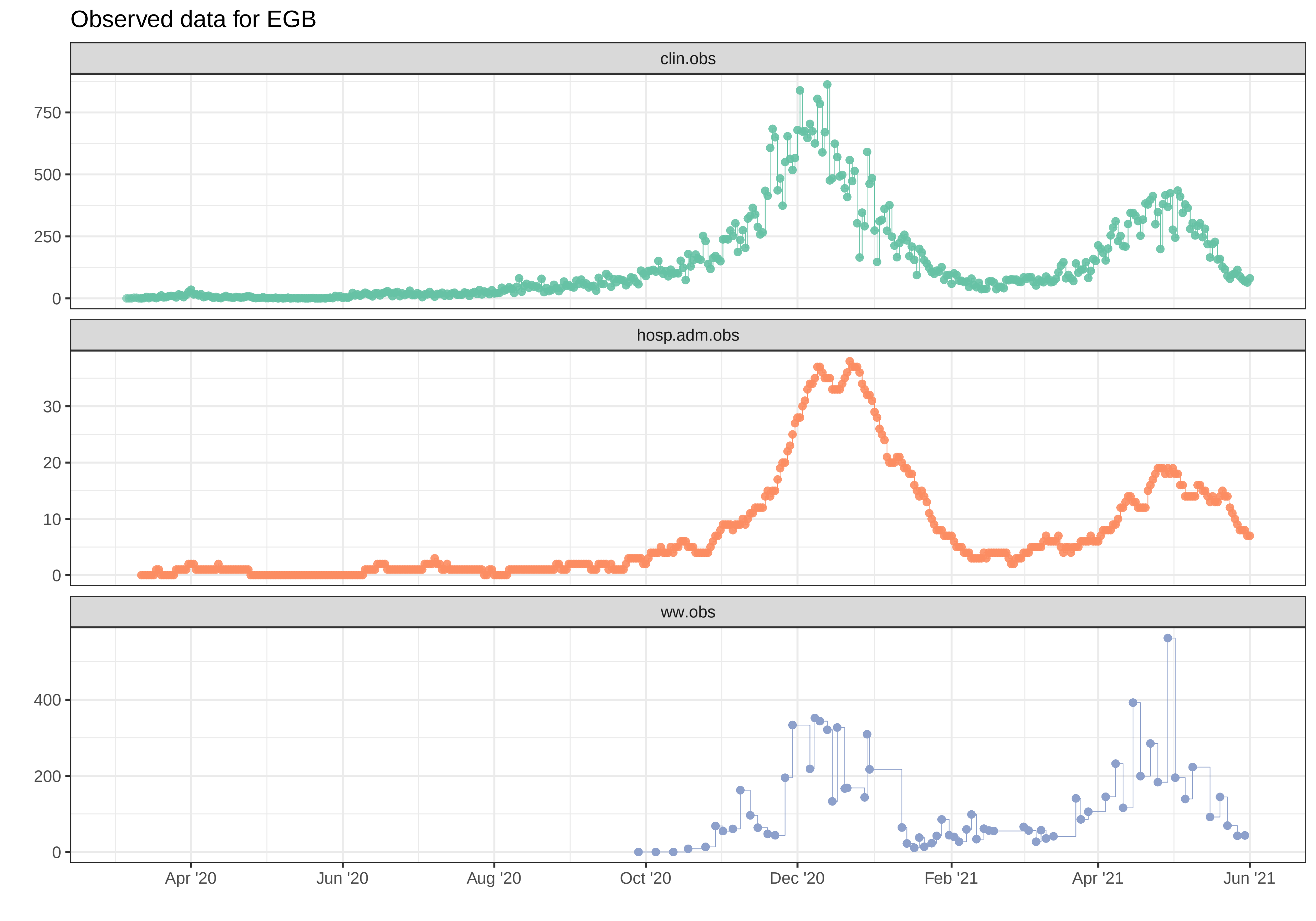
Observed data for EGB
clin.obs
hosp.adm.obs
ww.obs
0
250
500
750
0
10
20
30
0
200
400
Apr '20
Jun '20
Aug '20
Oct '20
Dec '20
Feb '21
Apr '21
Jun '21

Simulation with initial parameters for EGB

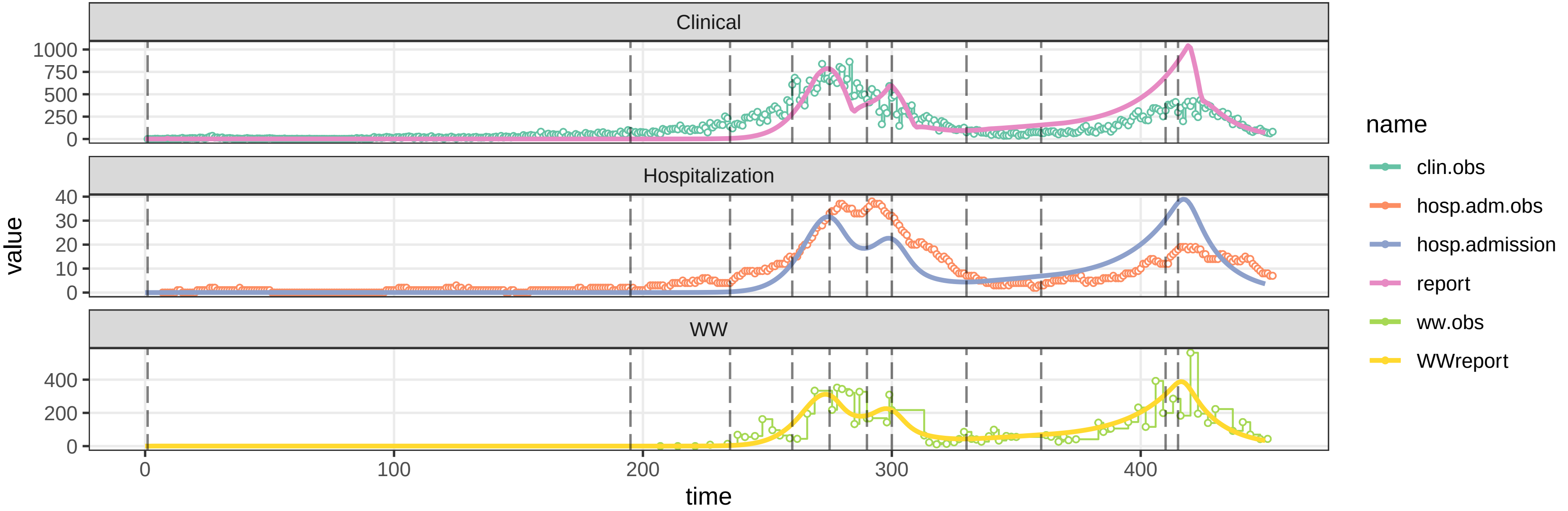


parameters/prm−EGB/prm−interv.csv

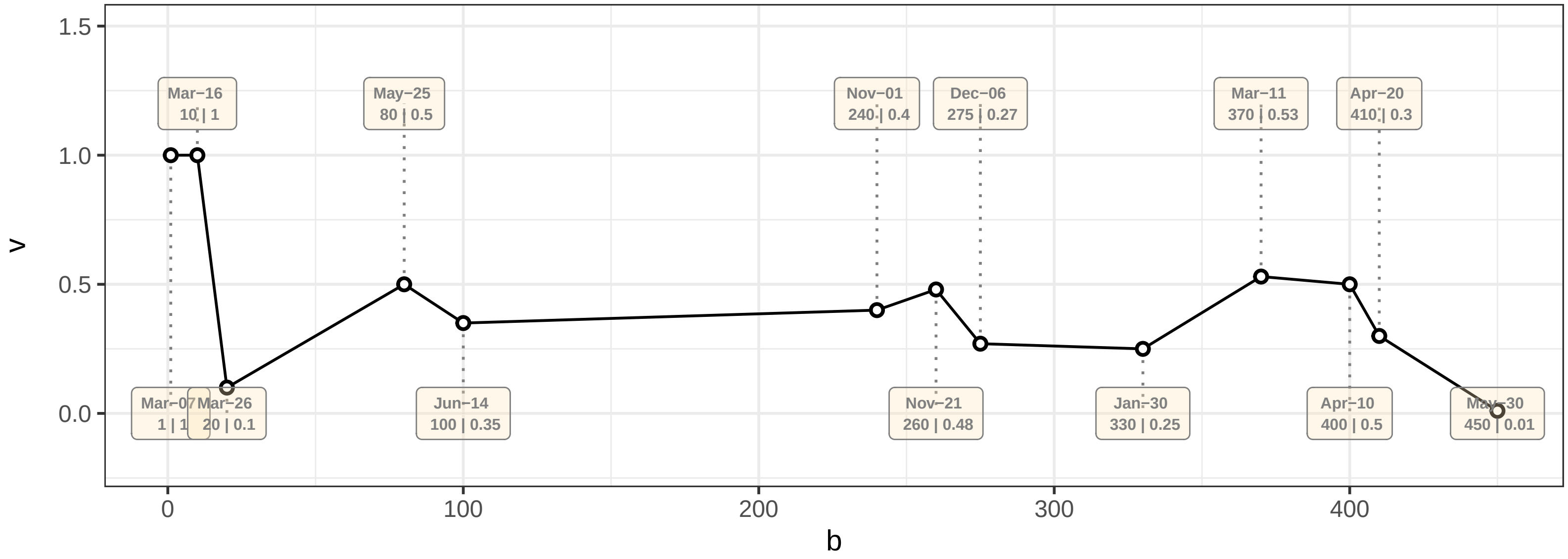

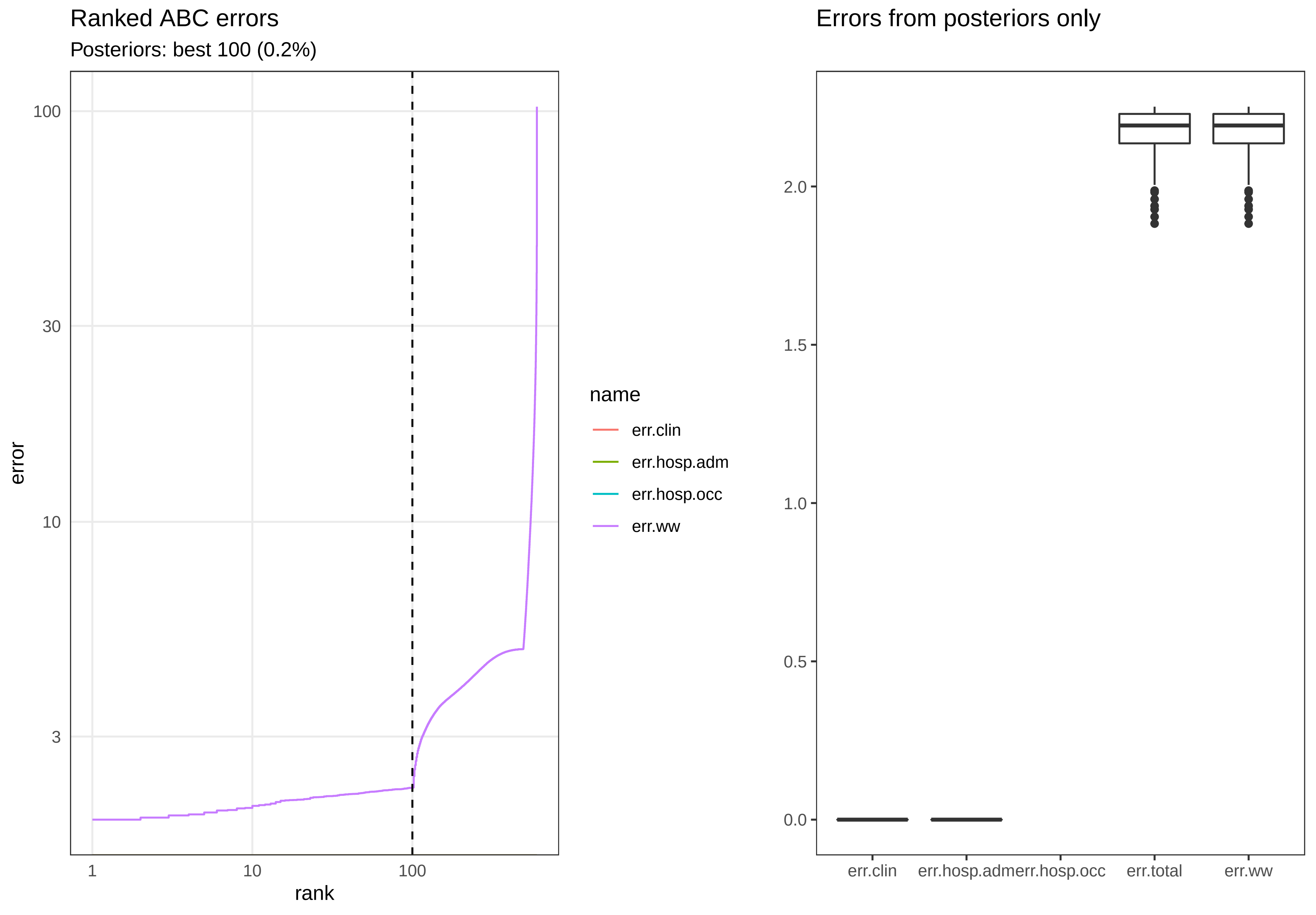

Ranked ABC errors
Posteriors: best 100 (0.2%)
error
100
30
10
3
1
10
100
rank
name
err.clin
err.hosp.adm
err.hosp.occ
err.ww
Errors from posteriors only
2.0
1.5
1.0
0.5
0.0
err.clin
err.hosp.adm
err.hosp.occ
err.total
err.ww

Posterior distributions

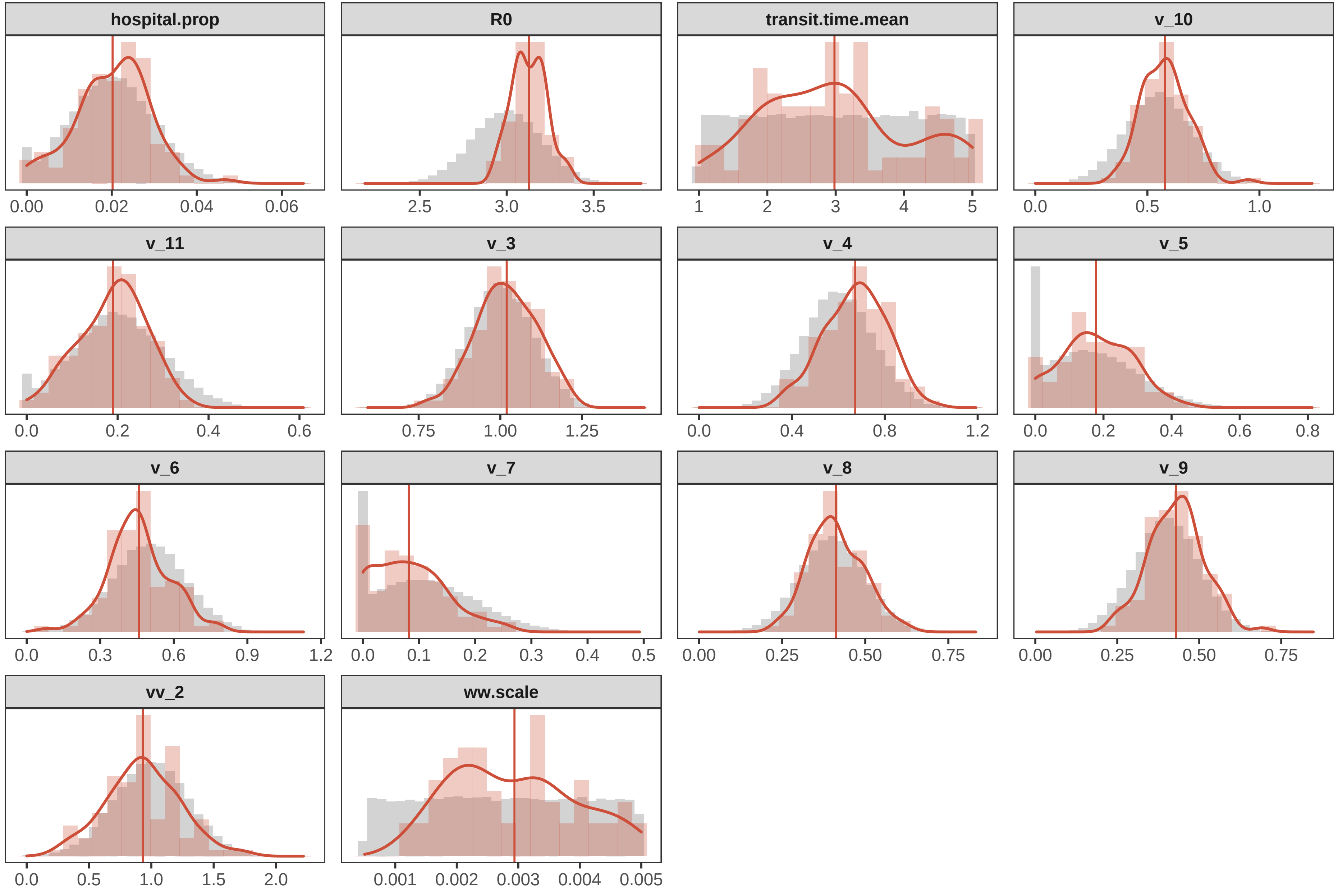

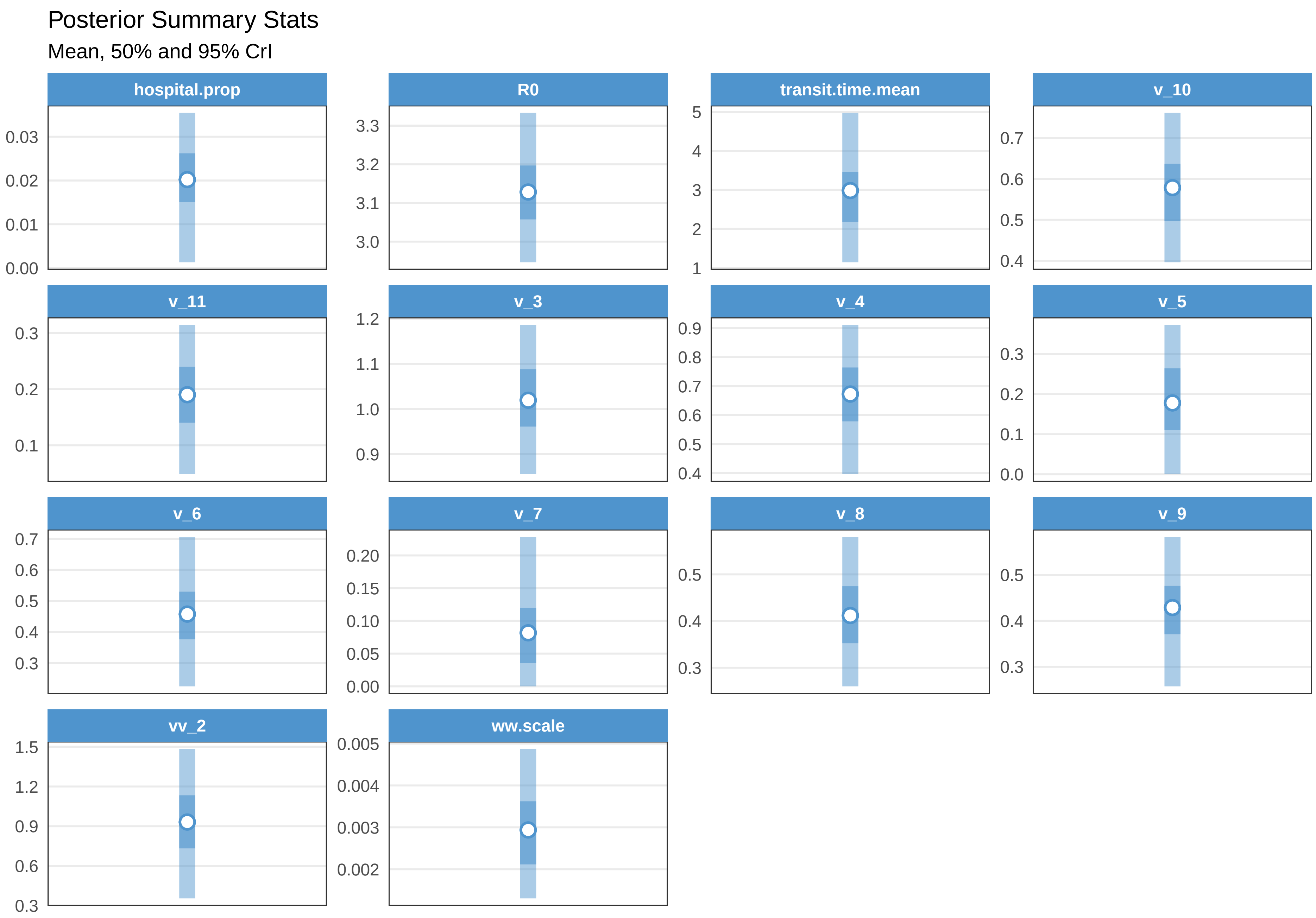
Posterior Summary Stats
Mean, 50% and 95% CrI
hospital.prop
0.03
0.02
0.01
0.00
R0
3.3
3.2
3.1
3.0
transit.time.mean
5
4
3
2
1
v_10
0.7
0.6
0.5
0.4
v_11
0.3
0.2
0.1
v_3
1.2
1.1
1.0
0.9
v_4
0.9
0.8
0.7
0.6
0.5
0.4
v_5
0.3
0.2
0.1
0.0
v_6
0.7
0.6
0.5
0.4
0.3
v_7
0.20
0.15
0.10
0.05
0.00
v_8
0.5
0.4
0.3
v_9
0.5
0.4
0.3
vv_2
1.5
1.2
0.9
0.6
0.3
ww.scale
0.005
0.004
0.003
0.002

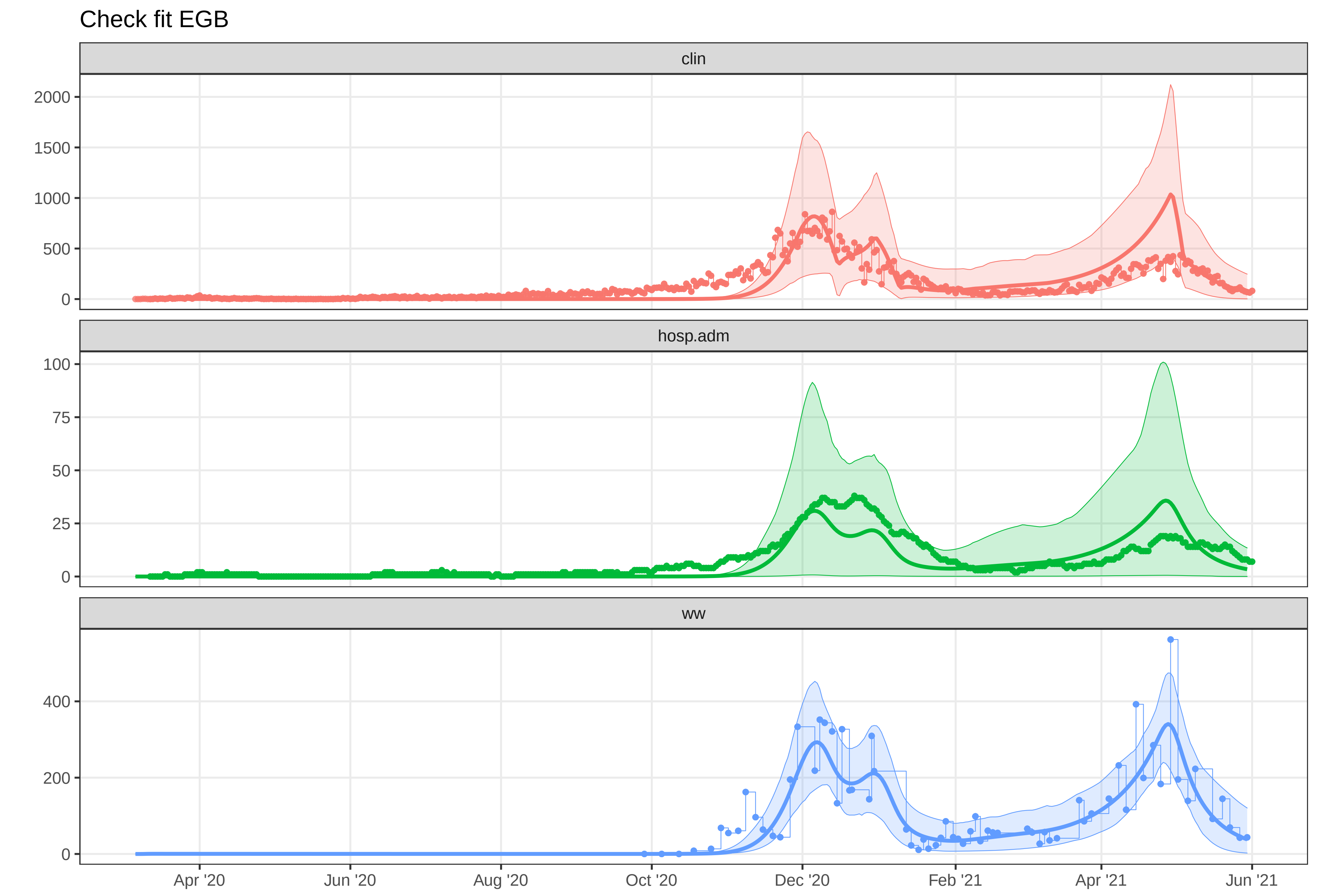

Check fit EGB
clin
2000
1500
1000
500
0
hosp.adm
100
75
50
25
0
ww
400
200
0
Apr '20
Jun '20
Aug '20
Oct '20
Dec '20
Feb '21
Apr '21
Jun '21
