## Supplementary material for "A Wastewater-Based Epidemic Model for SARS-CoV-2 with Application to Three Canadian Cities": fit outputs: plot-fit-EGB-07-09-0804-clin.pdf

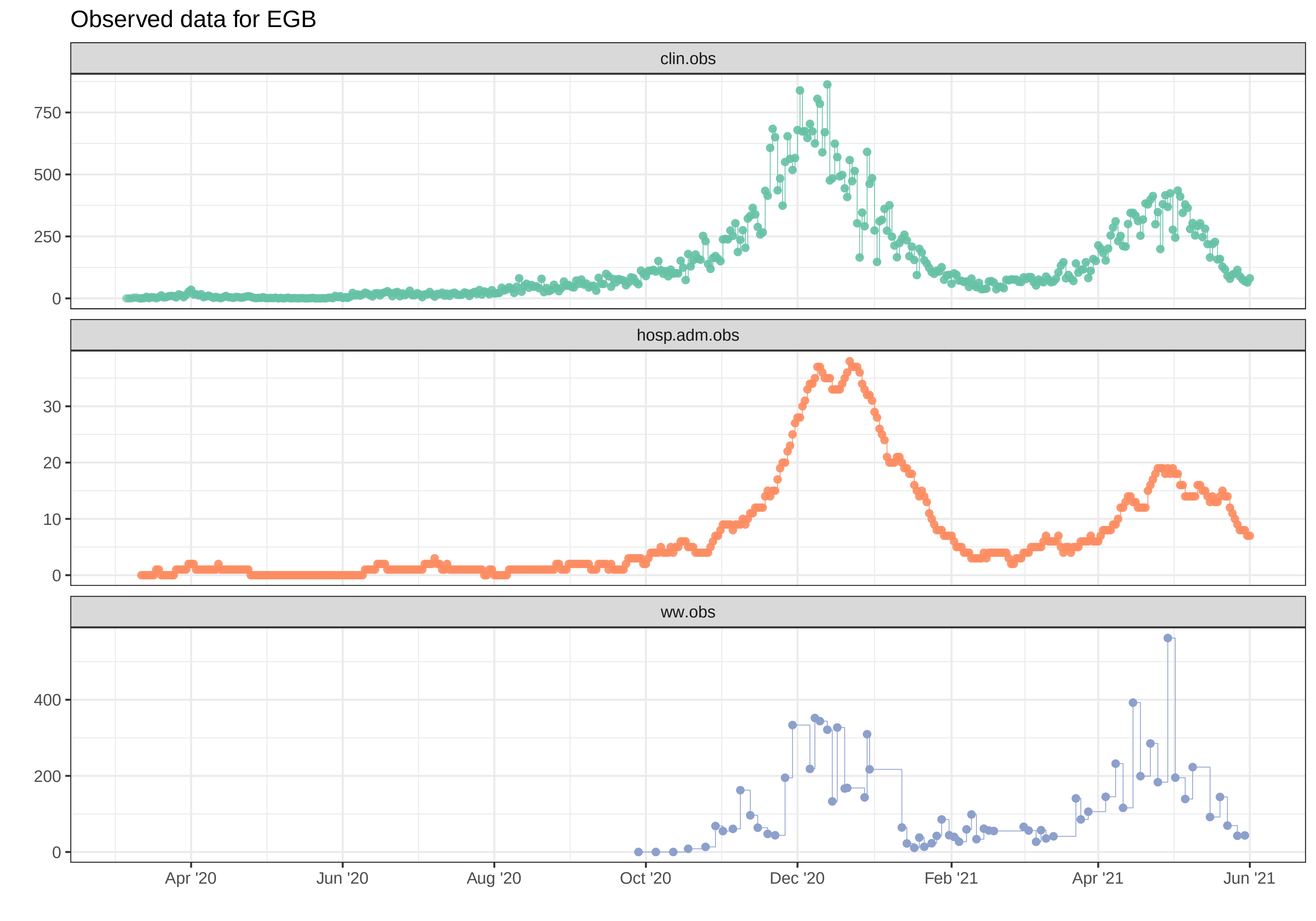

Observed data for EGB
clin.obs
750
500
250
0
hosp.adm.obs
30
20
10
0
ww.obs
400
200
0
Apr '20
Jun '20
Aug '20
Oct '20
Dec '20
Feb '21
Apr '21
Jun '21

#### Simulation with initial parameters for EGB

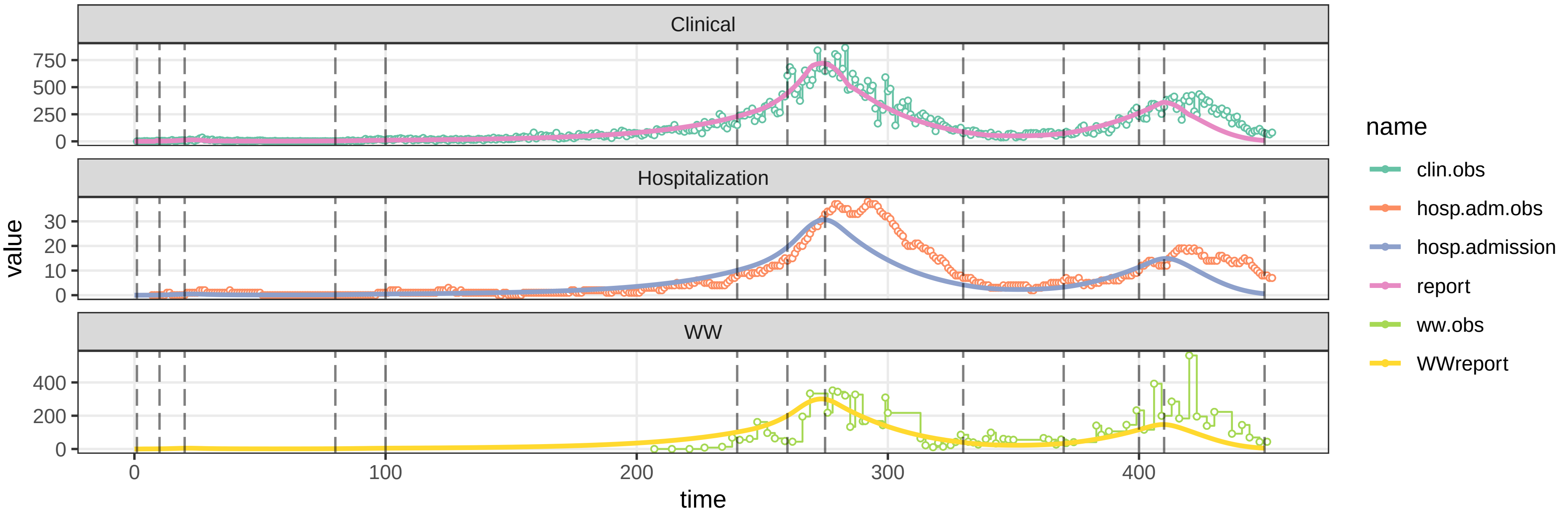


#### parameters/prm−EGB/prm−interv.csv

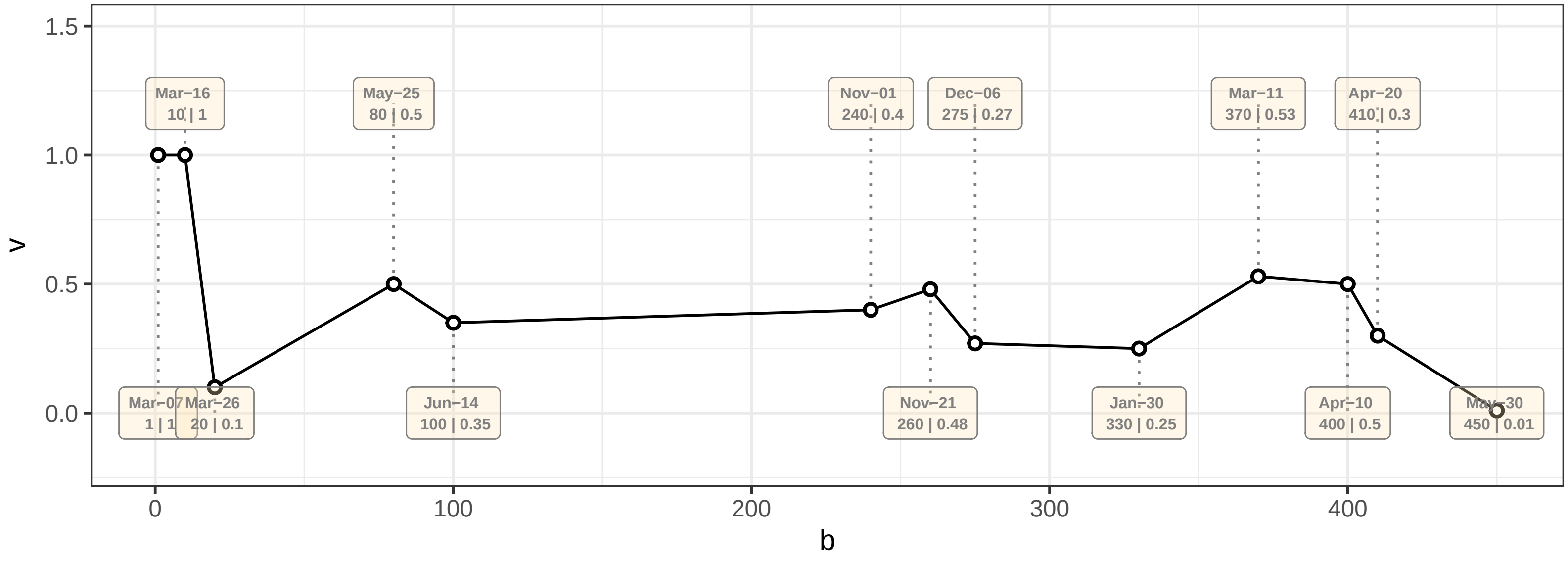

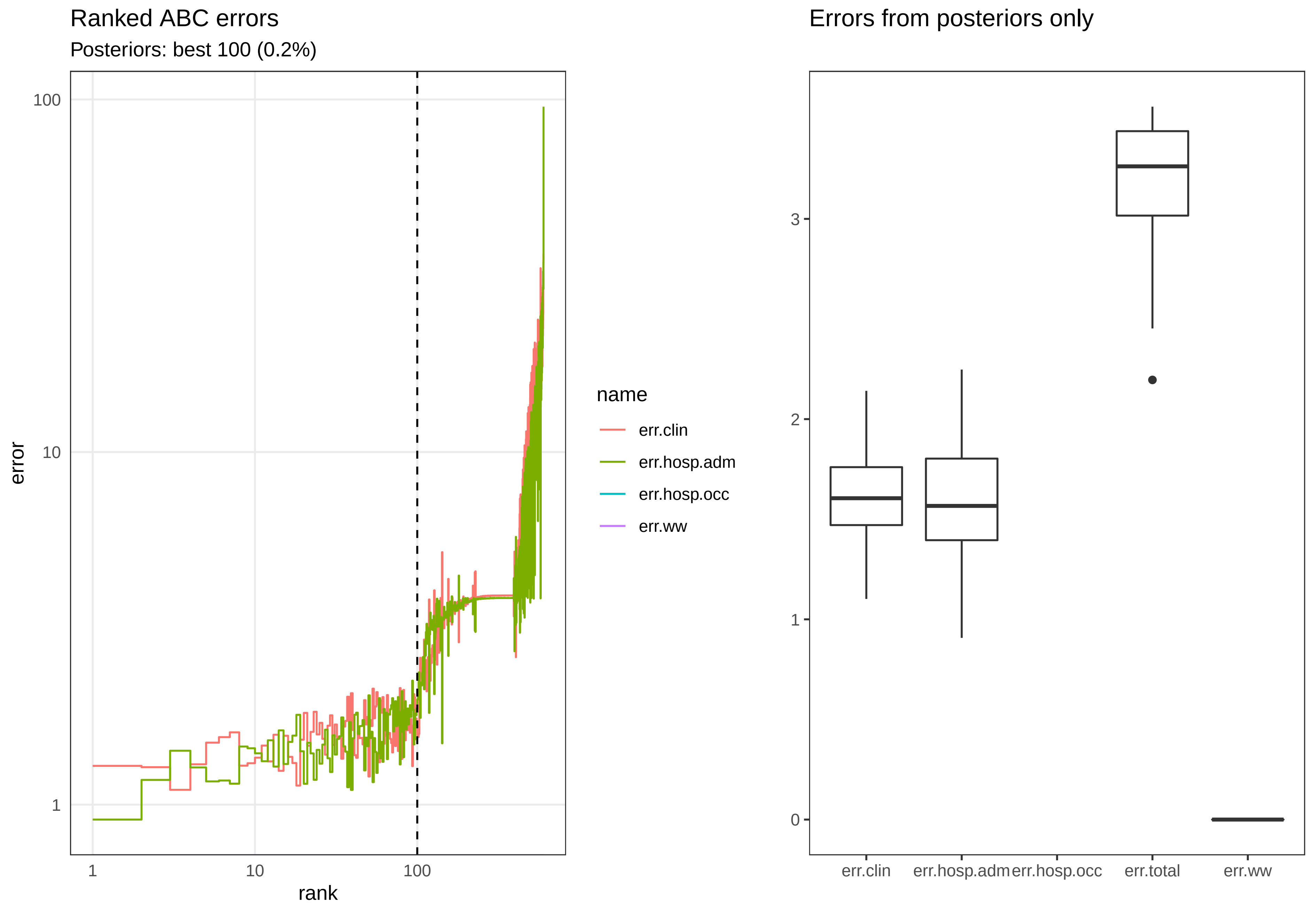

Ranked ABC errors
Posteriors: best 100 (0.2%)
error
100
10
1
1
10
100
rank
name
err.clin
err.hosp.adm
err.hosp.occ
err.ww
Errors from posteriors only
3
2
1
0
err.clin
err.hosp.adm
err.hosp.occ
err.total
err.ww

### Posterior distributions

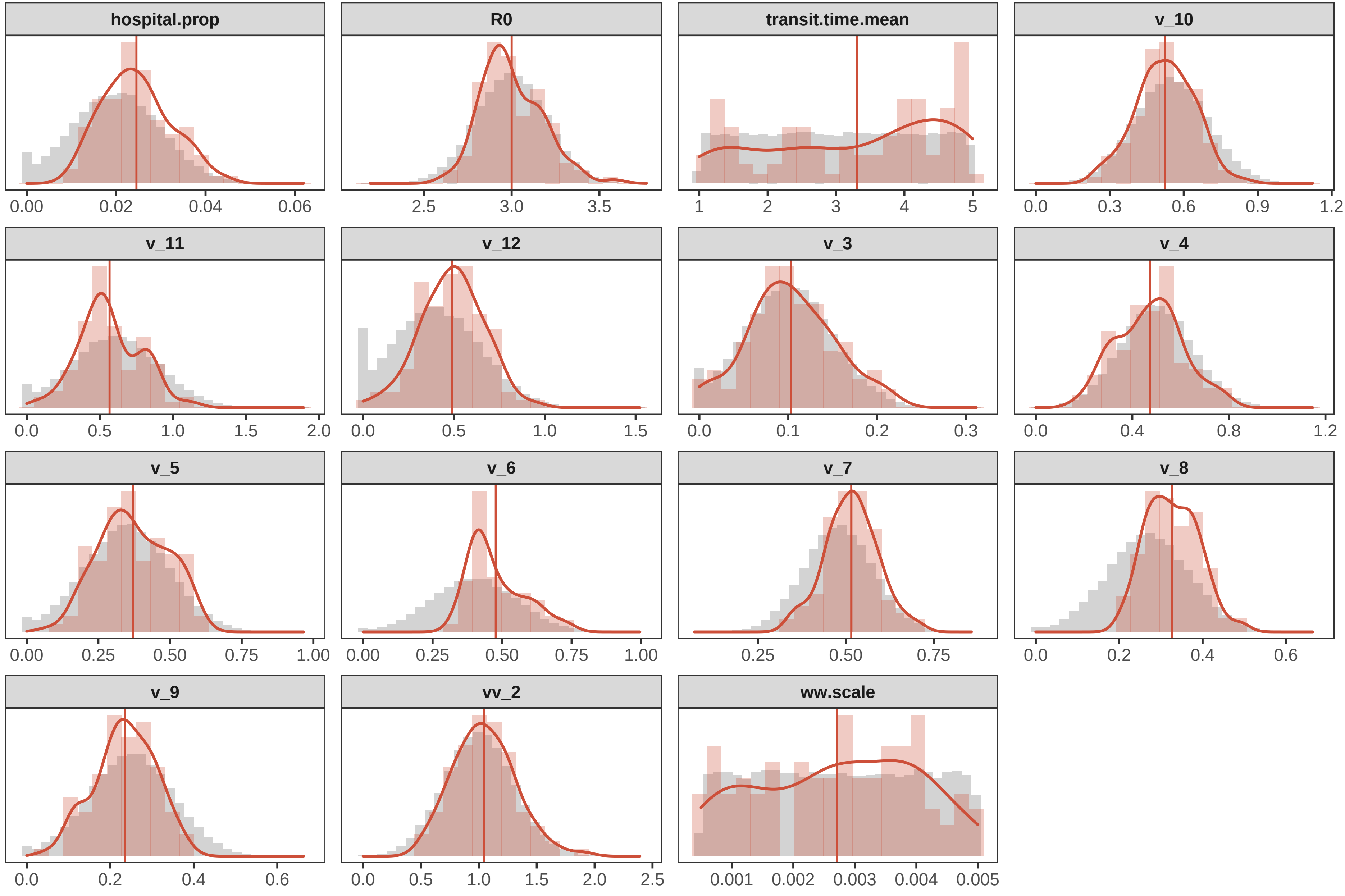

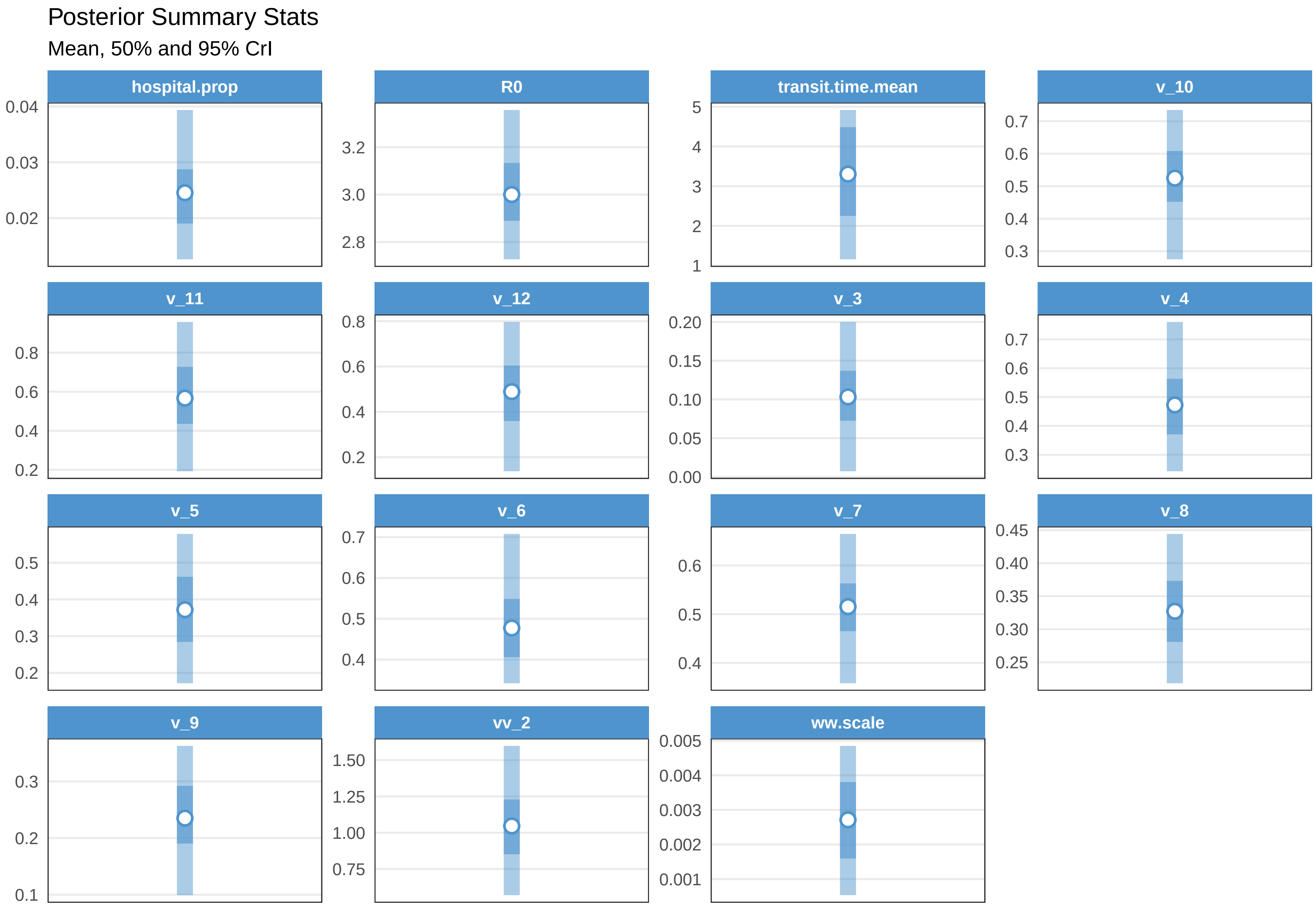

Posterior Summary Stats
Mean, 50% and 95% CrI
hospital.prop
R0
transit.time.mean
v_10
v_11
v_12
v_3
v_4
v_5
v_6
v_7
v_8
v_9
vv_2
ww.scale
0.04
0.03
0.02
3.2
3.0
2.8
5
4
3
2
1
0.7
0.6
0.5
0.4
0.3
0.8
0.6
0.4
0.2
0.8
0.6
0.4
0.2
0.20
0.15
0.10
0.05
0.00
0.7
0.6
0.5
0.4
0.3
0.5
0.4
0.3
0.2
0.7
0.6
0.5
0.4
0.6
0.5
0.4
0.45
0.40
0.35
0.30
0.25
0.3
0.2
0.1
1.50
1.25
1.00
0.75
0.005
0.004
0.003
0.002
0.001

Check fit EGB

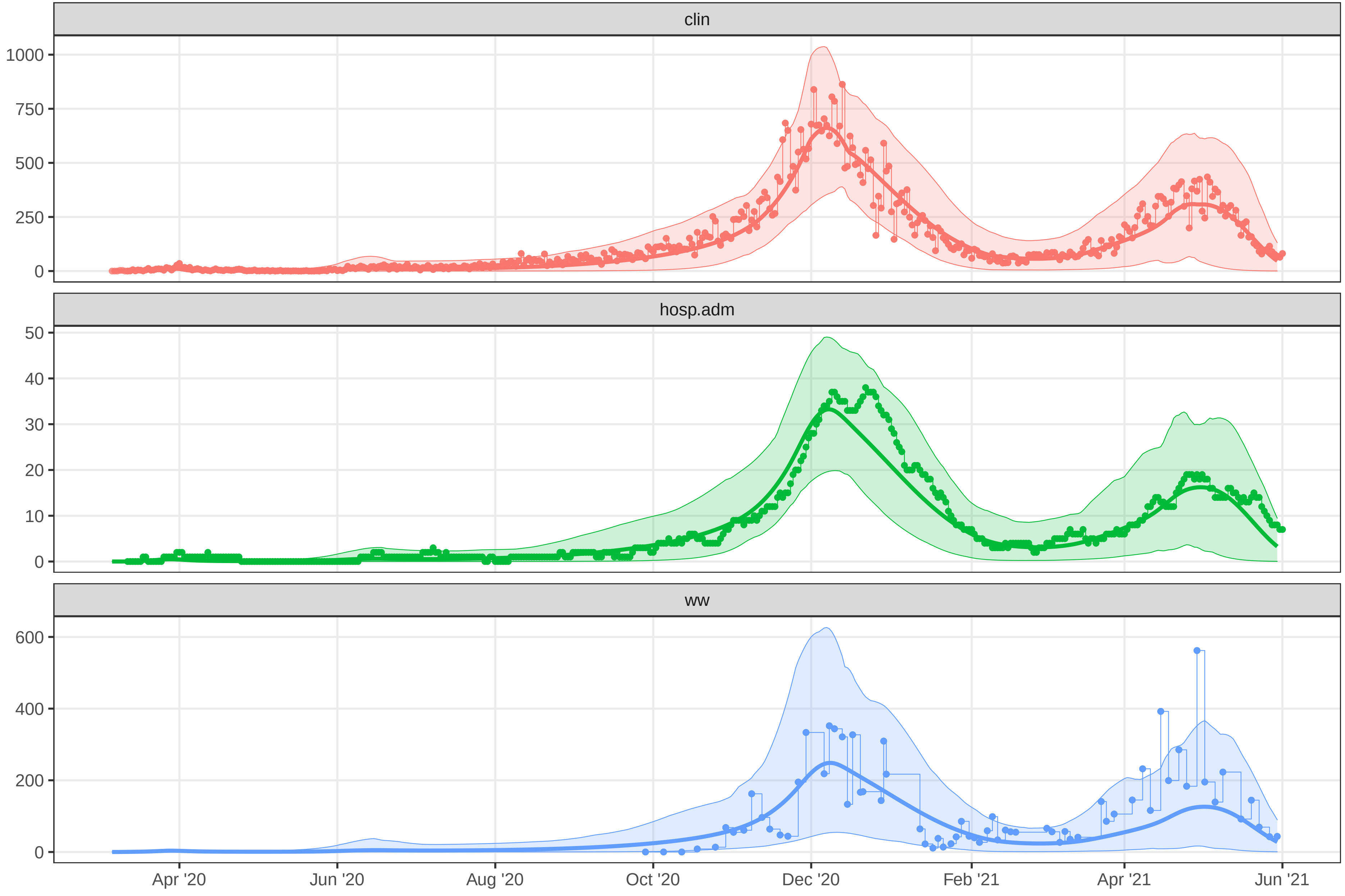
