## Supplementary material for "A Wastewater-Based Epidemic Model for SARS-CoV-2 with Application to Three Canadian Cities": fit outputs: plot-fit-EGB-07-09-0831-clinww.pdf

Observed data for EGB

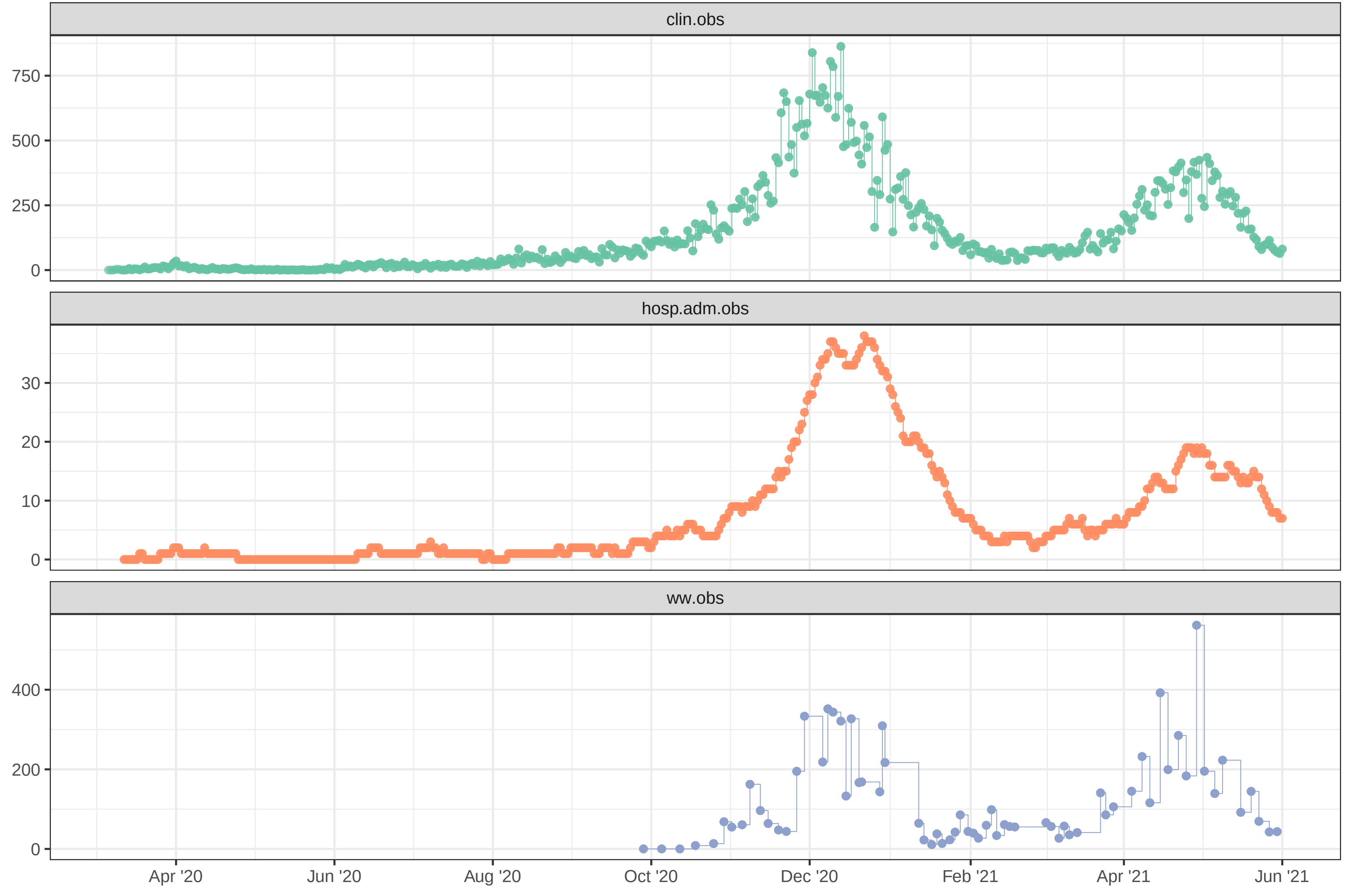

### Simulation with initial parameters for EGB

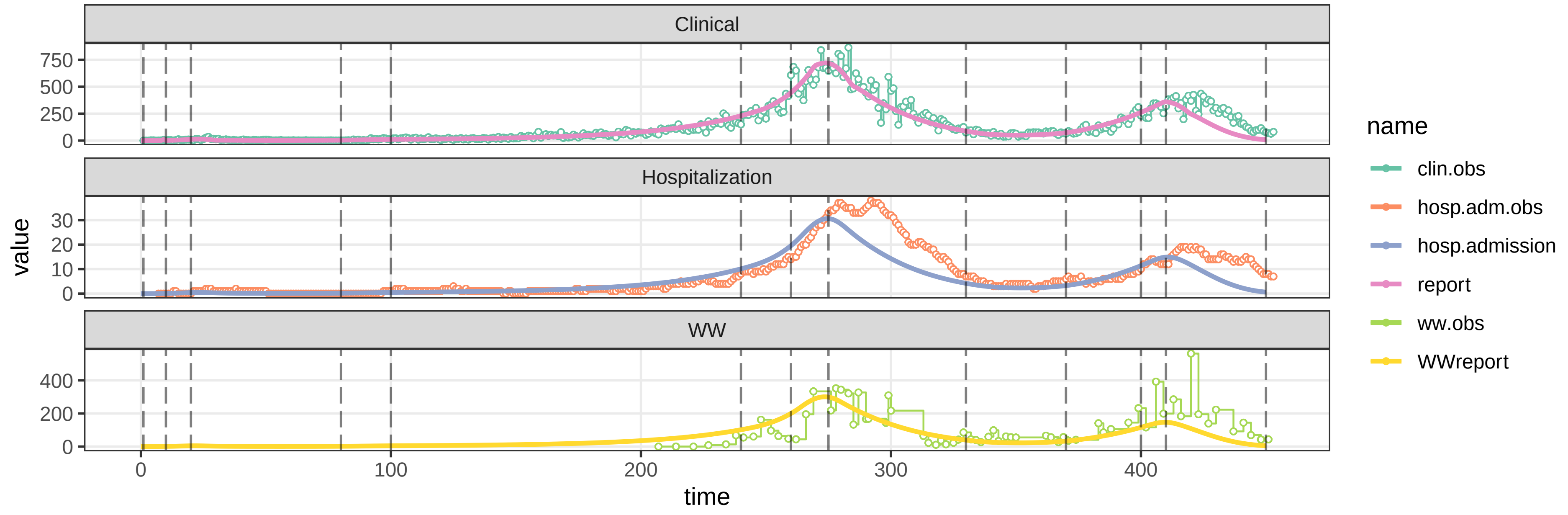


### parameters/prm−EGB/prm−interv.csv

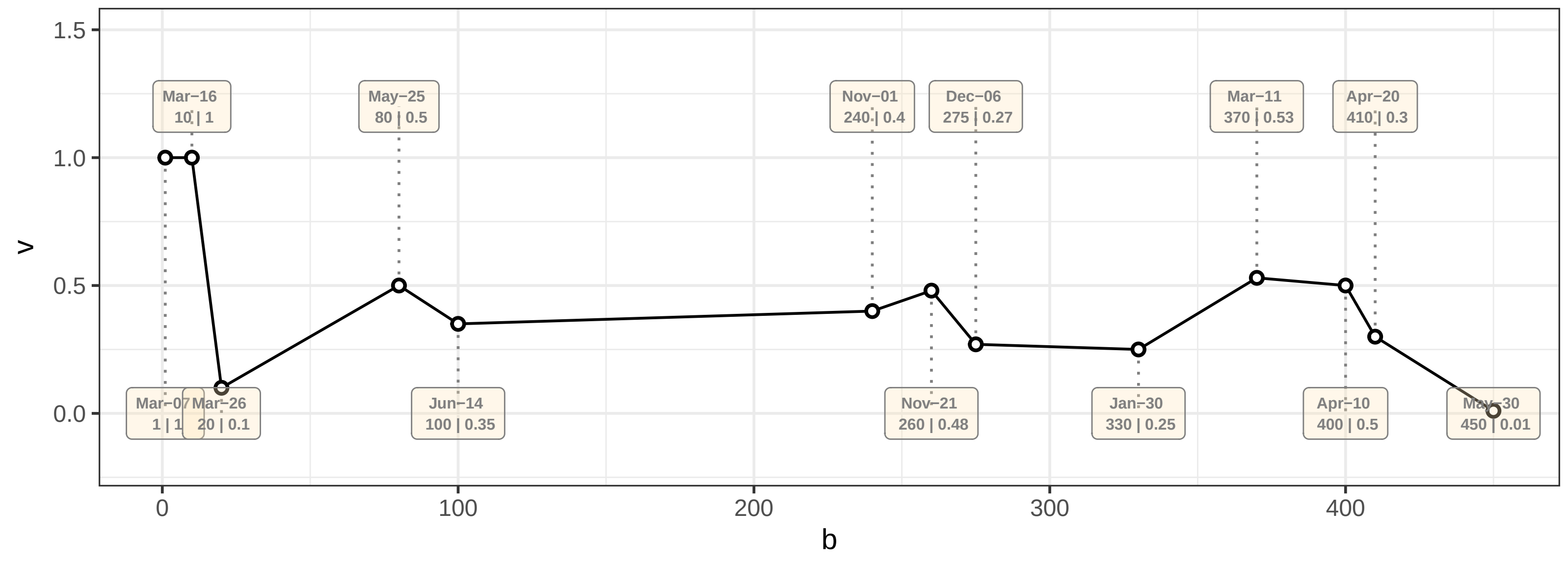

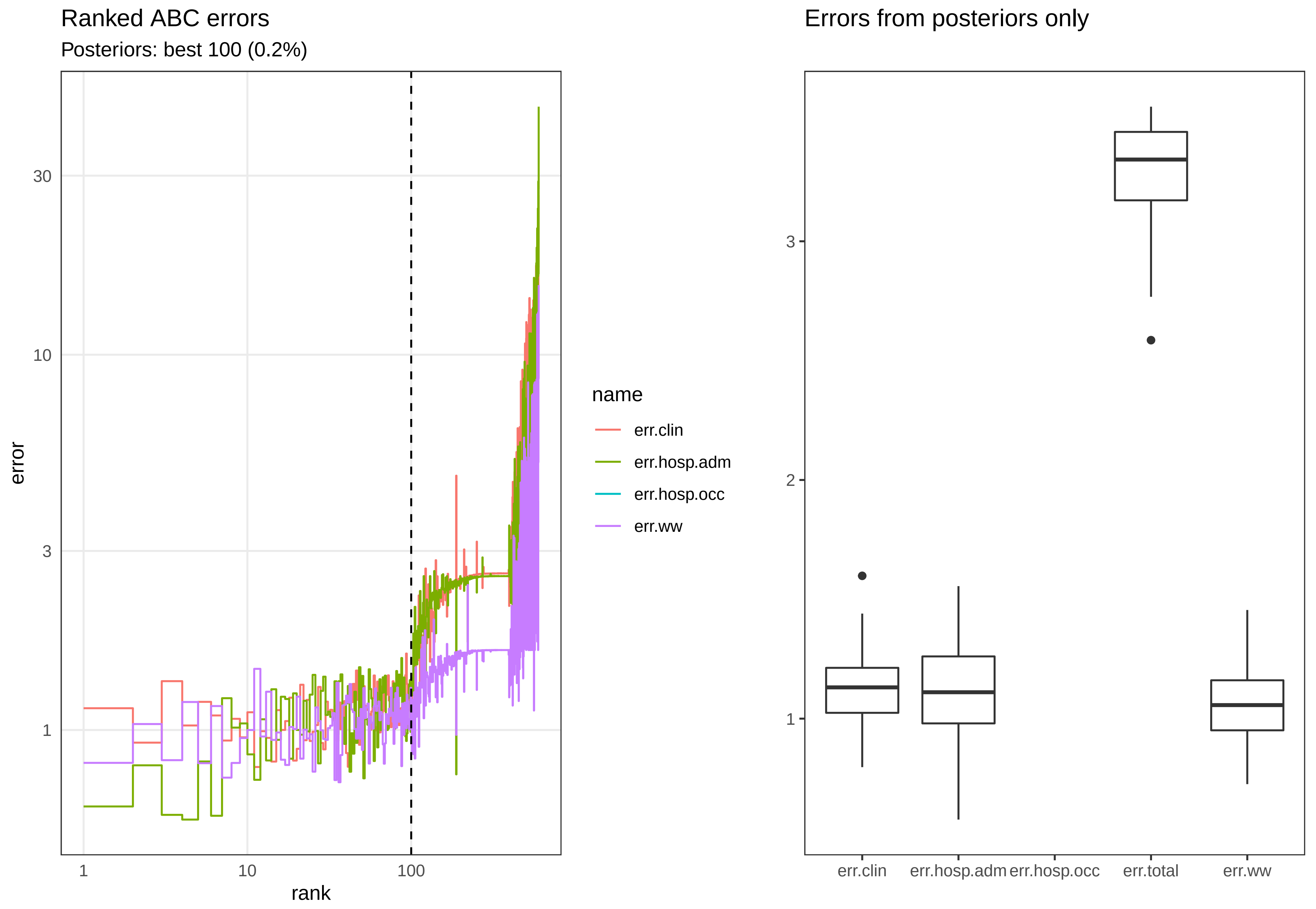
Ranked ABC errors
Posteriors: best 100 (0.2%)
error
30
10
3
1
1
10
100
rank
name
err.clin
err.hosp.adm
err.hosp.occ
err.ww
Errors from posteriors only
3
2
1
err.clin
err.hosp.adm
err.hosp.occ
err.total
err.ww

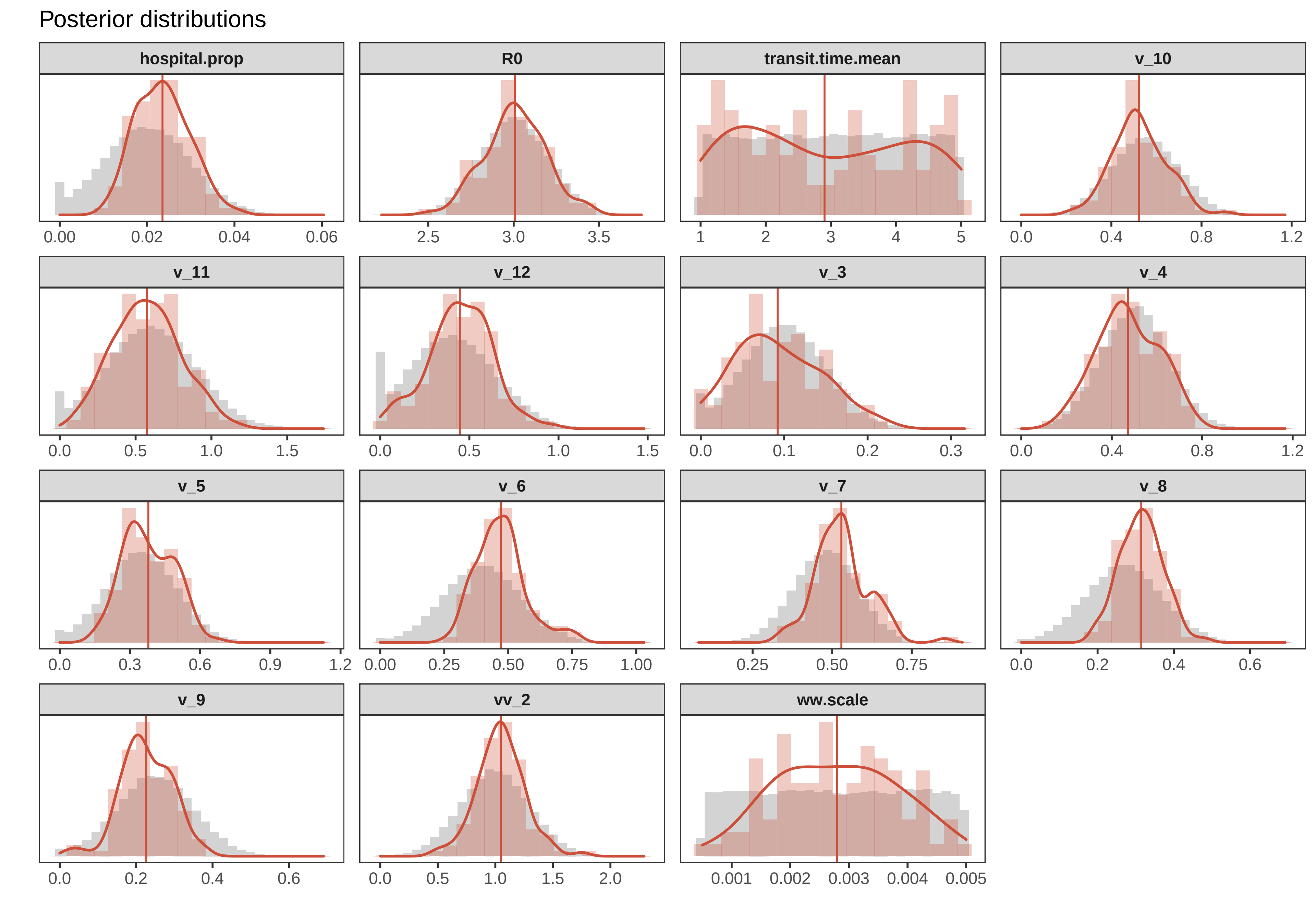

Posterior distributions
hospital.prop
0.00
0.02
0.04
0.06
R0
2.5
3.0
3.5
transit.time.mean
1
2
3
4
5
v_10
0.0
0.4
0.8
1.2
v_11
0.0
0.5
1.0
1.5
v_12
0.0
0.5
1.0
1.5
v_3
0.0
0.1
0.2
0.3
v_4
0.0
0.4
0.8
1.2
v_5
0.0
0.3
0.6
0.9
1.2
v_6
0.00
0.25
0.50
0.75
1.00
v_7
0.25
0.50
0.75
v_8
0.0
0.2
0.4
0.6
v_9
0.0
0.2
0.4
0.6
vv_2
0.0
0.5
1.0
1.5
2.0
ww.scale
0.001
0.002
0.003
0.004
0.005

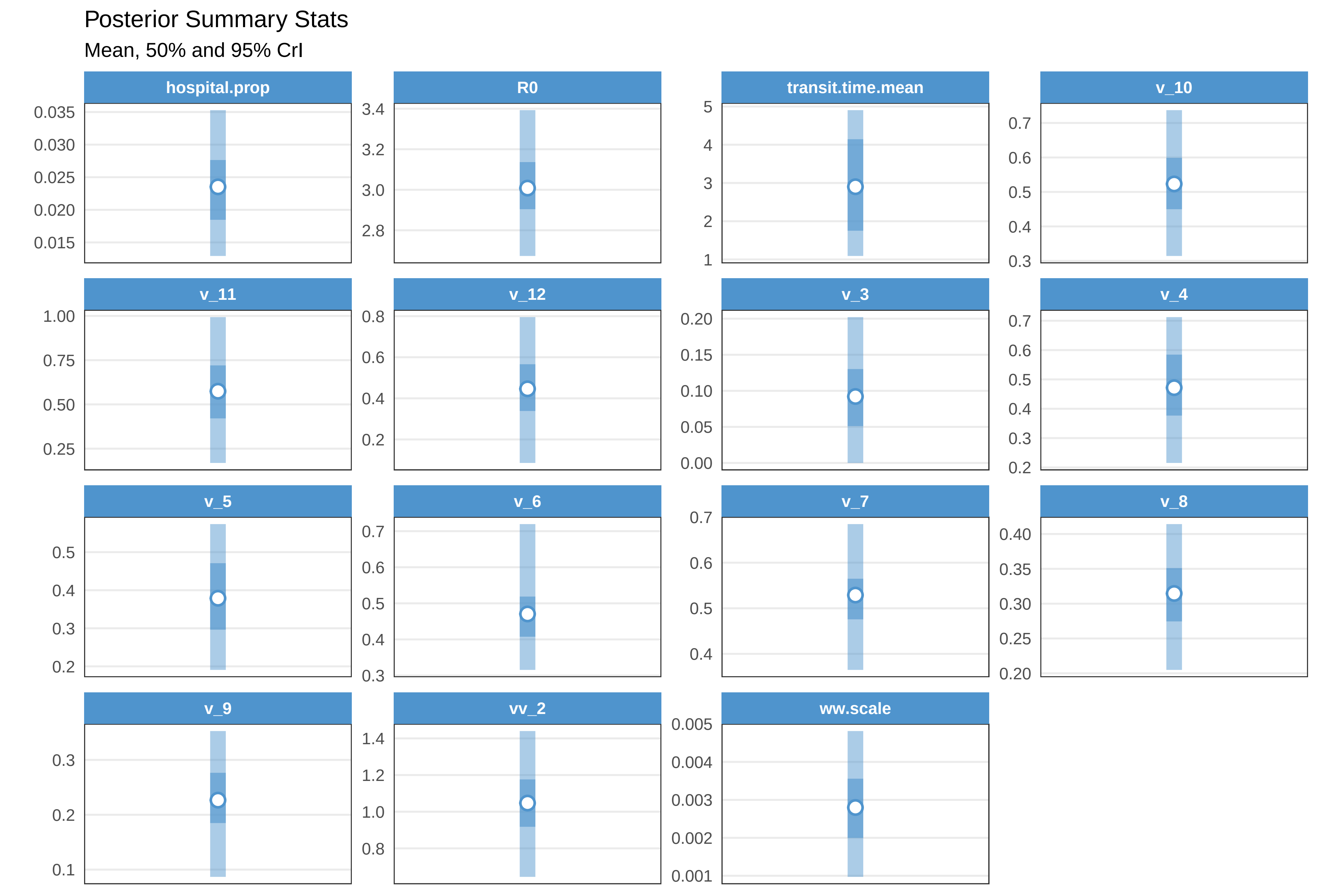

Posterior Summary Stats
Mean, 50% and 95% CrI
hospital.prop
0.035
0.030
0.025
0.020
0.015
R0
3.4
3.2
3.0
2.8
transit.time.mean
5
4
3
2
1
v_10
0.7
0.6
0.5
0.4
0.3
v_11
1.00
0.75
0.50
0.25
v_12
0.8
0.6
0.4
0.2
v_3
0.20
0.15
0.10
0.05
0.00
v_4
0.7
0.6
0.5
0.4
0.3
0.2
v_5
0.5
0.4
0.3
0.2
v_6
0.7
0.6
0.5
0.4
0.3
v_7
0.7
0.6
0.5
0.4
v_8
0.40
0.35
0.30
0.25
0.20
v_9
0.3
0.2
0.1
vv_2
1.4
1.2
1.0
0.8
ww.scale
0.005
0.004
0.003
0.002
0.001

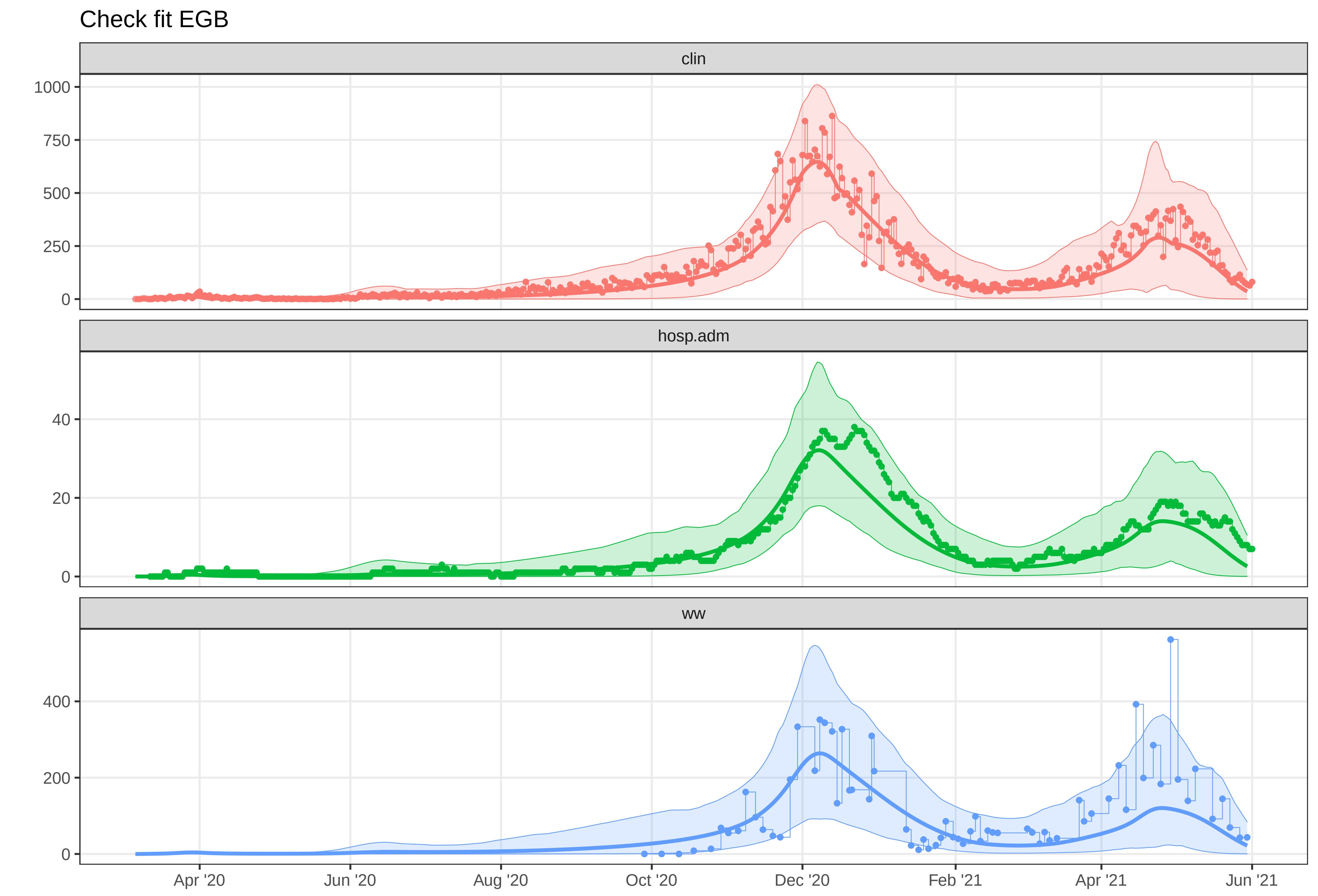
Check fit EGB
clin
hosp.adm
ww
1000
750
500
250
0
40
20
0
400
200
0
Apr '20
Jun '20
Aug '20
Oct '20
Dec '20
Feb '21
Apr '21
Jun '21
