## Supplementary material for "A Wastewater-Based Epidemic Model for SARS-CoV-2 with Application to Three Canadian Cities": fit outputs: plot-fit-OTW-07-09-0924-clin.pdf

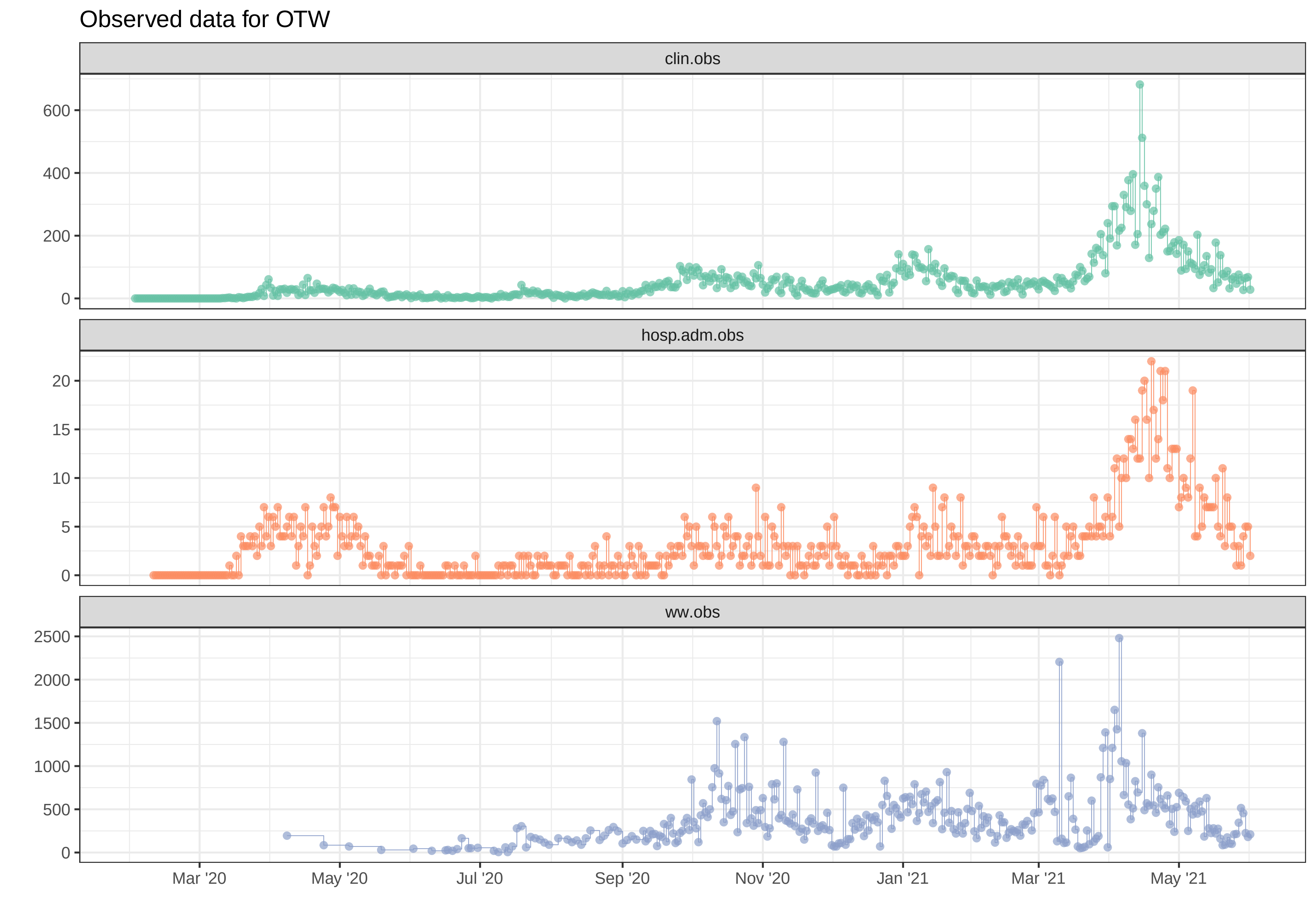

Observed data for OTW
clin.obs
0
200
400
600
hosp.adm.obs
0
5
10
15
20
ww.obs
0
500
1000
1500
2000
2500
Mar '20
May '20
Jul '20
Sep '20
Nov '20
Jan '21
Mar '21
May '21

Simulation with initial parameters for OTW

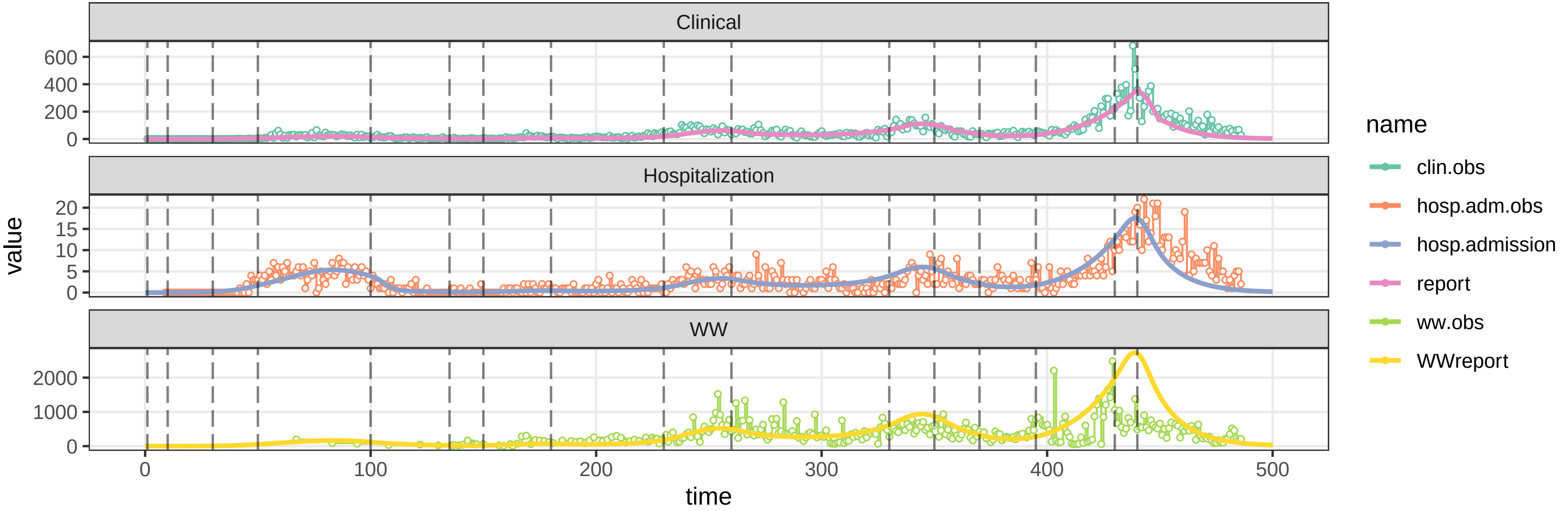


parameters/prm−OTW/prm−interv.csv

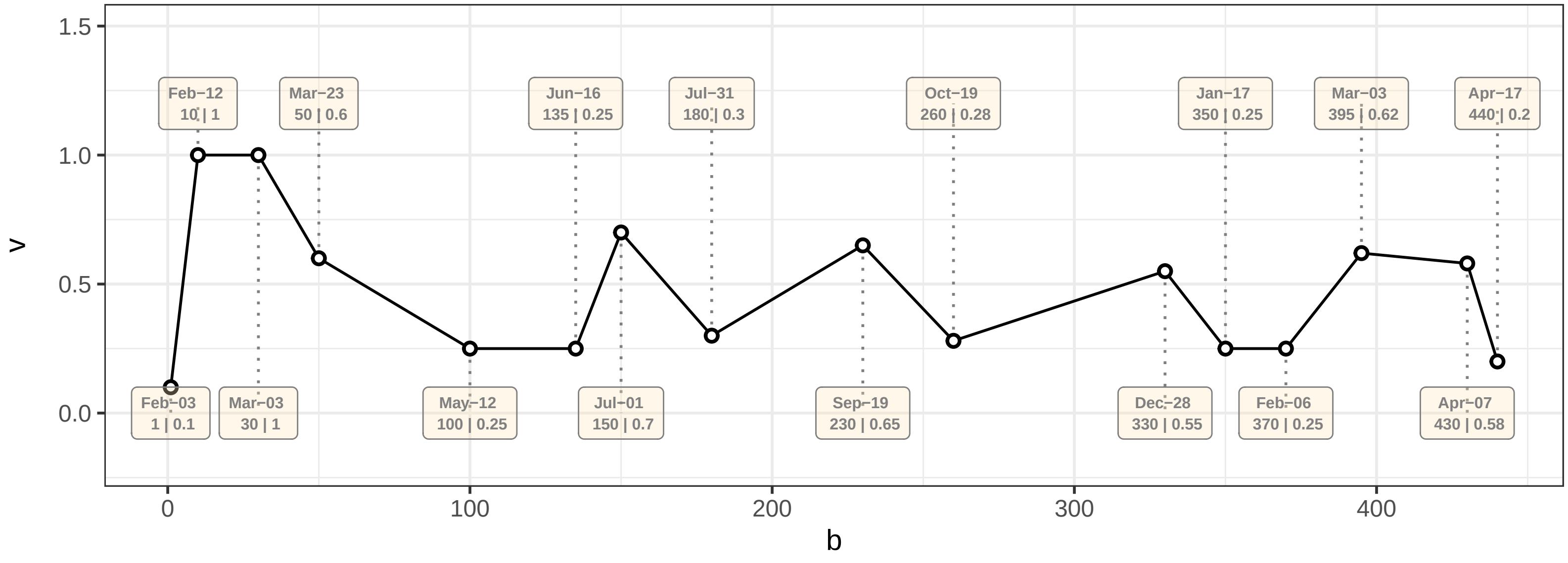

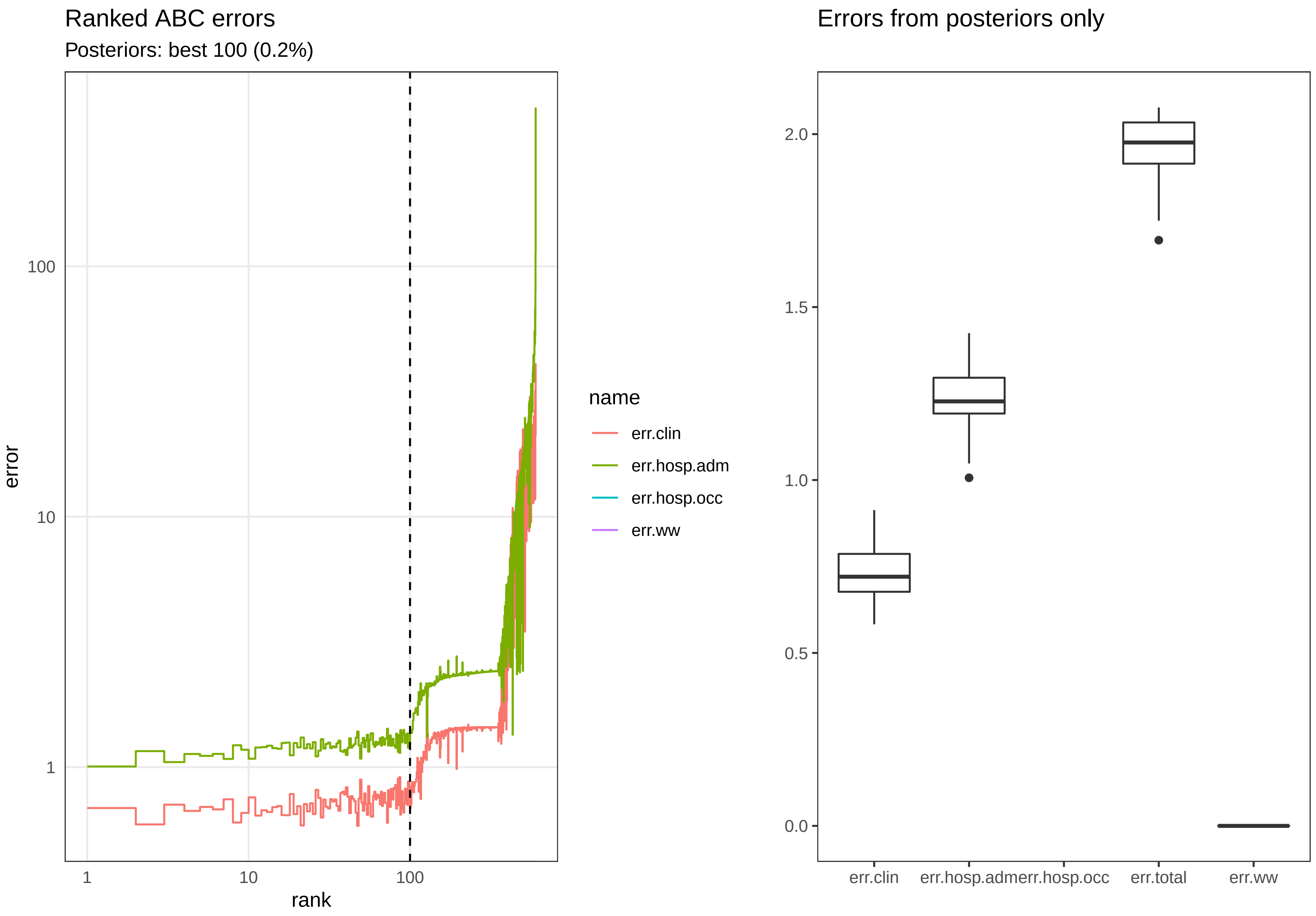

Ranked ABC errors
Posteriors: best 100 (0.2%)
error
100
10
1
1
10
100
rank
name
err.clin
err.hosp.adm
err.hosp.occ
err.ww
Errors from posteriors only
2.0
1.5
1.0
0.5
0.0
err.clin
err.hosp.adm
err.hosp.occ
err.total
err.ww

Posterior distributions

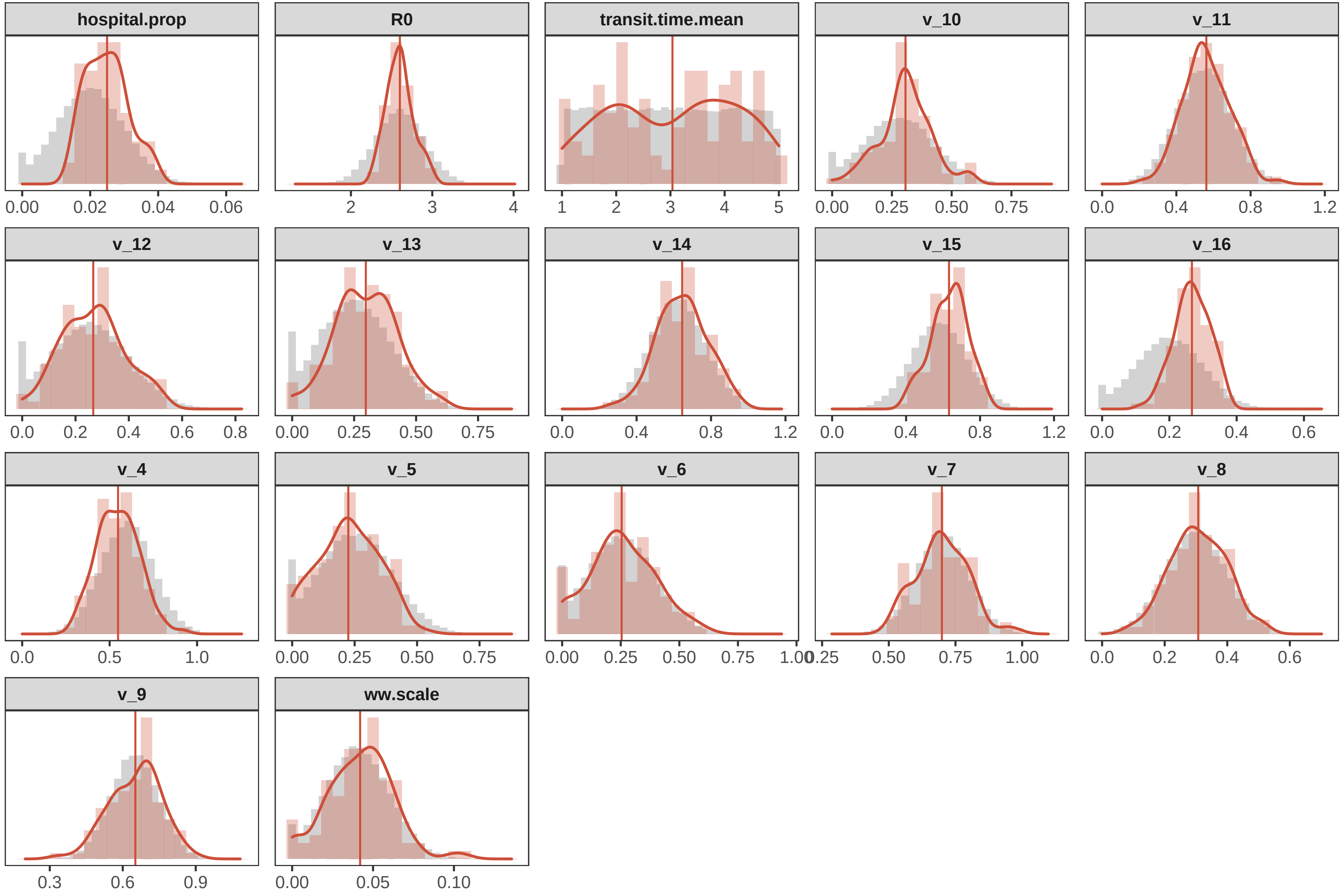

Posterior Summary Stats

Mean, 50% and 95% CrI

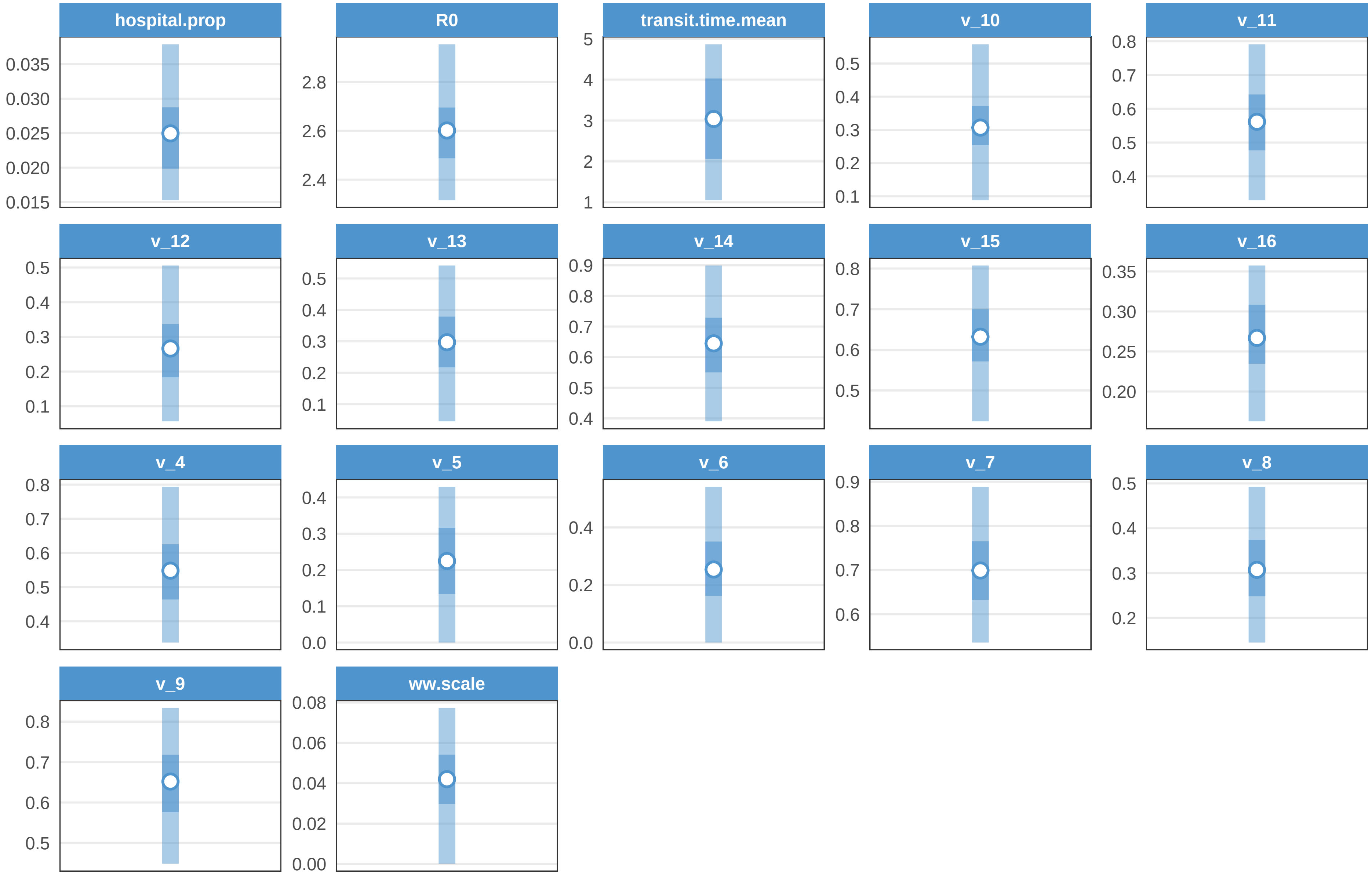

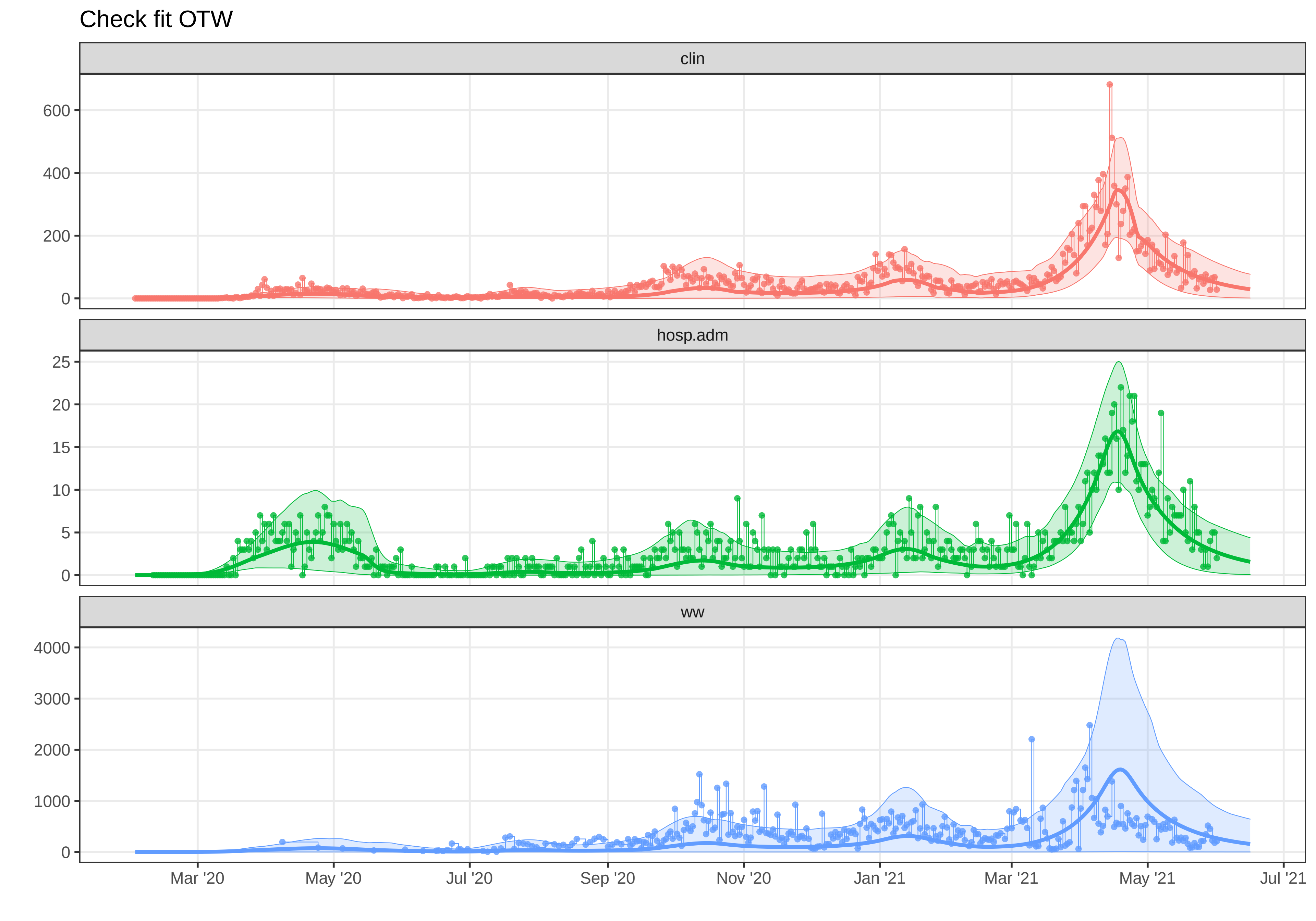

Check fit OTW
clin
600
400
200
0
hosp.adm
25
20
15
10
5
0
ww
4000
3000
2000
1000
0
Mar '20
May '20
Jul '20
Sep '20
Nov '20
Jan '21
Mar '21
May '21
Jul '21
