## Supplementary material for "A Wastewater-Based Epidemic Model for SARS-CoV-2 with Application to Three Canadian Cities": fit outputs: plot-fit-TAB-07-06-clin.pdf

### Observed data for TAB

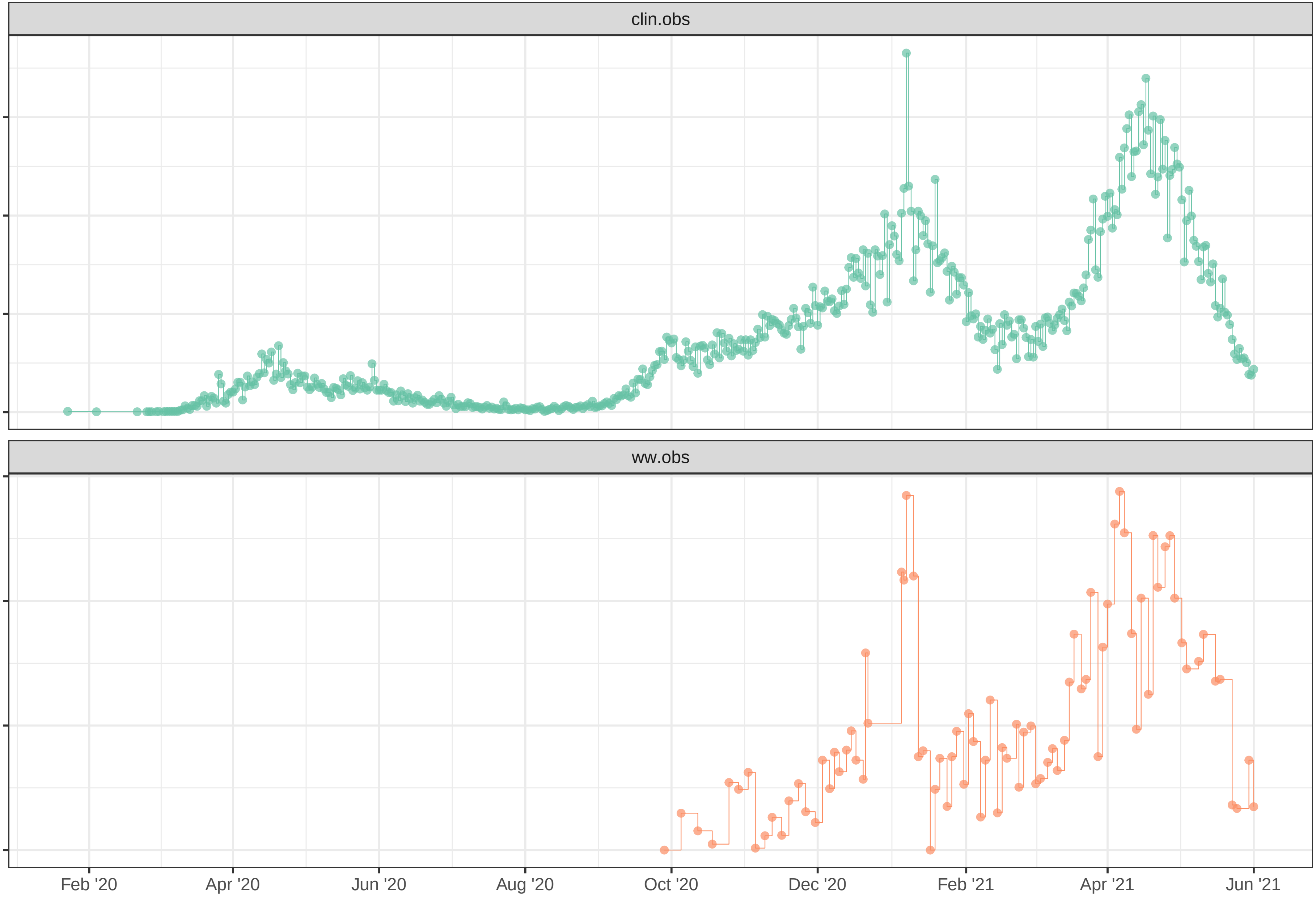

Simulation with initial parameters for TAB

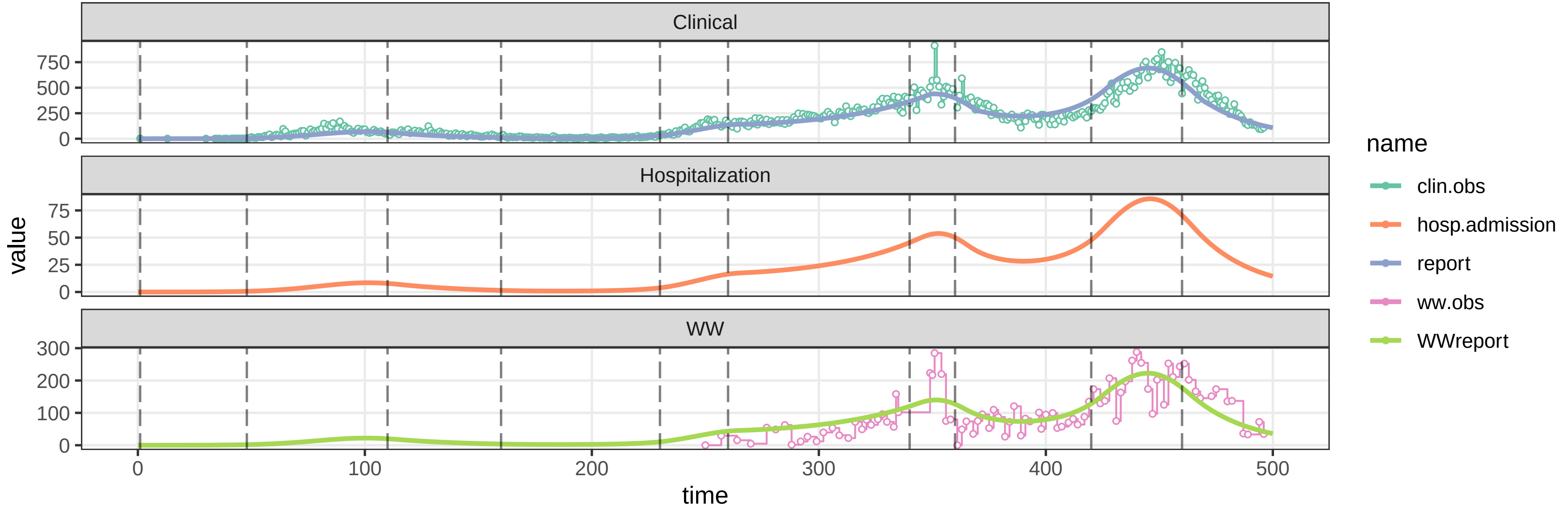


parameters/prm−TAB/prm−interv.csv

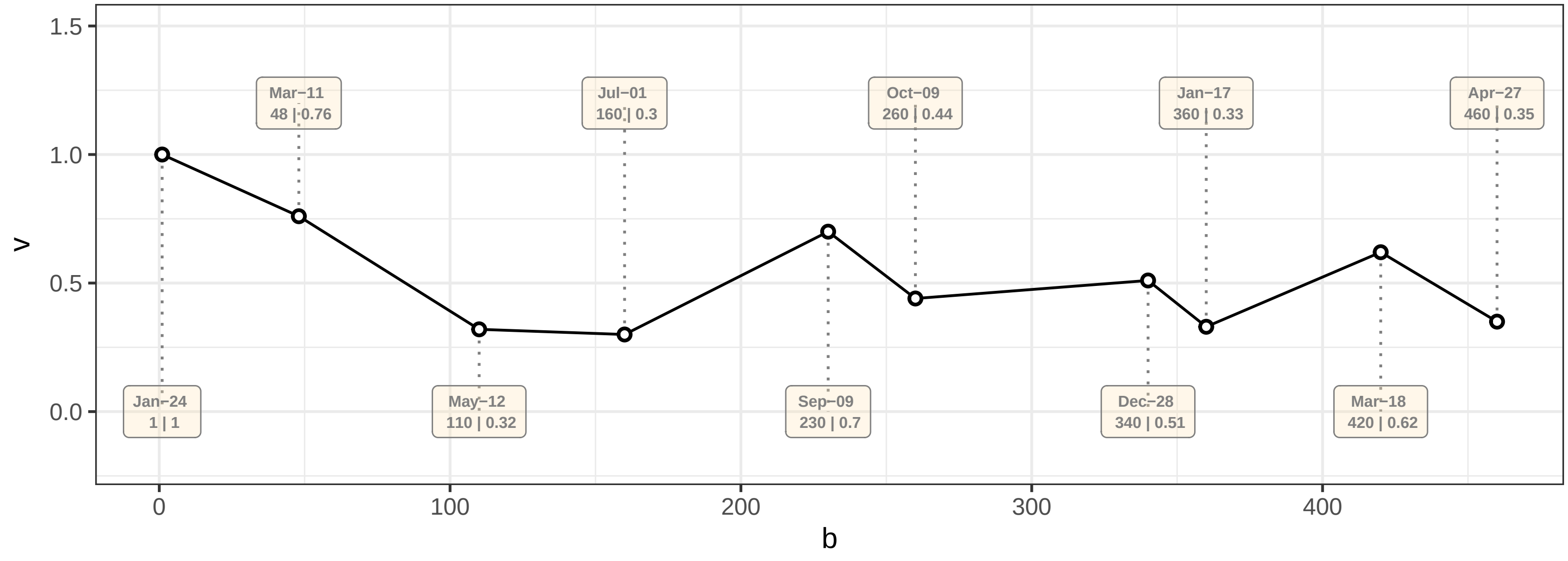

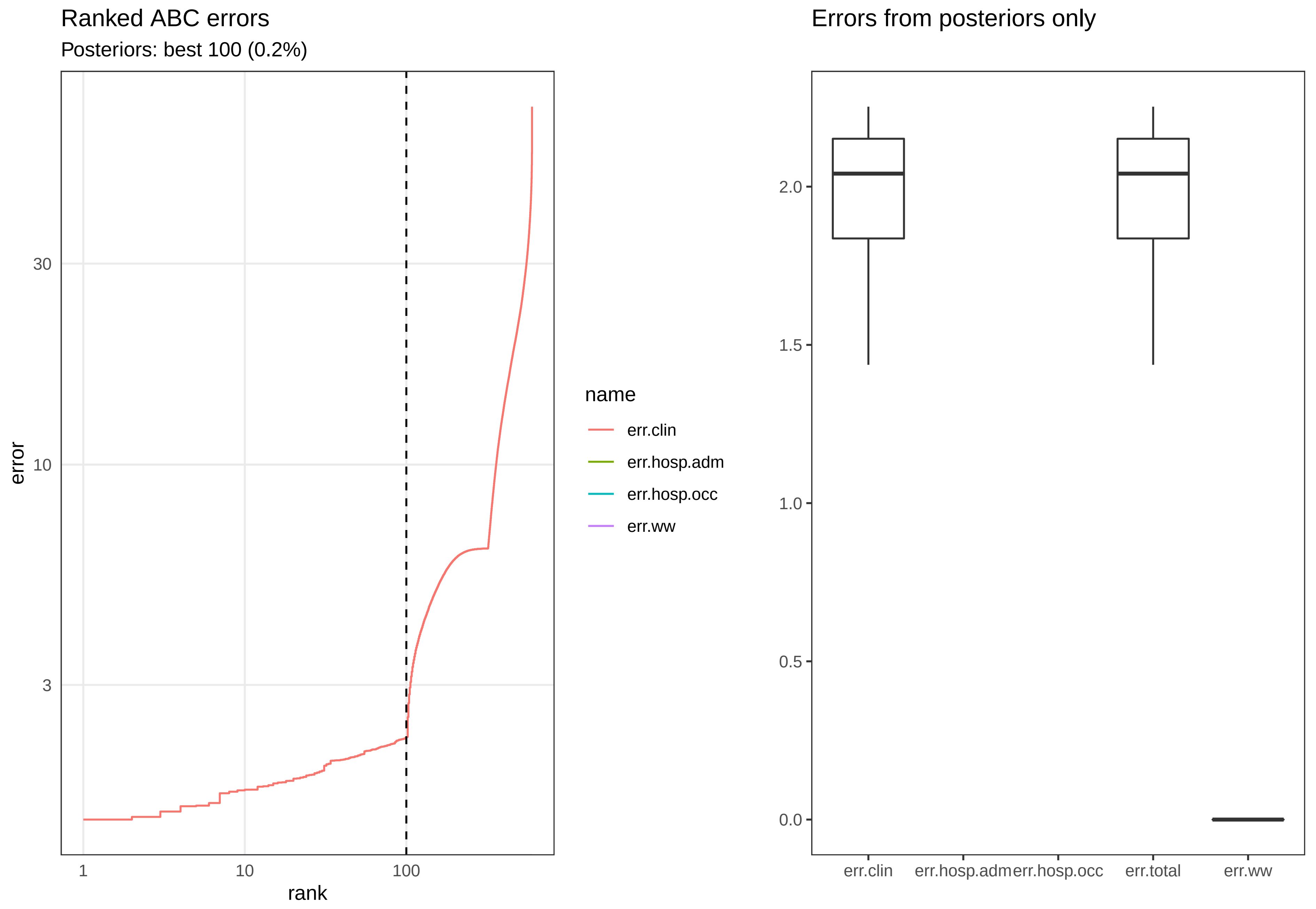
Ranked ABC errors
Posteriors: best 100 (0.2%)
error
30
10
3
1
10
100
rank
name
err.clin
err.hosp.adm
err.hosp.occ
err.ww
Errors from posteriors only
2.0
1.5
1.0
0.5
0.0
err.clin
err.hosp.adm
err.hosp.occ
err.total
err.ww

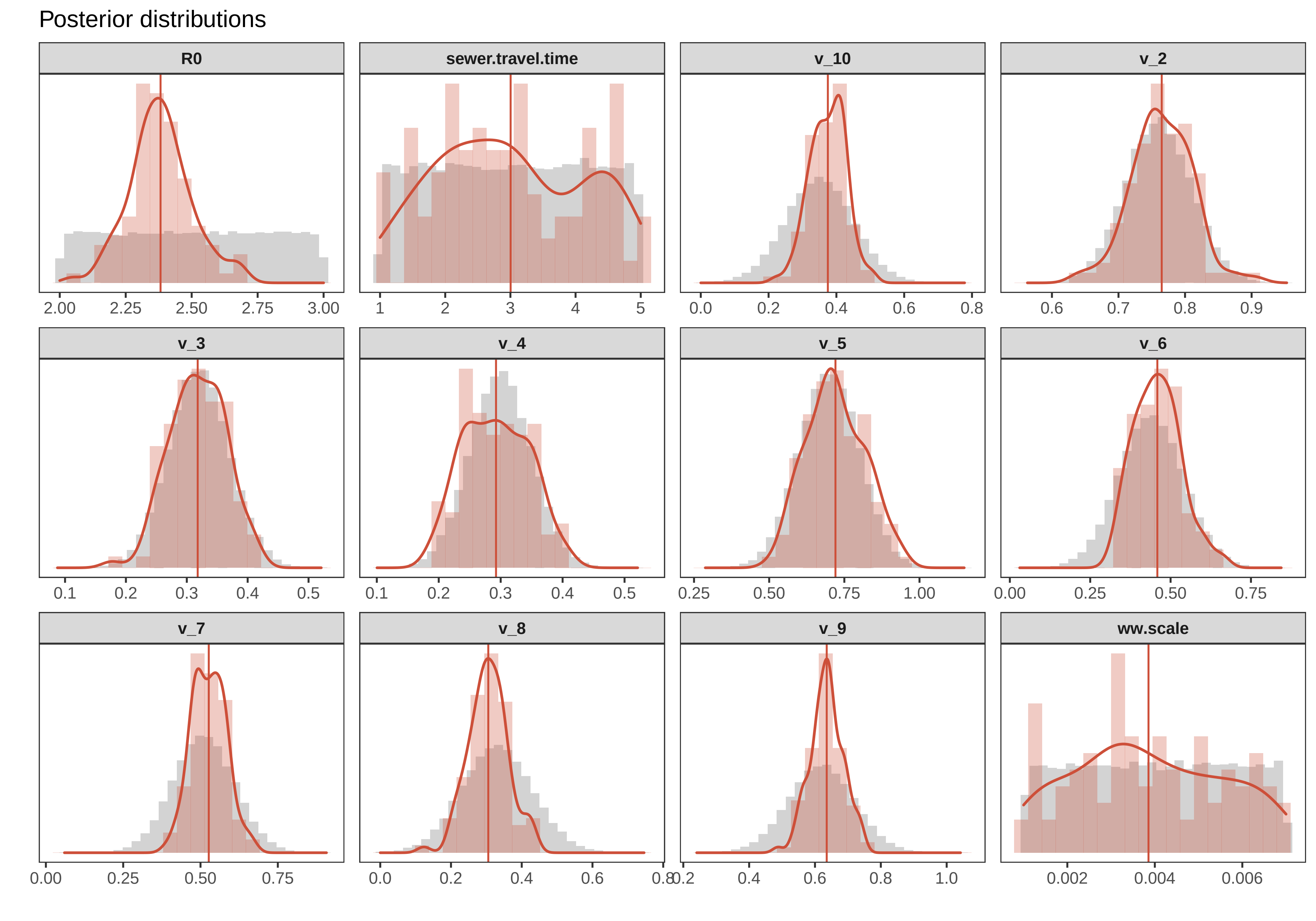
Posterior distributions
R0
sewer.travel.time
v_10
v_2
v_3
v_4
v_5
v_6
v_7
v_8
v_9
ww.scale
2.00 2.25 2.50 2.75 3.00
1 2 3 4 5
0.0 0.2 0.4 0.6 0.8
0.6 0.7 0.8 0.9
0.1 0.2 0.3 0.4 0.5
0.1 0.2 0.3 0.4 0.5
0.25 0.50 0.75 1.00
0.00 0.25 0.50 0.75
0.00 0.25 0.50 0.75
0.0 0.2 0.4 0.6 0.8
0.2 0.4 0.6 0.8 1.0
0.002 0.004 0.006

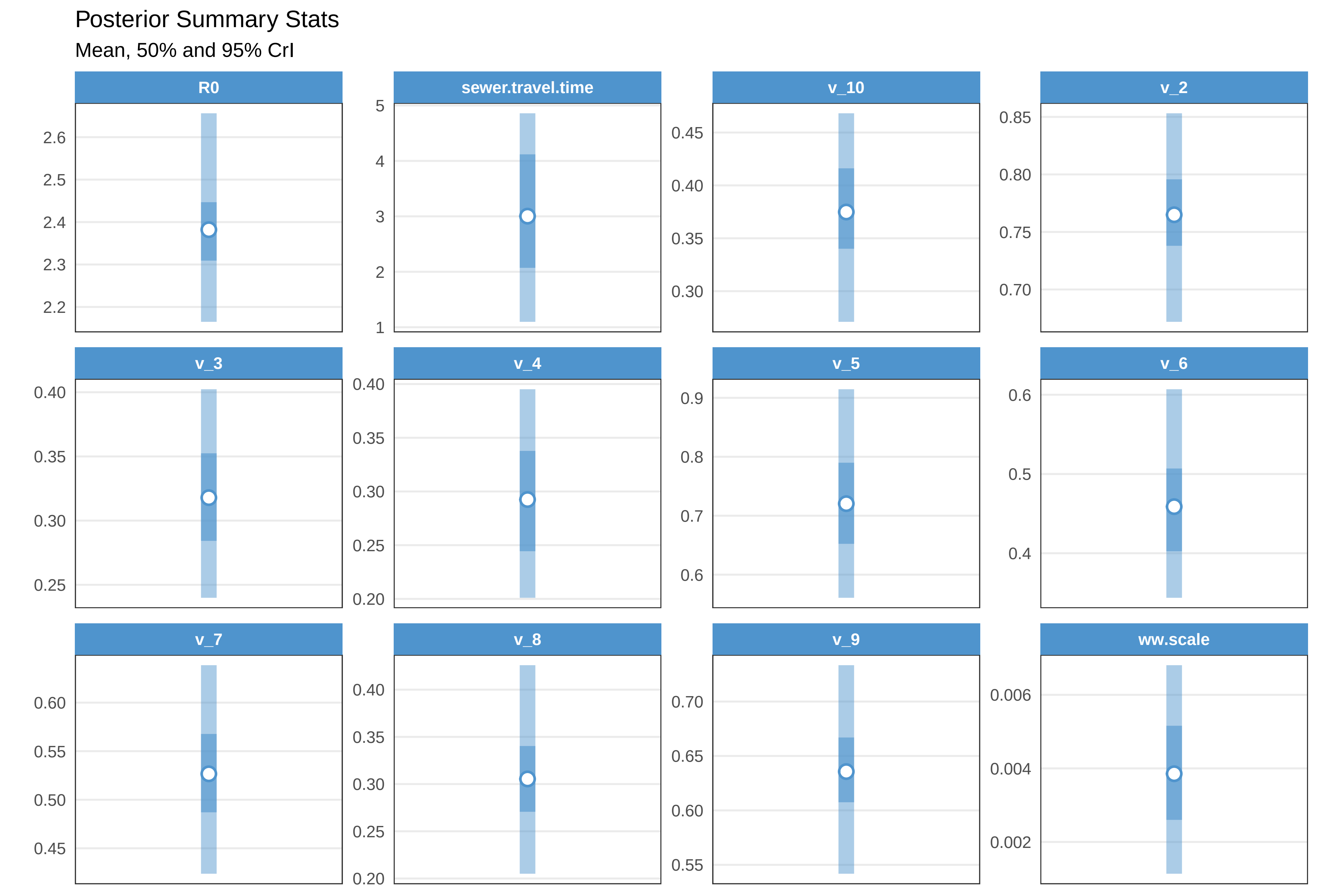
Posterior Summary Stats
Mean, 50% and 95% CrI
R0
2.6
2.5
2.4
2.3
2.2
sewer.travel.time
5
4
3
2
1
v_10
0.45
0.40
0.35
0.30
v_2
0.85
0.80
0.75
0.70
v_3
0.40
0.35
0.30
0.25
v_4
0.40
0.35
0.30
0.25
0.20
v_5
0.9
0.8
0.7
0.6
v_6
0.6
0.5
0.4
v_7
0.60
0.55
0.50
0.45
v_8
0.40
0.35
0.30
0.25
0.20
v_9
0.70
0.65
0.60
0.55
ww.scale
0.006
0.004
0.002

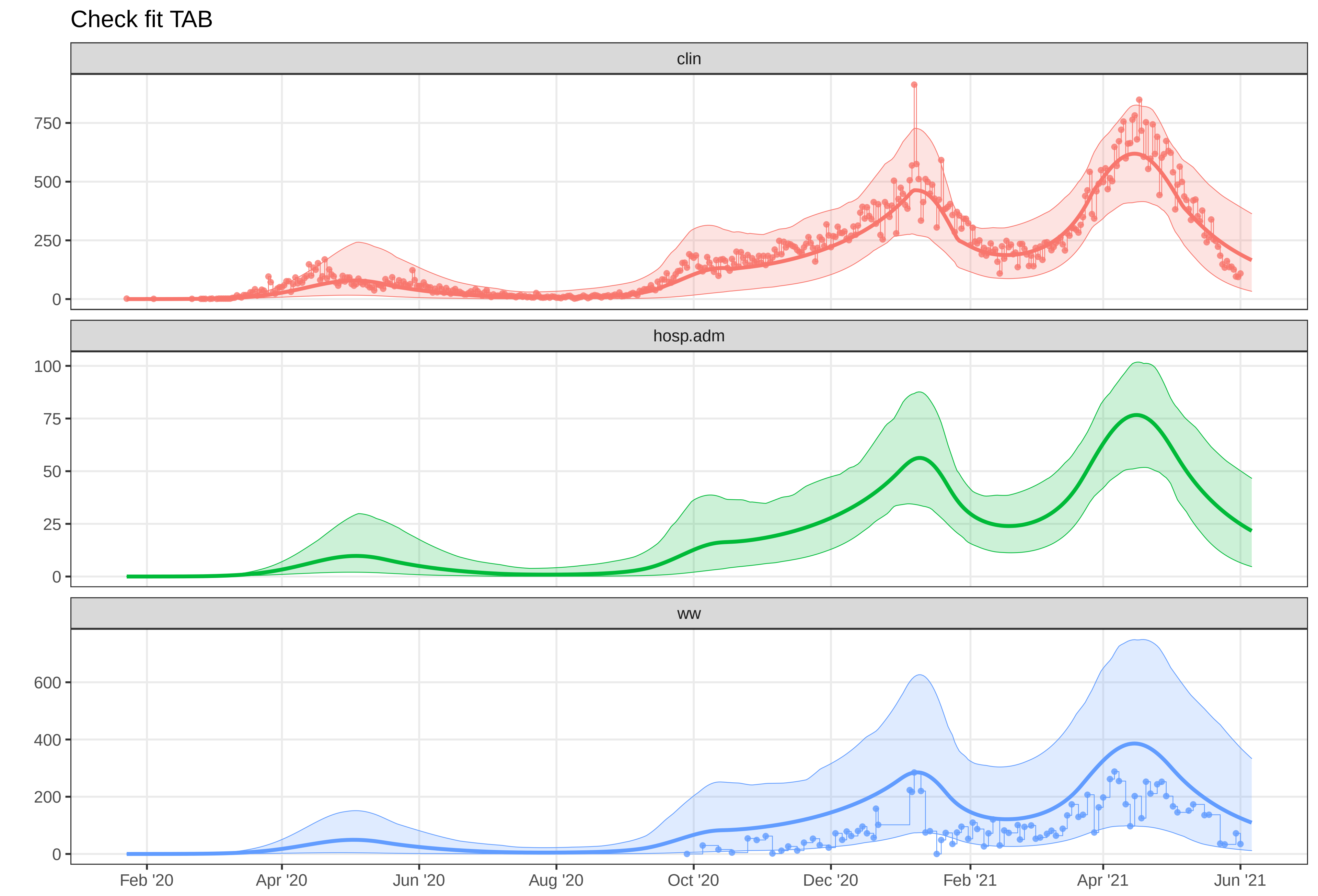

Check fit TAB
clin
750
500
250
0
hosp.adm
100
75
50
25
0
ww
600
400
200
0
Feb '20
Apr '20
Jun '20
Aug '20
Oct '20
Dec '20
Feb '21
Apr '21
Jun '21
