## Supplementary material for "A Wastewater-Based Epidemic Model for SARS-CoV-2 with Application to Three Canadian Cities": fit outputs: plot-fit-THU-07-06-clinww.pdf

Observed data for THU

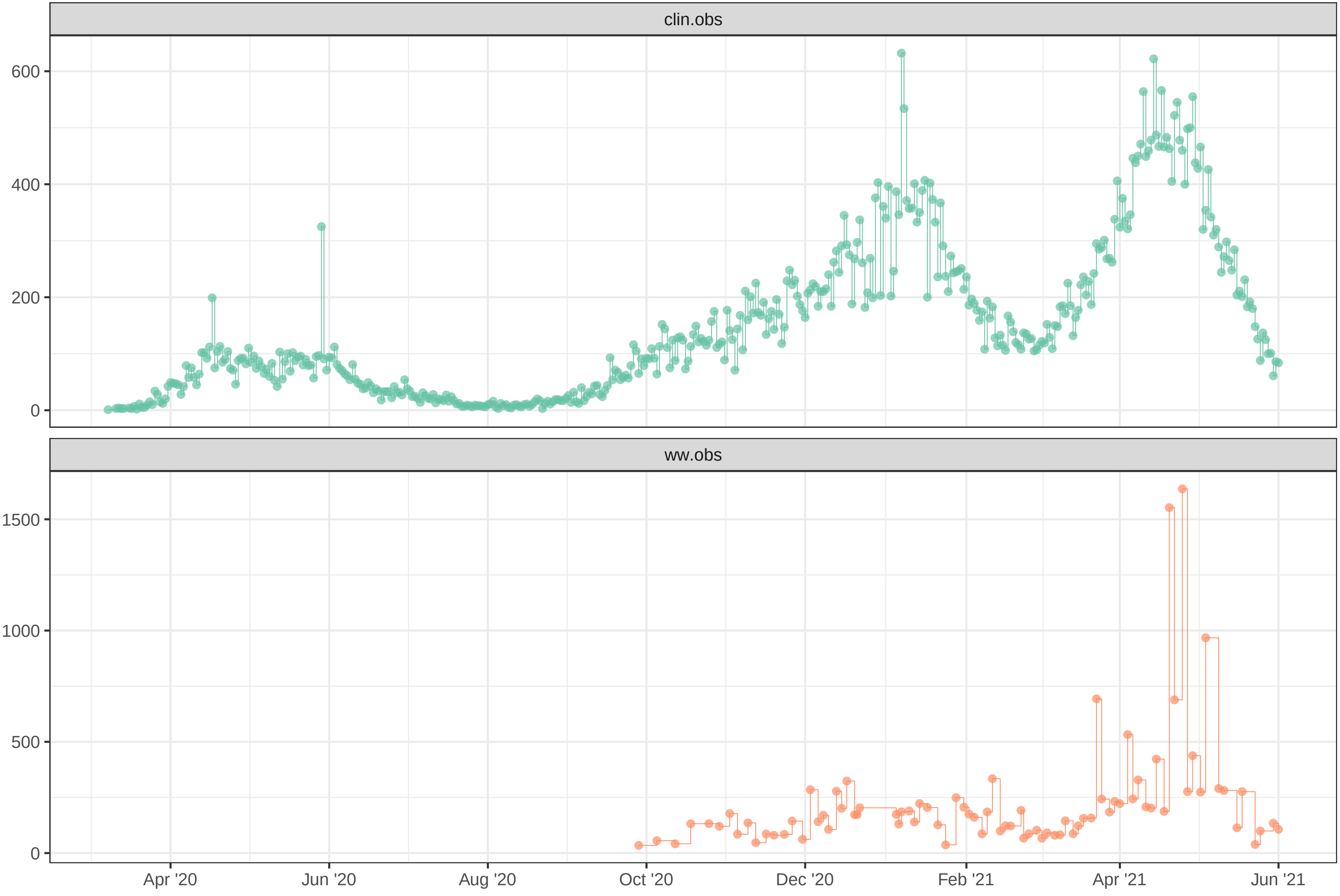

Simulation with initial parameters for THU

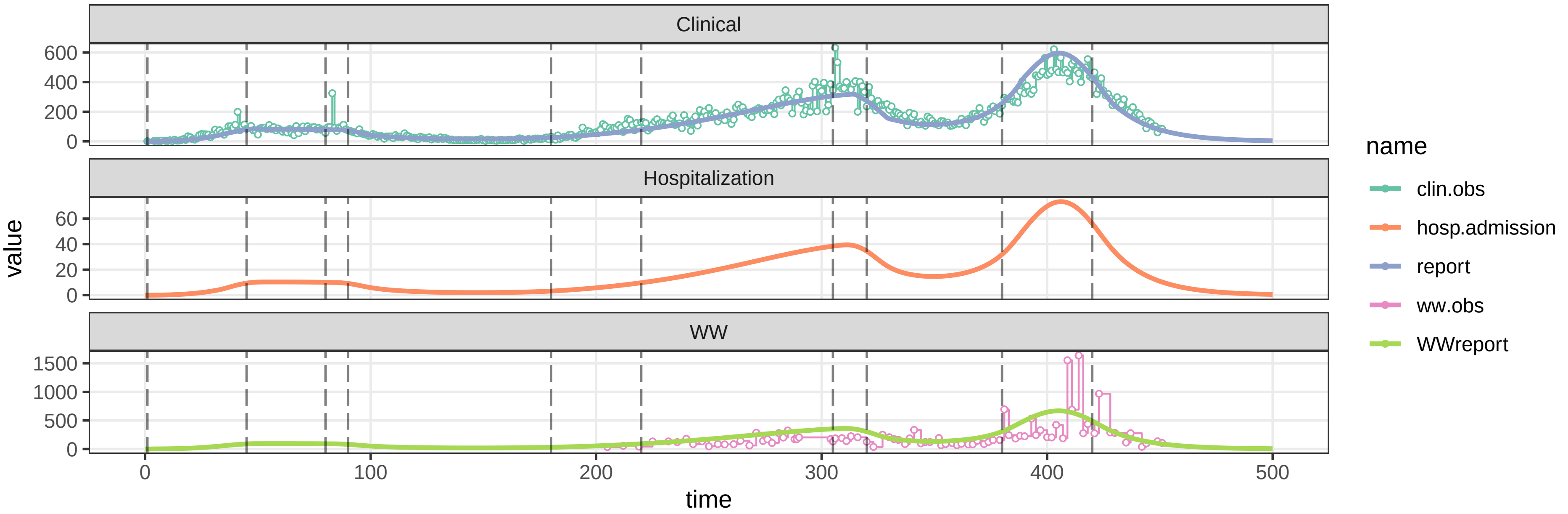


parameters/prm−THU/prm−interv.csv

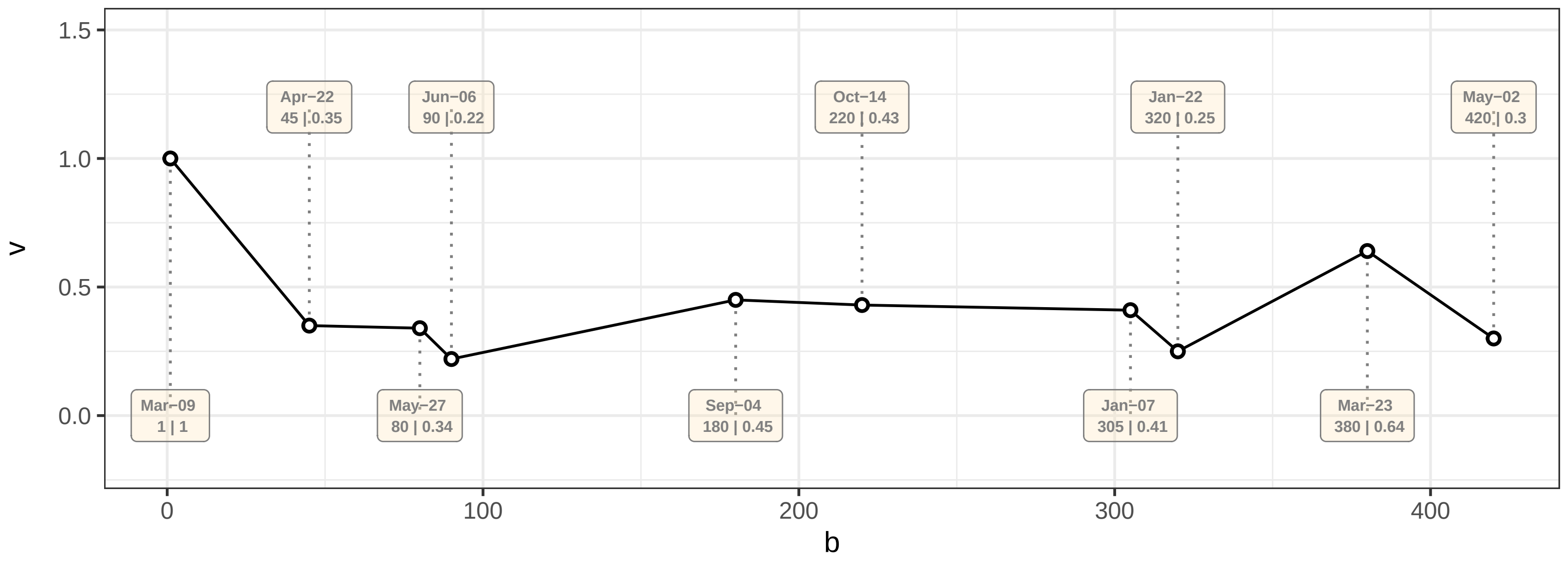

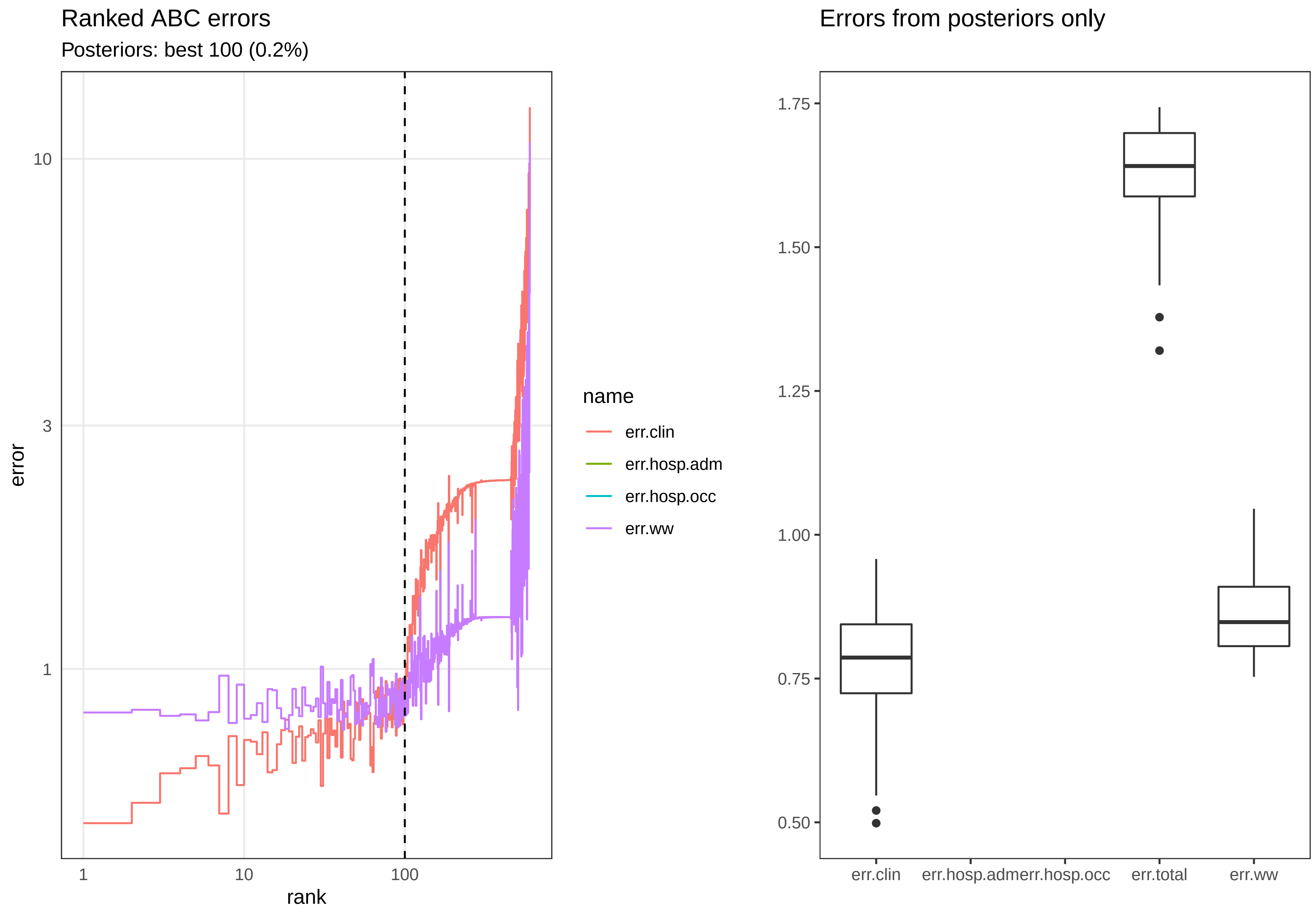
Ranked ABC errors
Posteriors: best 100 (0.2%)
error
10
3
1
1
10
100
rank
name
err.clin
err.hosp.adm
err.hosp.occ
err.ww
Errors from posteriors only
1.75
1.50
1.25
1.00
0.75
0.50
err.clin
err.hosp.adm
err.hosp.occ
err.total
err.ww

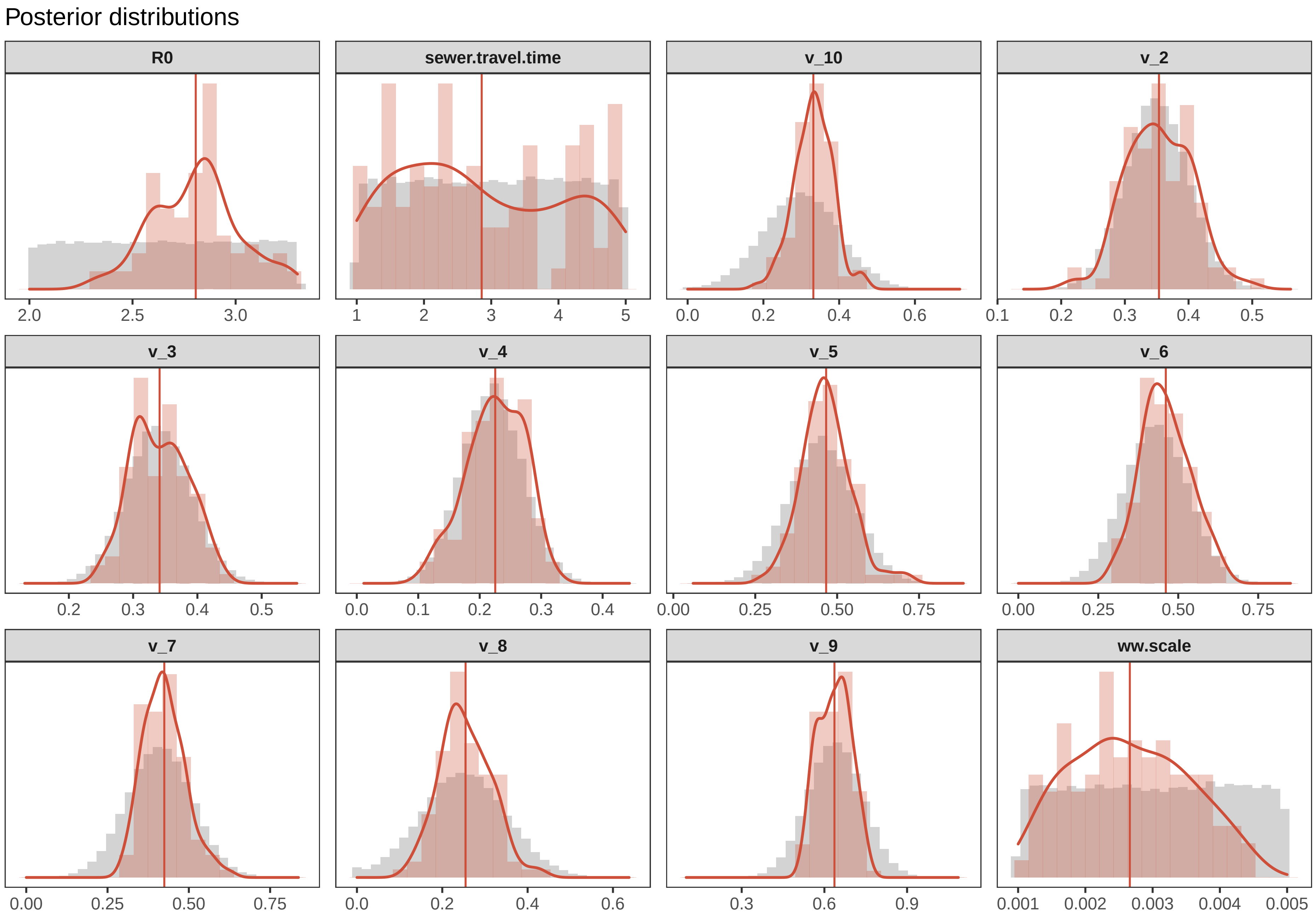

Posterior distributions
R0
sewer.travel.time
v_10
v_2
v_3
v_4
v_5
v_6
v_7
v_8
v_9
ww.scale
2.0
2.5
3.0
1
2
3
4
5
0.0
0.2
0.4
0.6
0.1
0.2
0.3
0.4
0.5
0.2
0.3
0.4
0.5
0.0
0.1
0.2
0.3
0.4
0.00
0.25
0.50
0.75
0.00
0.25
0.50
0.75
0.00
0.25
0.50
0.75
0.0
0.2
0.4
0.6
0.3
0.6
0.9
0.001
0.002
0.003
0.004
0.005

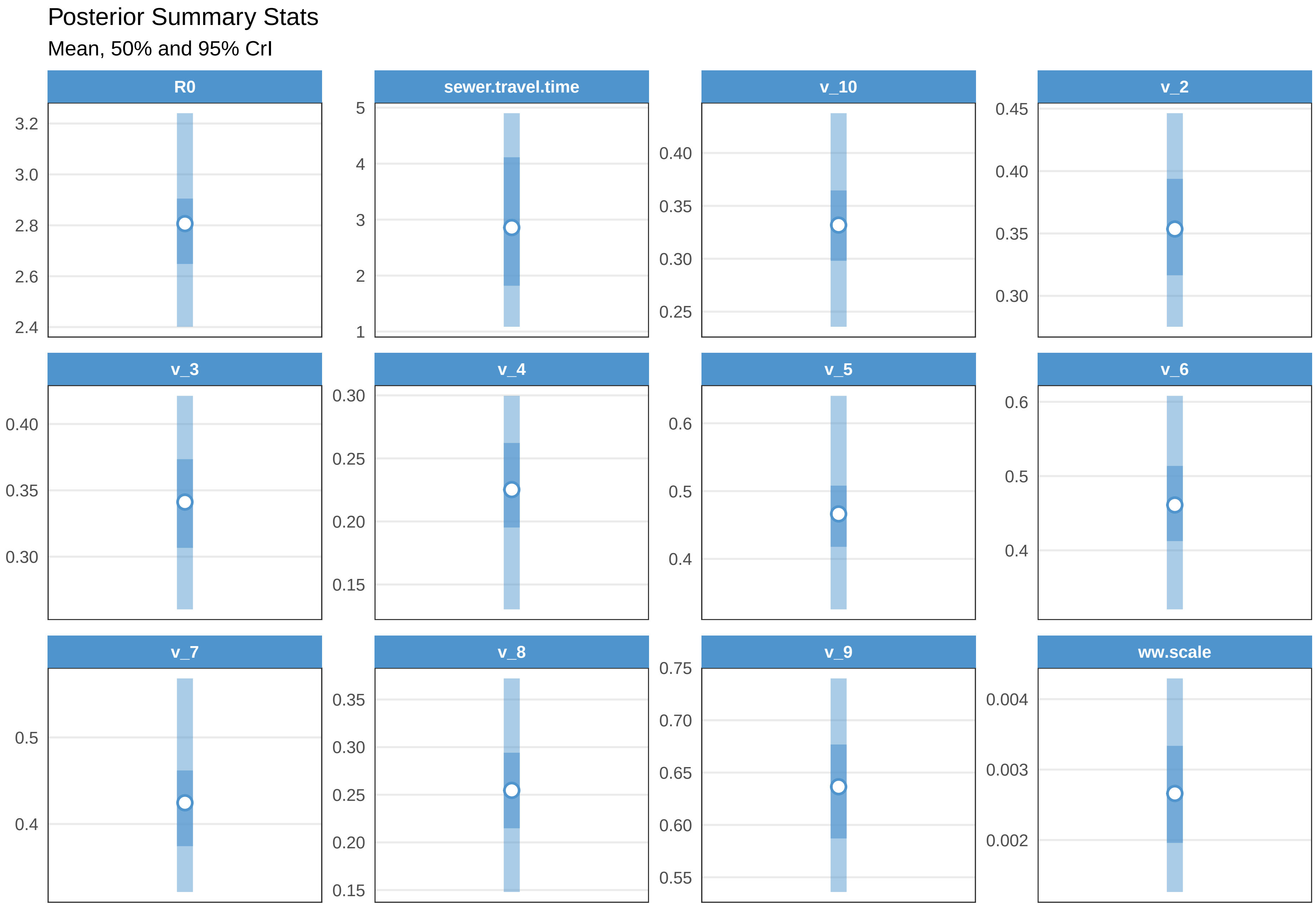
Posterior Summary Stats
Mean, 50% and 95% CrI
R0
3.2
3.0
2.8
2.6
2.4
sewer.travel.time
5
4
3
2
1
v_10
0.40
0.35
0.30
0.25
v_2
0.45
0.40
0.35
0.30
v_3
0.40
0.35
0.30
v_4
0.30
0.25
0.20
0.15
v_5
0.6
0.5
0.4
v_6
0.6
0.5
0.4
v_7
0.5
0.4
v_8
0.35
0.30
0.25
0.20
0.15
v_9
0.75
0.70
0.65
0.60
0.55
ww.scale
0.004
0.003
0.002

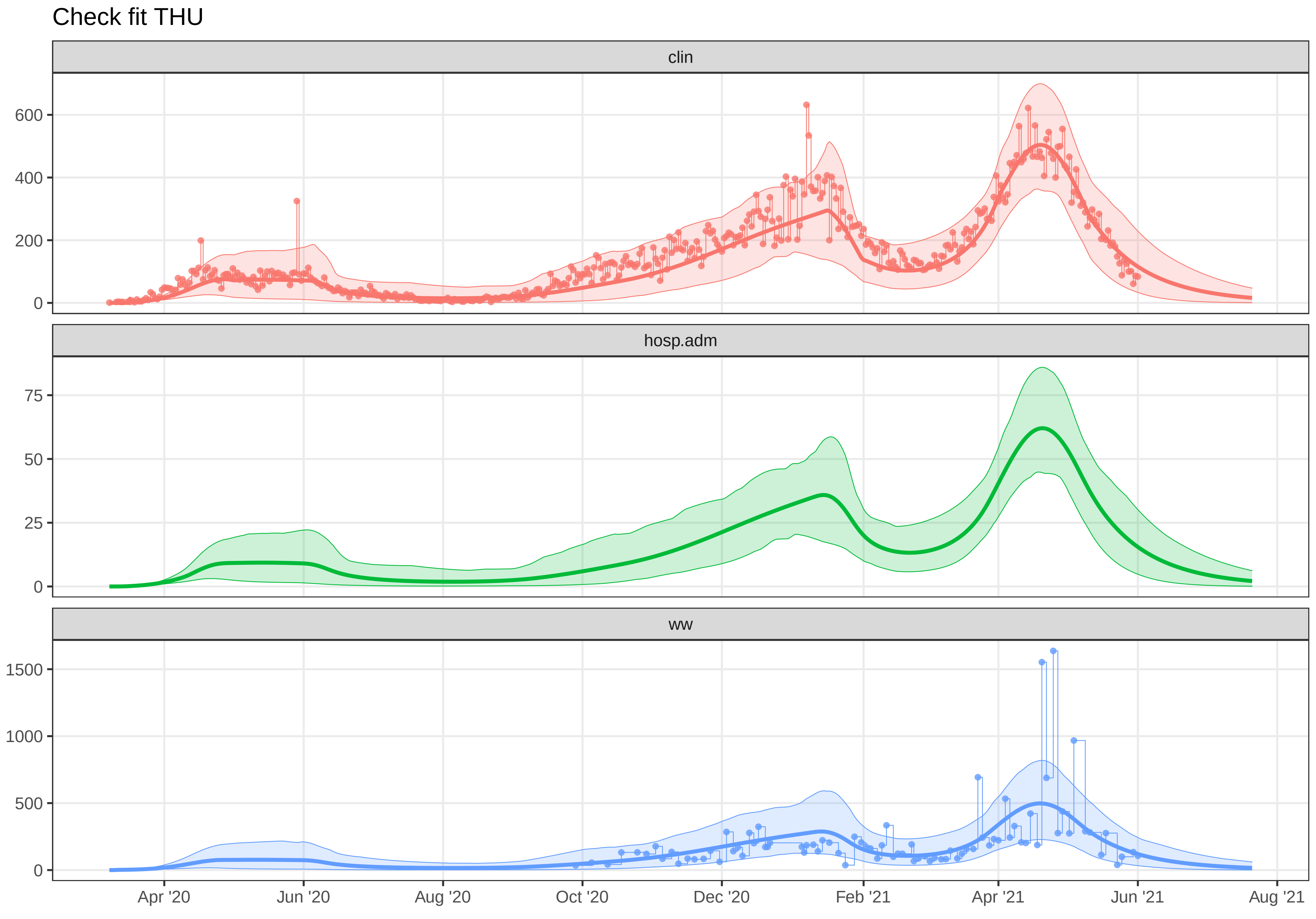
Check fit THU
clin
600
400
200
0
hosp.adm
75
50
25
0
ww
1500
1000
500
0
Apr '20
Jun '20
Aug '20
Oct '20
Dec '20
Feb '21
Apr '21
Jun '21
Aug '21
