## Supplementary material for "A Wastewater-Based Epidemic Model for SARS-CoV-2 with Application to Three Canadian Cities": fit outputs: plot-fit-TNT-07-08-clin.pdf

Observed data for TNT

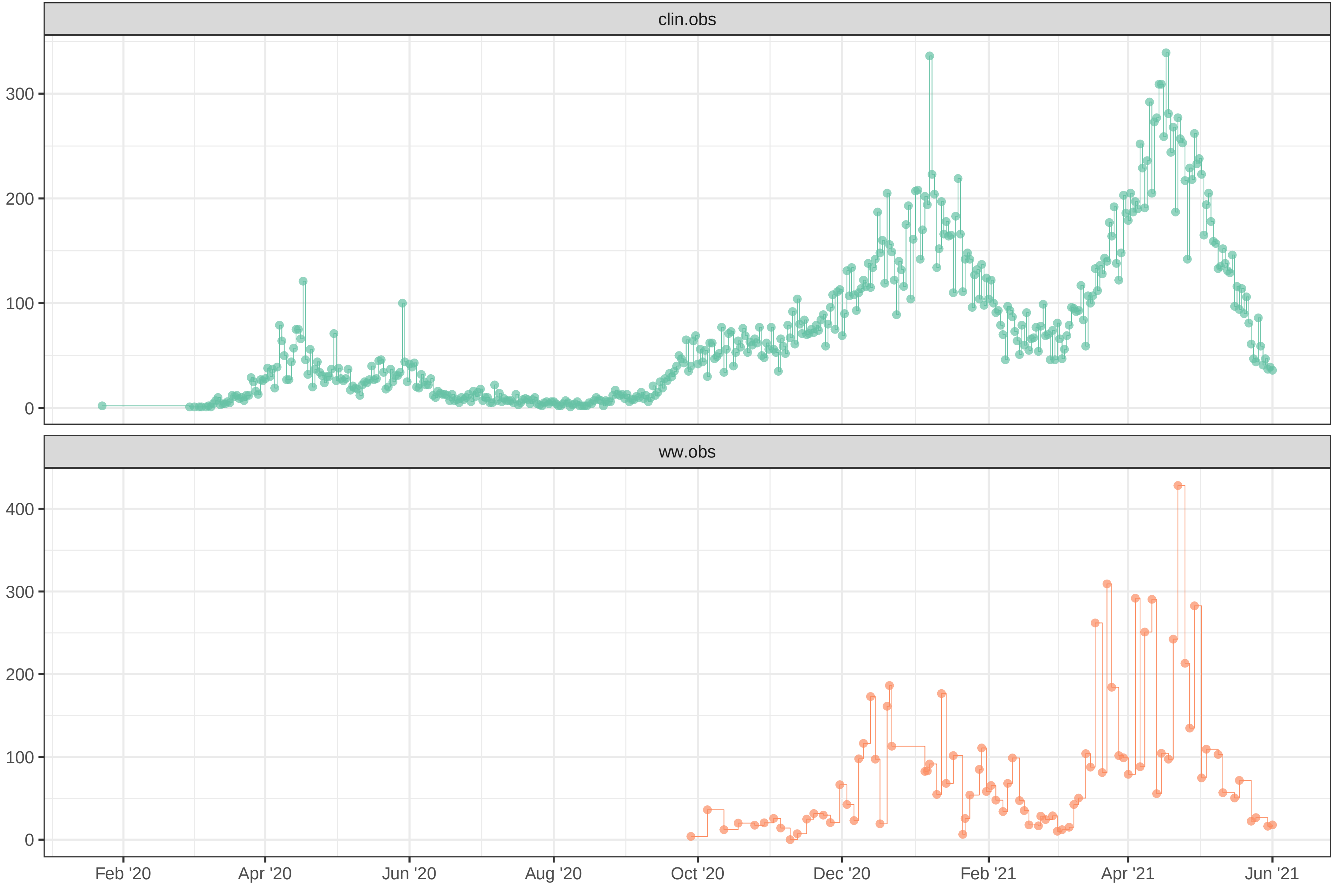

Simulation with initial parameters for TNT

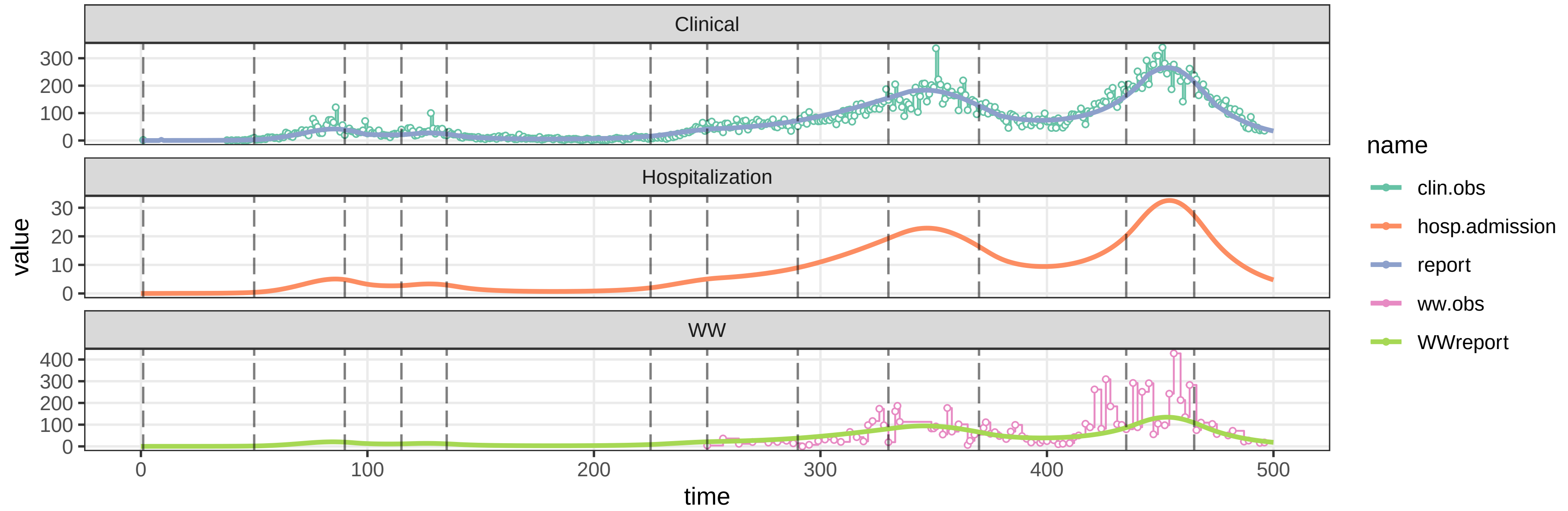


parameters/prm−TNT/prm−interv.csv

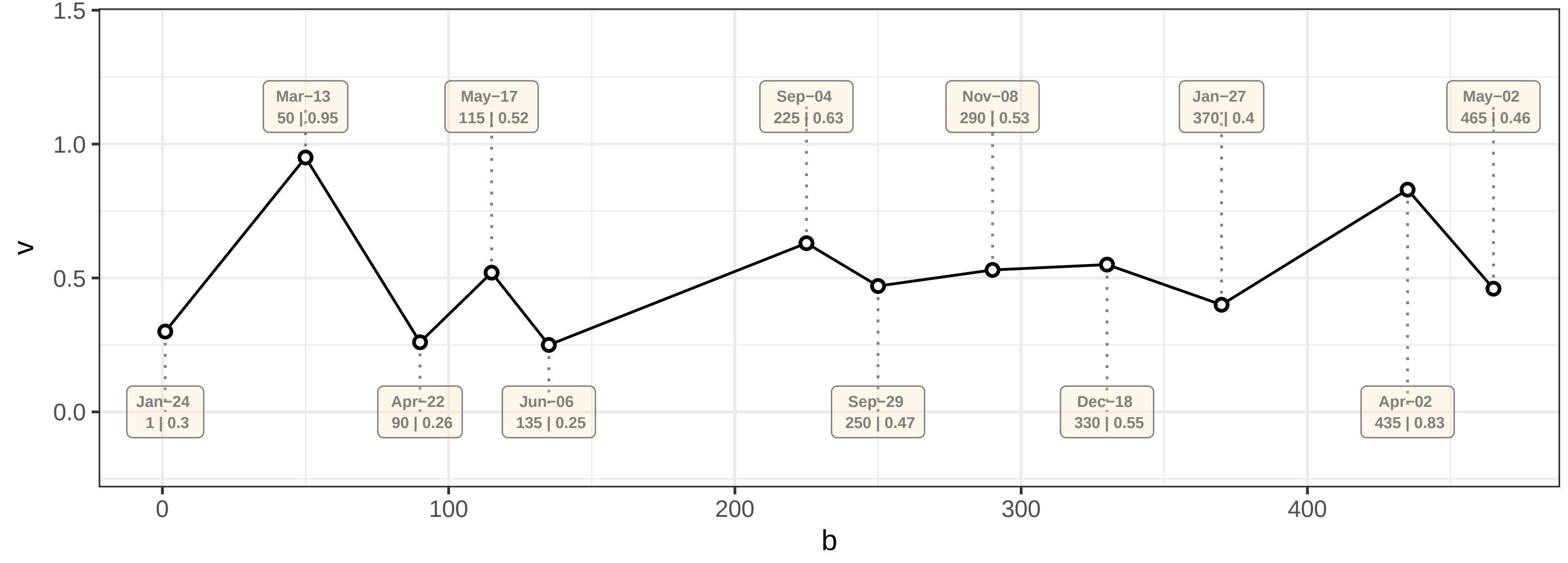

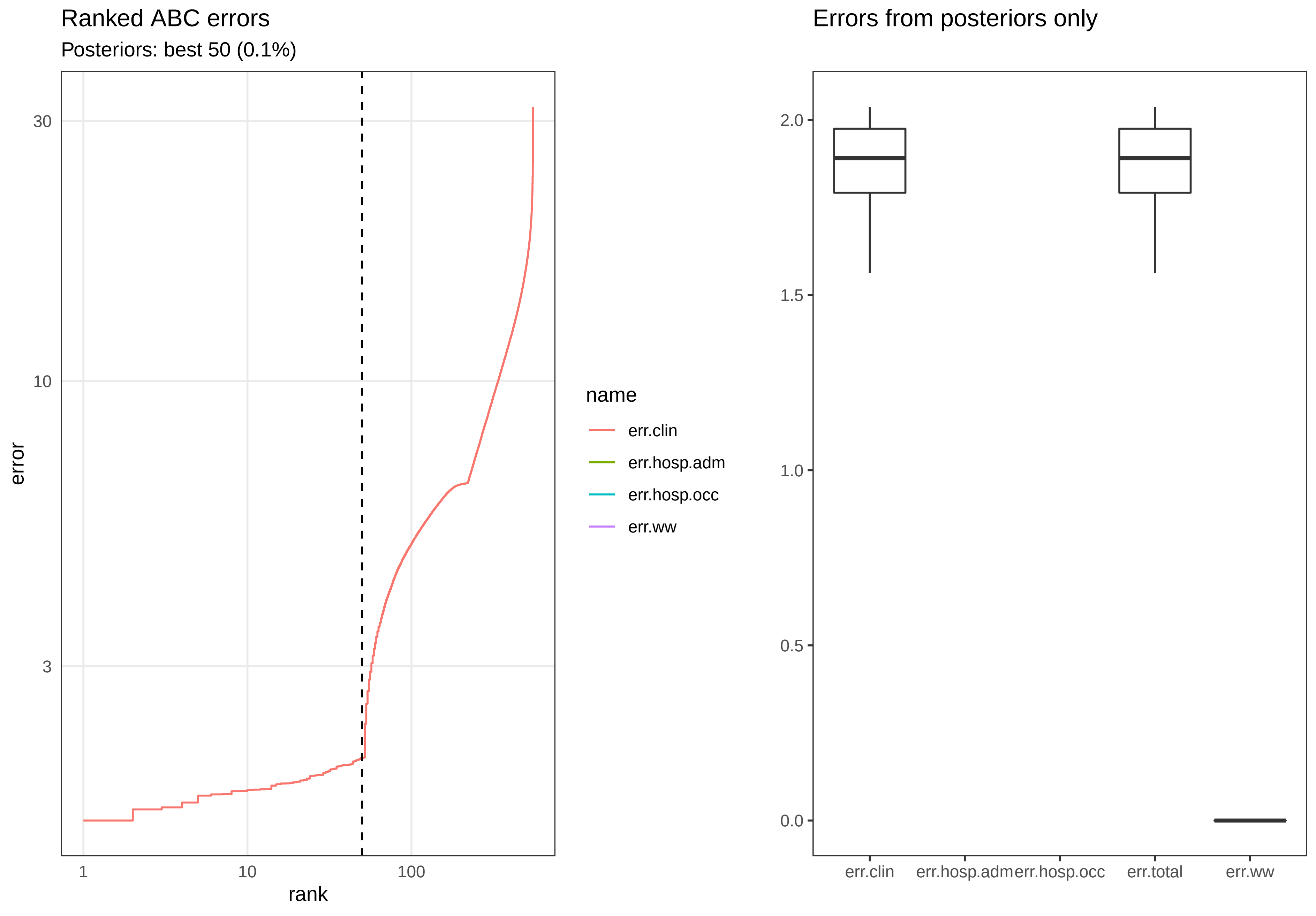

Ranked ABC errors
Posteriors: best 50 (0.1%)
30
10
3
error
1
10
100
rank
name
err.clin
err.hosp.adm
err.hosp.occ
err.ww
Errors from posteriors only
2.0
1.5
1.0
0.5
0.0
err.clin
err.hosp.adm
err.hosp.occ
err.total
err.ww

Posterior distributions

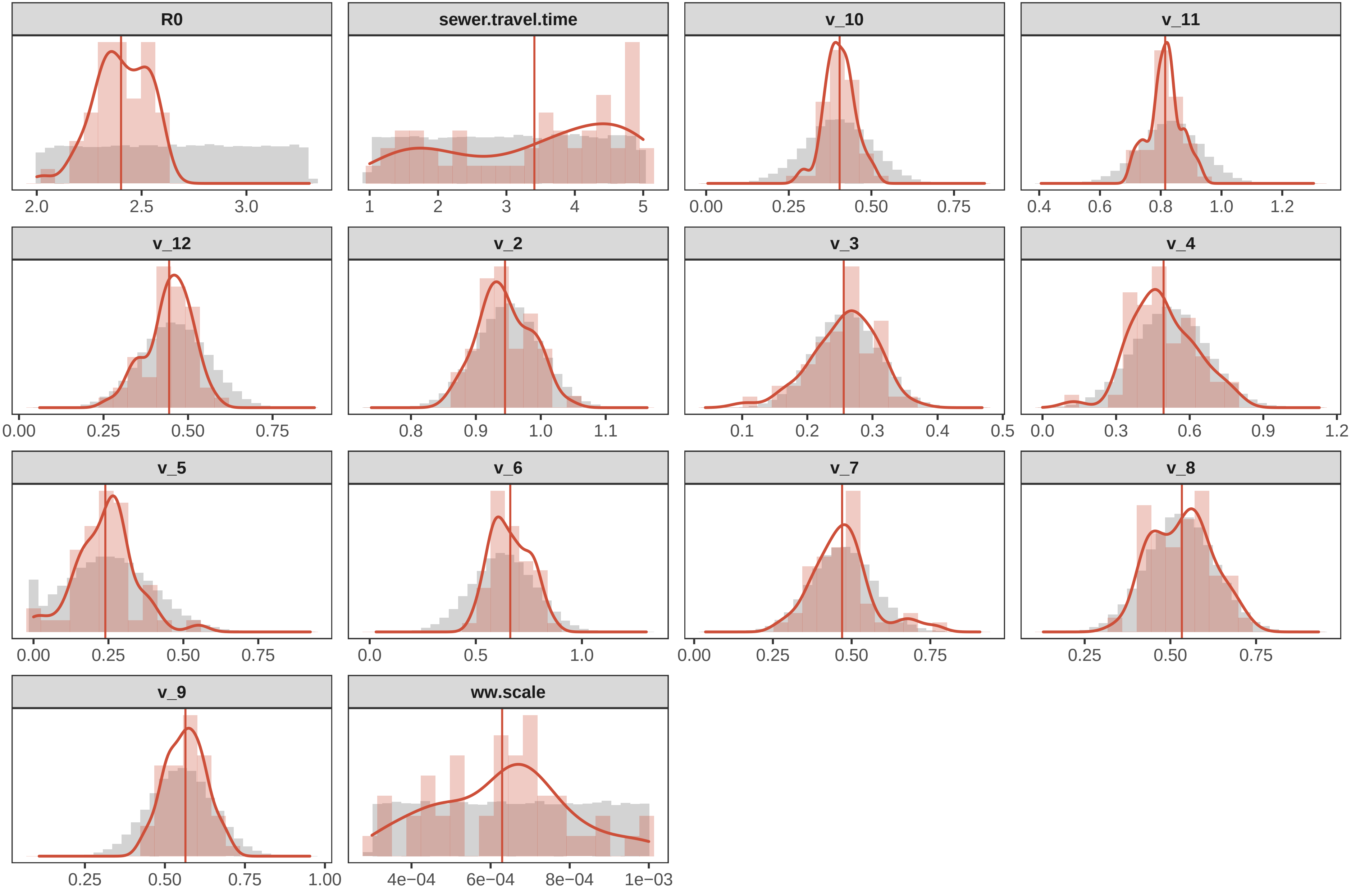

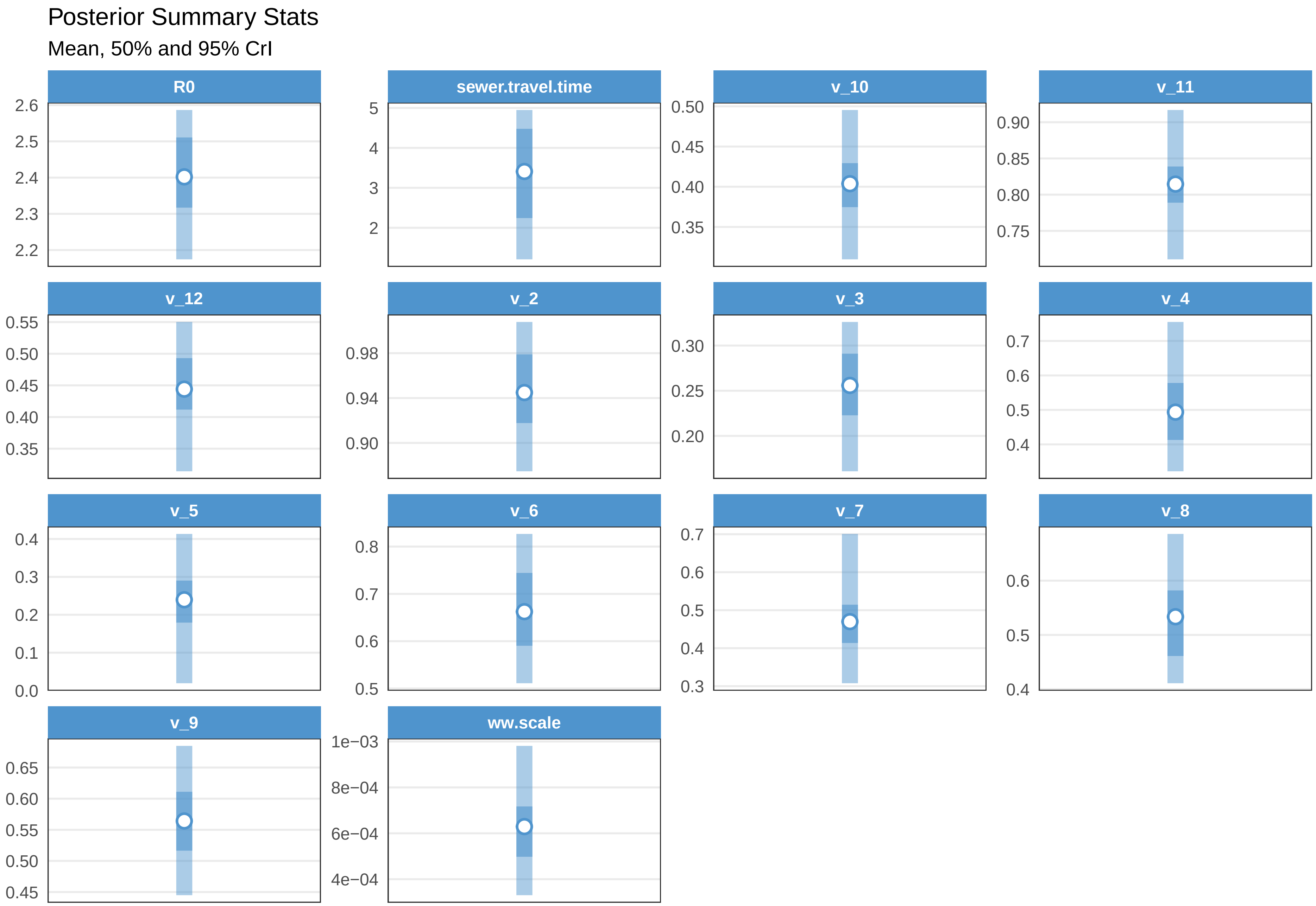

Posterior Summary Stats
Mean, 50% and 95% CrI
R0
2.6
2.5
2.4
2.3
2.2
sewer.travel.time
5
4
3
2
v_10
0.50
0.45
0.40
0.35
v_11
0.90
0.85
0.80
0.75
v_12
0.55
0.50
0.45
0.40
0.35
v_2
0.98
0.94
0.90
v_3
0.30
0.25
0.20
v_4
0.7
0.6
0.5
0.4
v_5
0.4
0.3
0.2
0.1
0.0
v_6
0.8
0.7
0.6
0.5
v_7
0.7
0.6
0.5
0.4
0.3
v_8
0.6
0.5
0.4
v_9
0.65
0.60
0.55
0.50
0.45
ww.scale
1e−03
8e−04
6e−04
4e−04

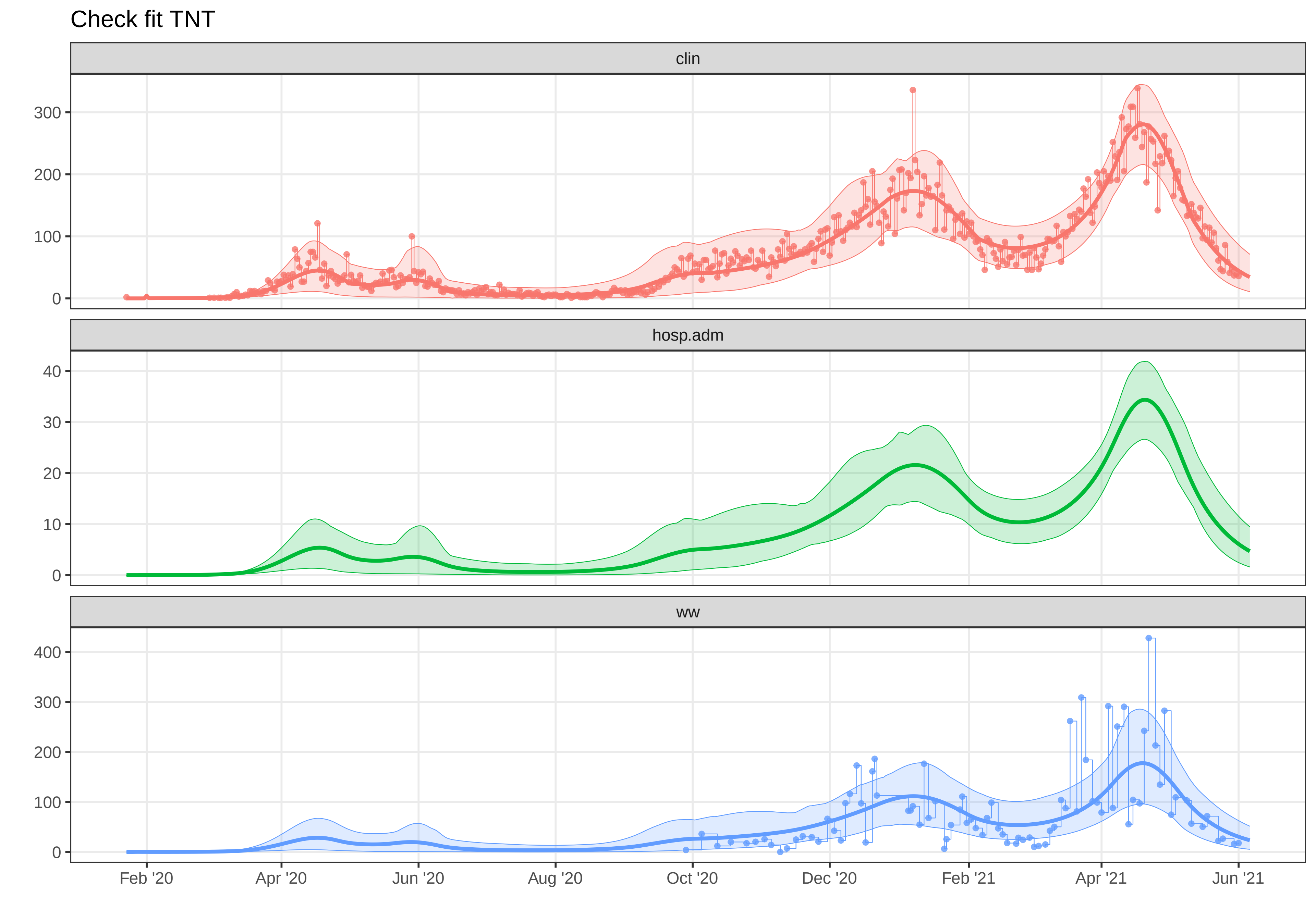
Check fit TNT
clin
hosp.adm
ww
300
200
100
0
40
30
20
10
0
400
300
200
100
0
Feb '20
Apr '20
Jun '20
Aug '20
Oct '20
Dec '20
Feb '21
Apr '21
Jun '21
