## Supplementary material for "A Wastewater-Based Epidemic Model for SARS-CoV-2 with Application to Three Canadian Cities": fit outputs: plot-fit-TNT-07-08-clinww.pdf

### Observed data for TNT

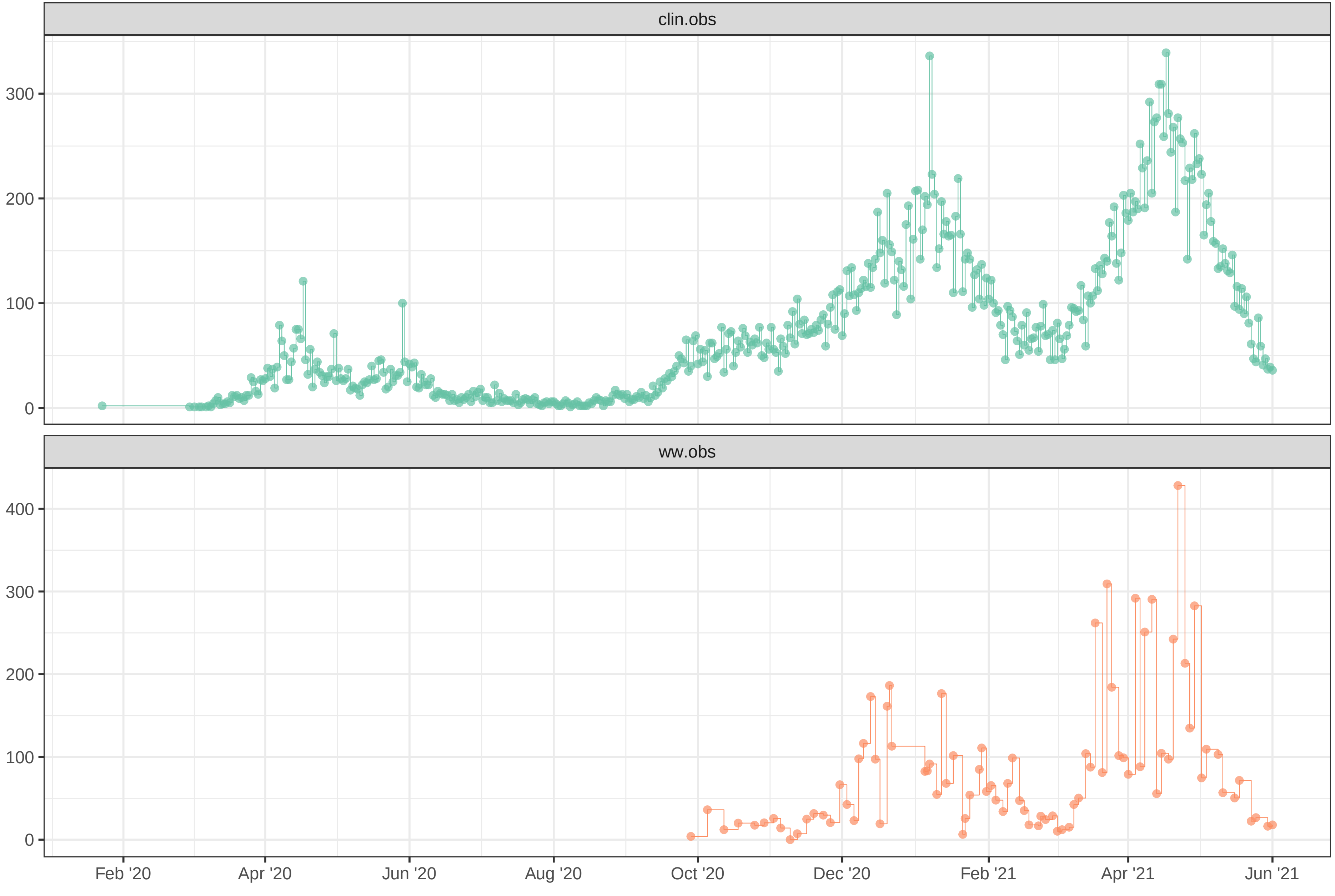

Simulation with initial parameters for TNT

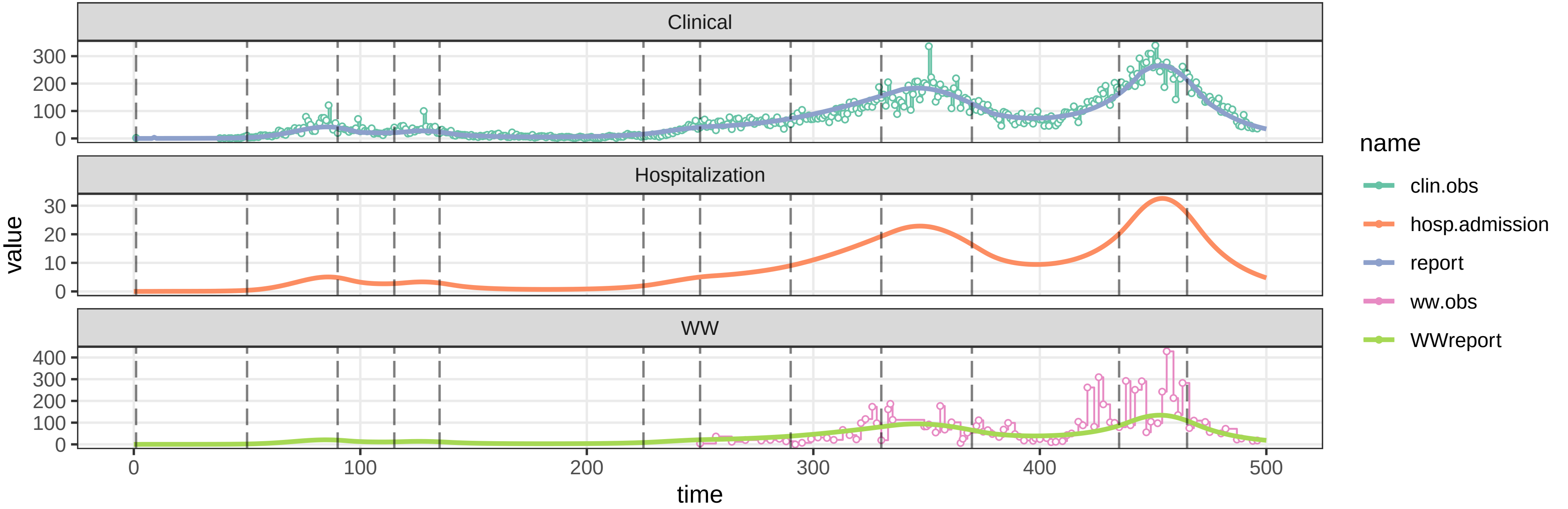


parameters/prm−TNT/prm−interv.csv

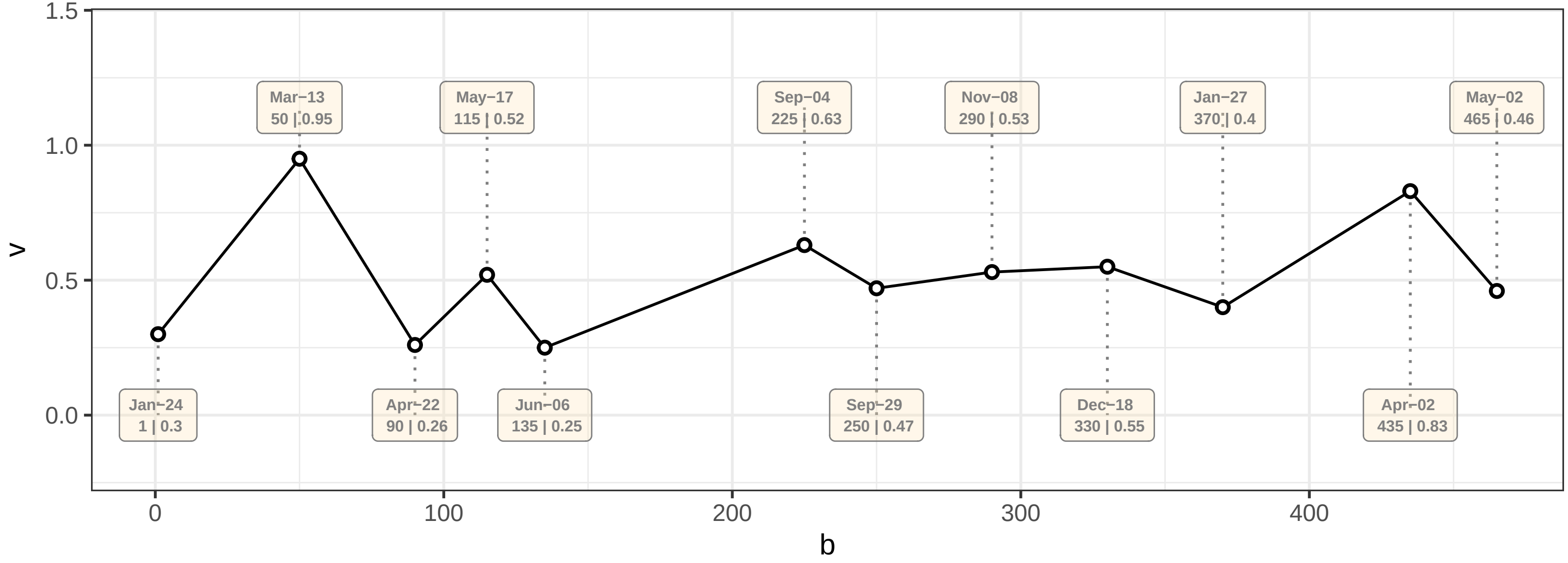

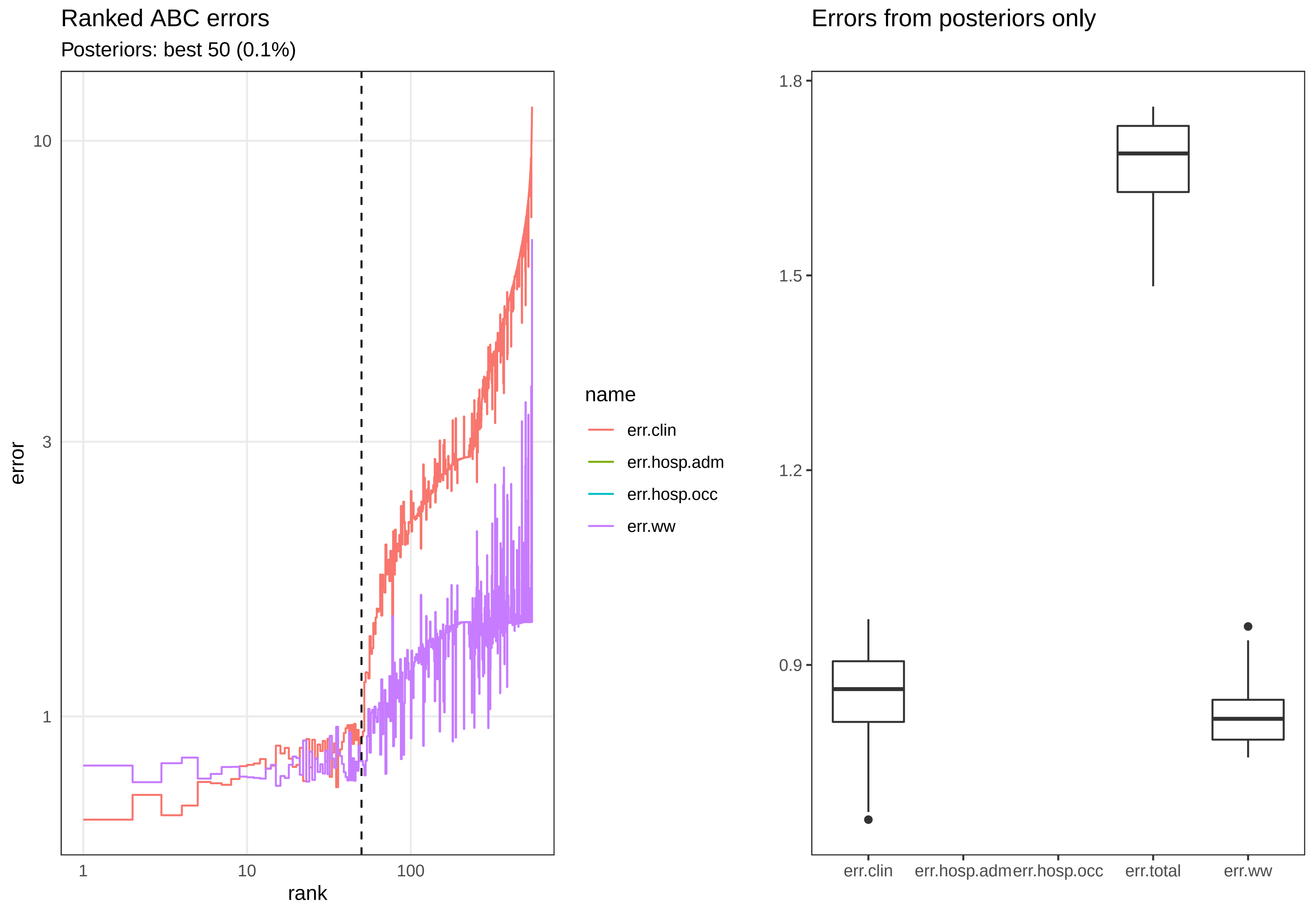

Ranked ABC errors
Posteriors: best 50 (0.1%)
10
3
1
error
1
10
100
rank
name
err.clin
err.hosp.adm
err.hosp.occ
err.ww
Errors from posteriors only
1.8
1.5
1.2
0.9
err.clin
err.hosp.adm
err.hosp.occ
err.total
err.ww

Posterior distributions

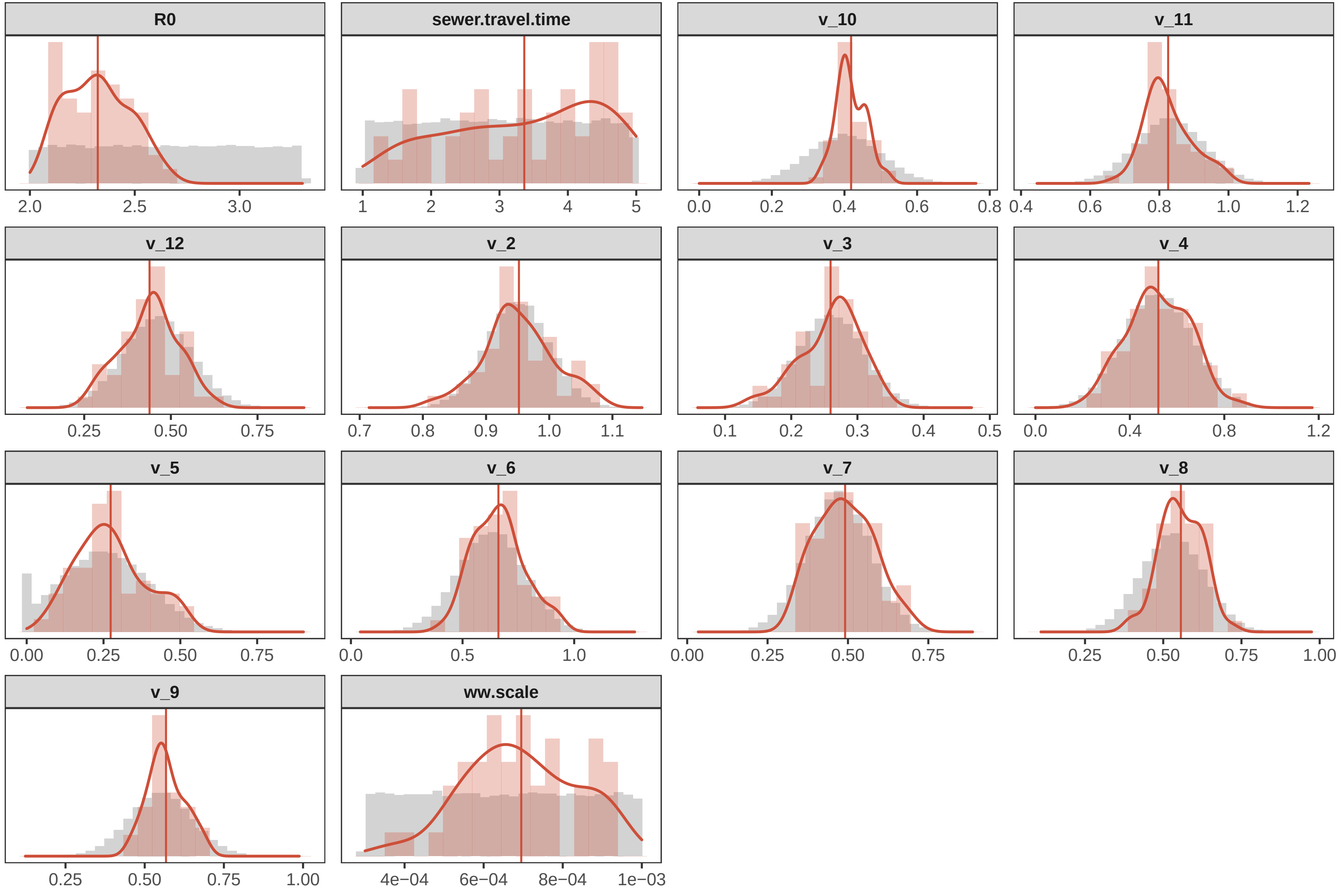

### Posterior Summary Stats

Mean, 50% and 95% CrI

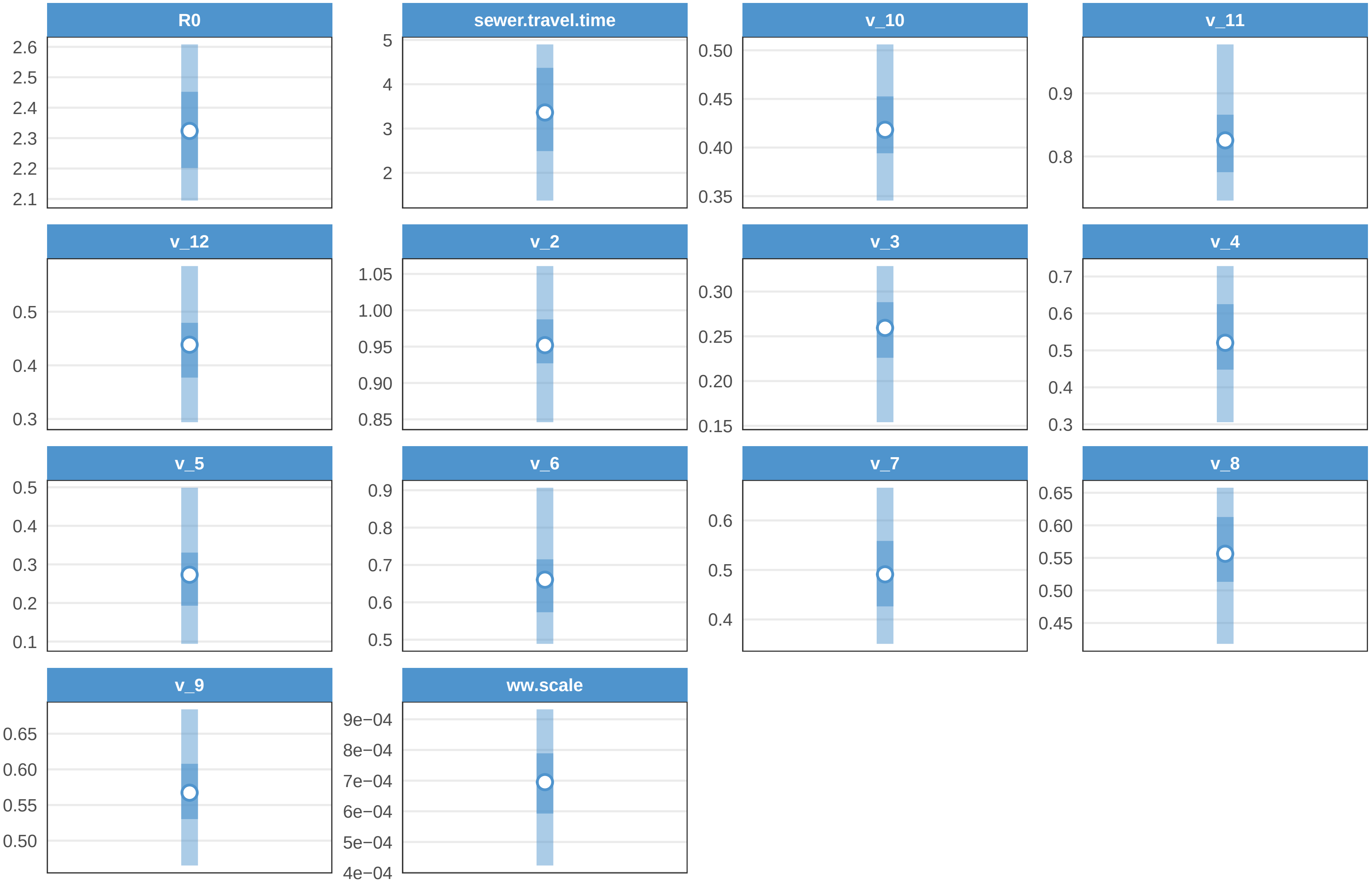

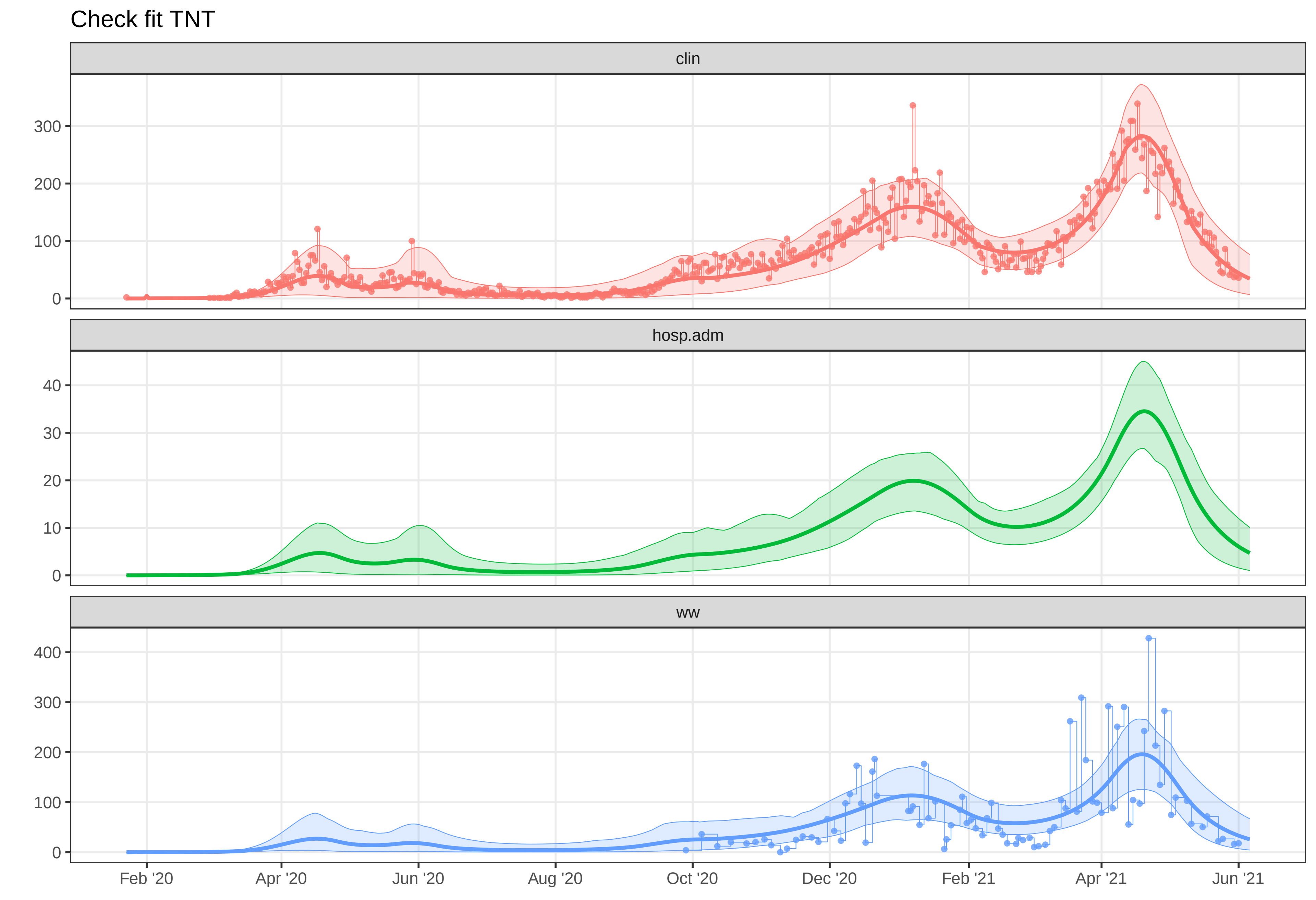
Check fit TNT
clin
300
200
100
0
hosp.adm
40
30
20
10
0
ww
400
300
200
100
0
Feb '20
Apr '20
Jun '20
Aug '20
Oct '20
Dec '20
Feb '21
Apr '21
Jun '21
