## Supplementary figures and images for "A Wastewater-Based Epidemic Model for SARS-CoV-2 with Application to Three Canadian Cities"

### plot-fit-OTW-07-06-1017-ww.pdf

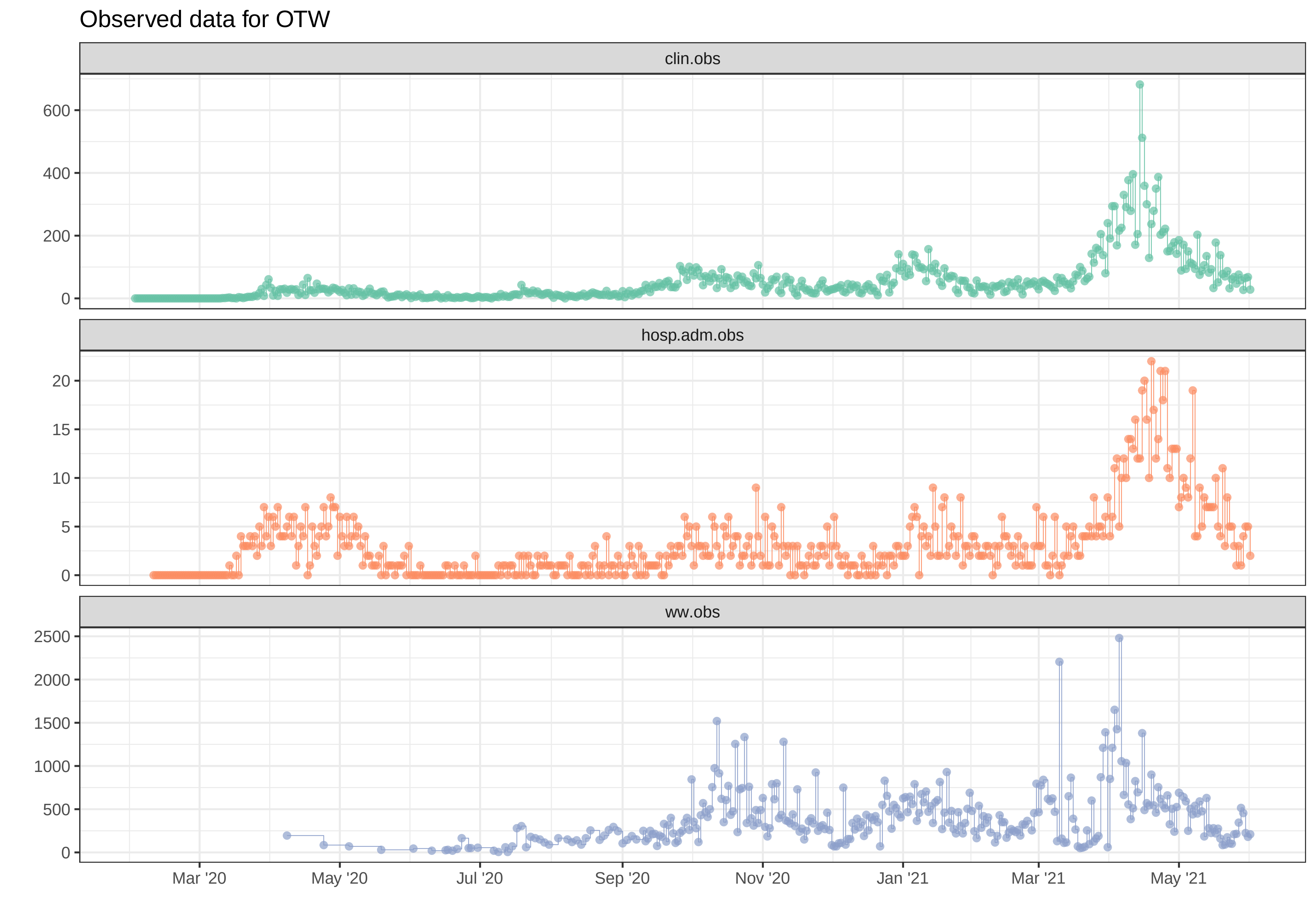

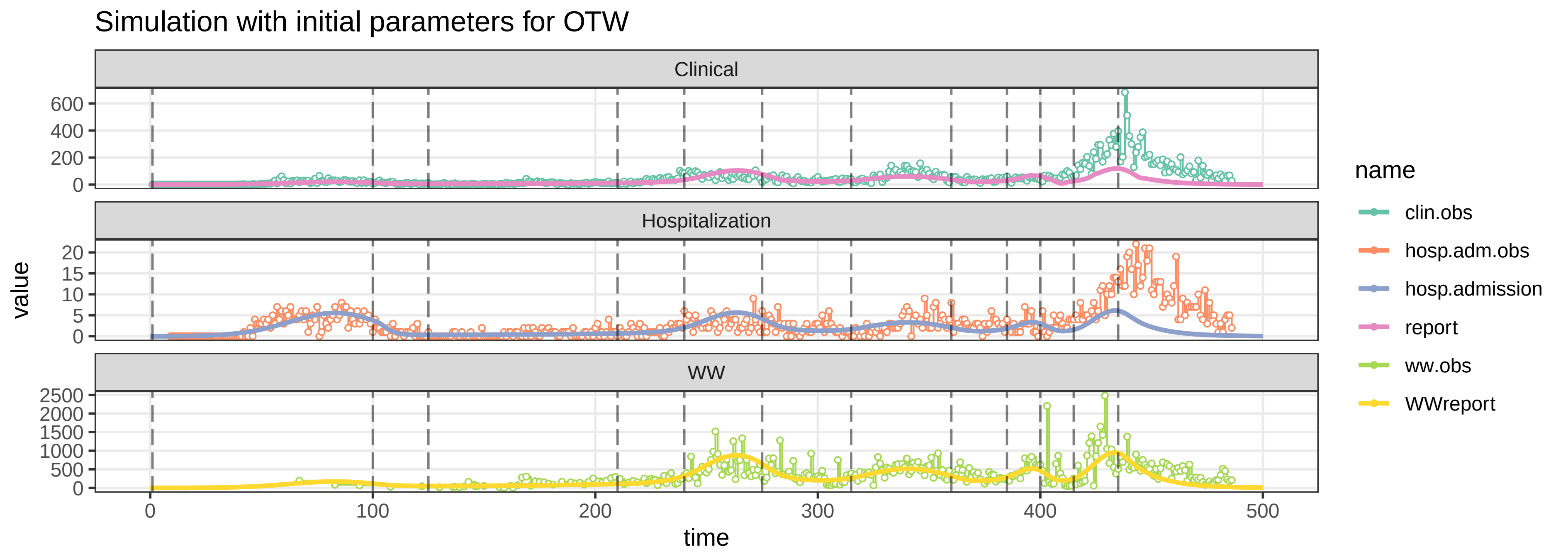

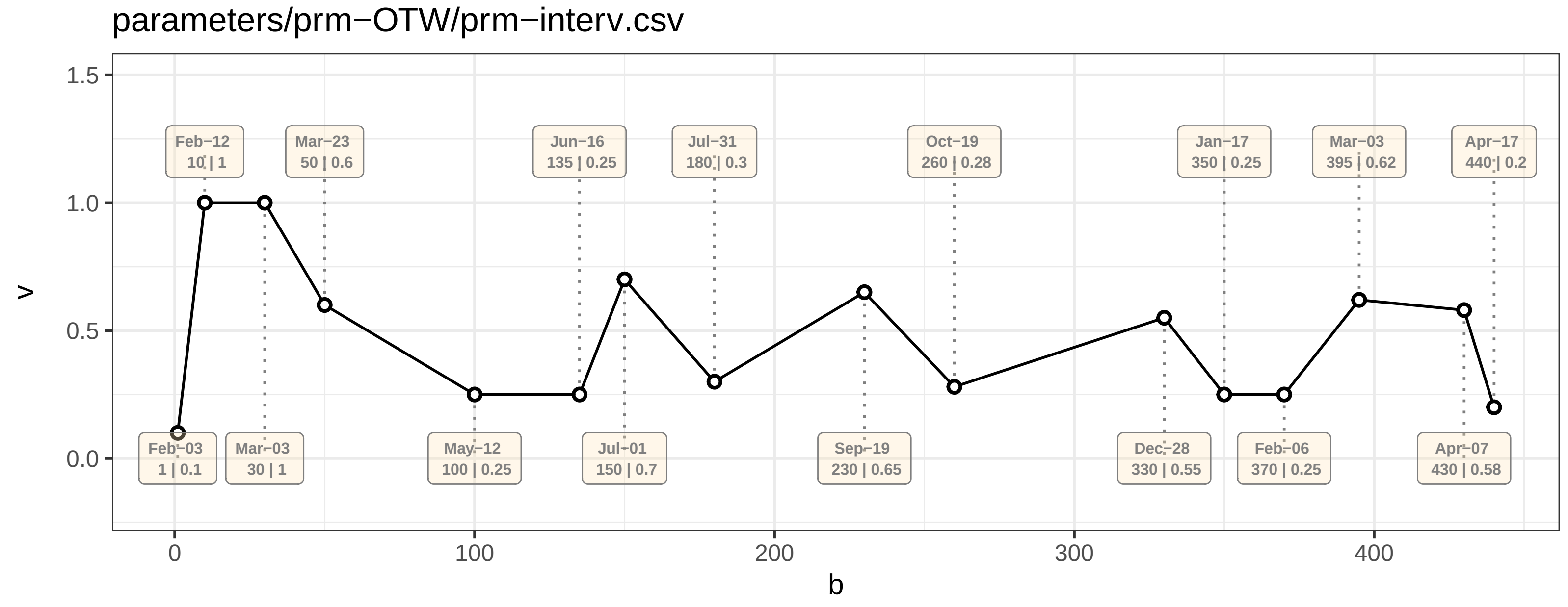

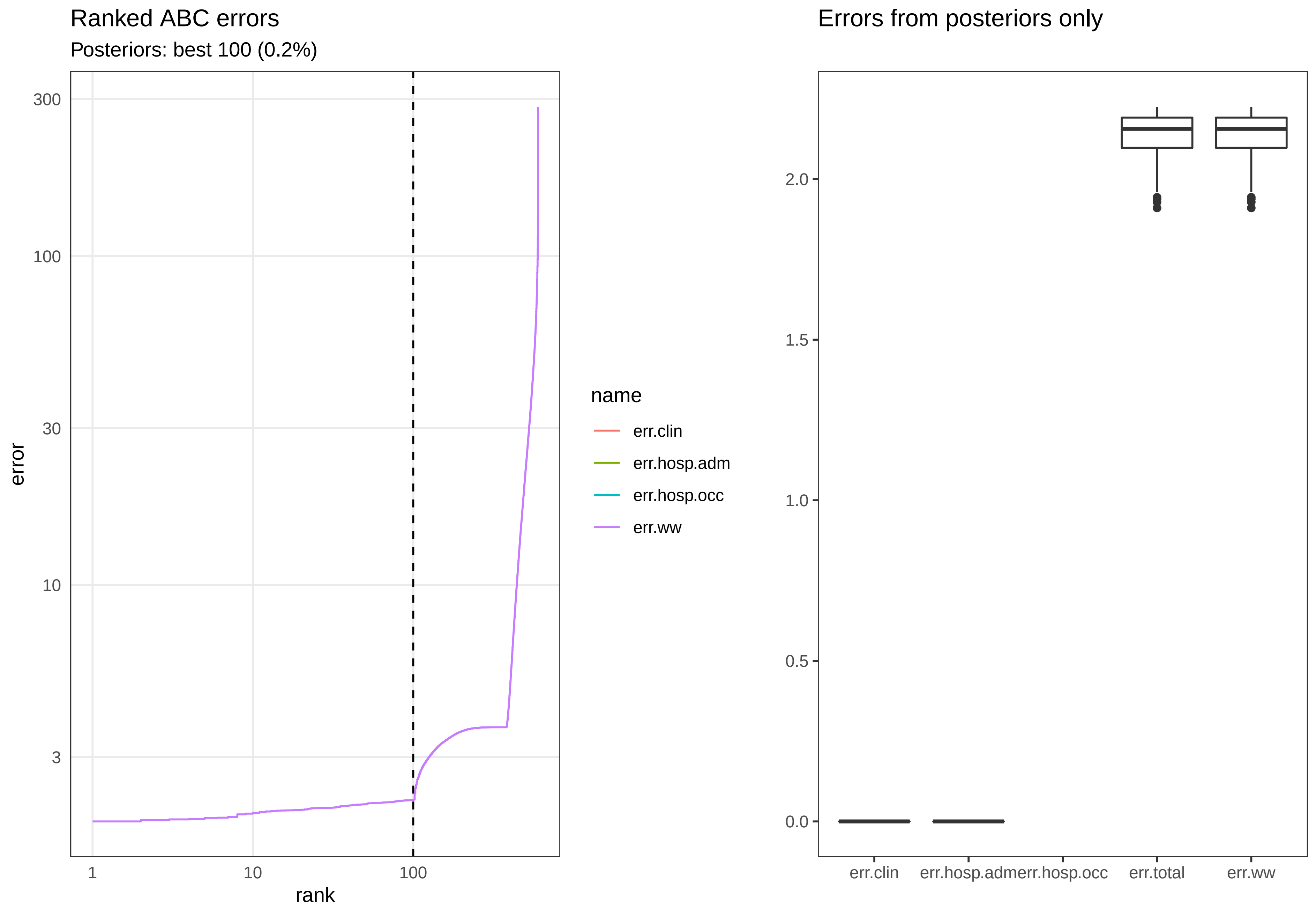

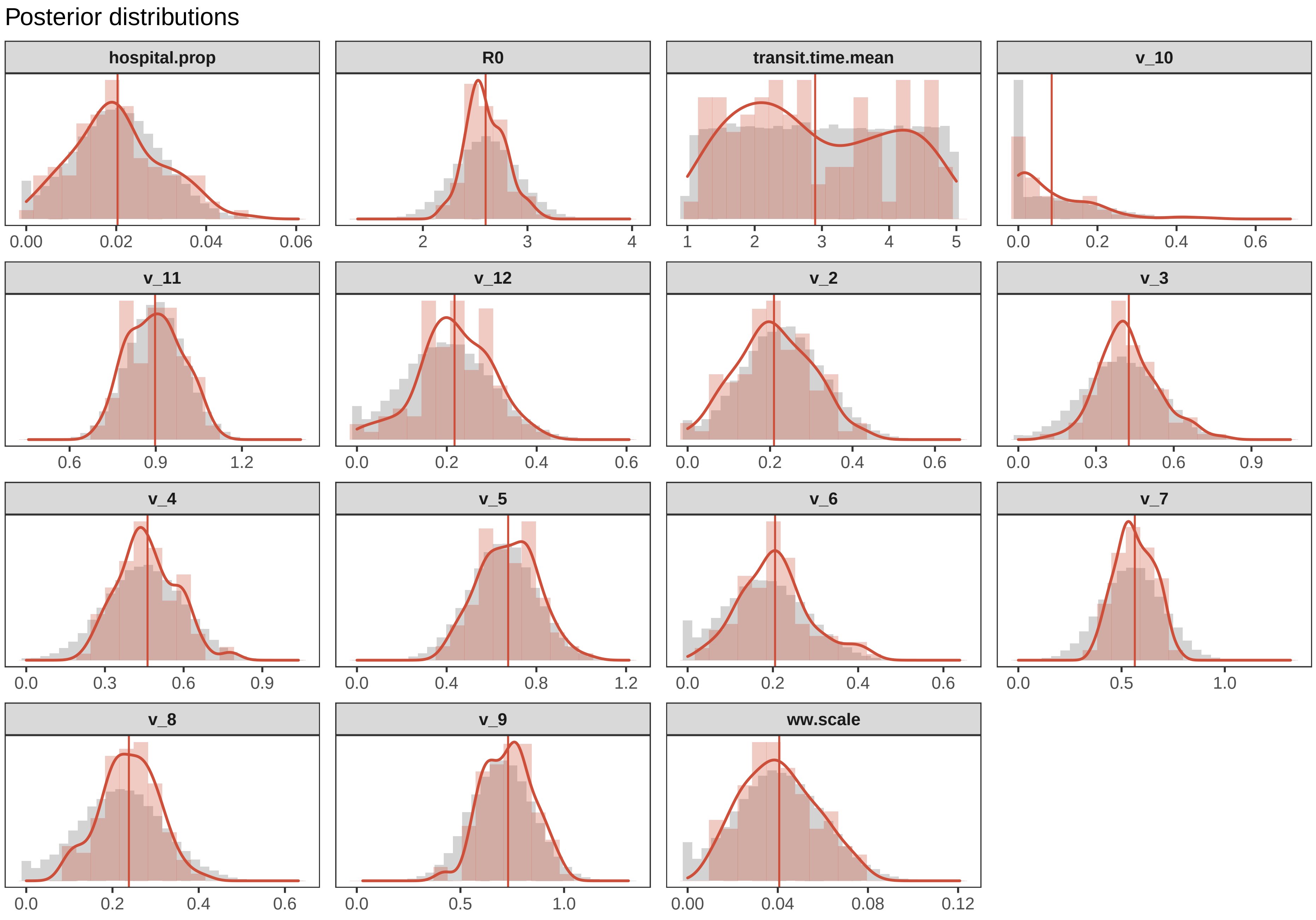

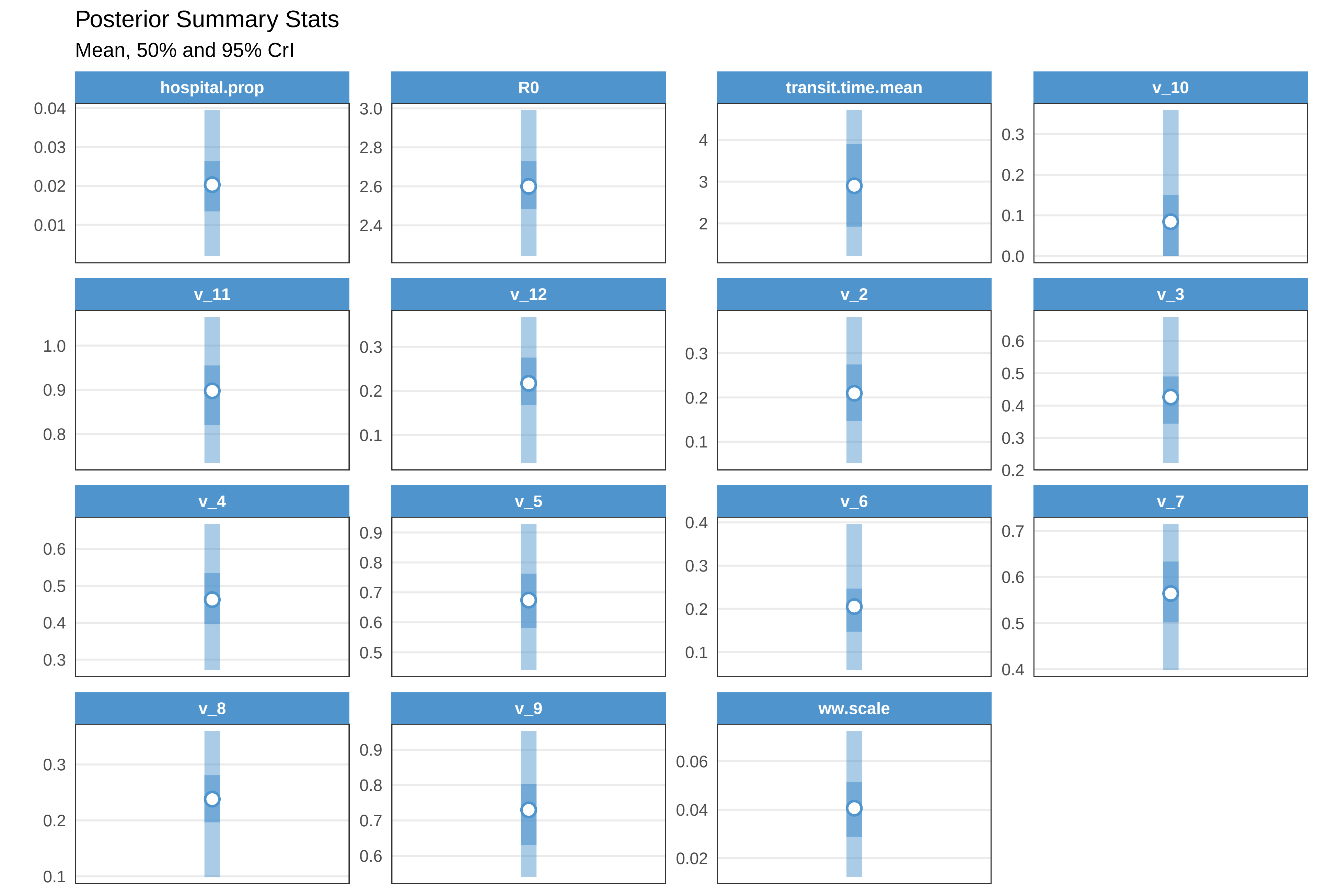

Check fit OTW

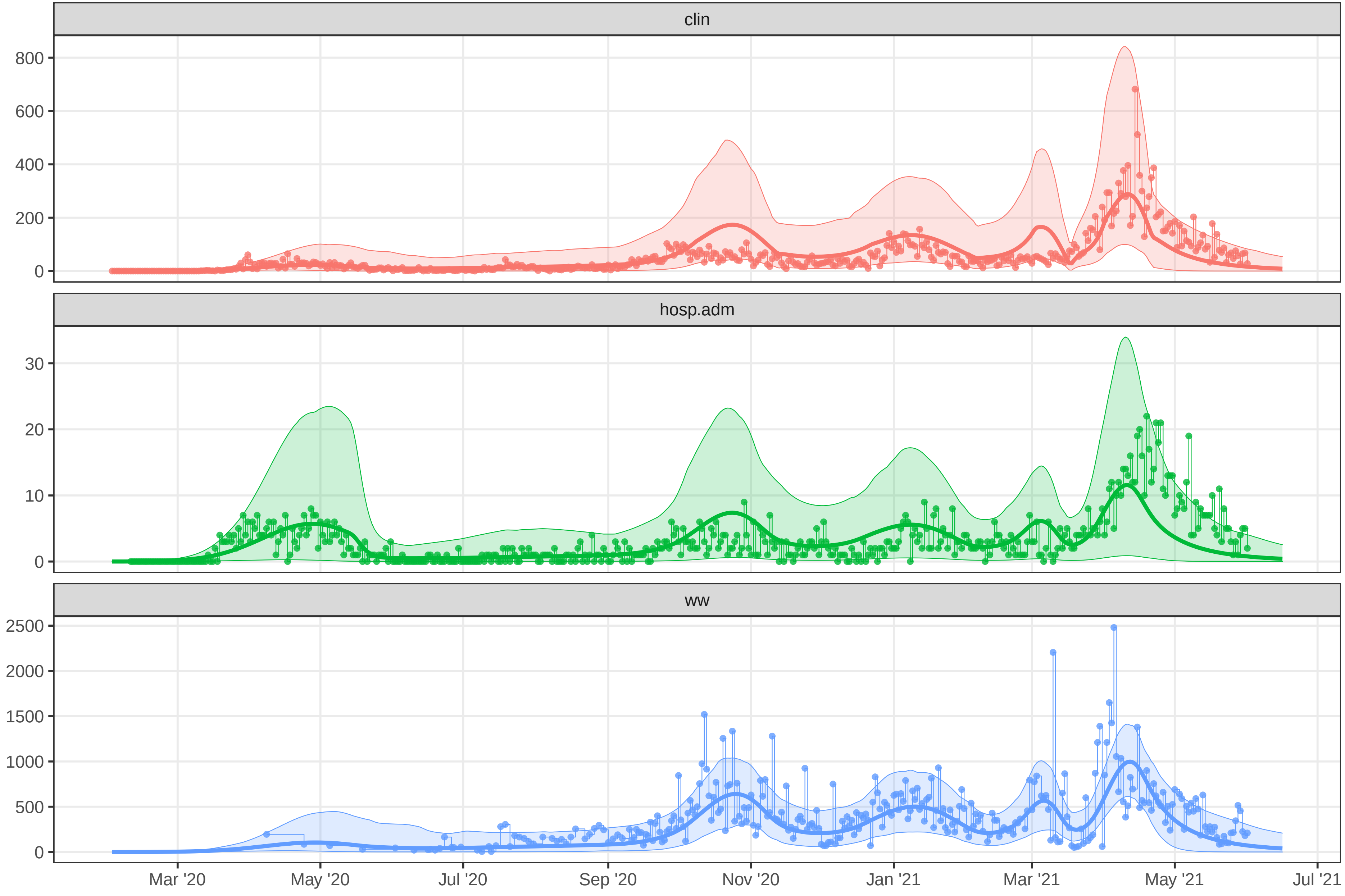

### plot-fit-OTW-07-09-0959-clinww.pdf

##### Observed data for OTW

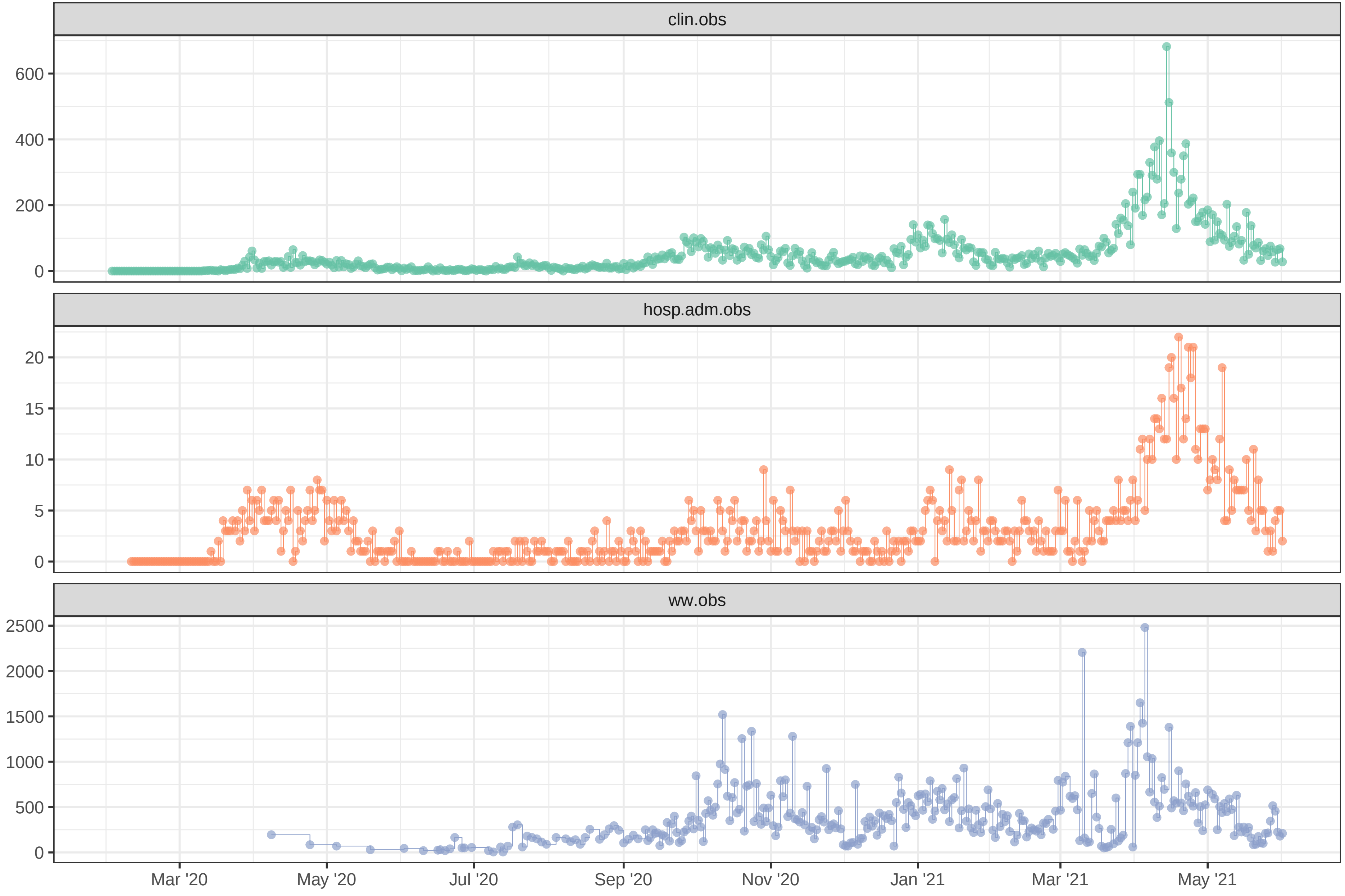

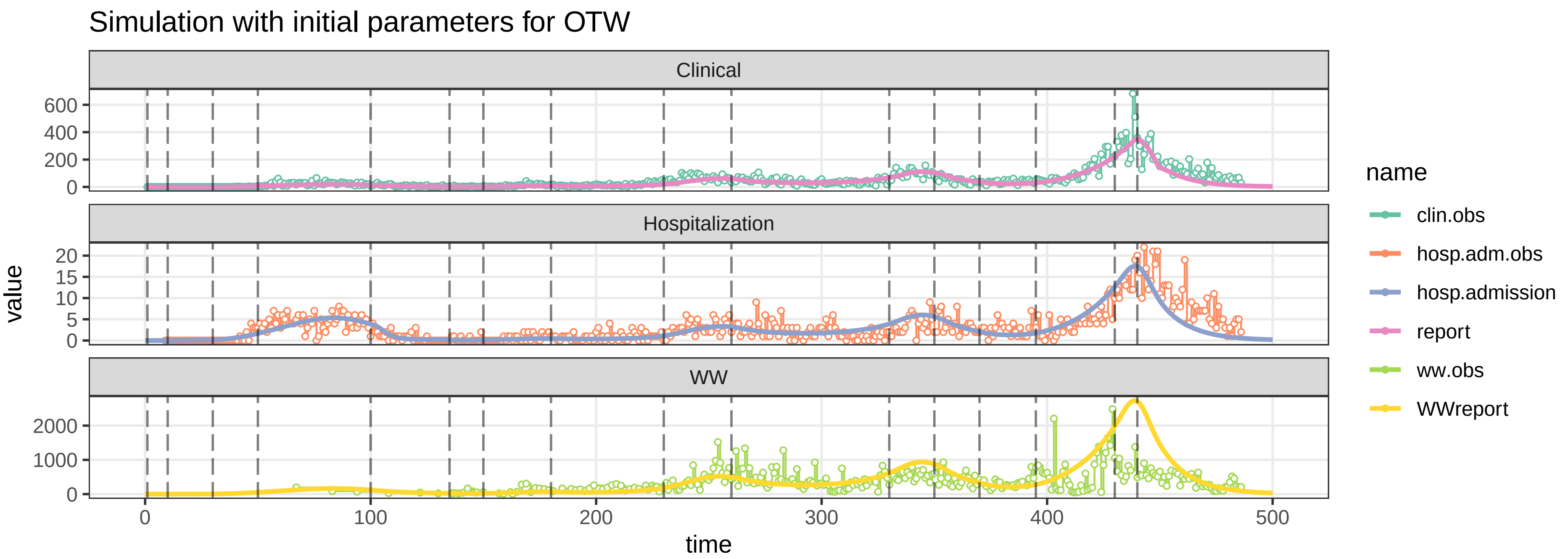

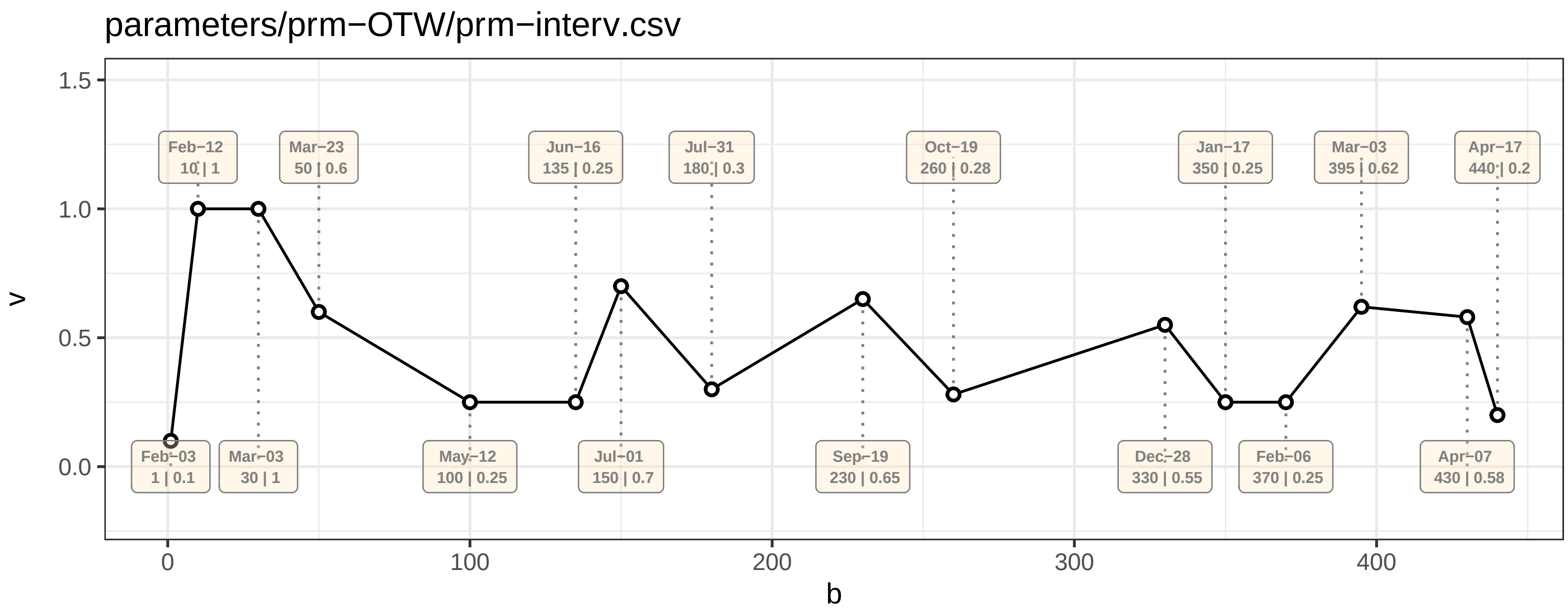

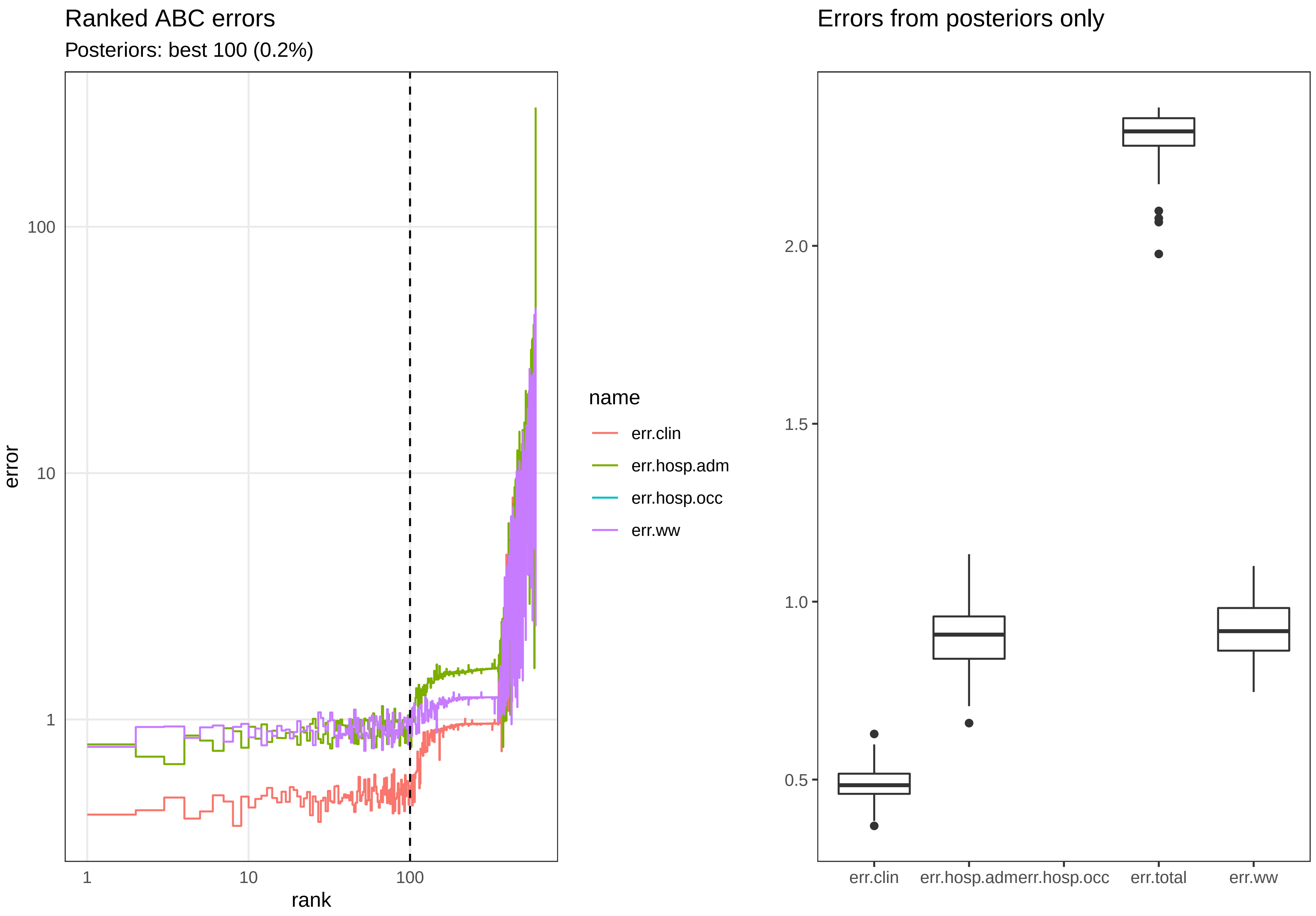

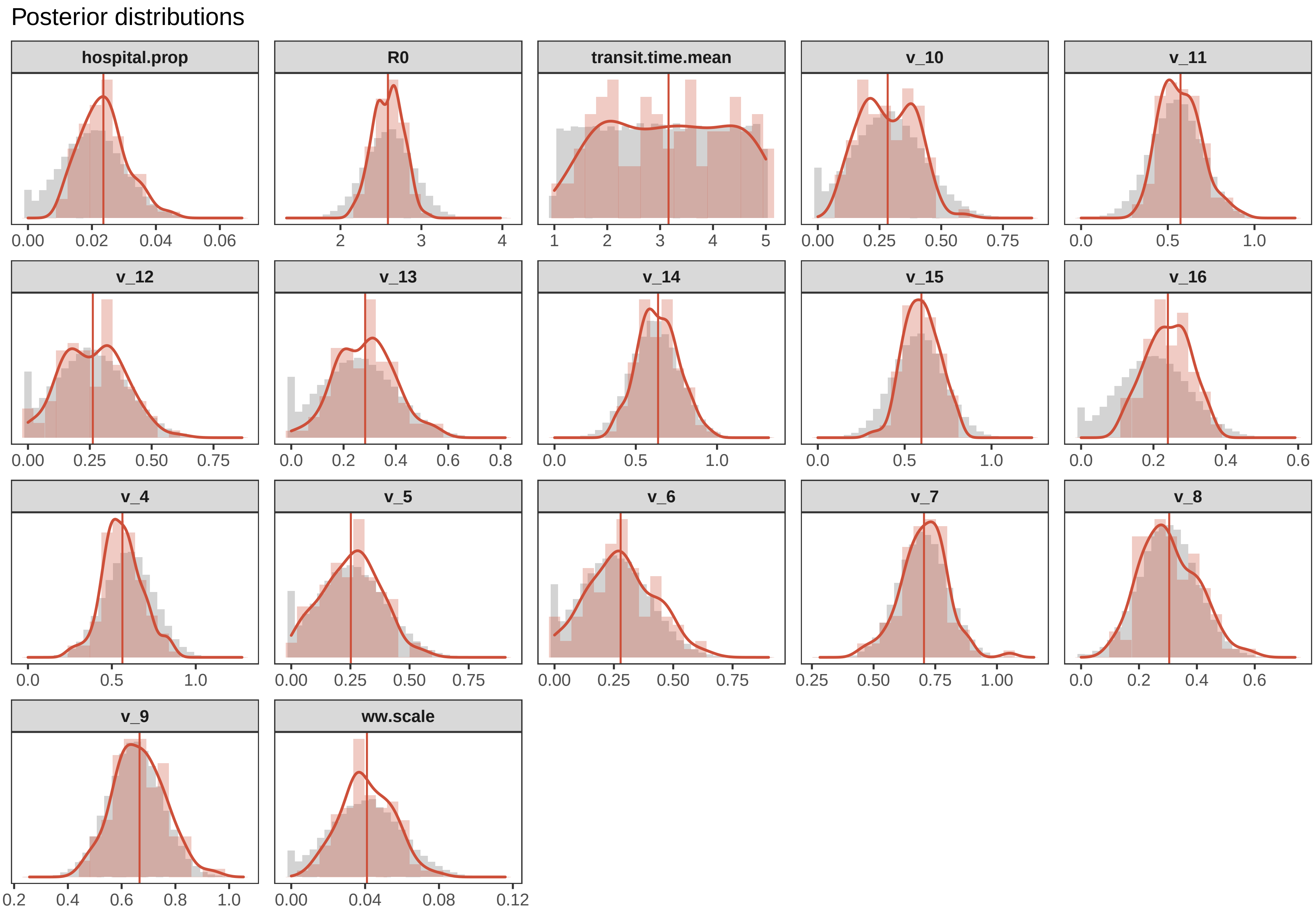

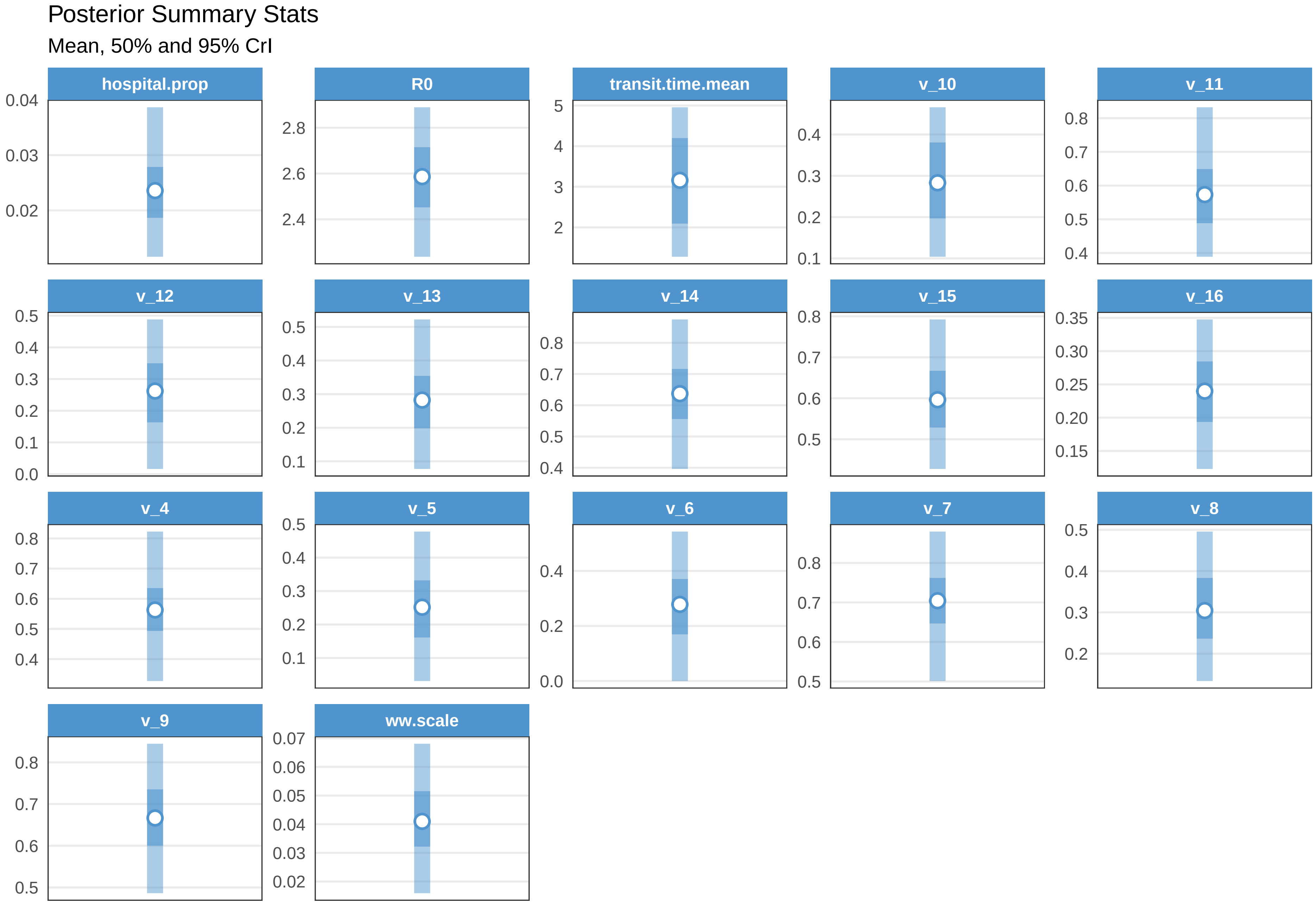

#### Check fit OTW

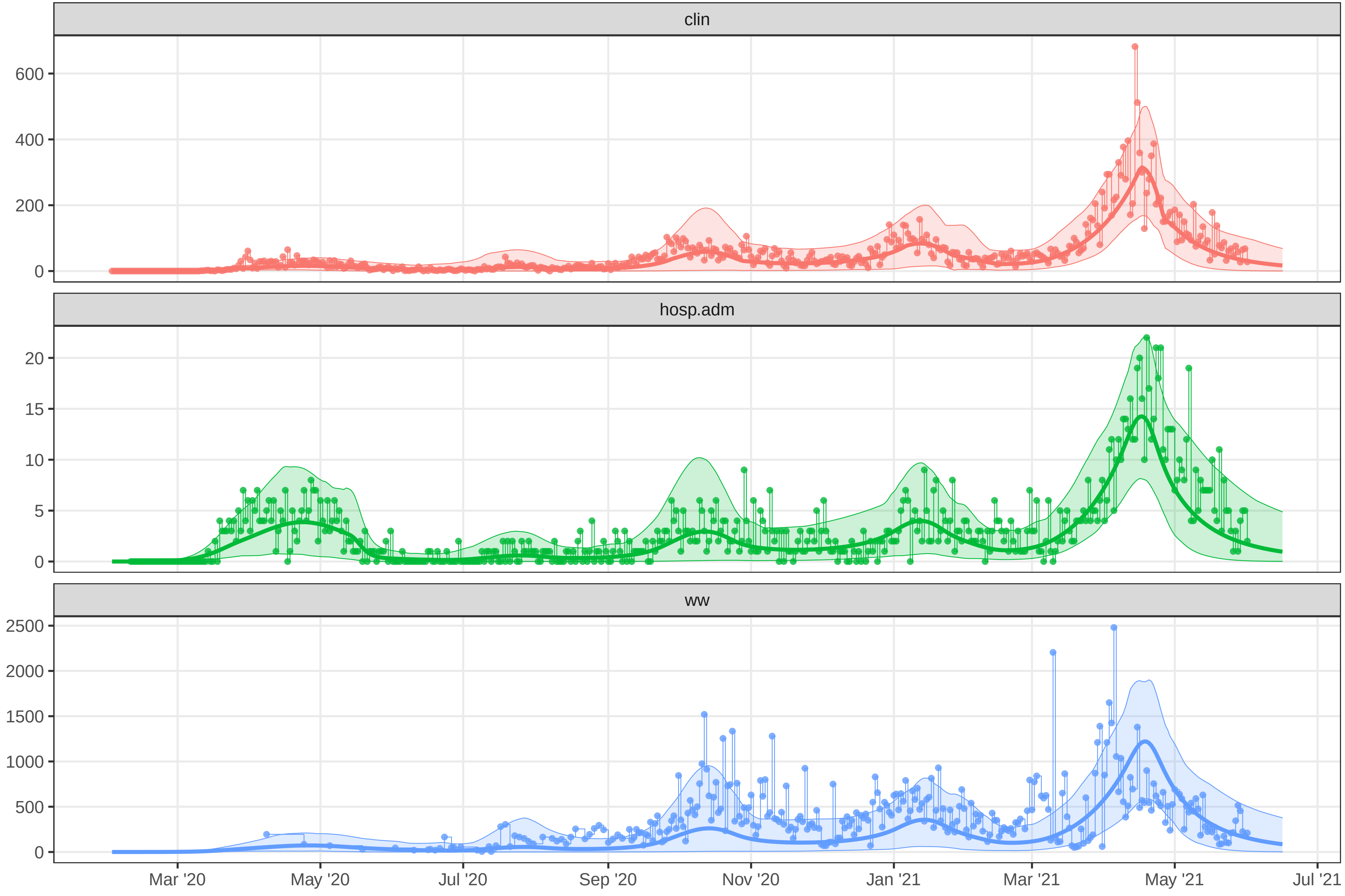

### plot-fit-TAB-07-06-clinww.pdf

Observed data for TAB

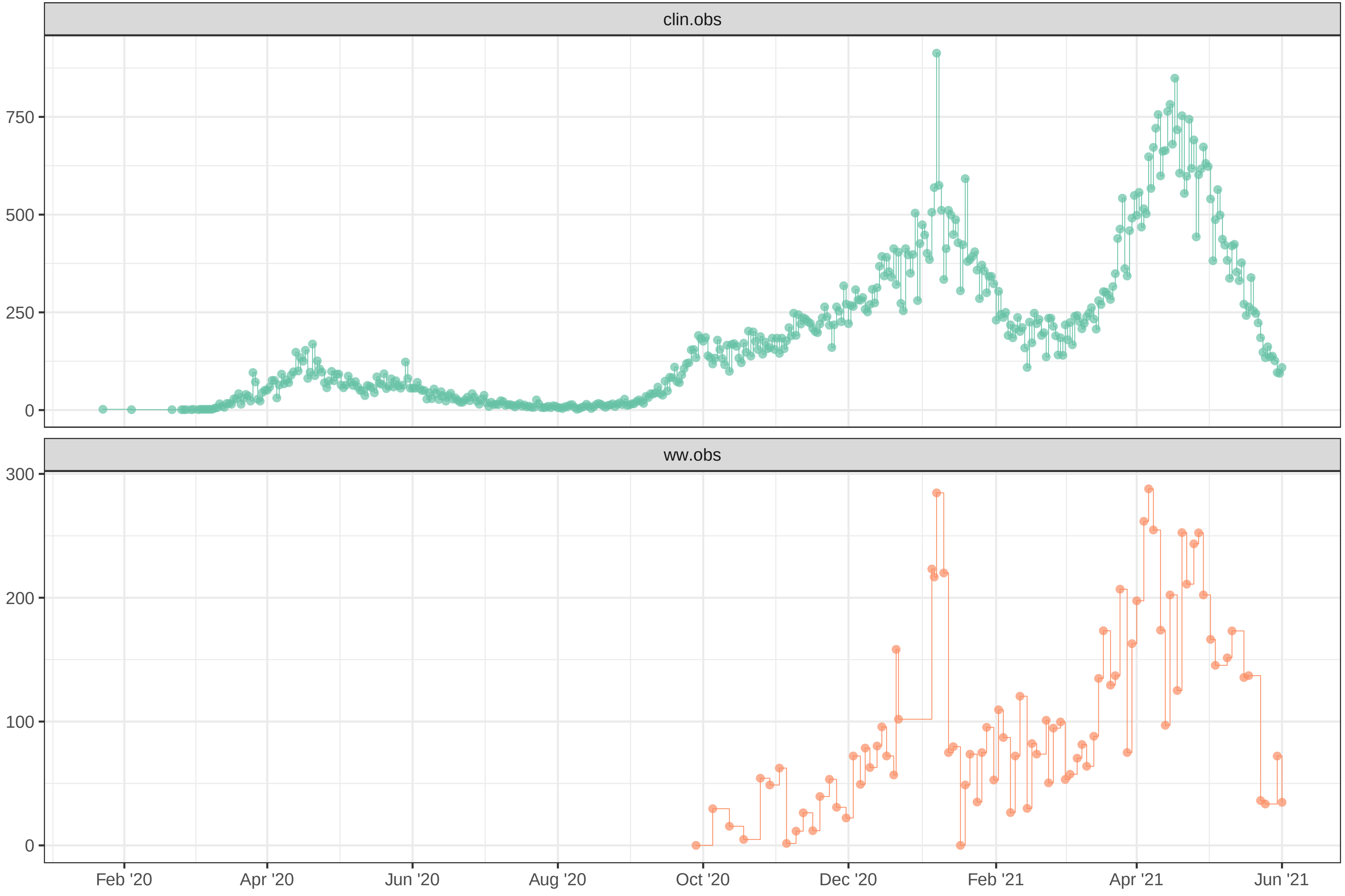

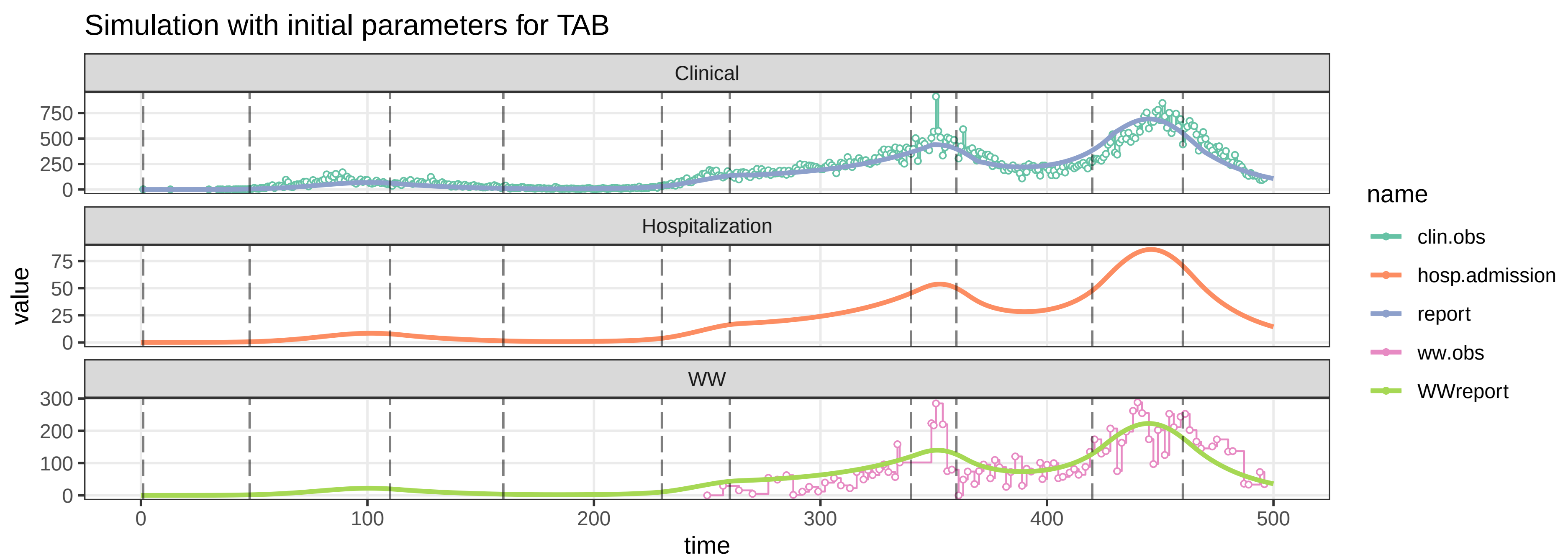

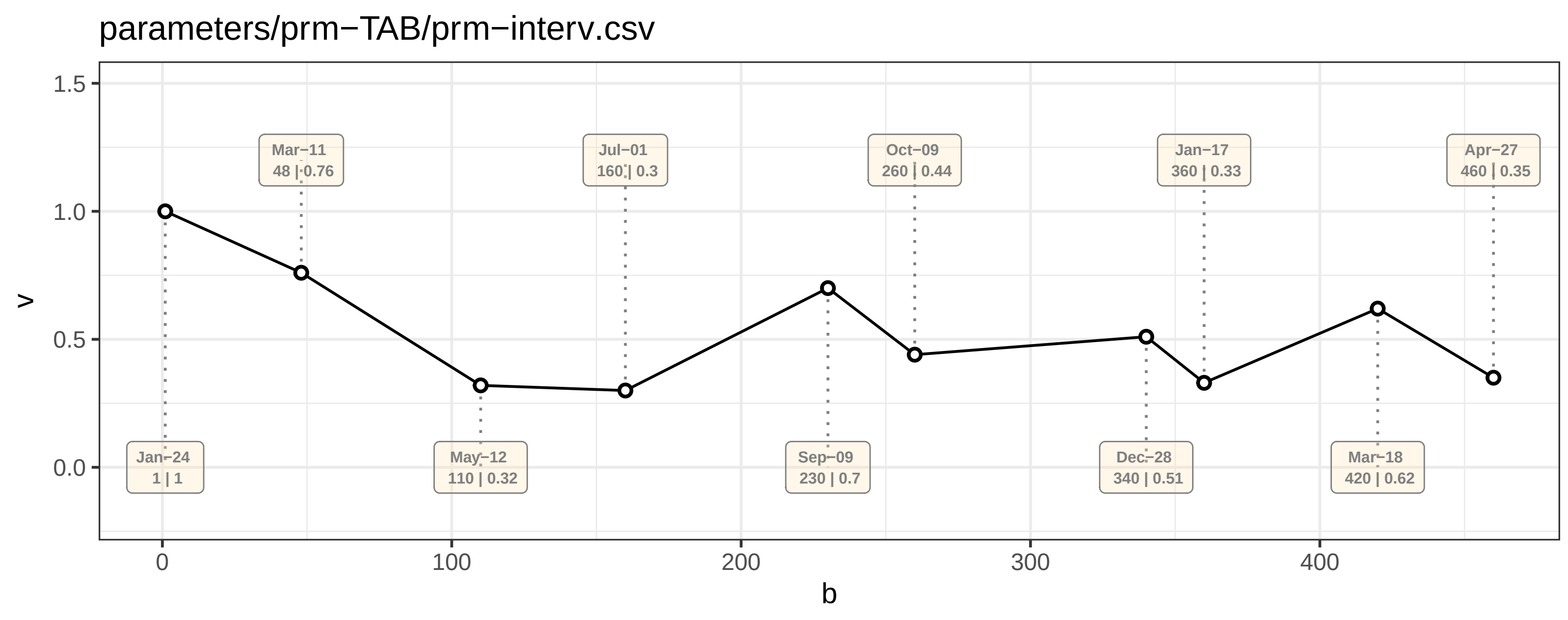

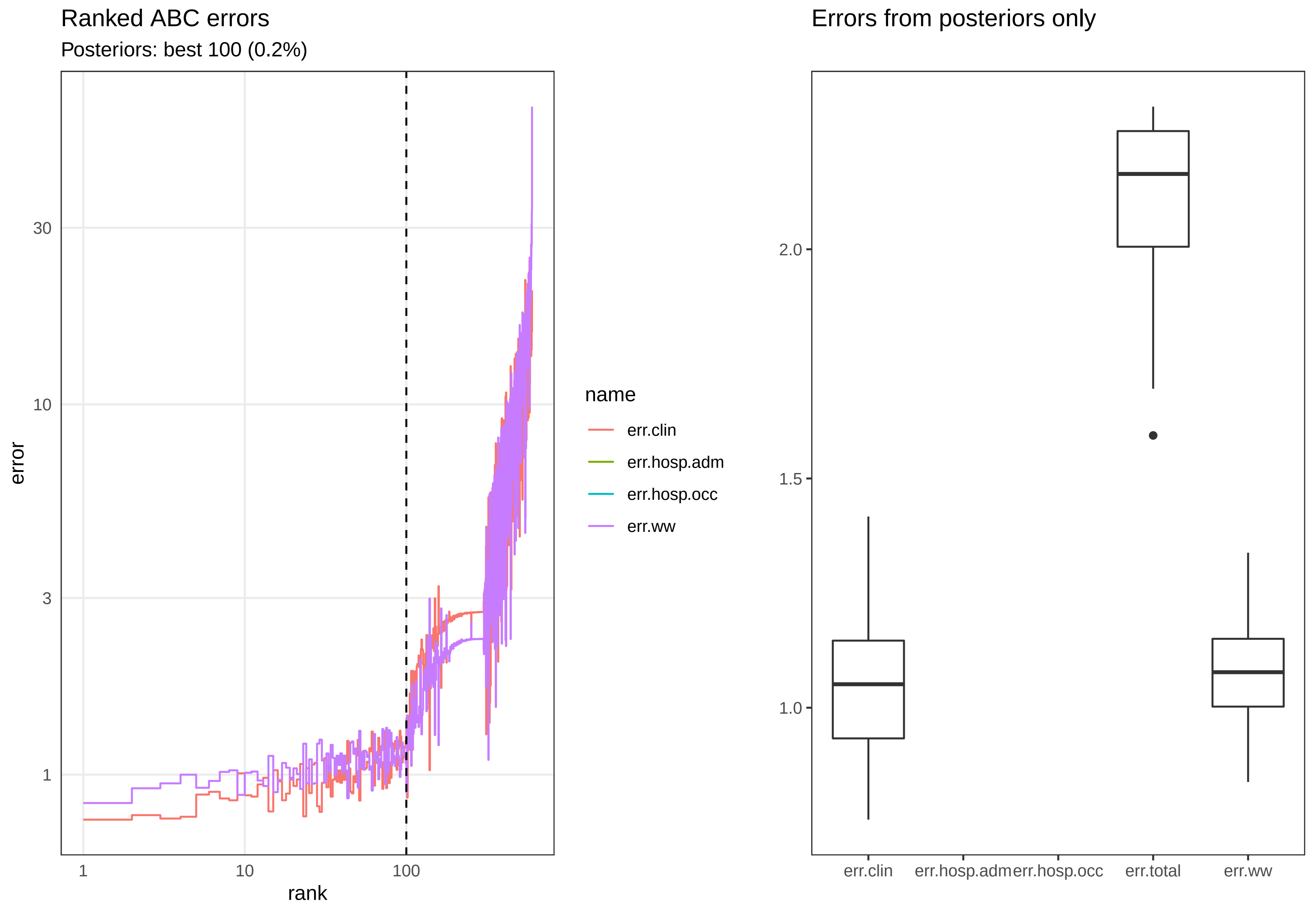

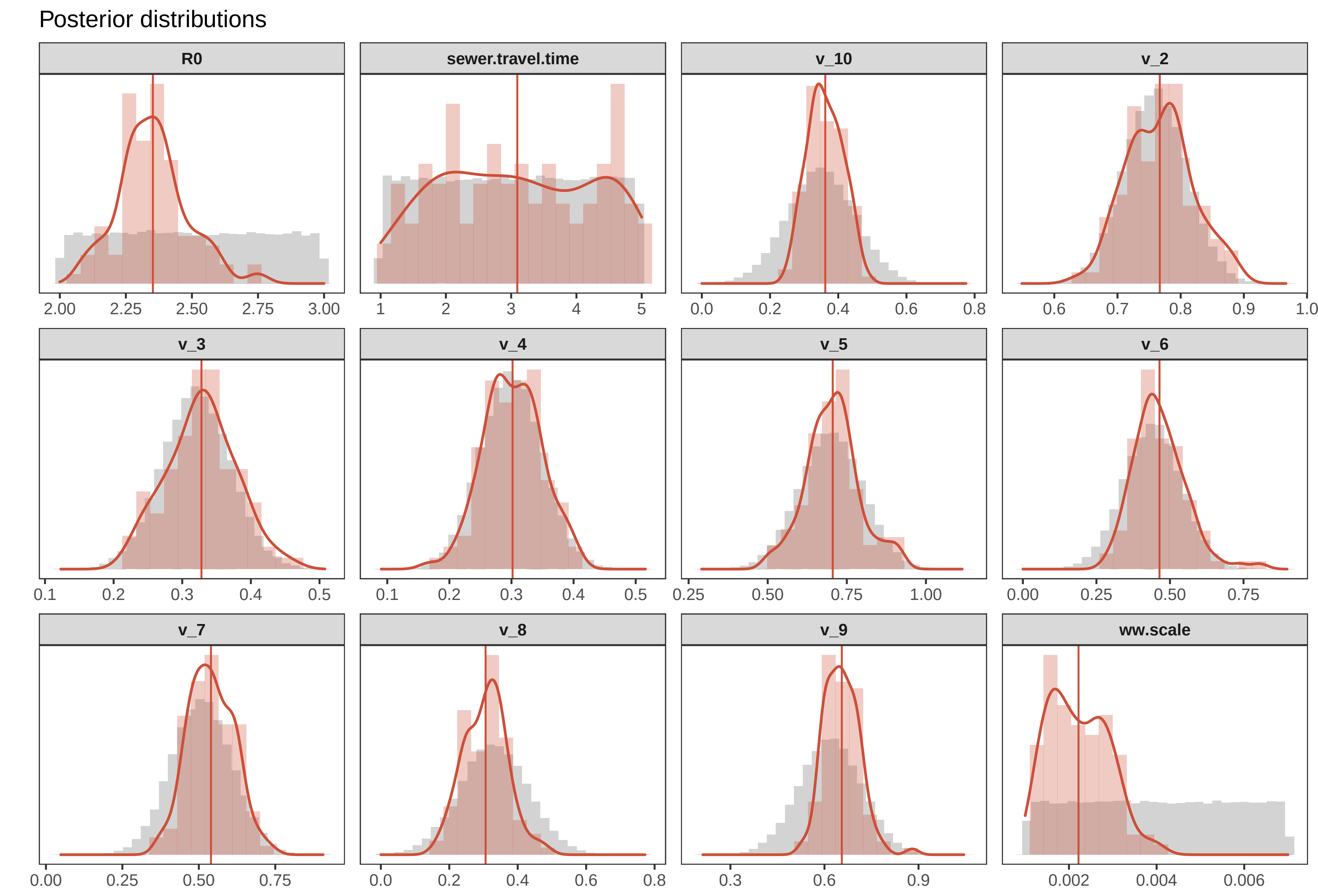

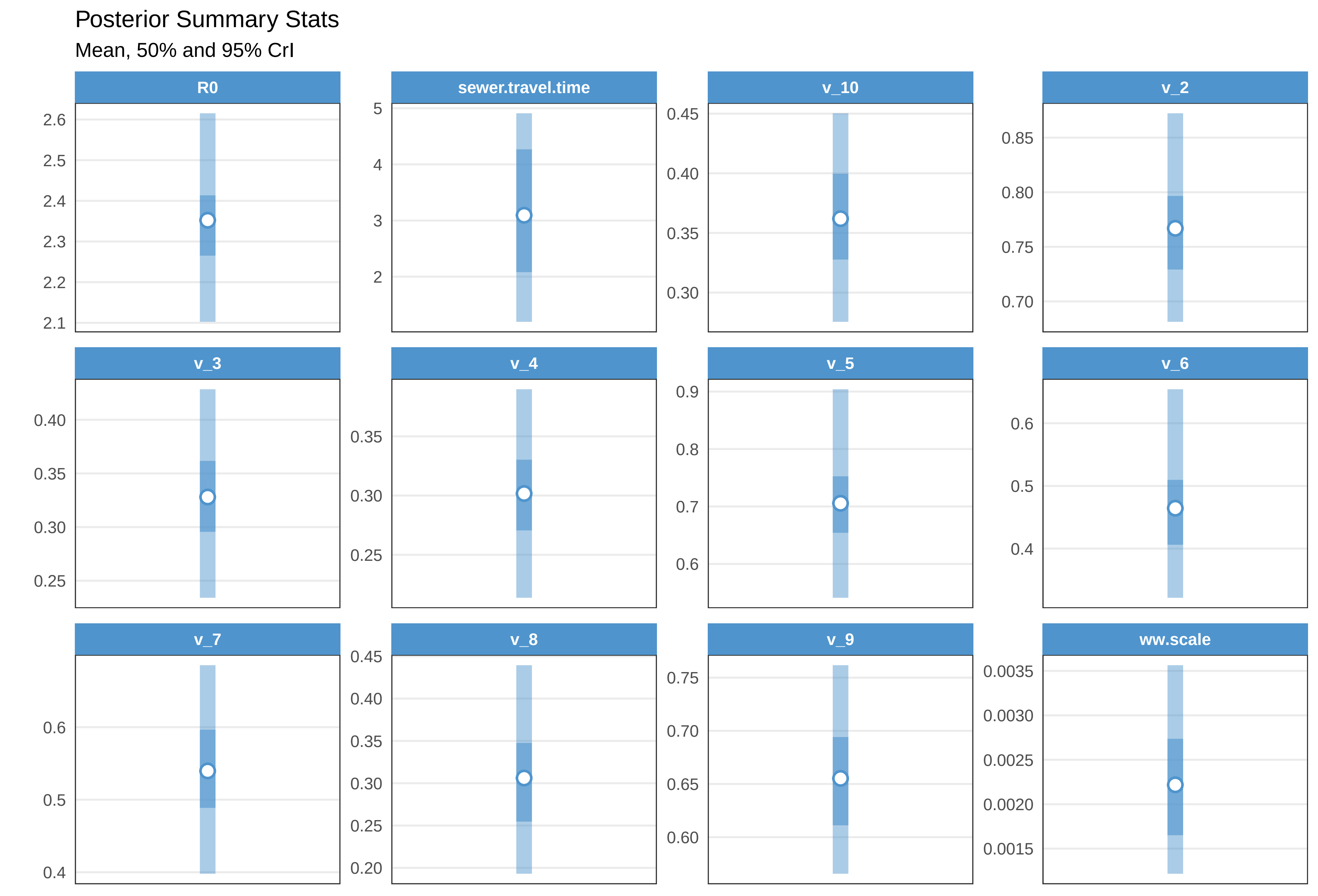

Check fit TAB

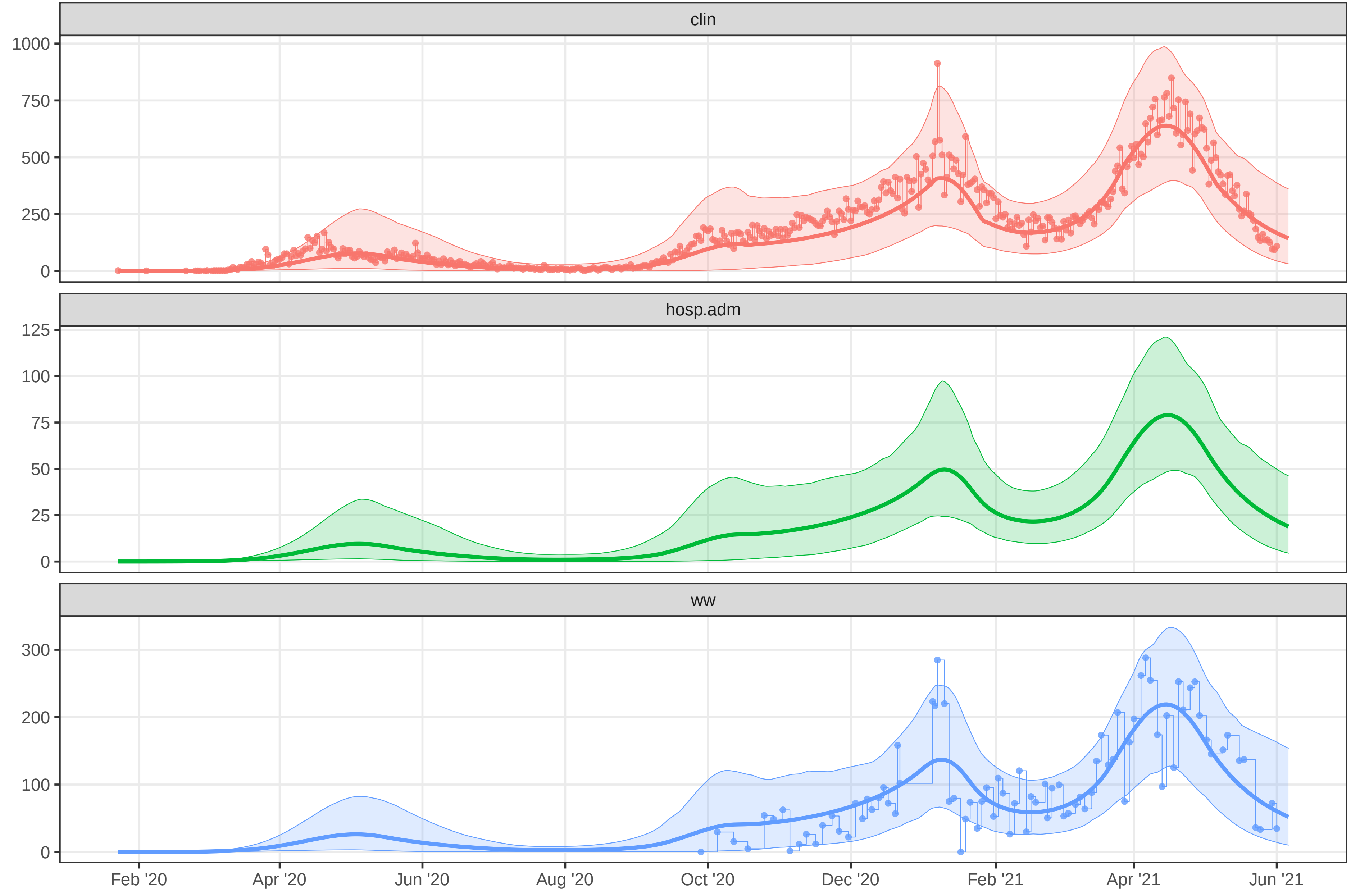

### plot-fit-TAB-07-06-ww.pdf

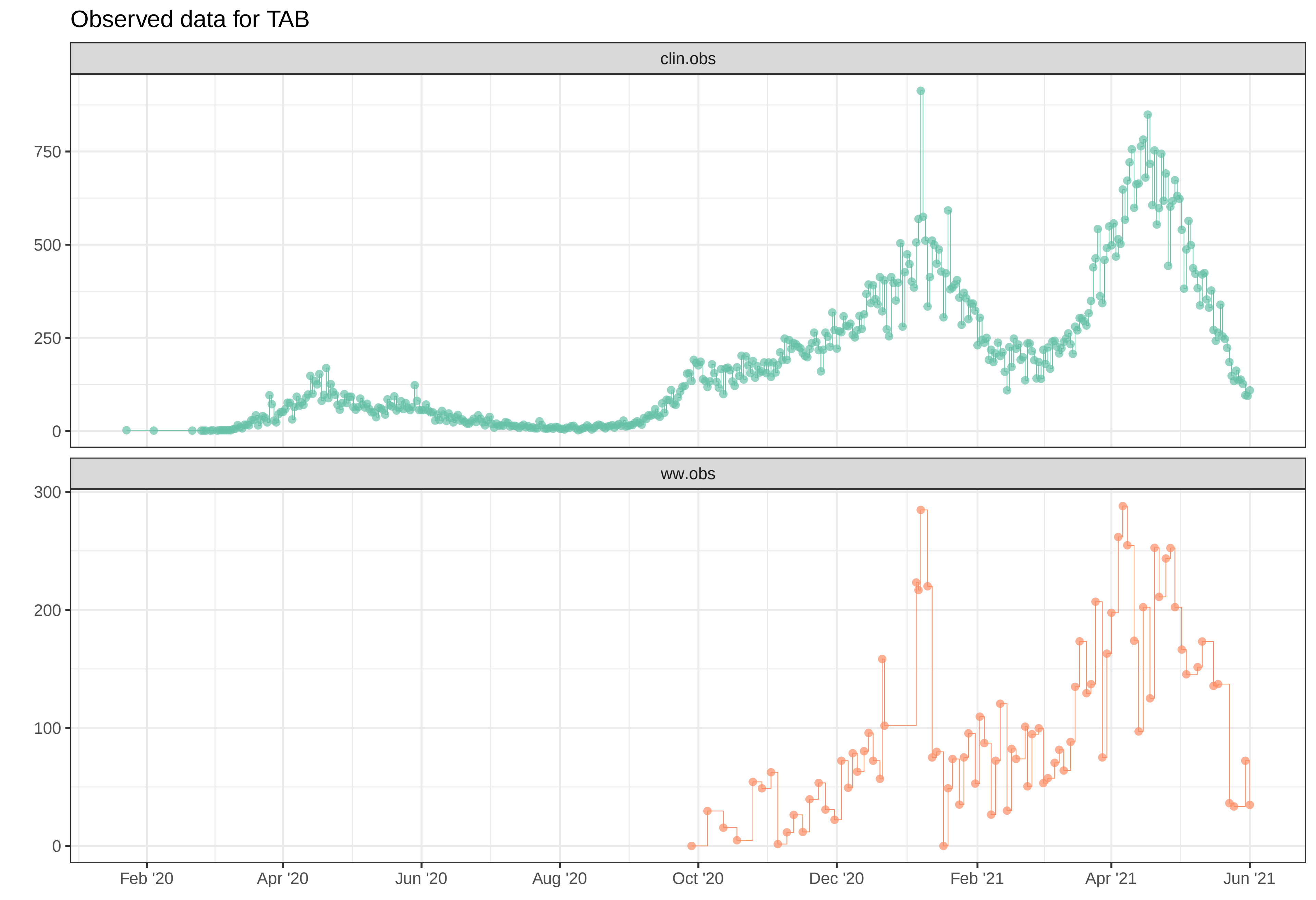

Simulation with initial parameters for TAB

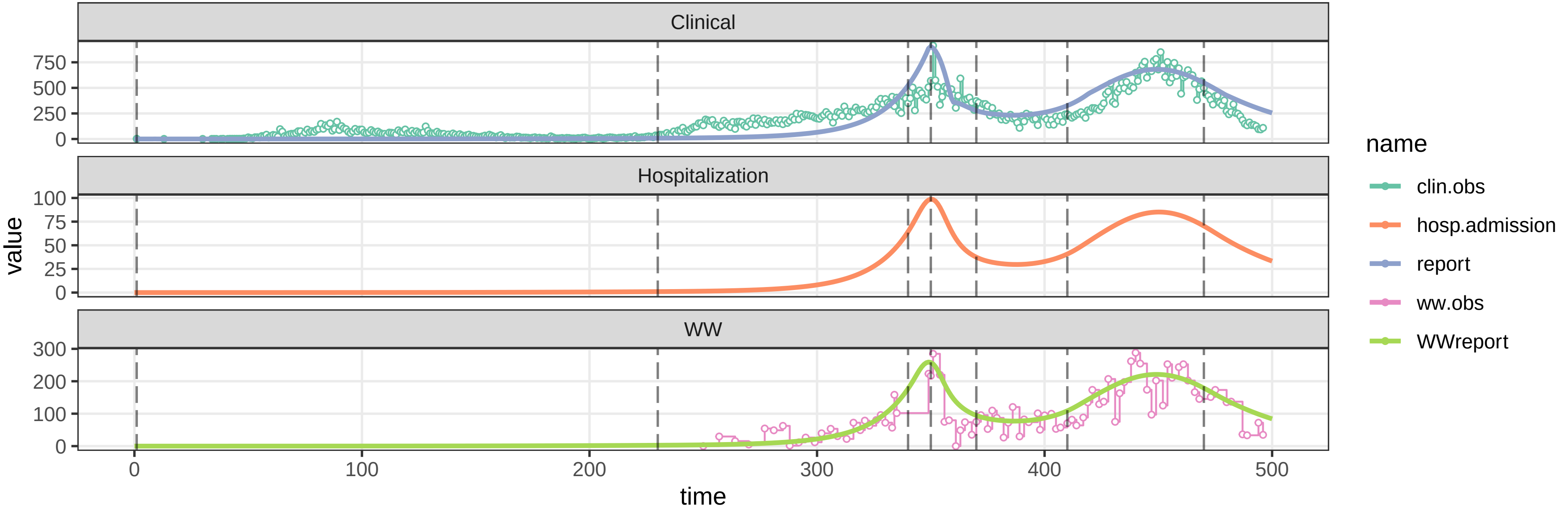


parameters/prm−TAB/prm−interv.csv

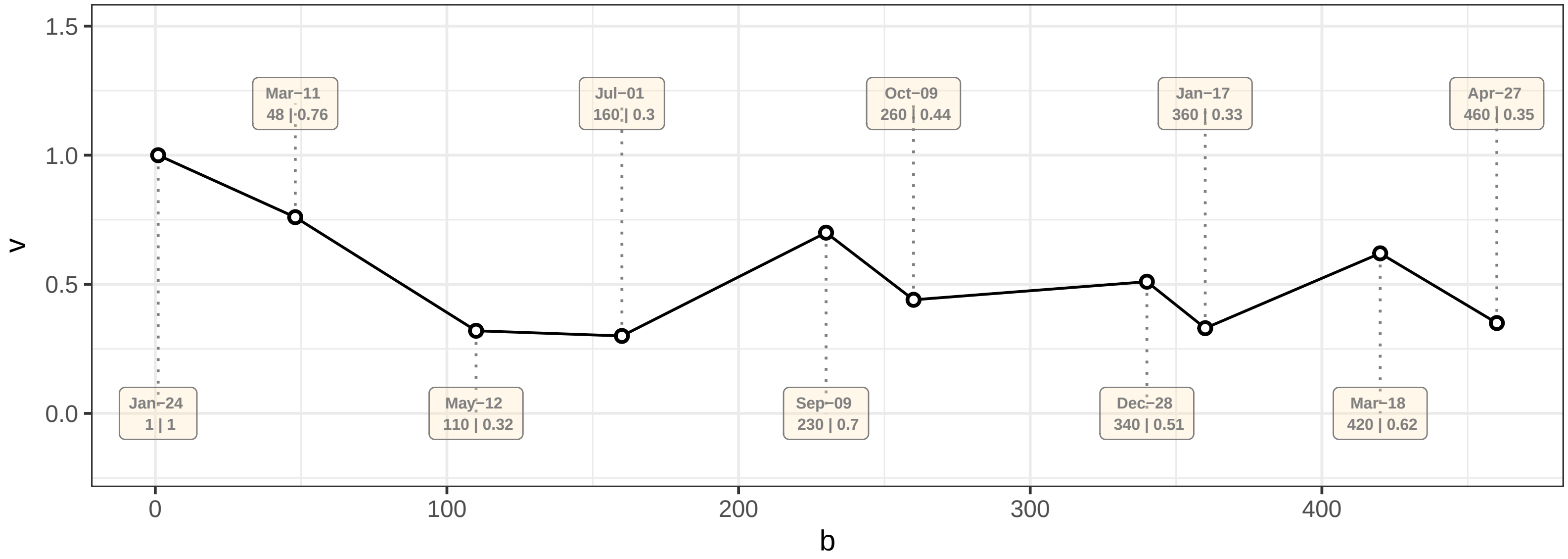

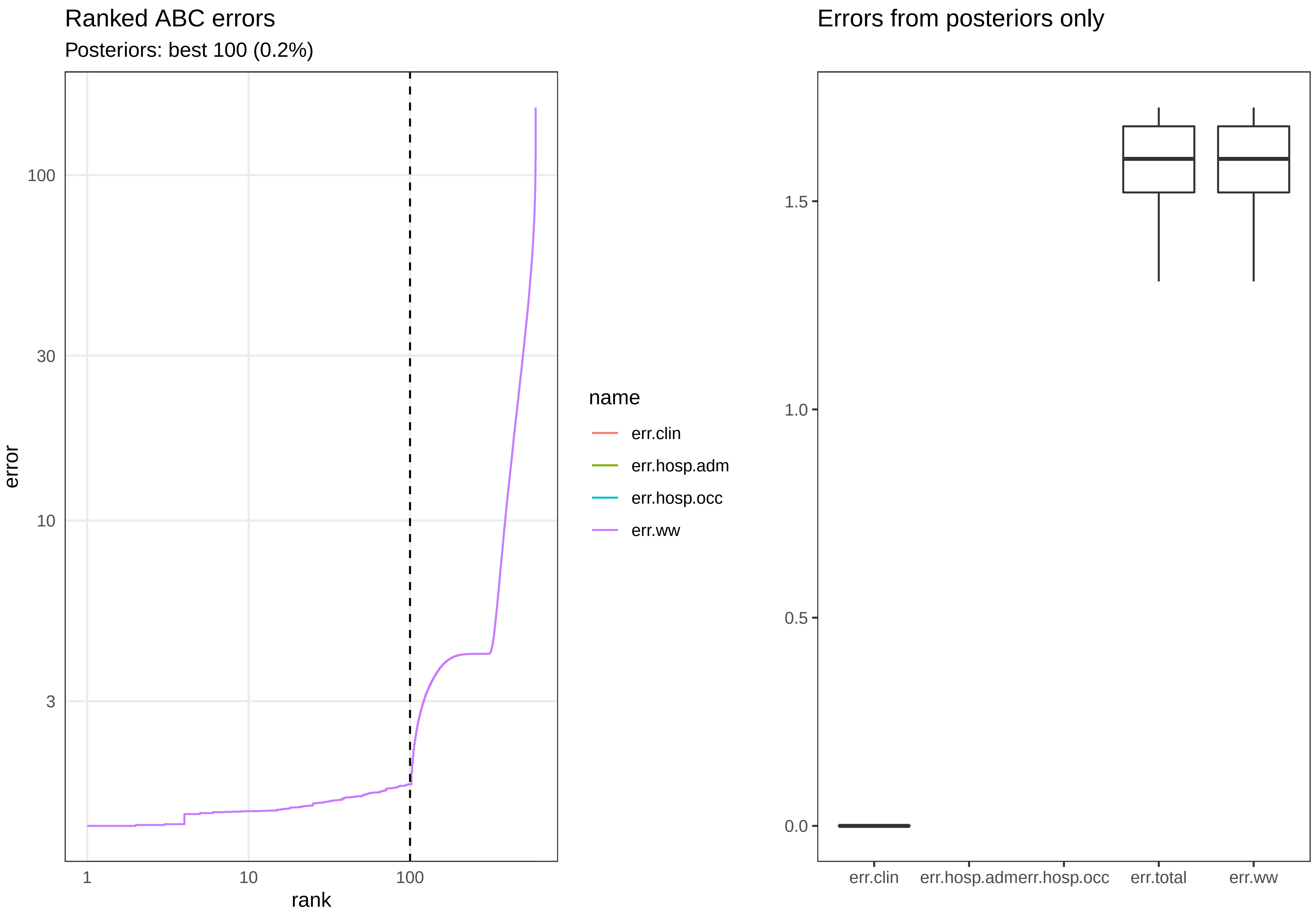

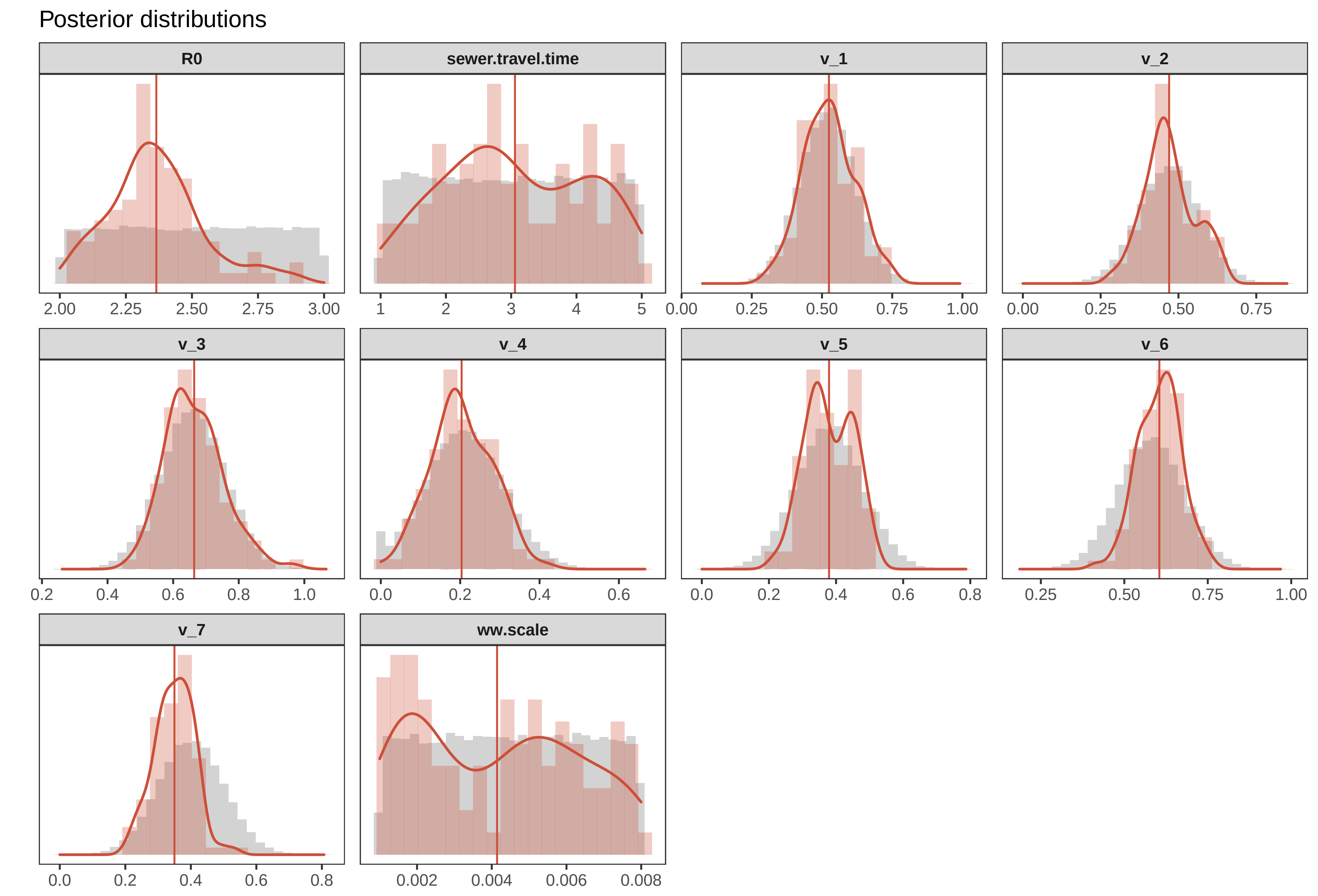

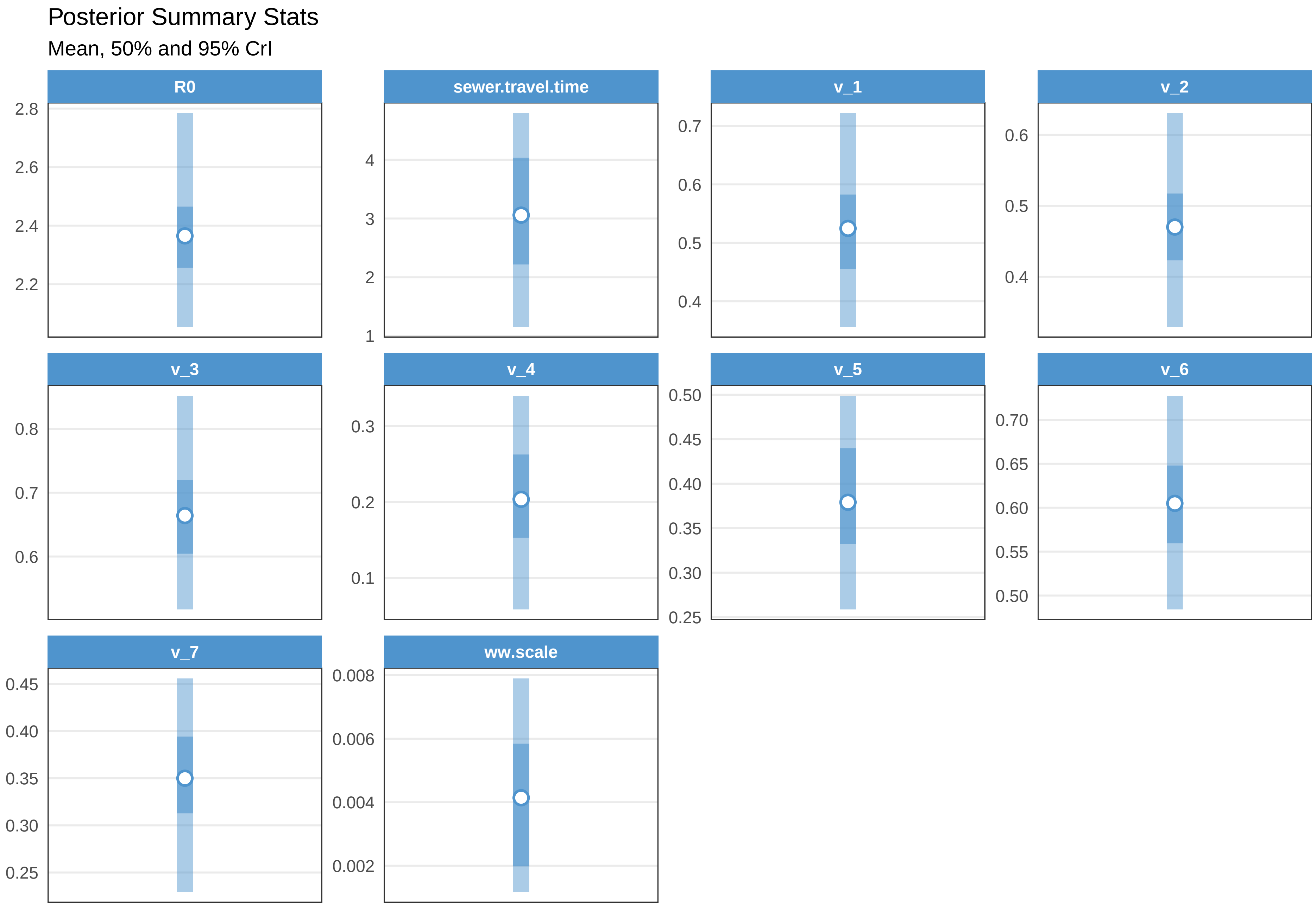

Check fit TAB

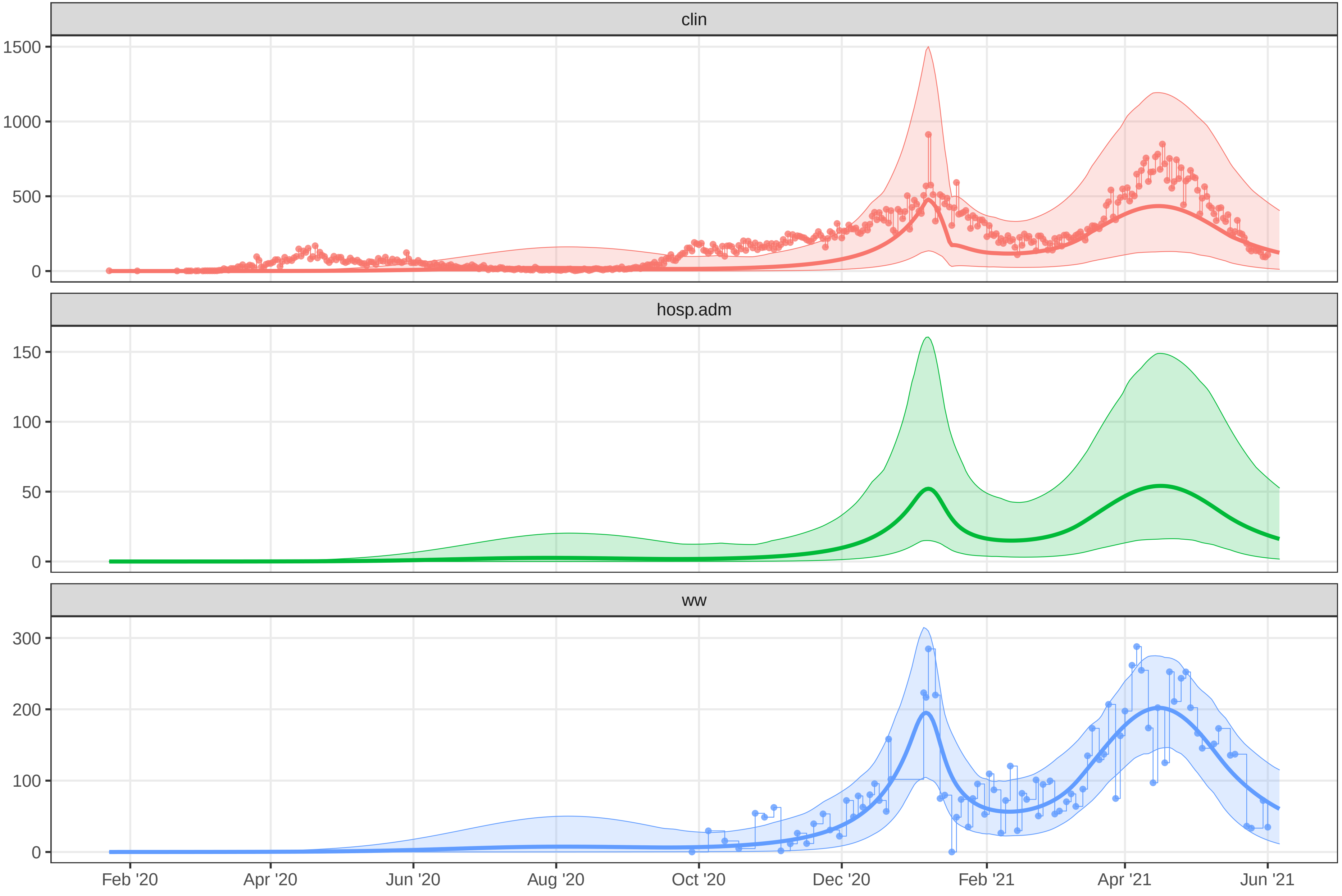

### plot-fit-THC-07-06-clin.pdf

Observed data for THC

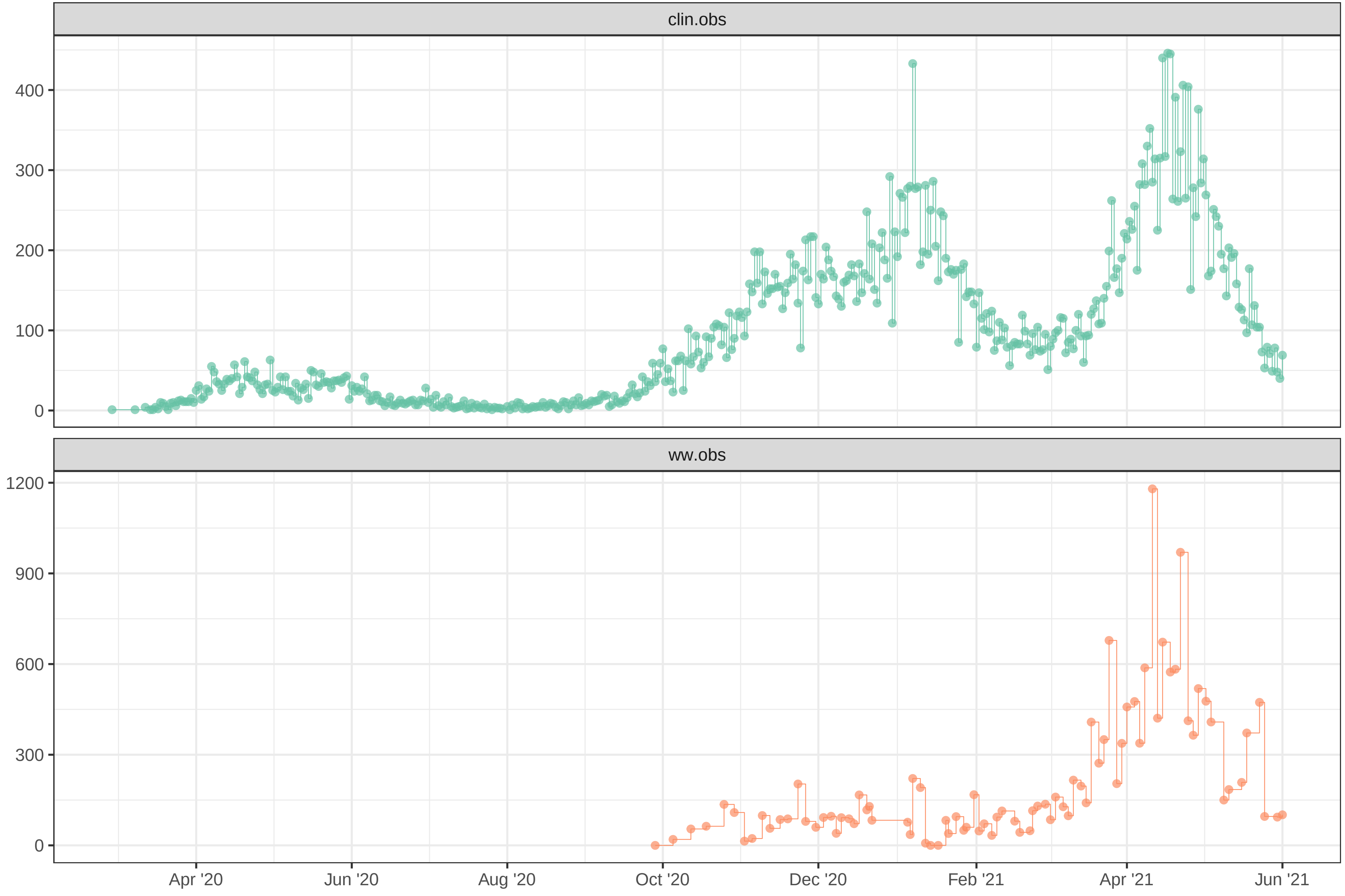

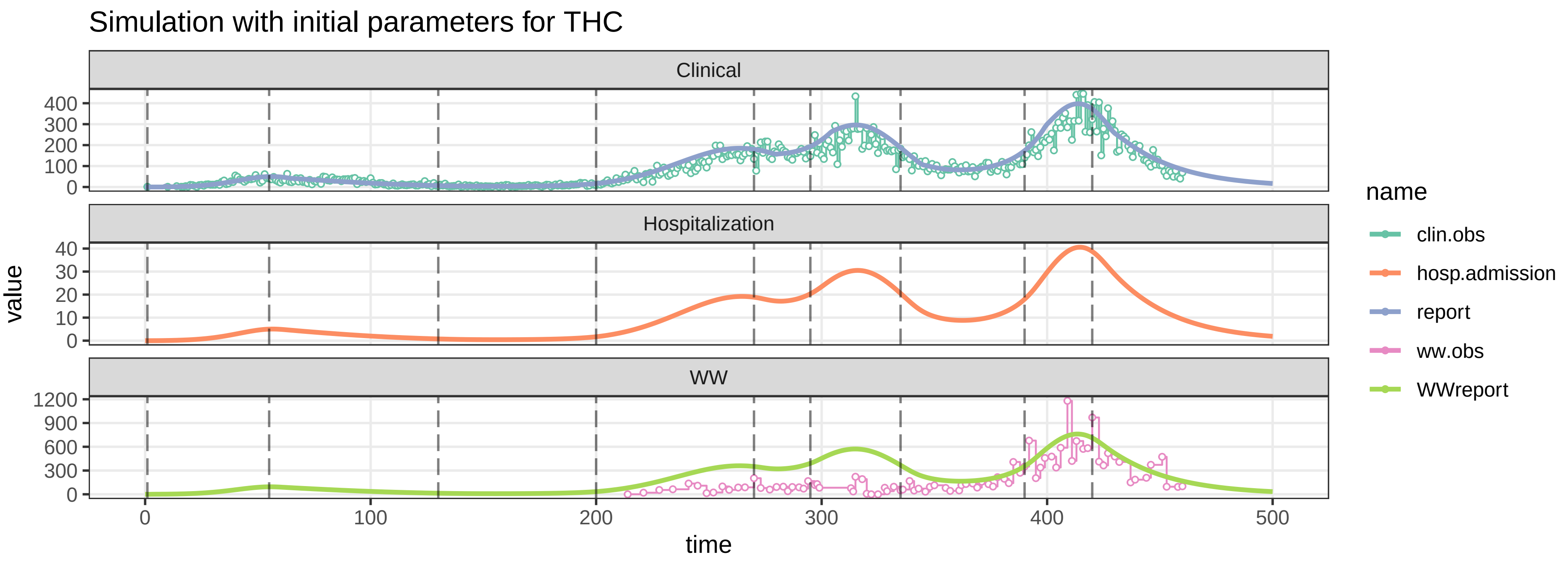

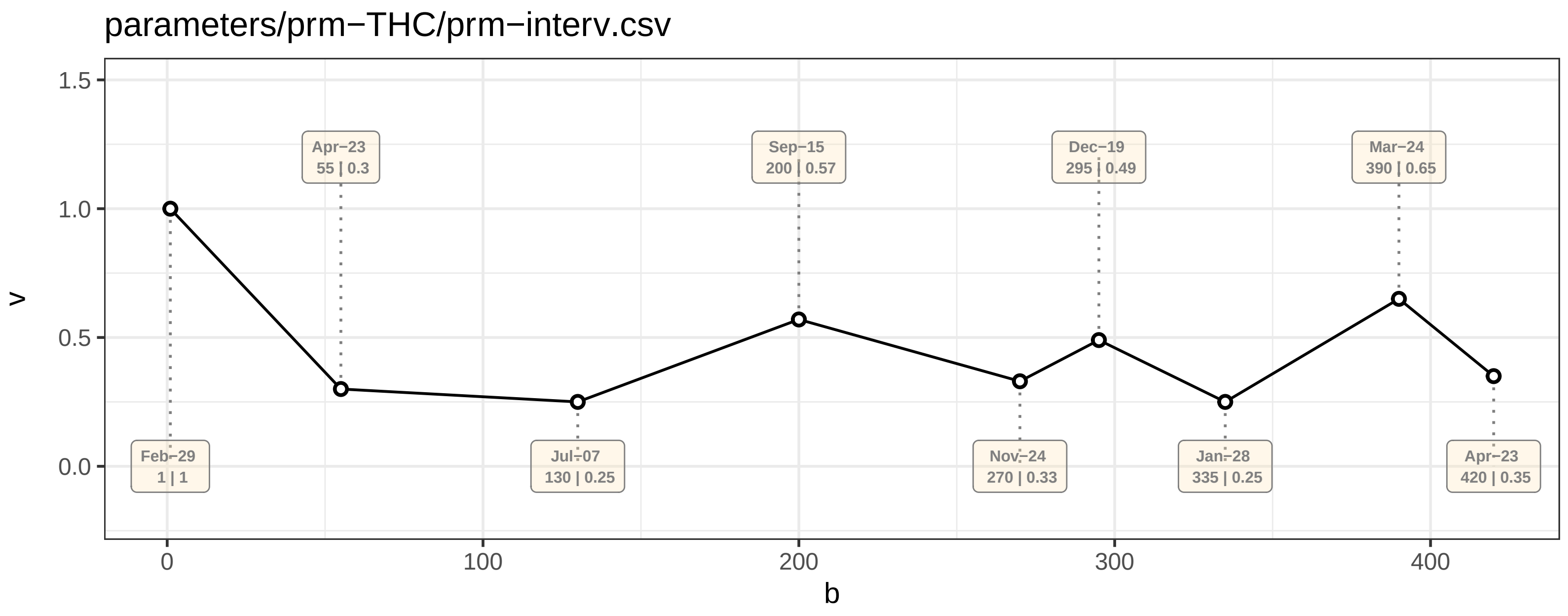

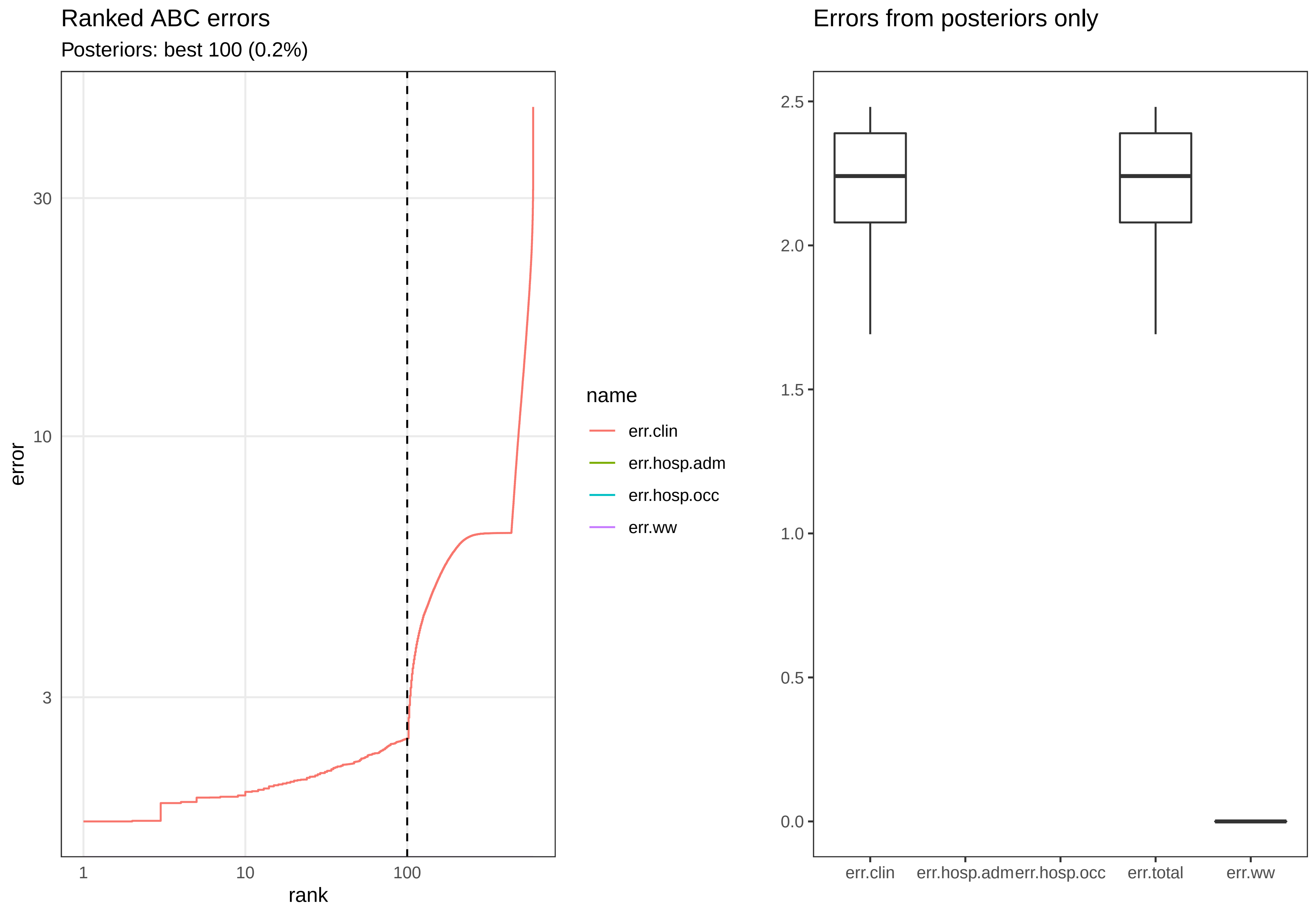

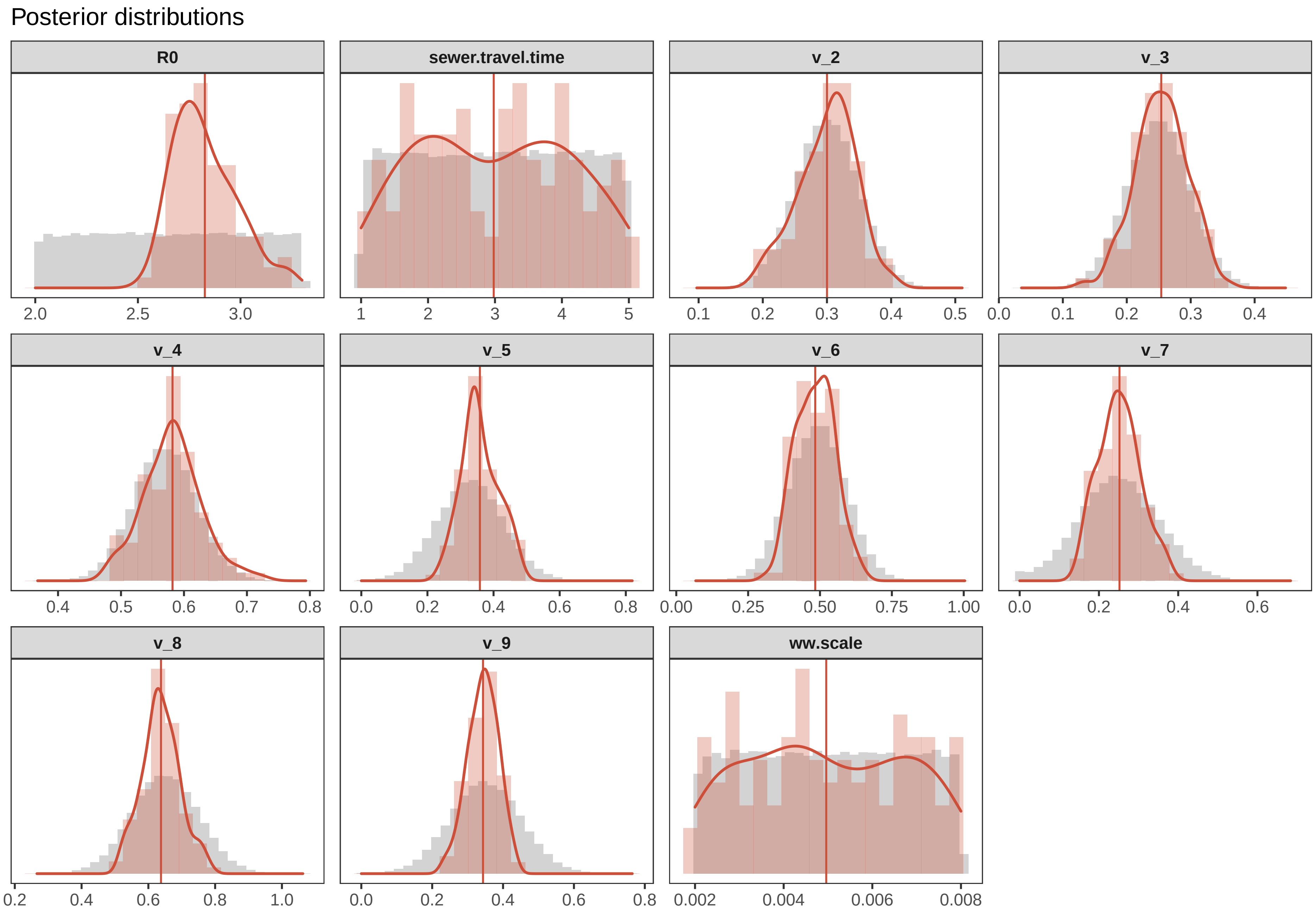

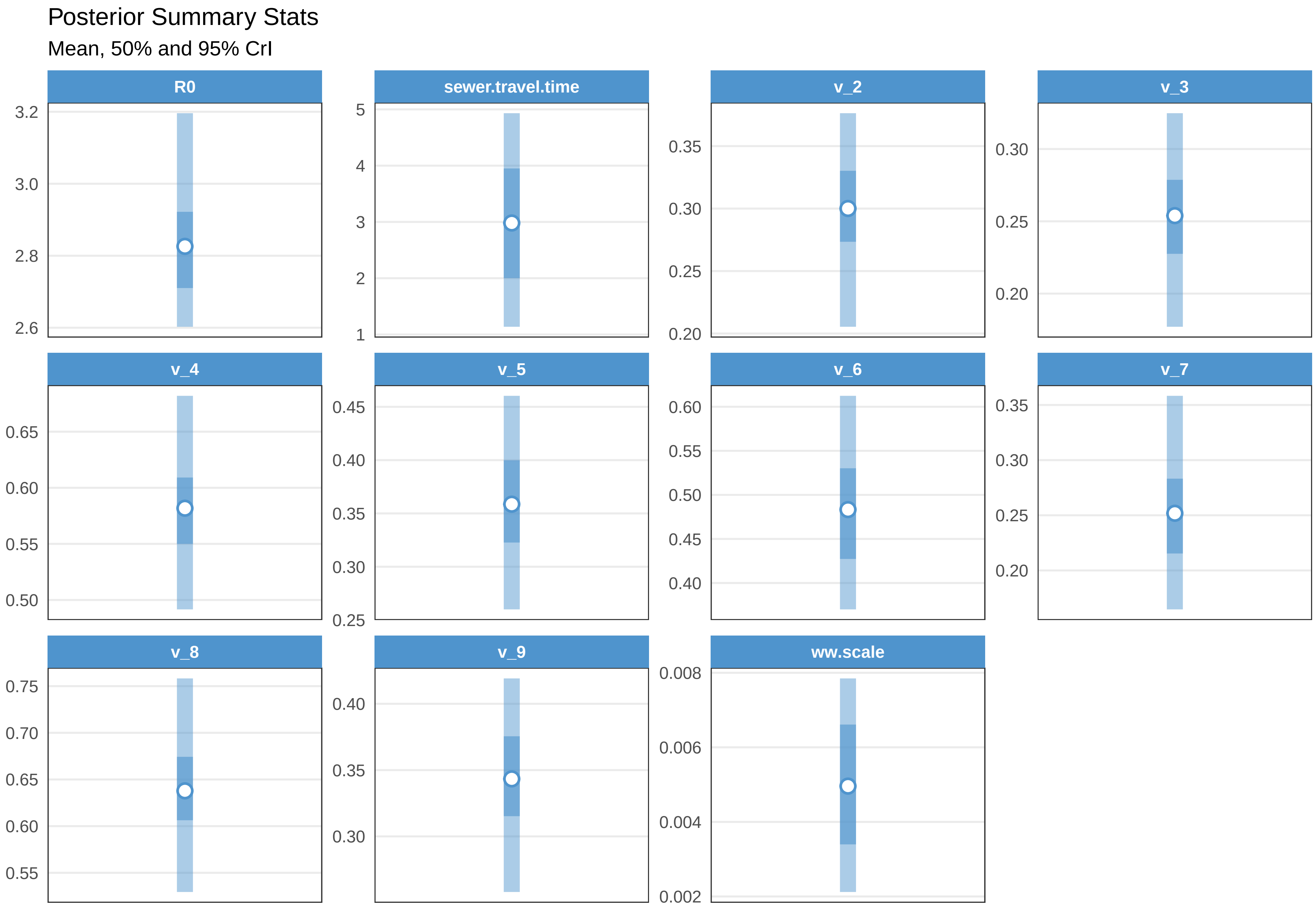

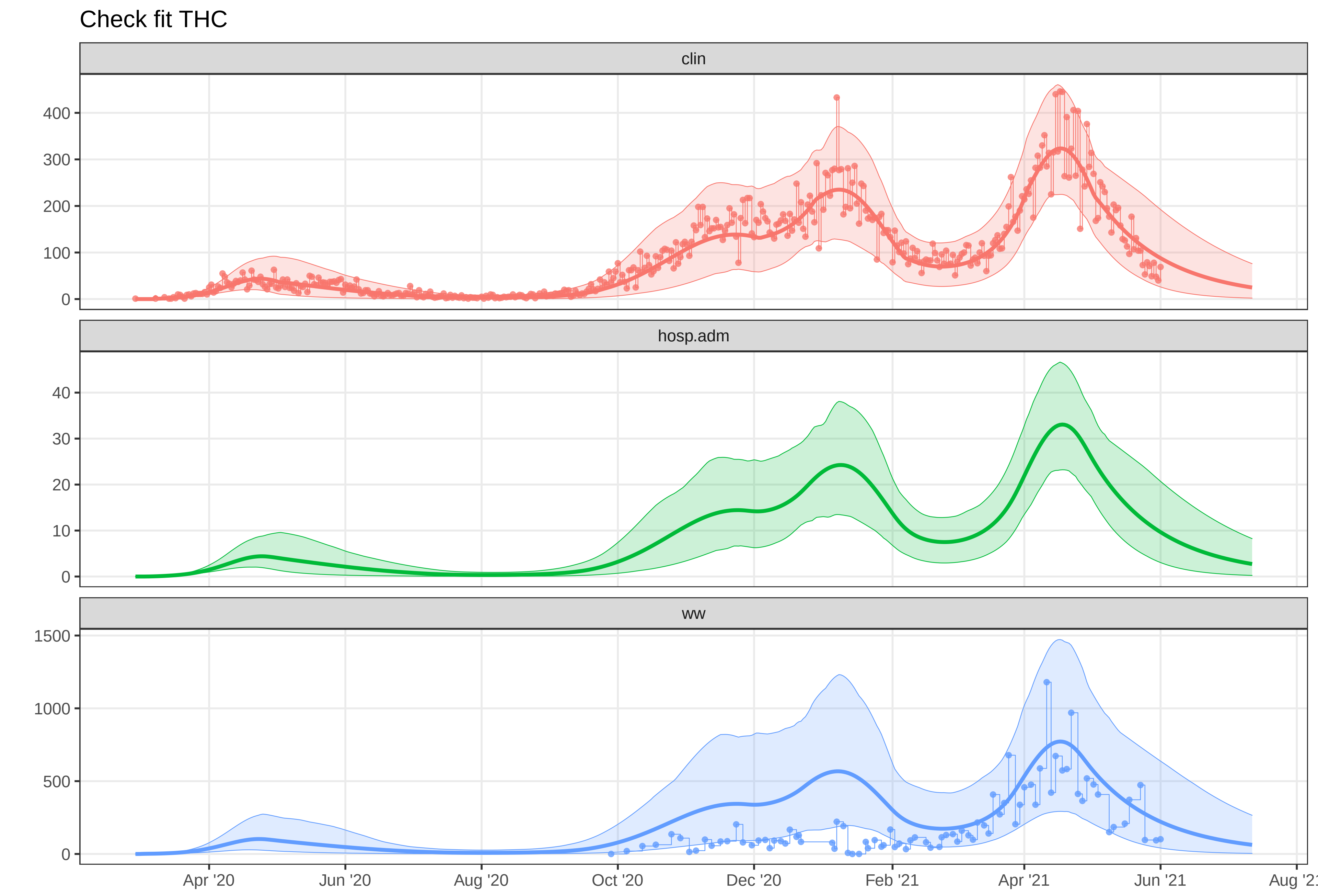

### plot-fit-THC-07-06-clinww.pdf

Observed data for THC

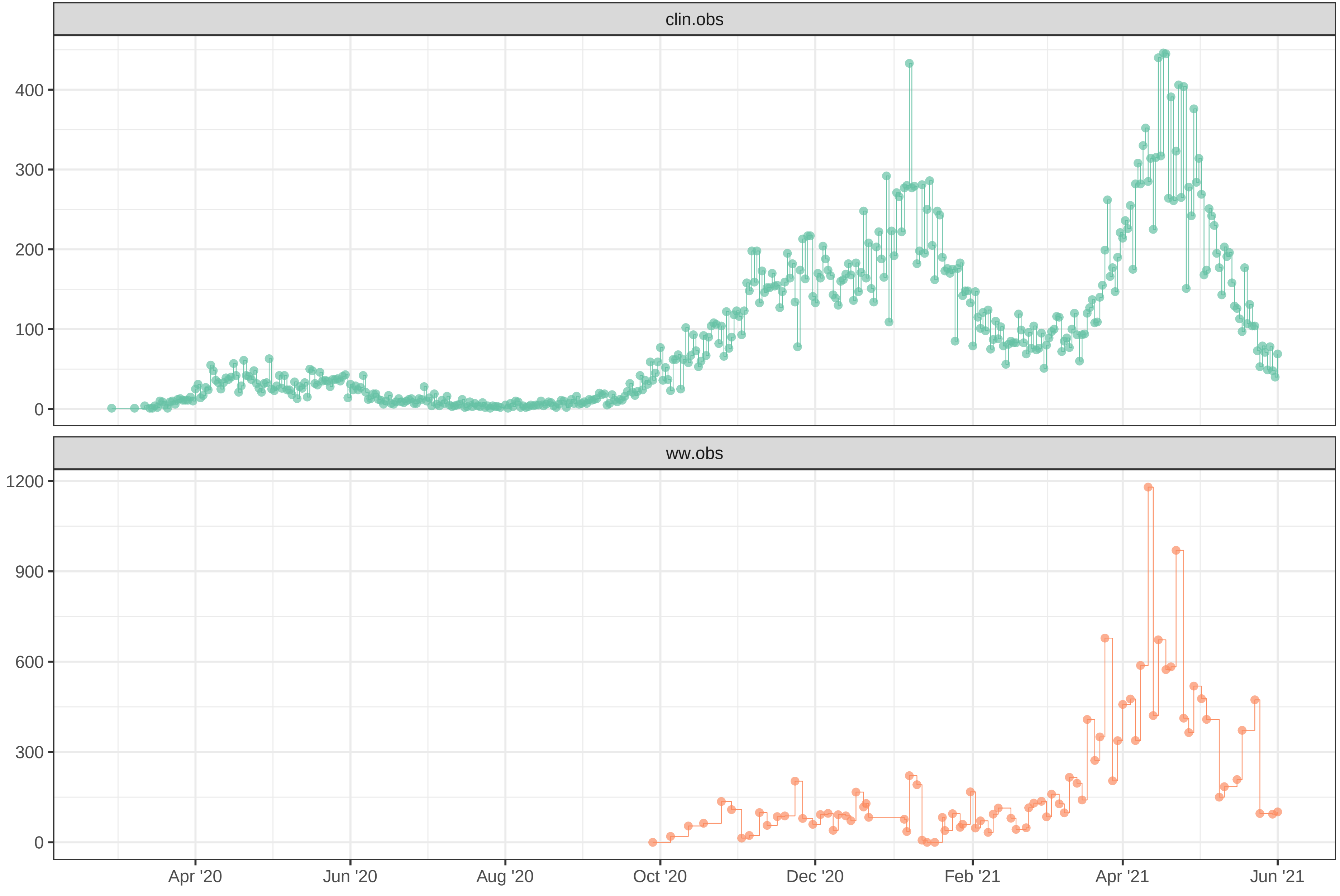

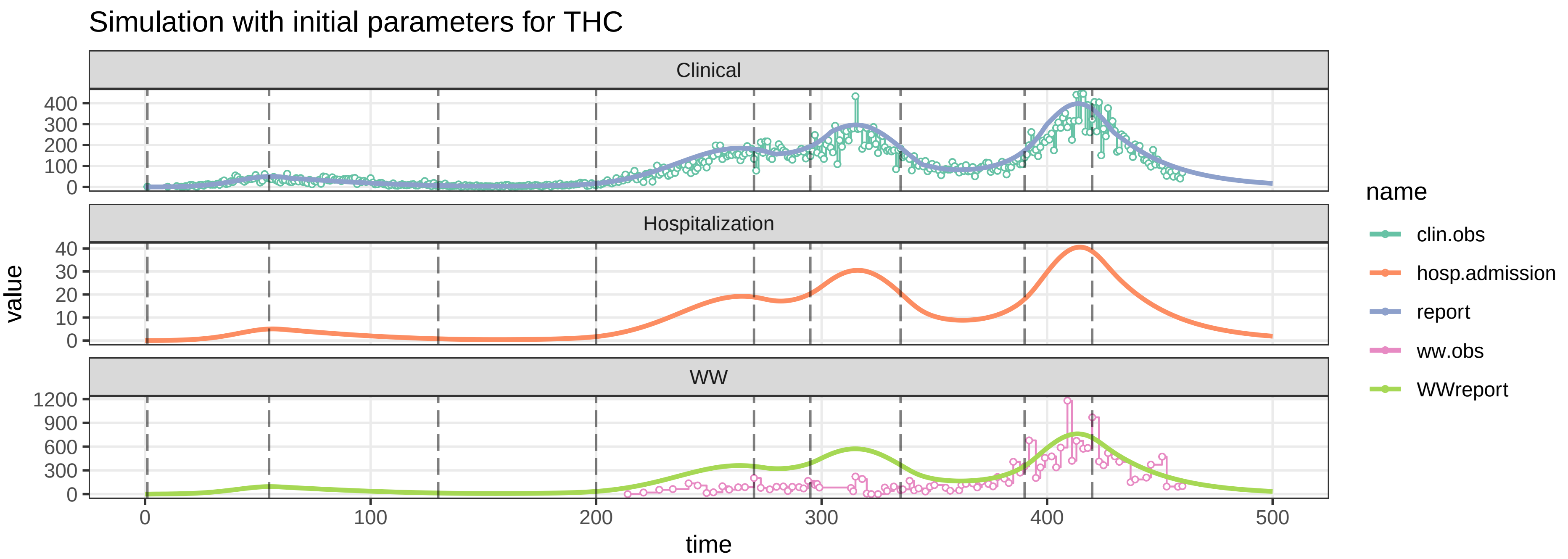

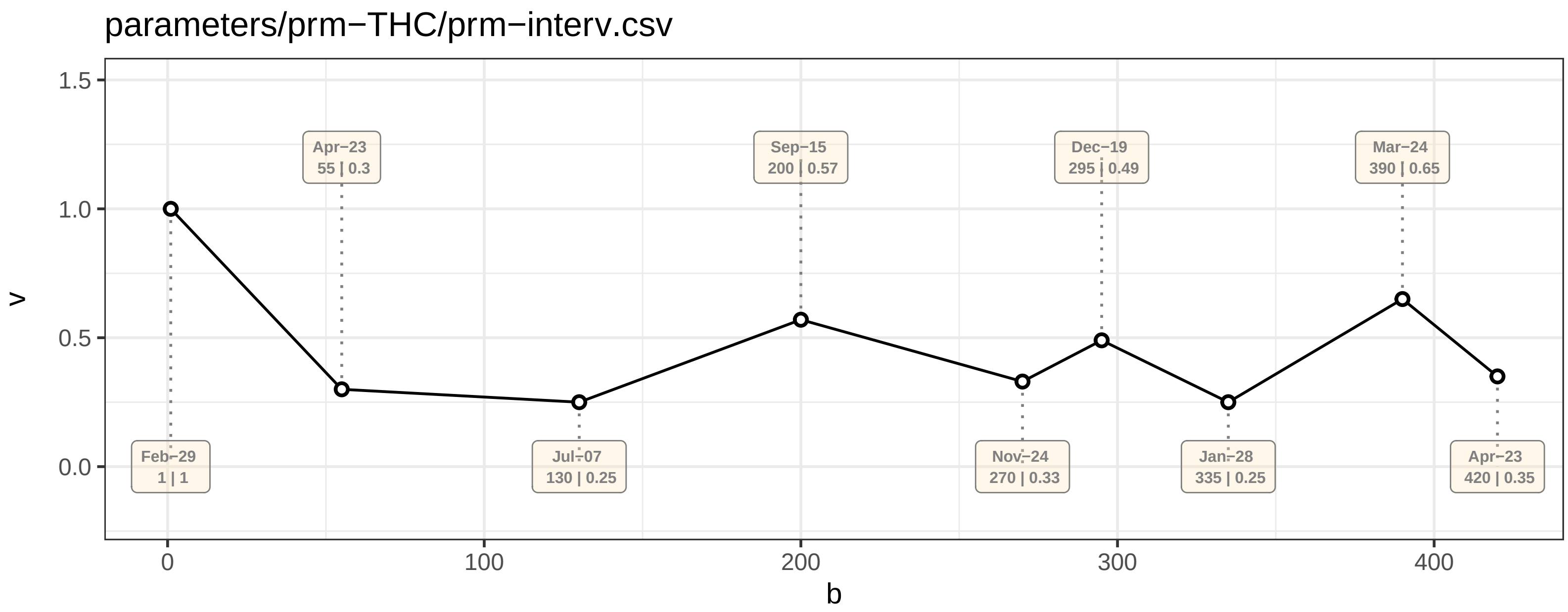

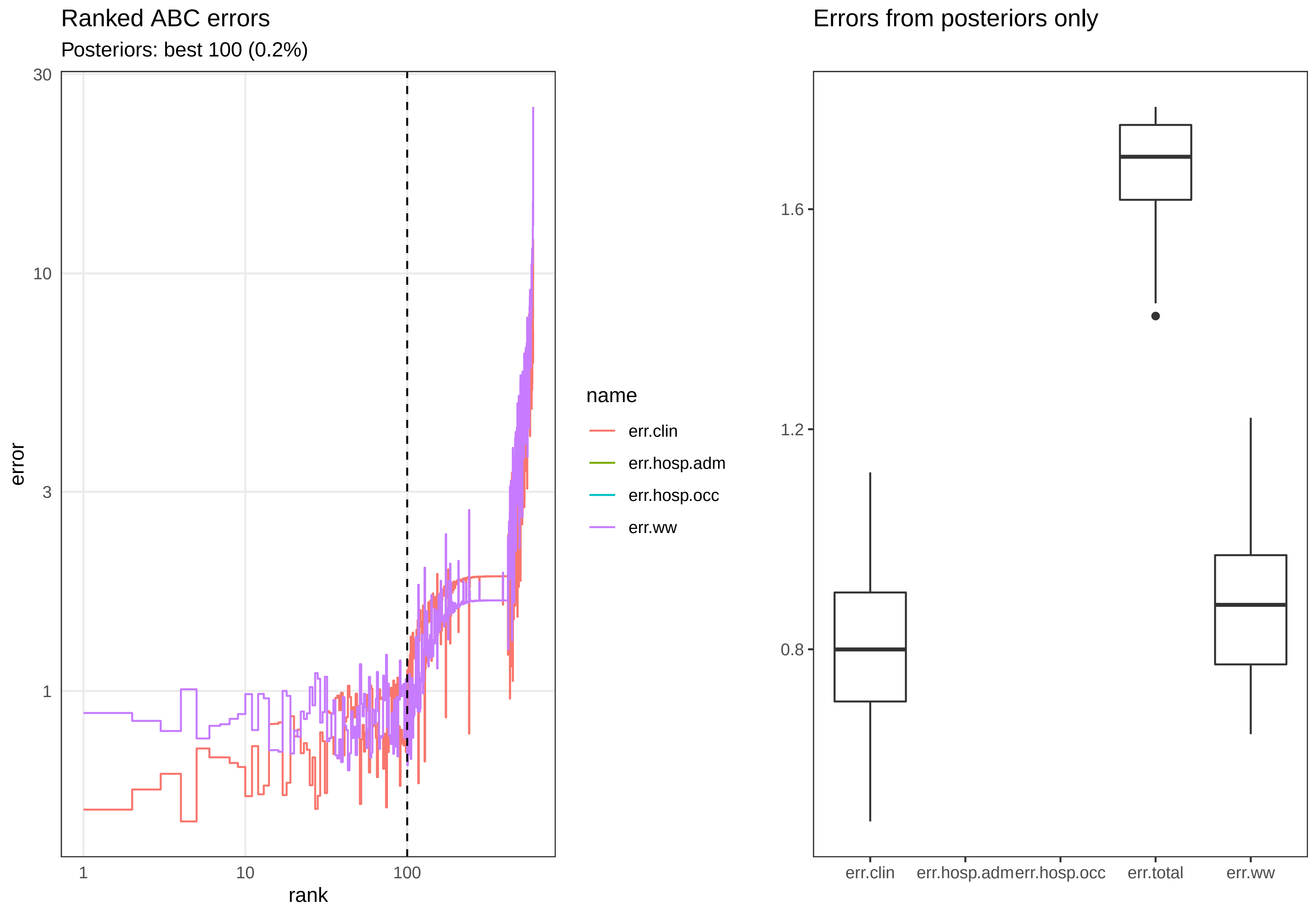

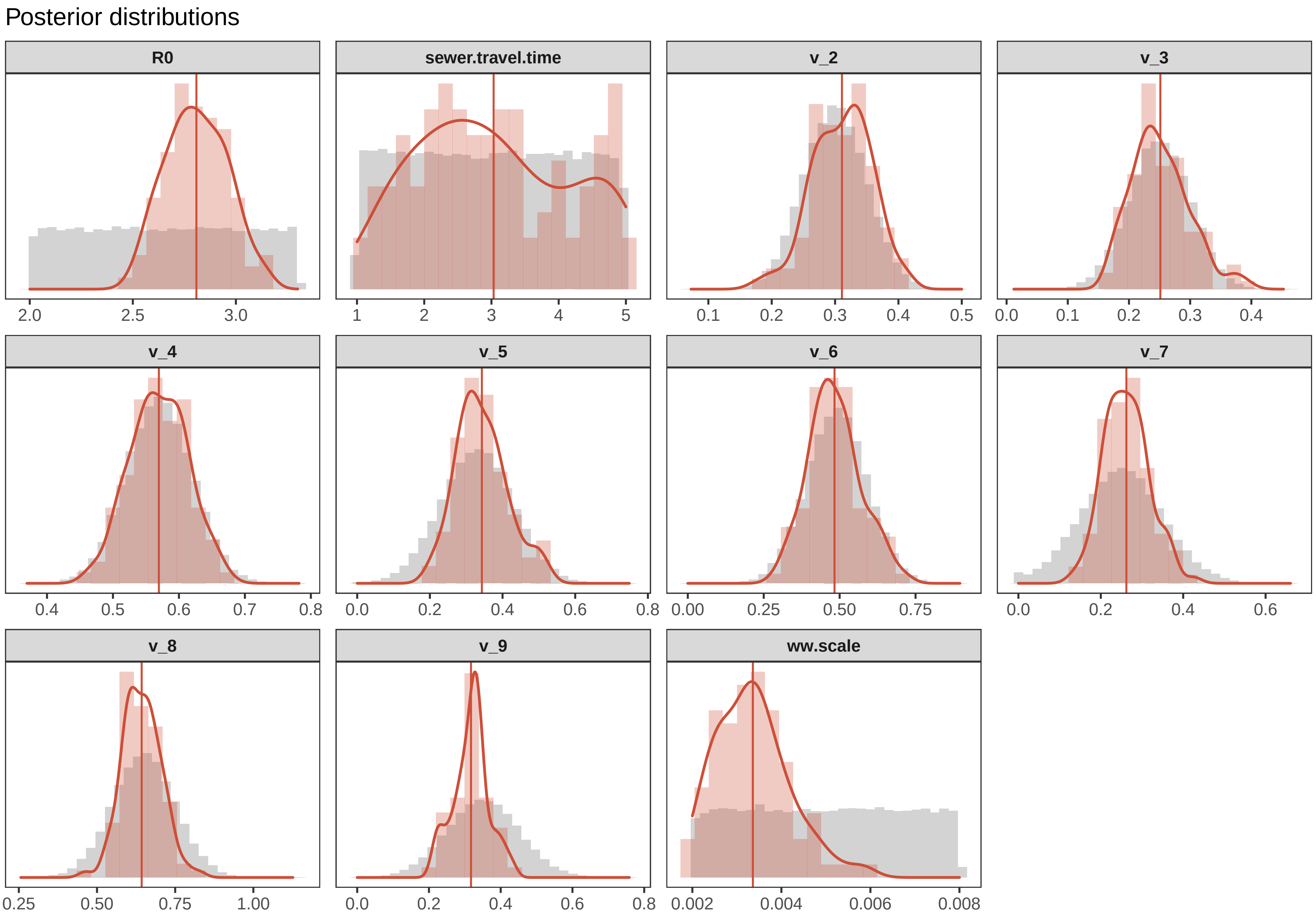

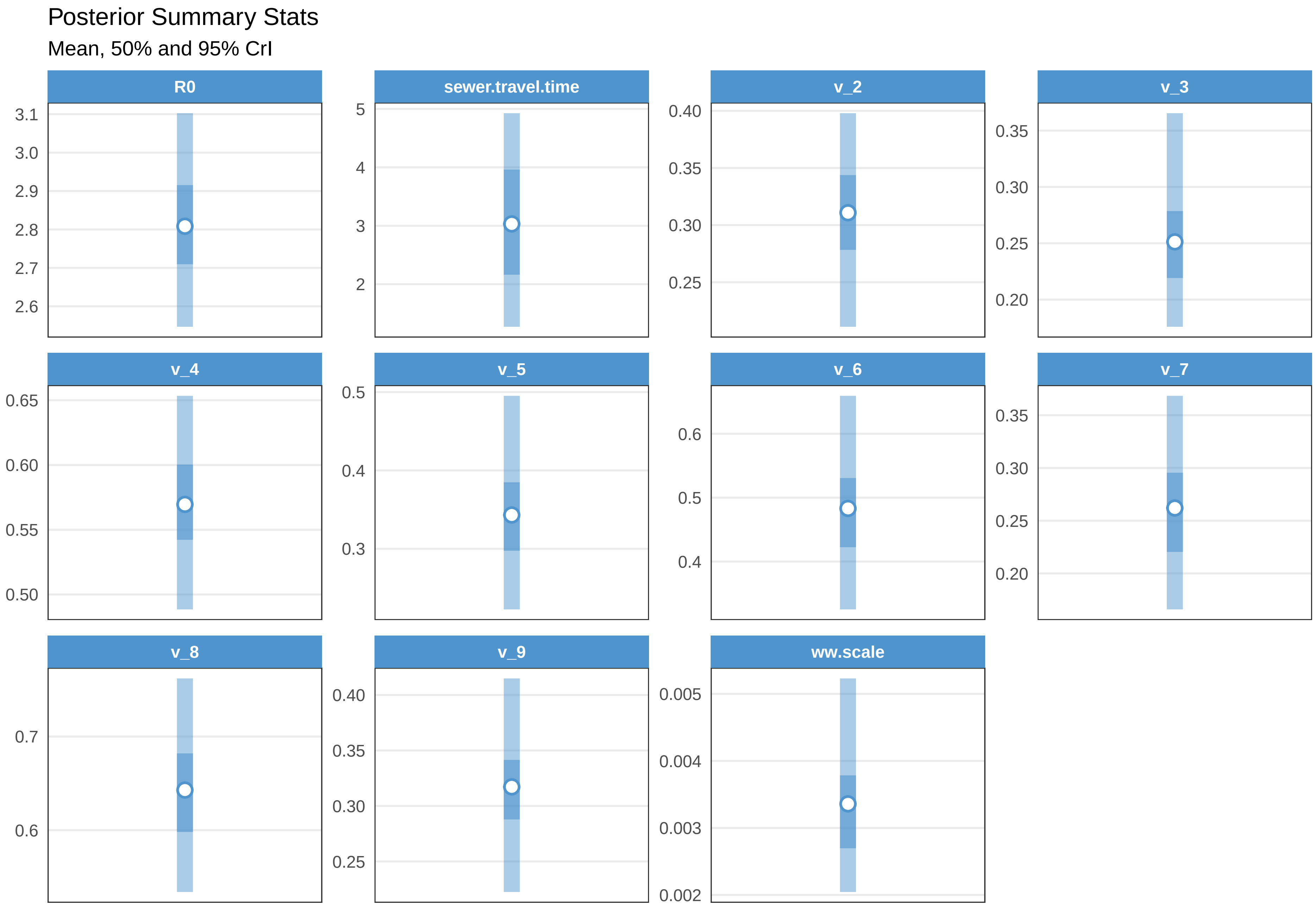

Check fit THC

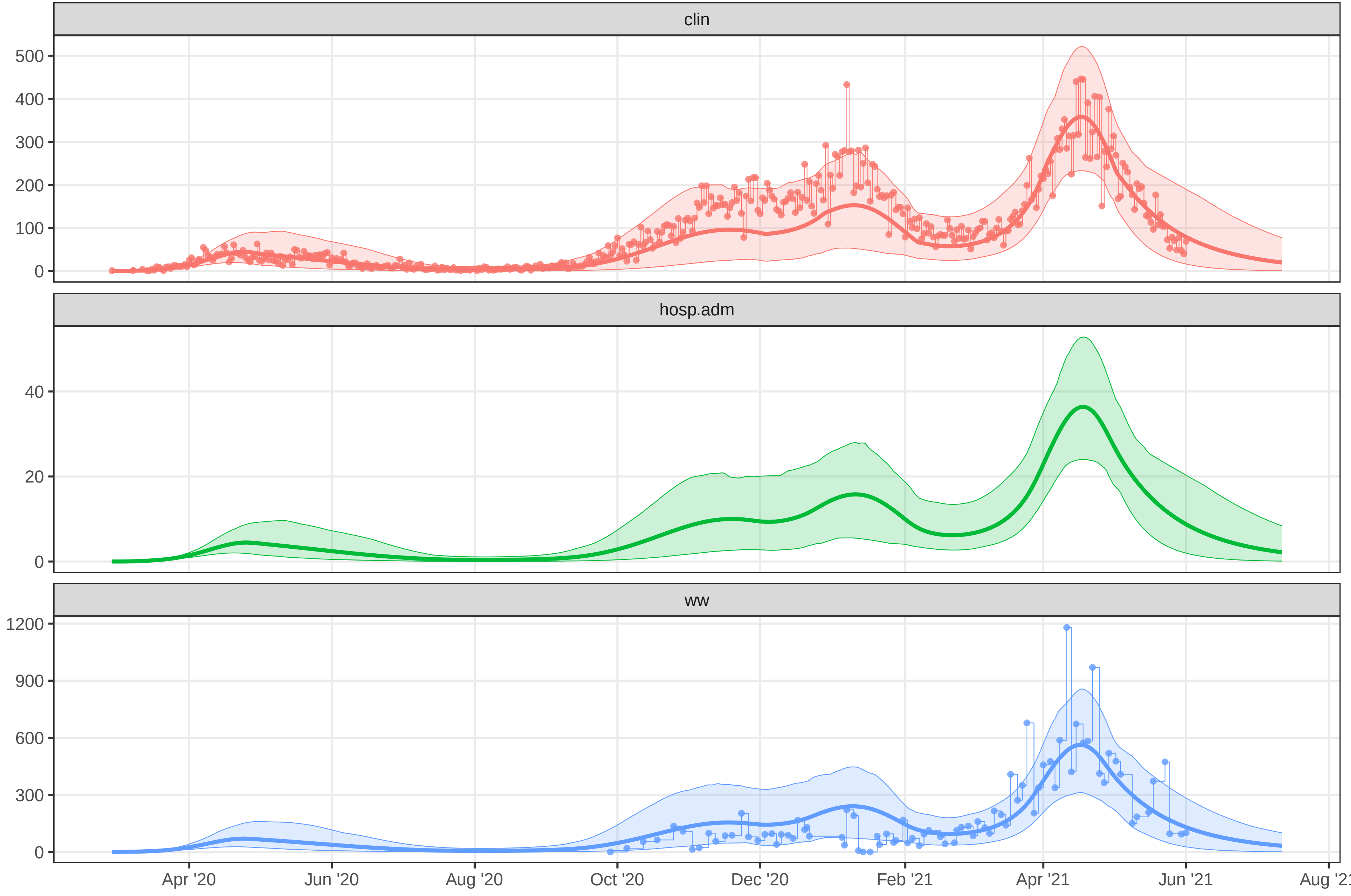

### plot-fit-THC-07-06-ww.pdf

Observed data for THC

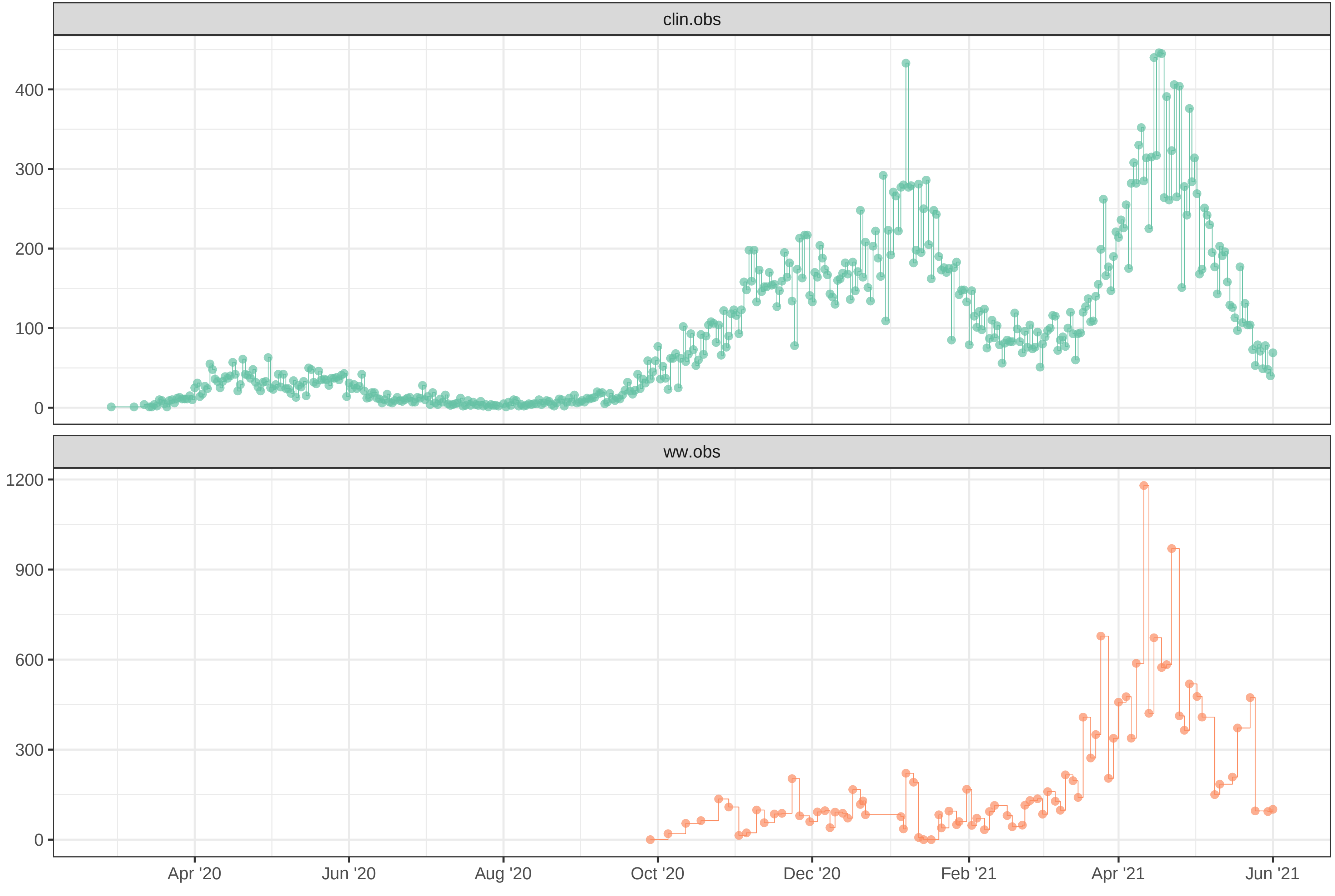

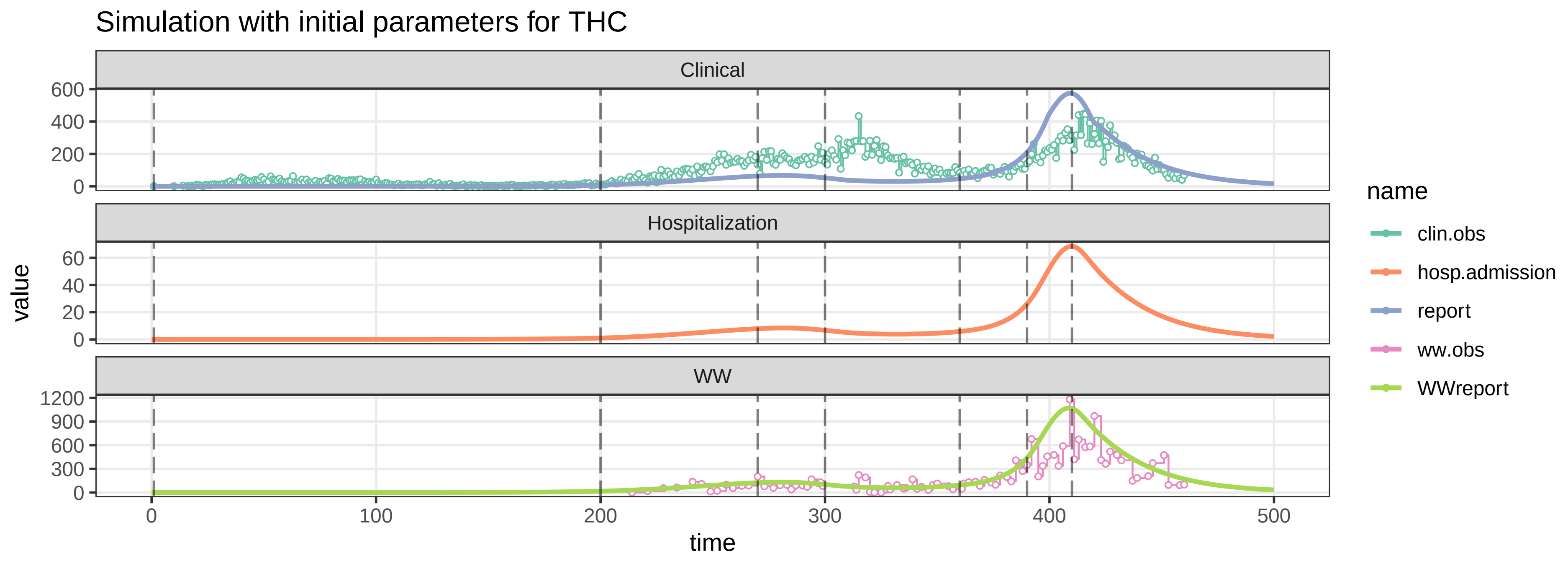

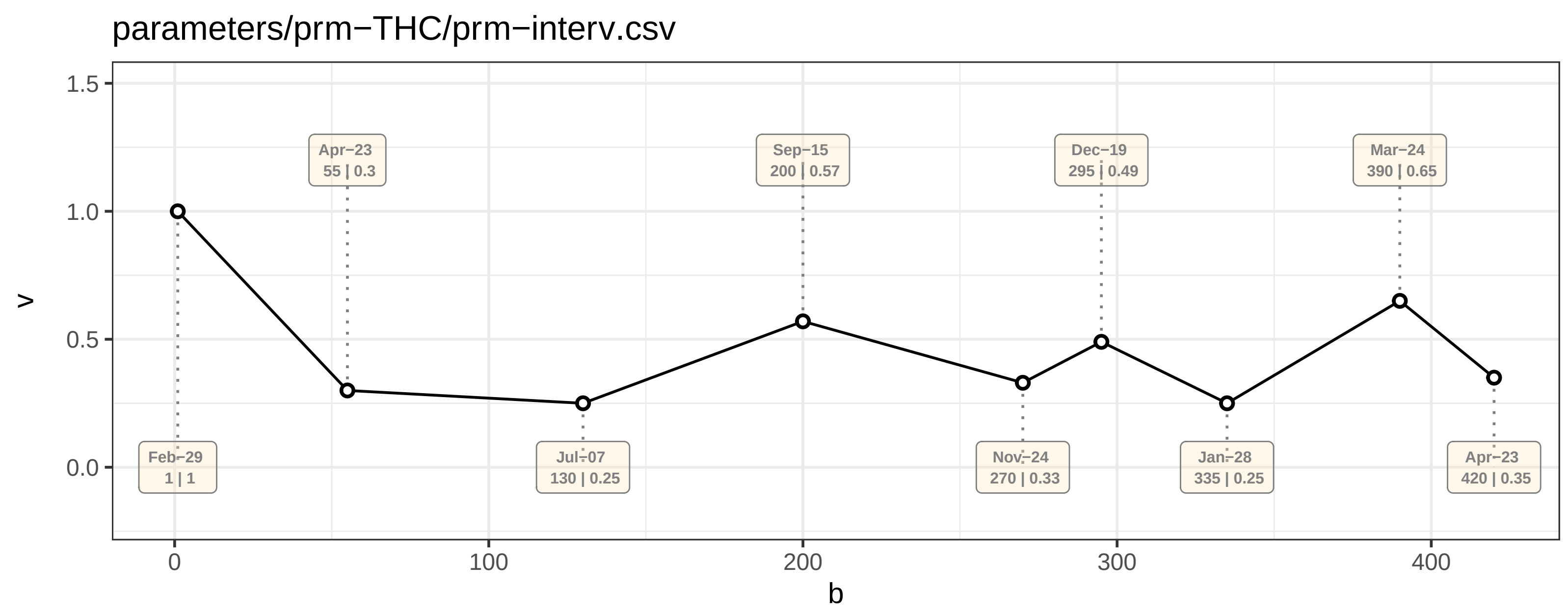

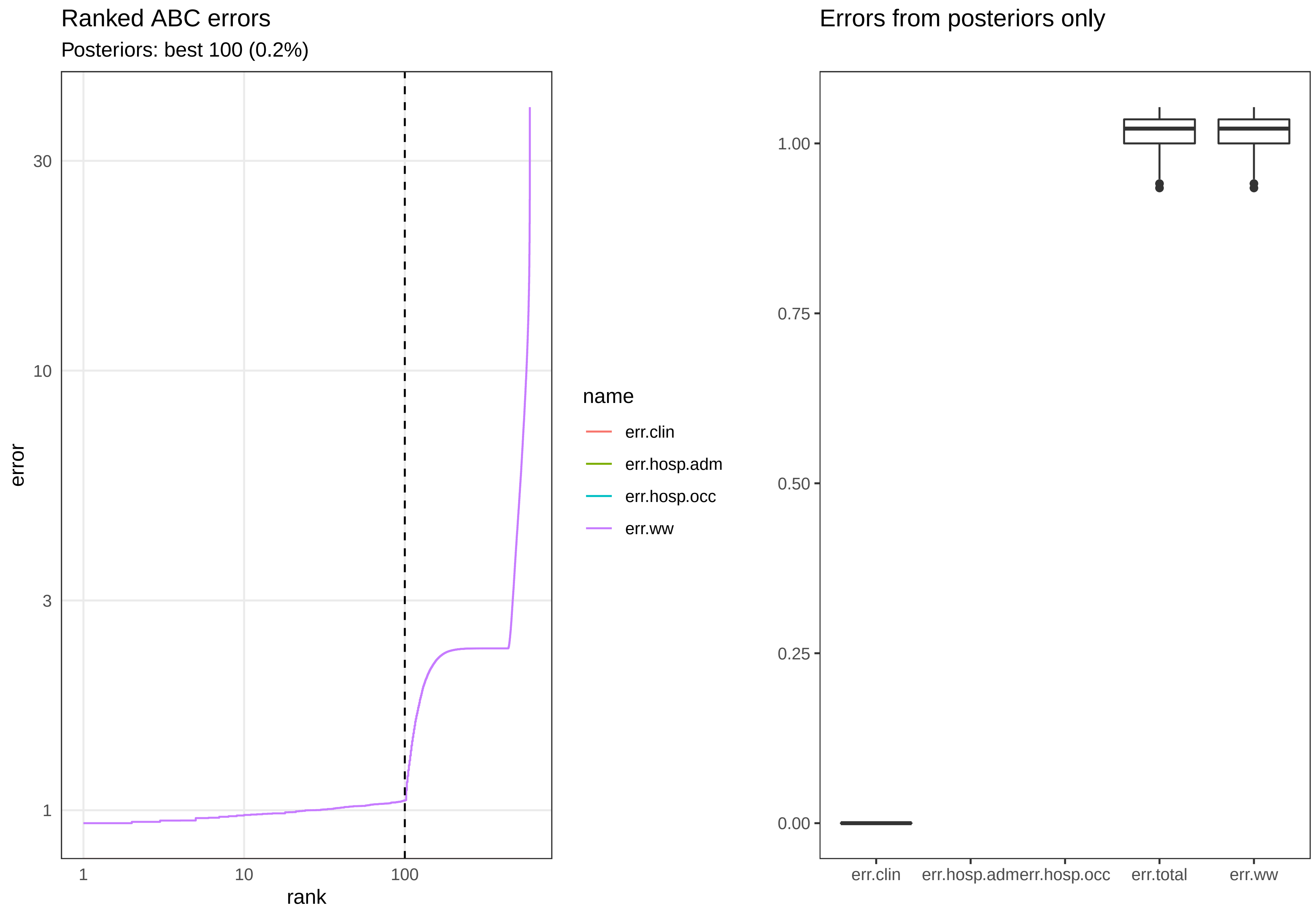

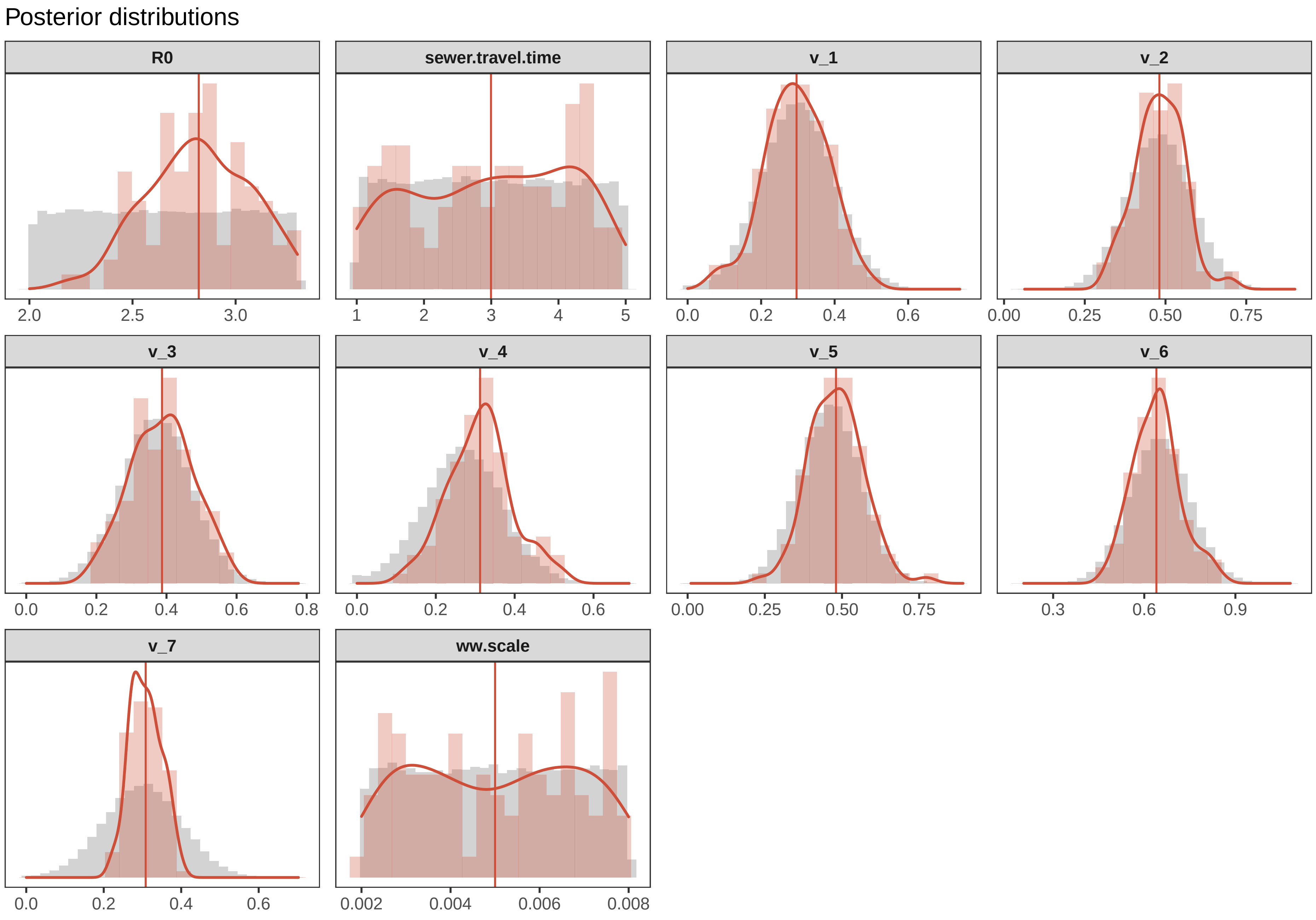

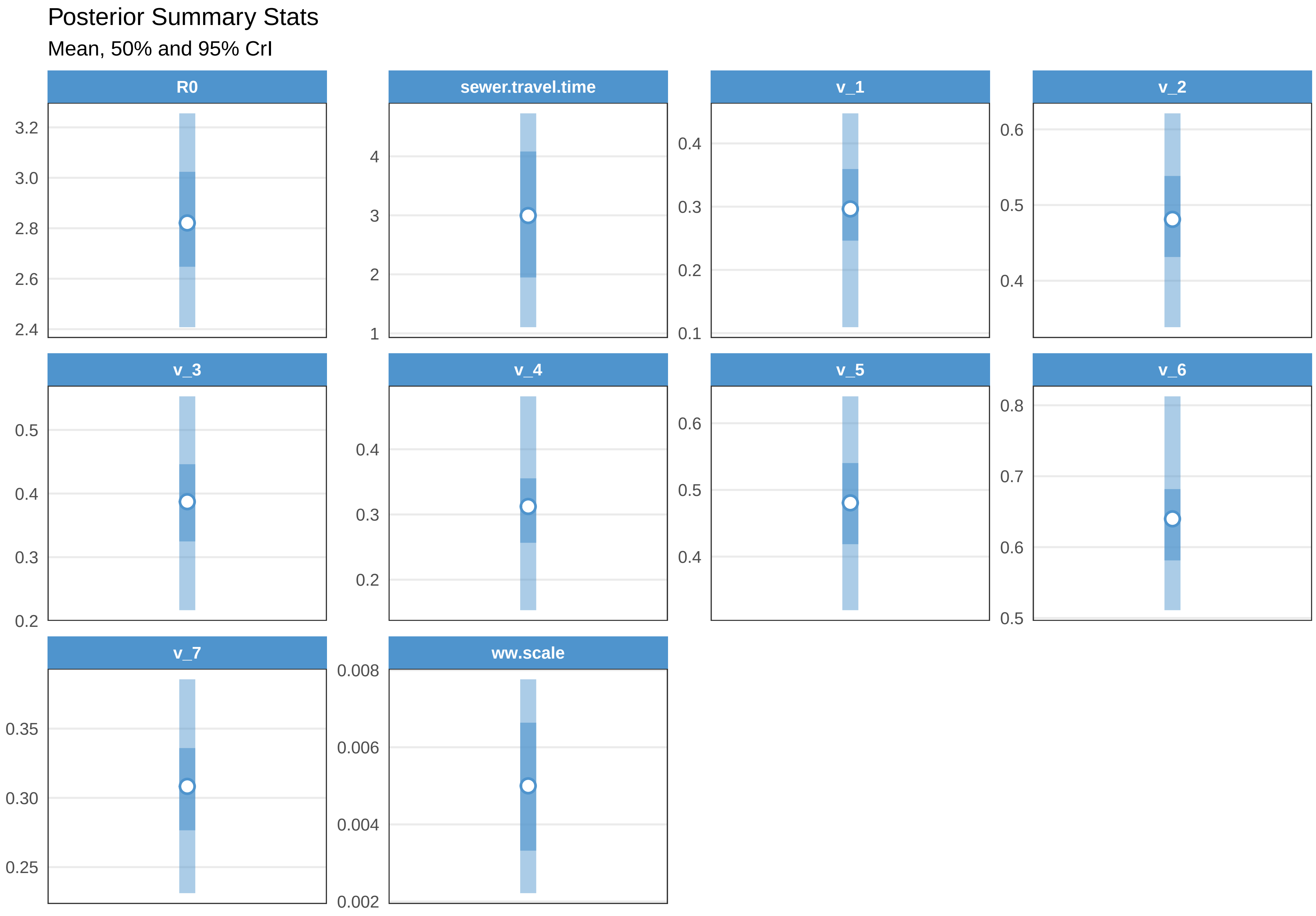

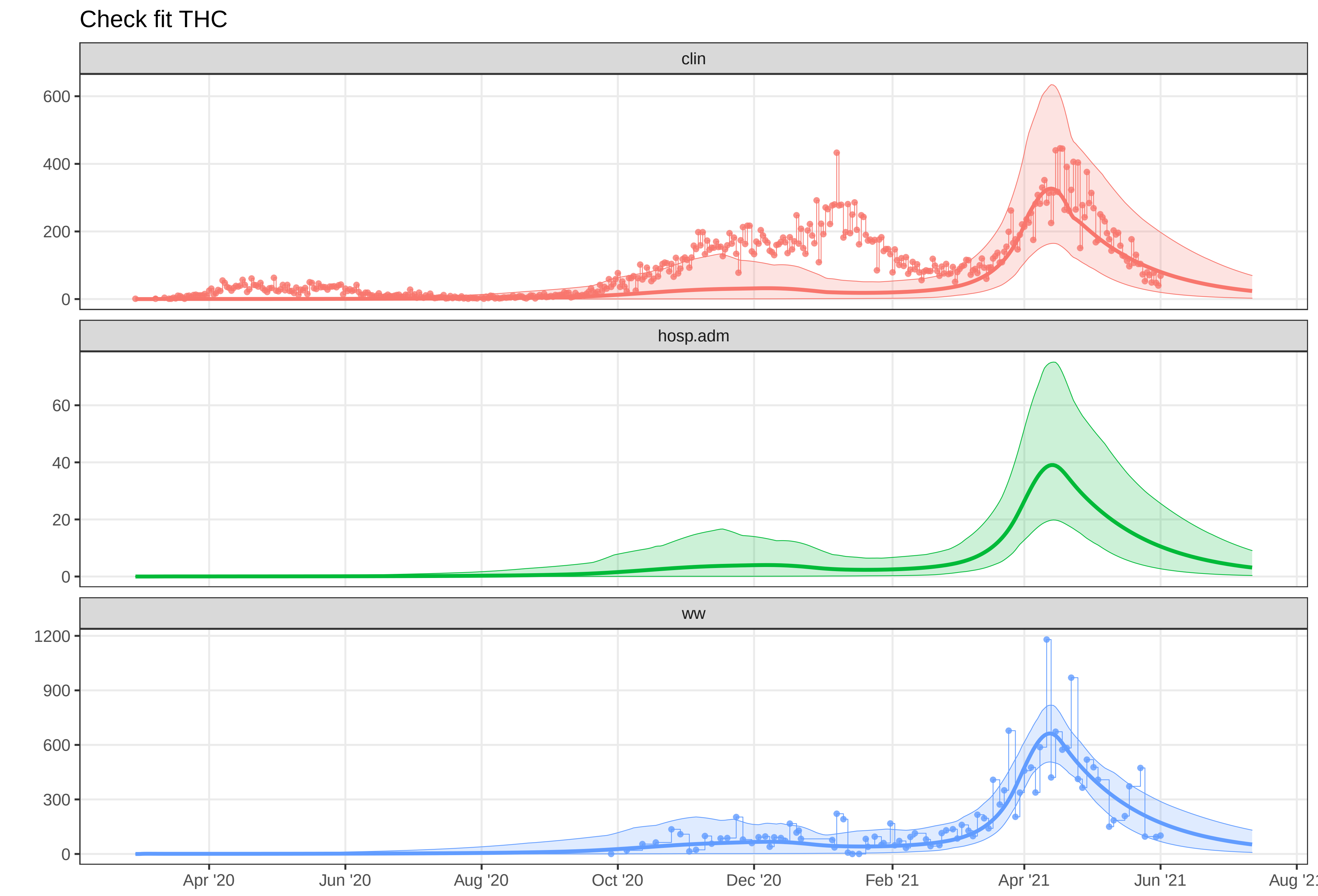

### plot-fit-THU-07-06-clin.pdf

#### Observed data for THU

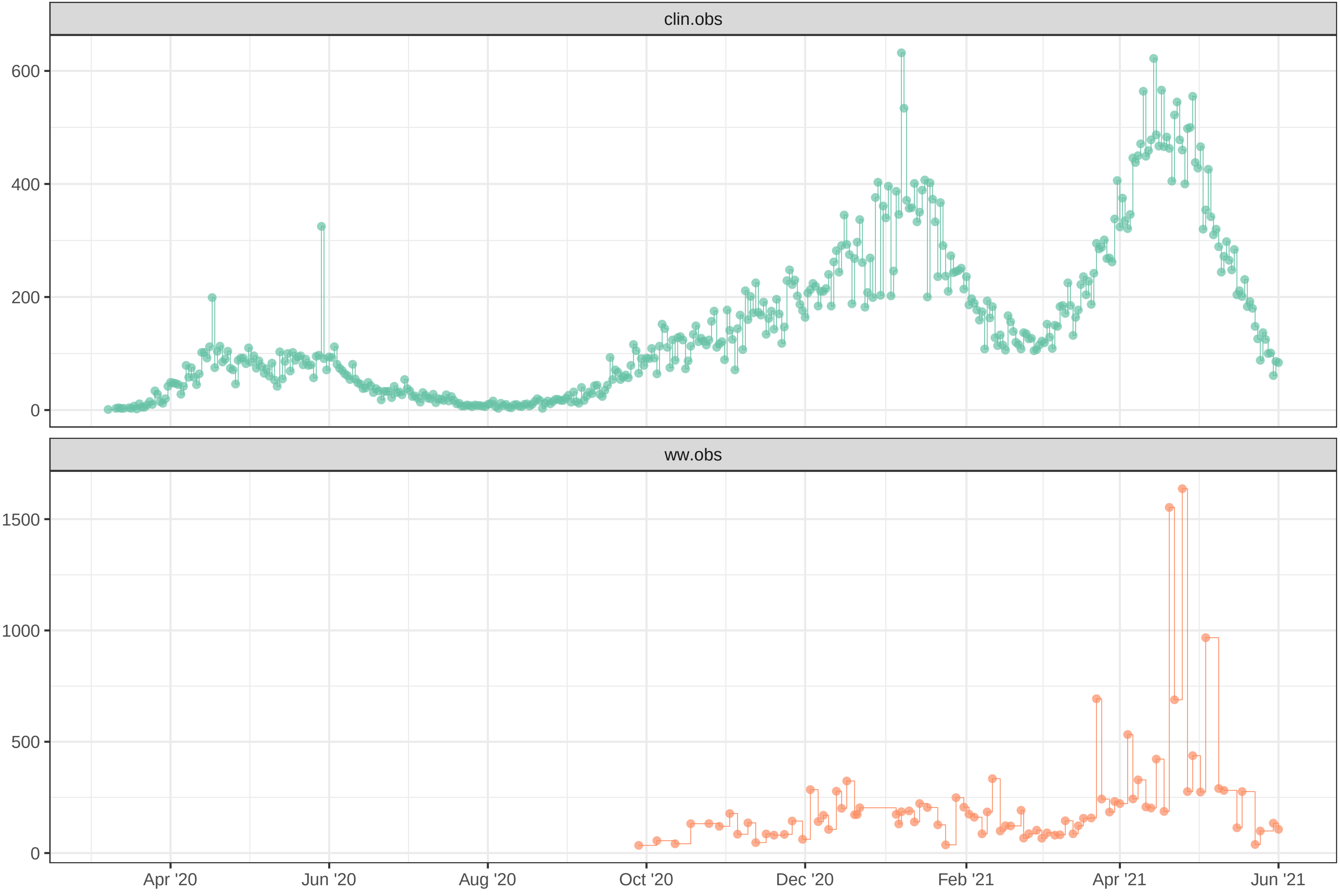

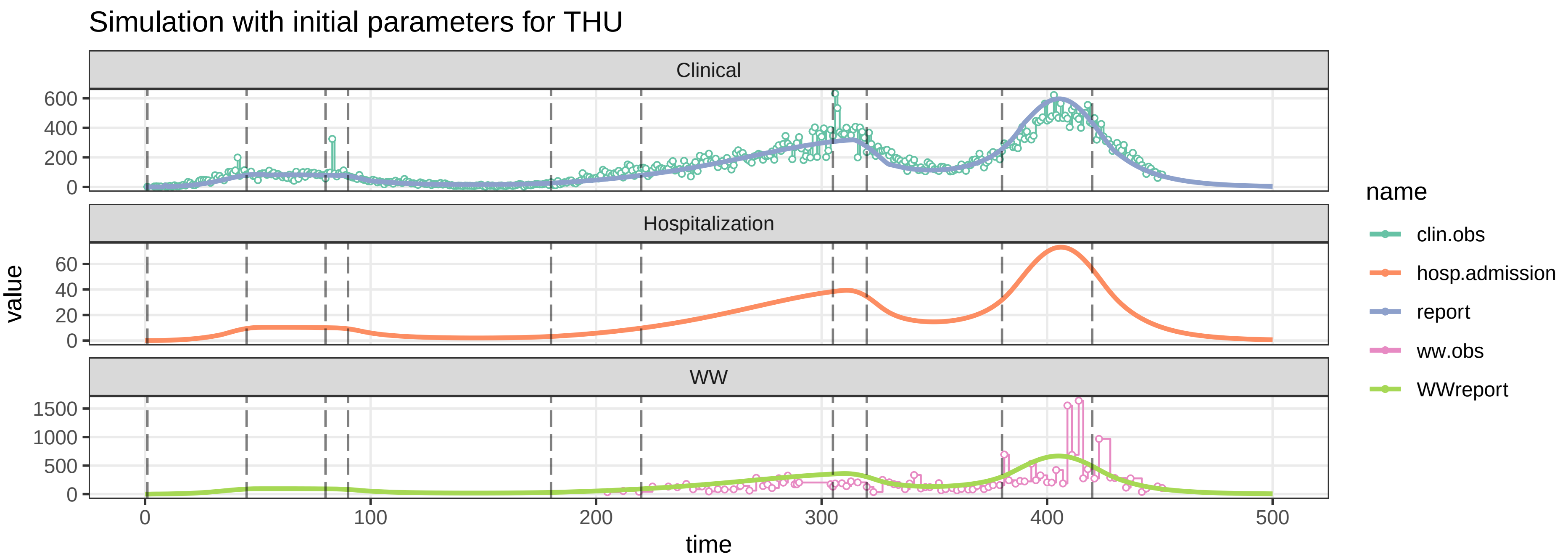

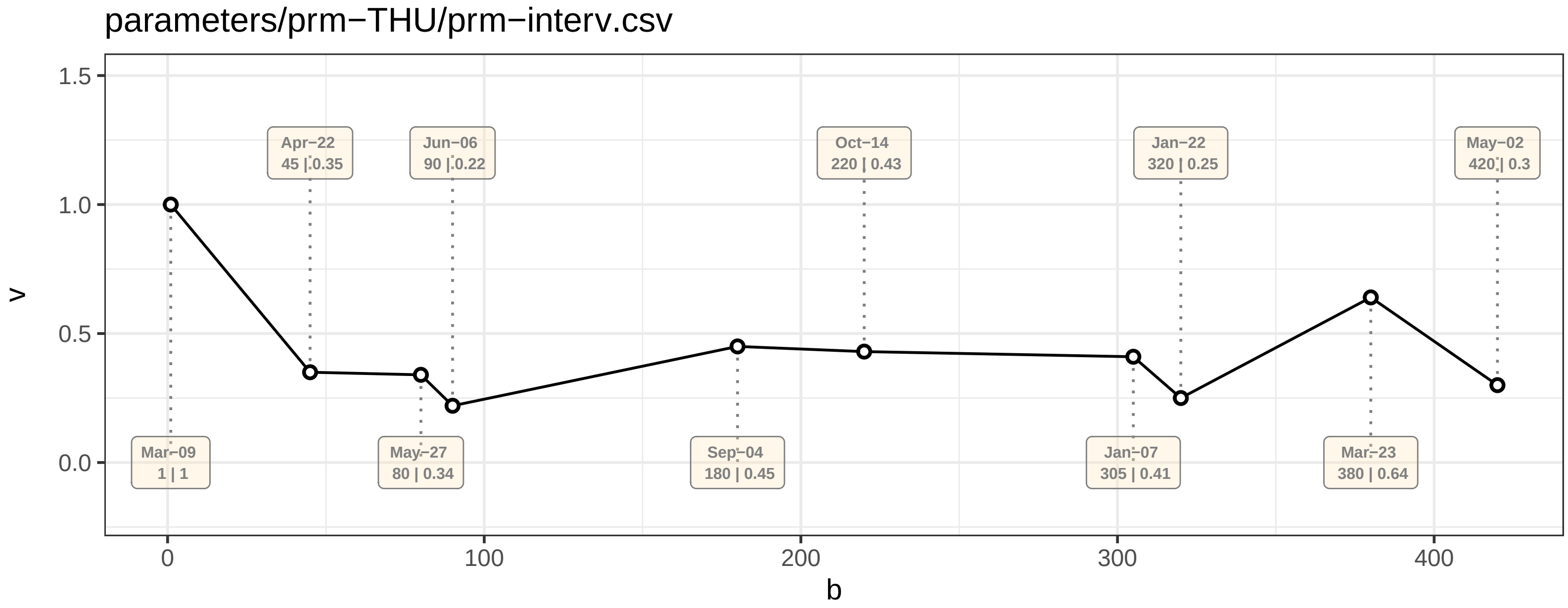

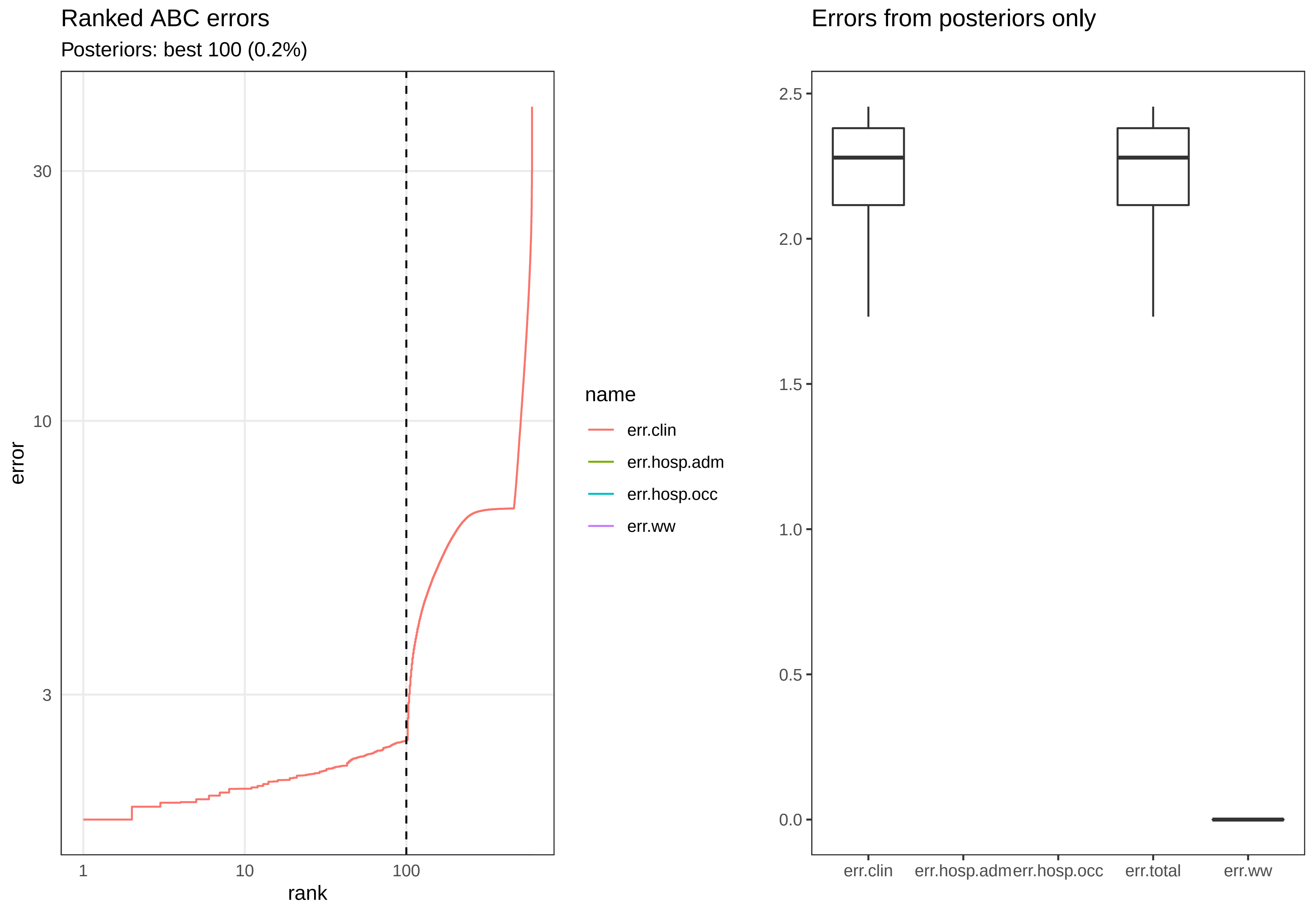

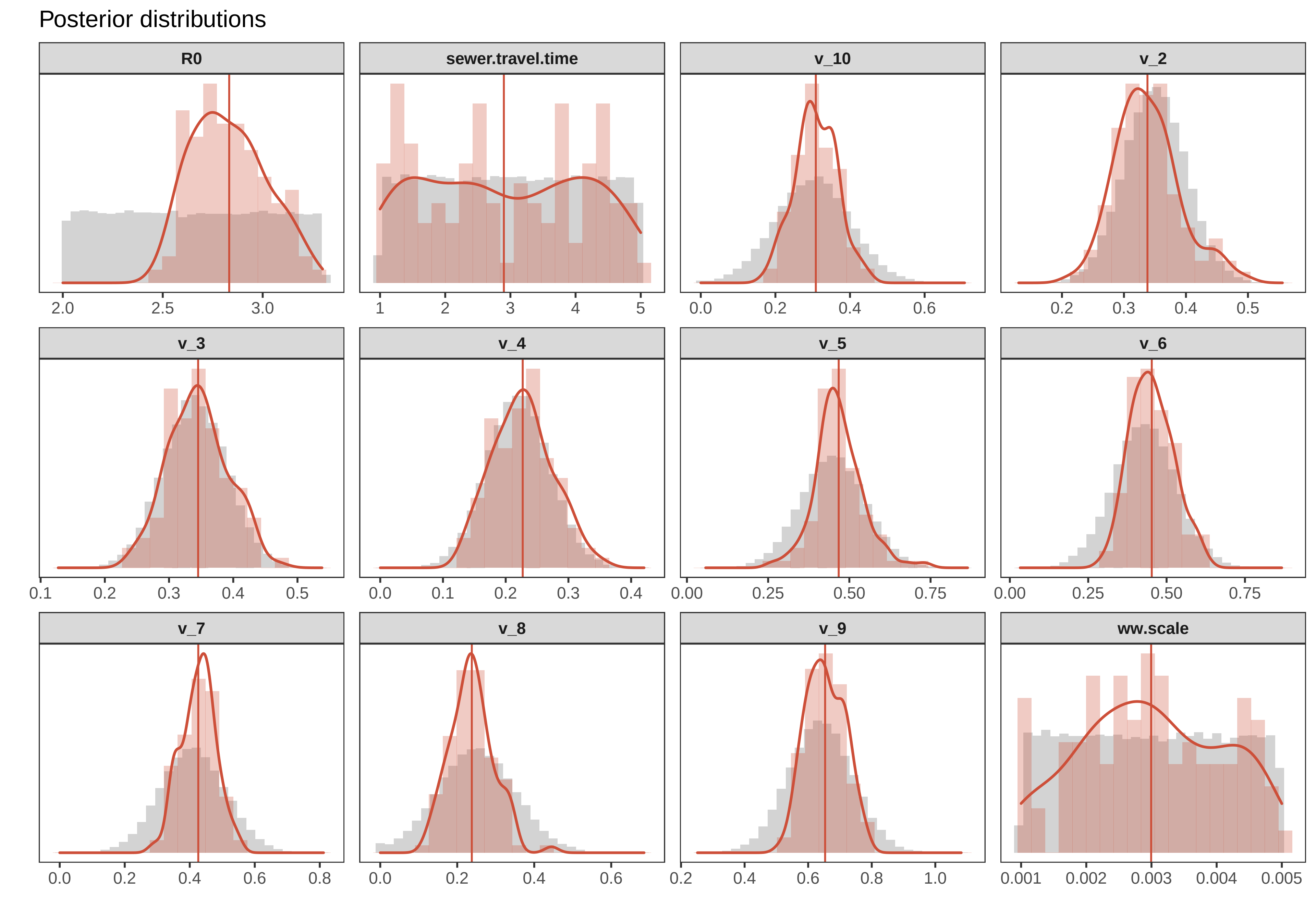

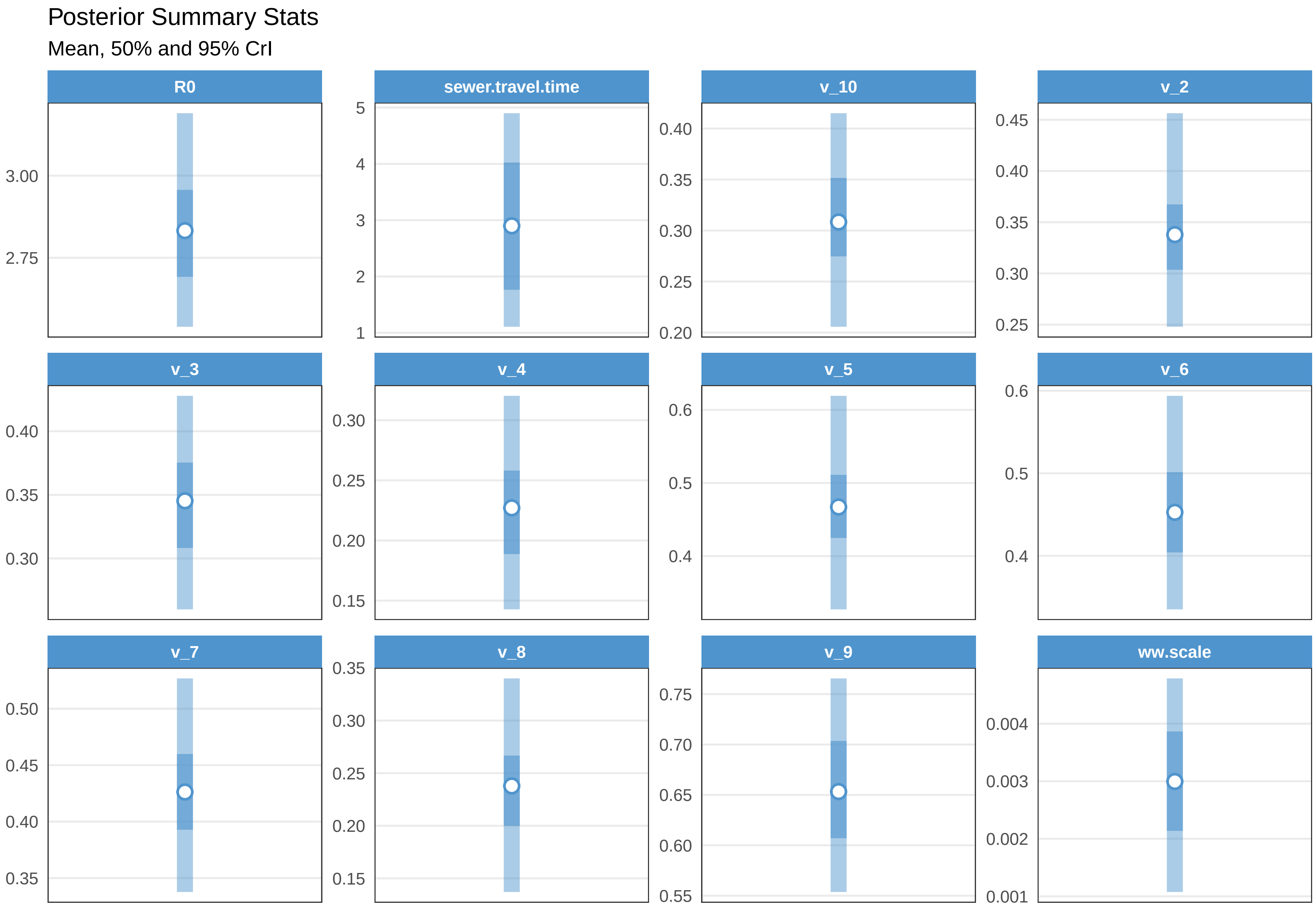

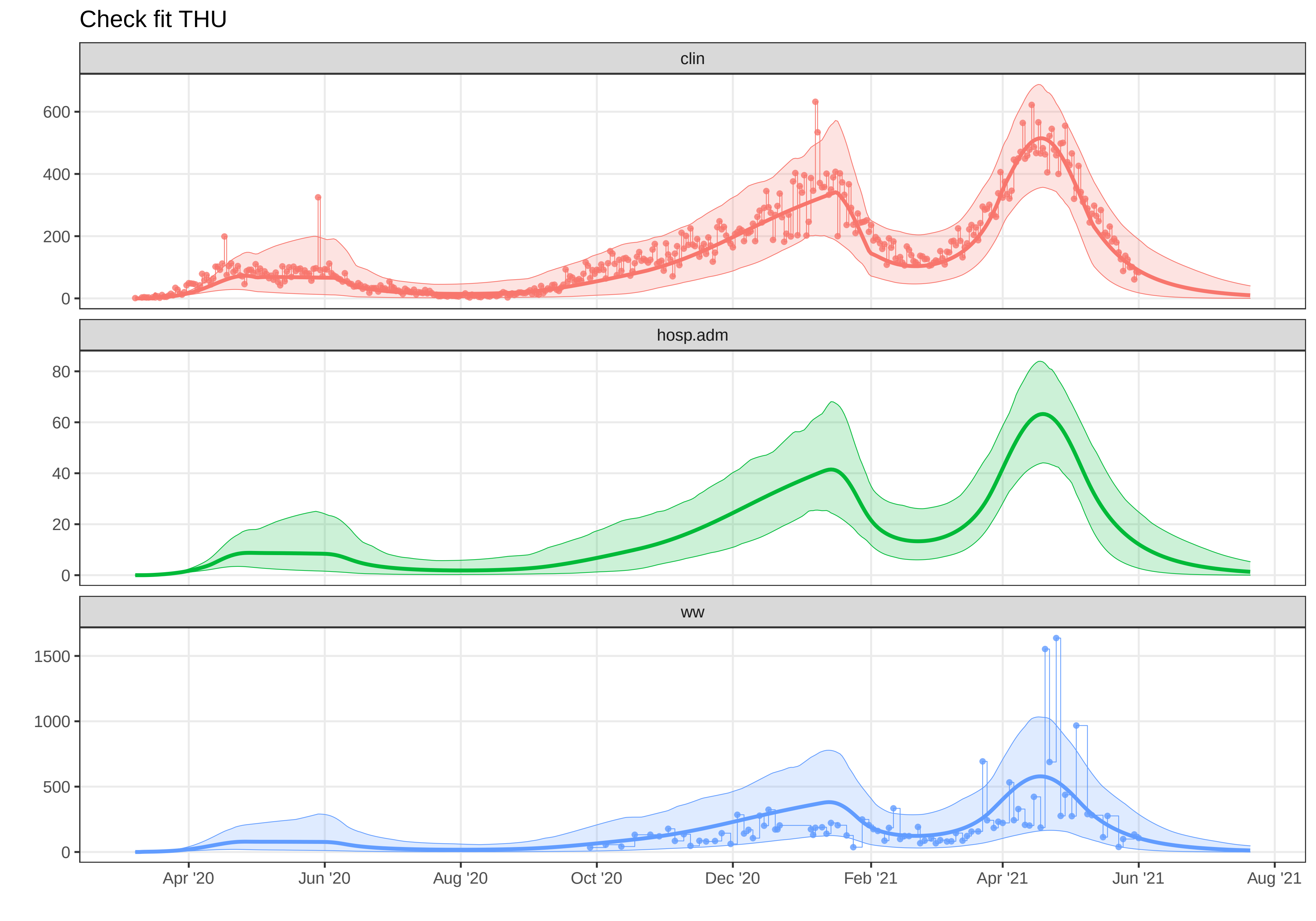

### plot-fit-THU-07-06-ww.pdf

Observed data for THU

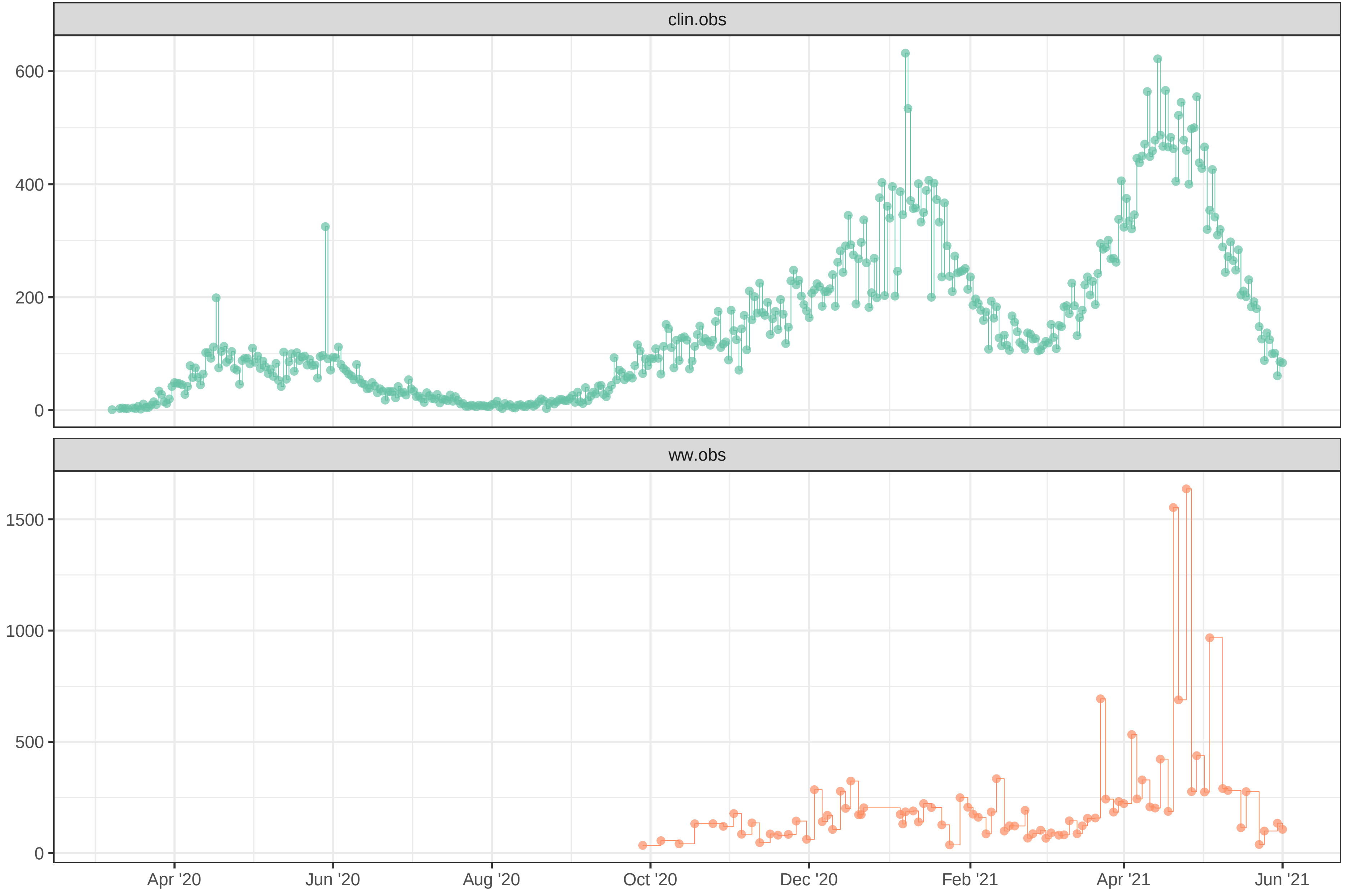

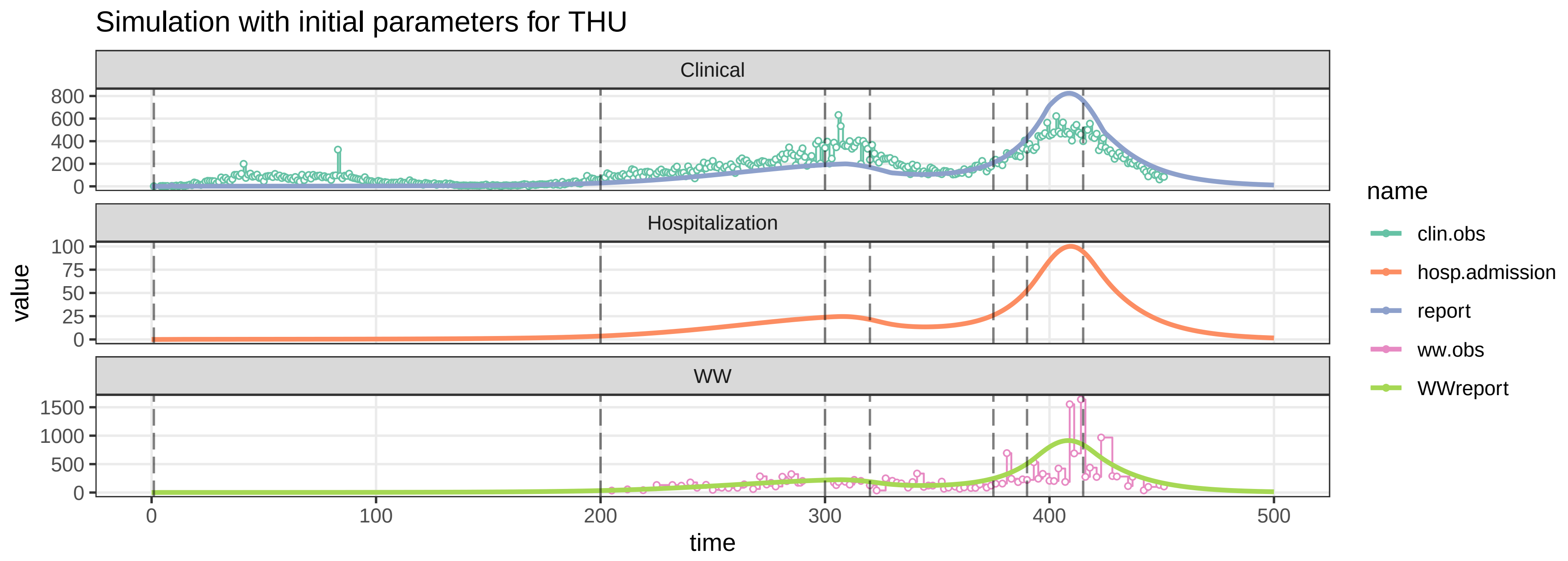

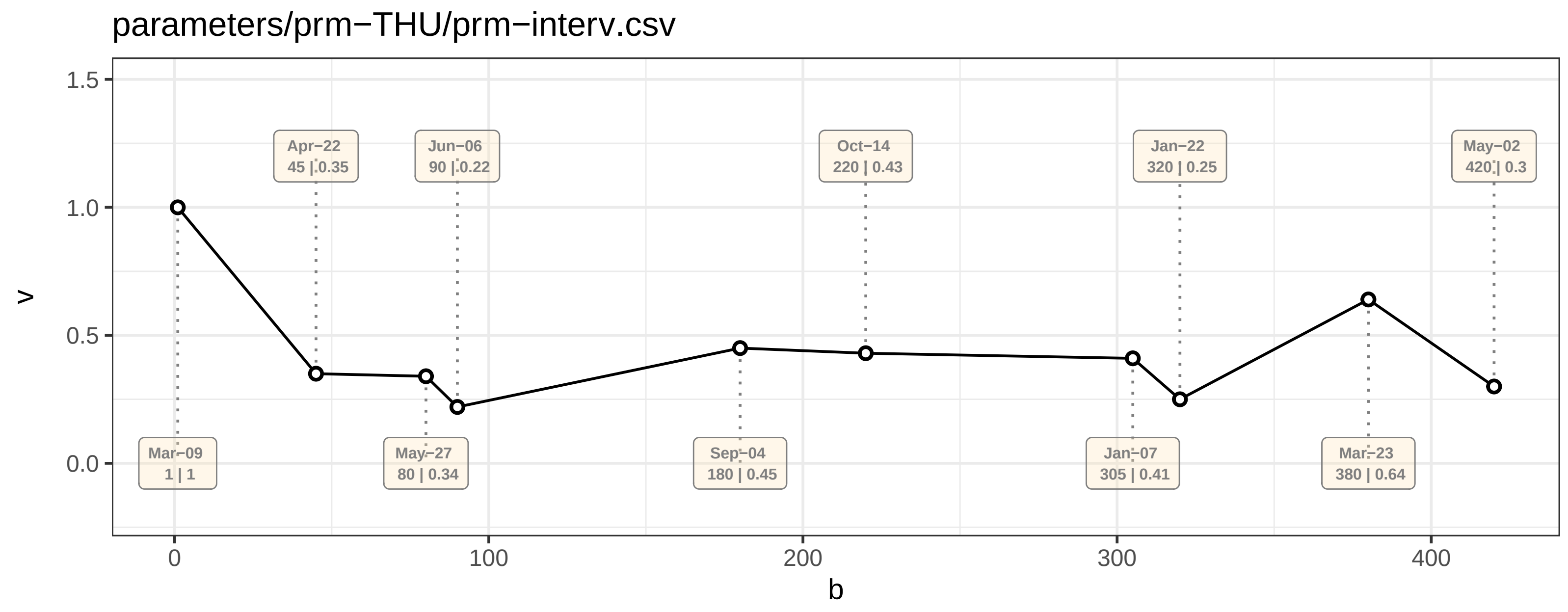

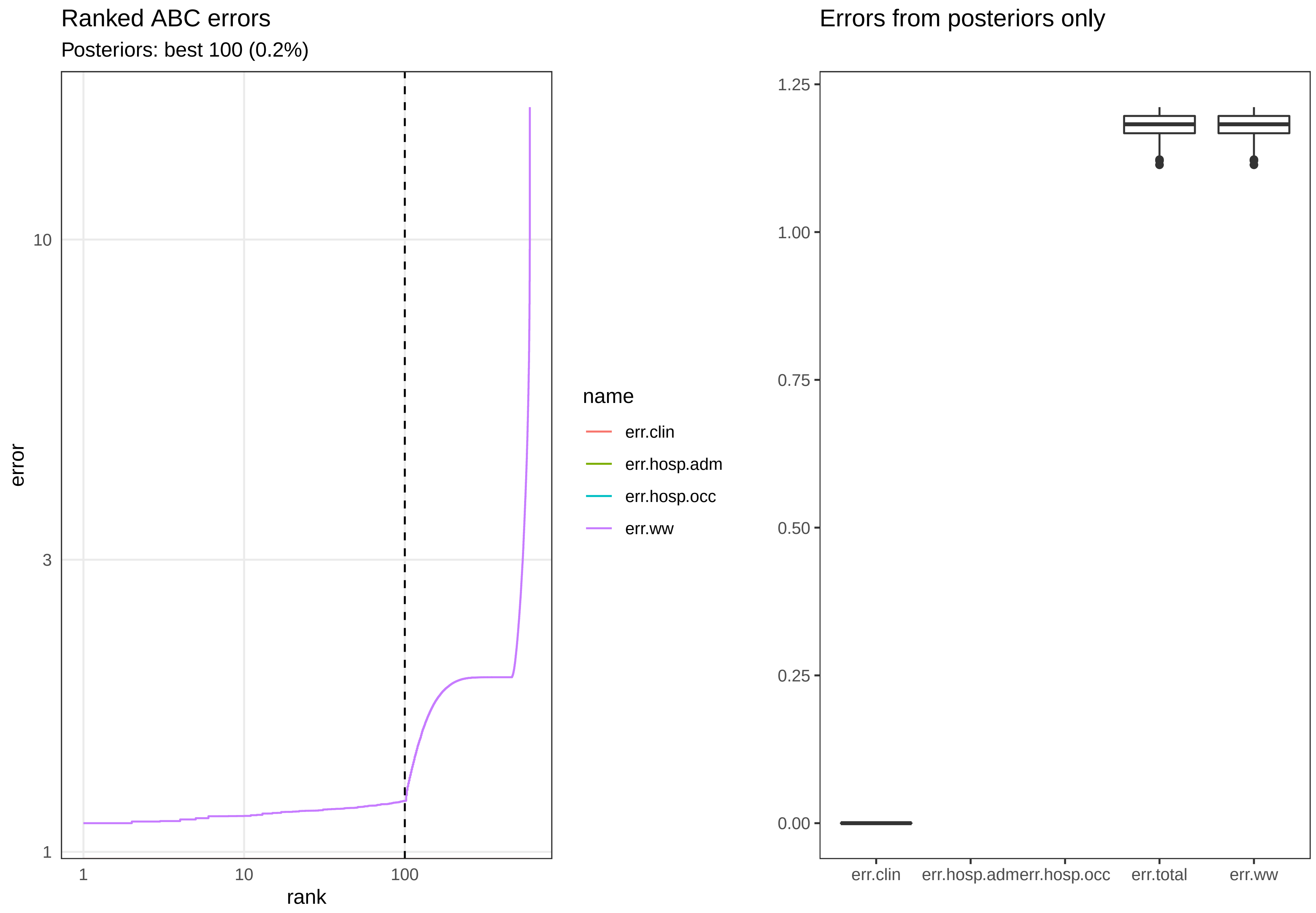

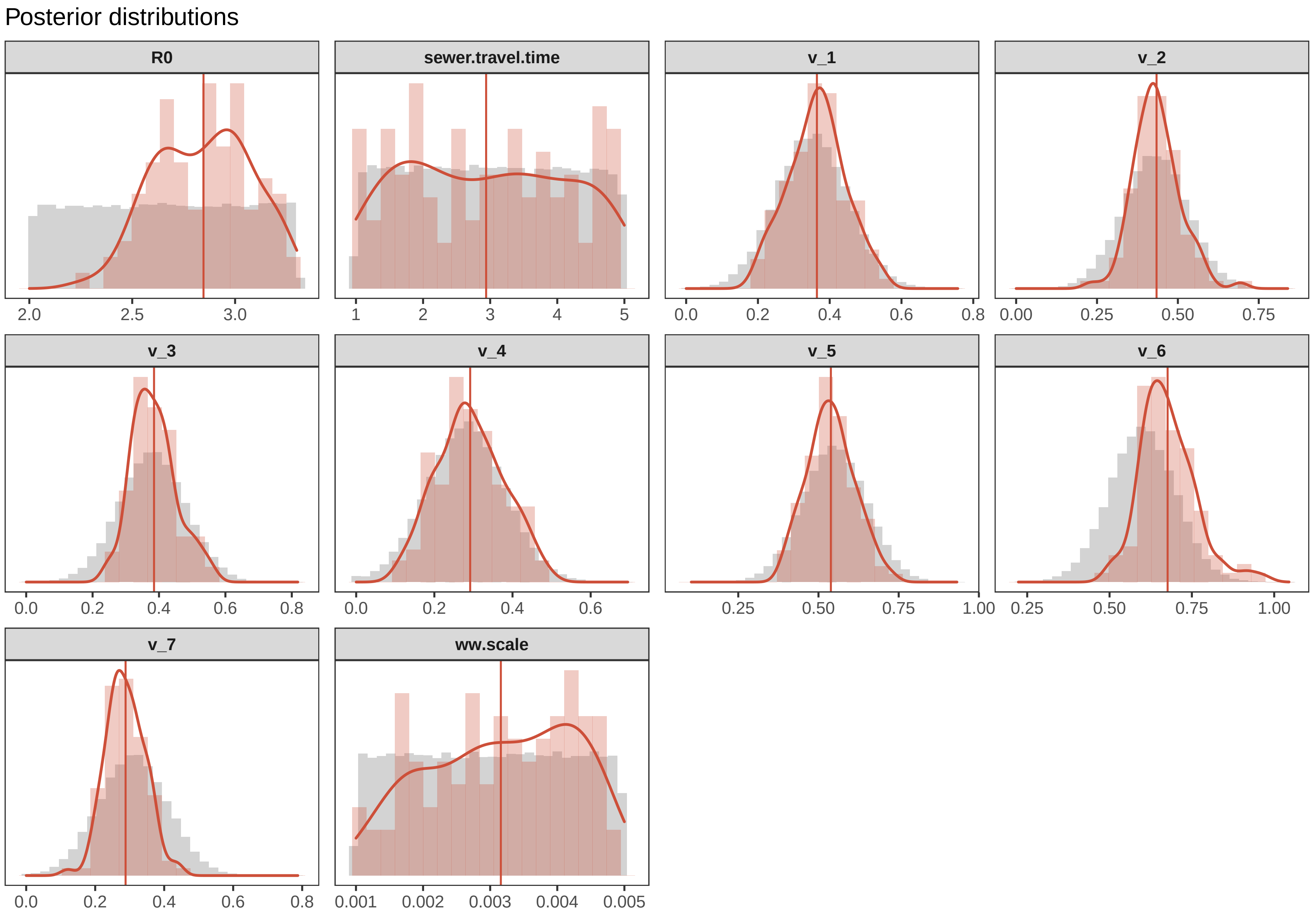

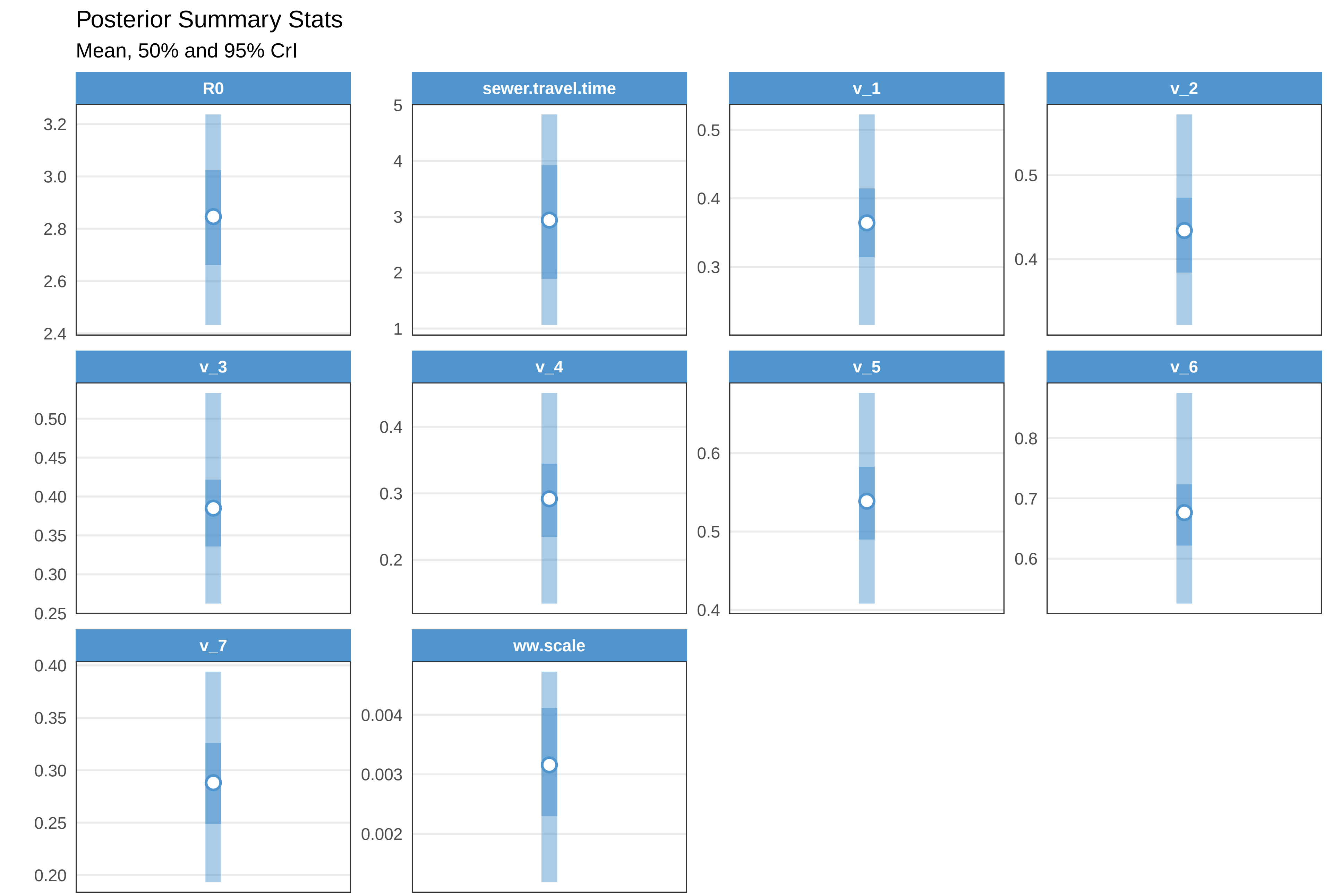

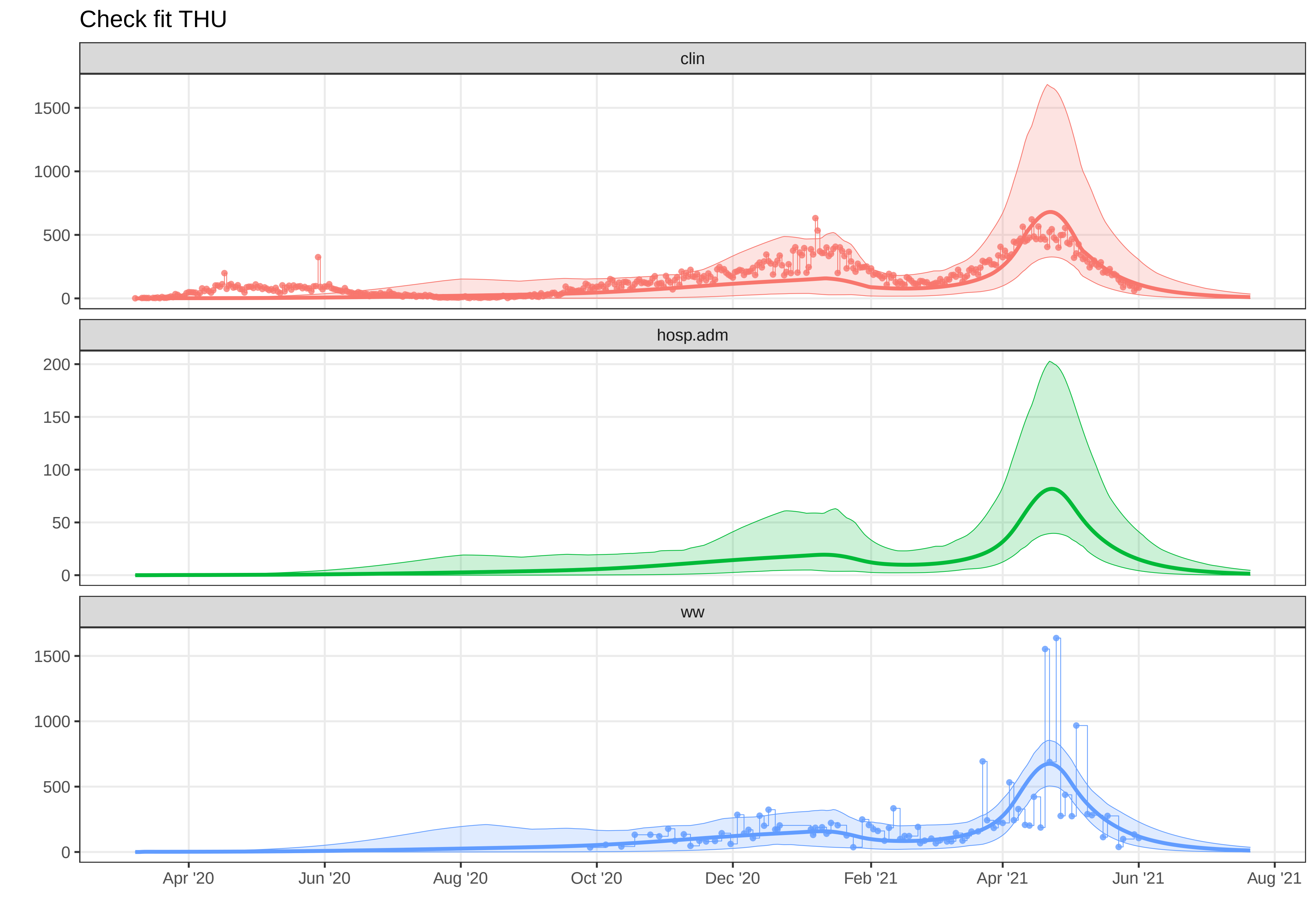

### plot-fit-TNT-07-08-ww.pdf

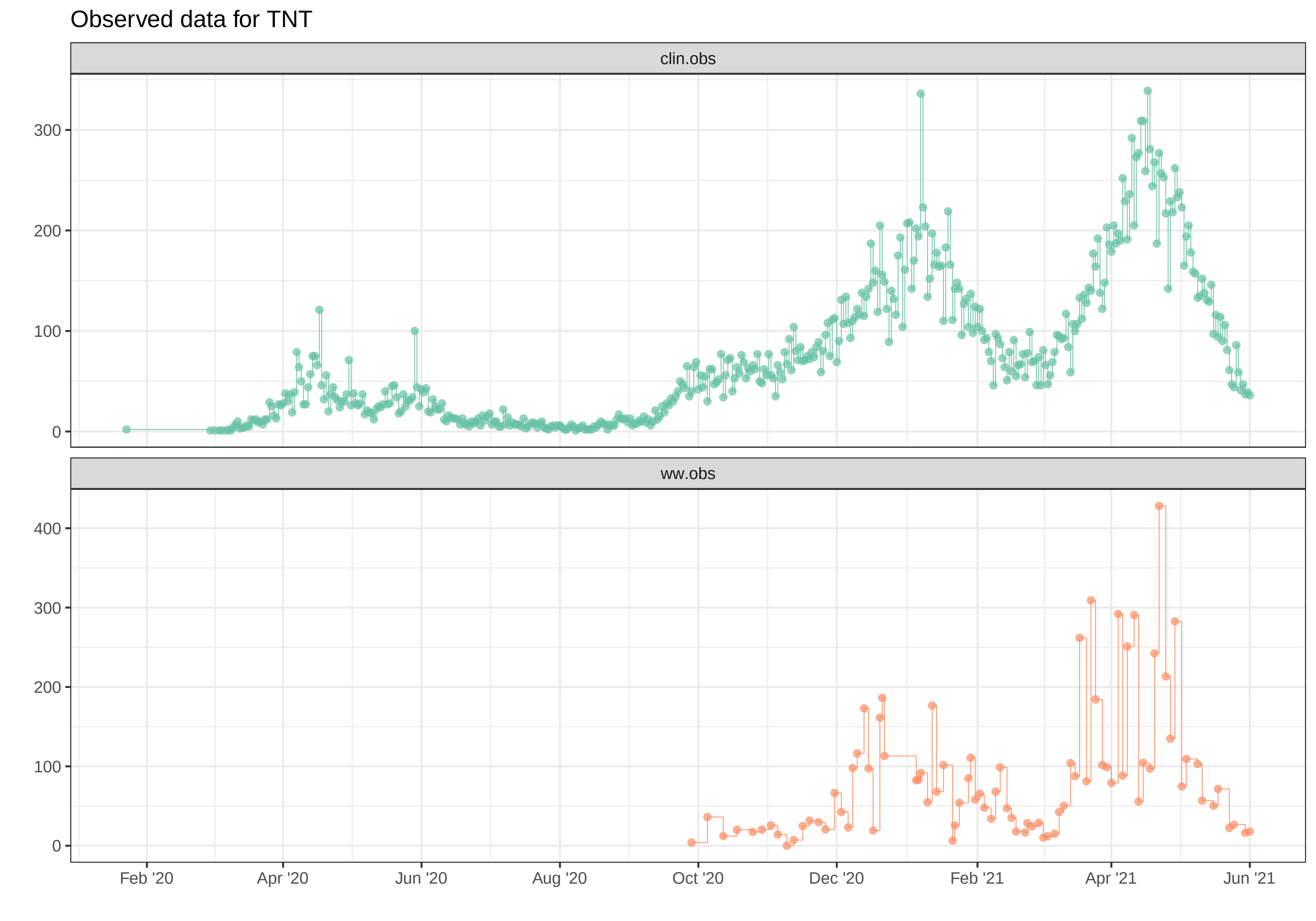

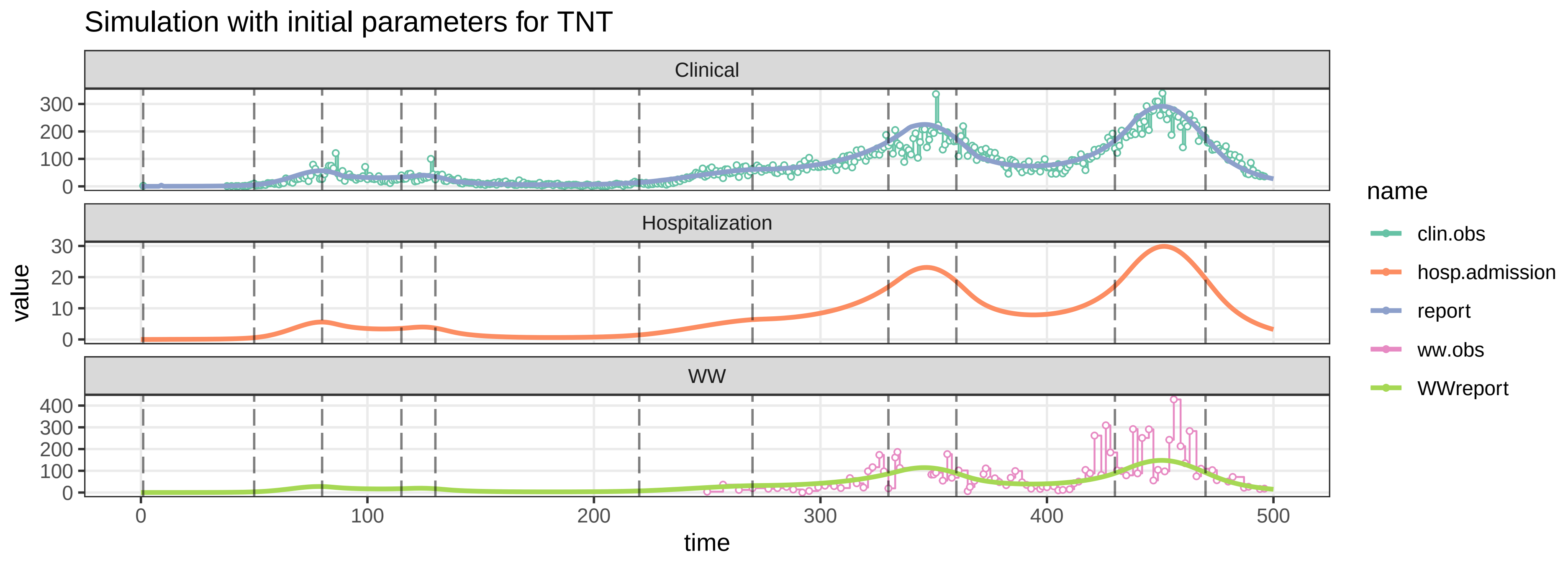

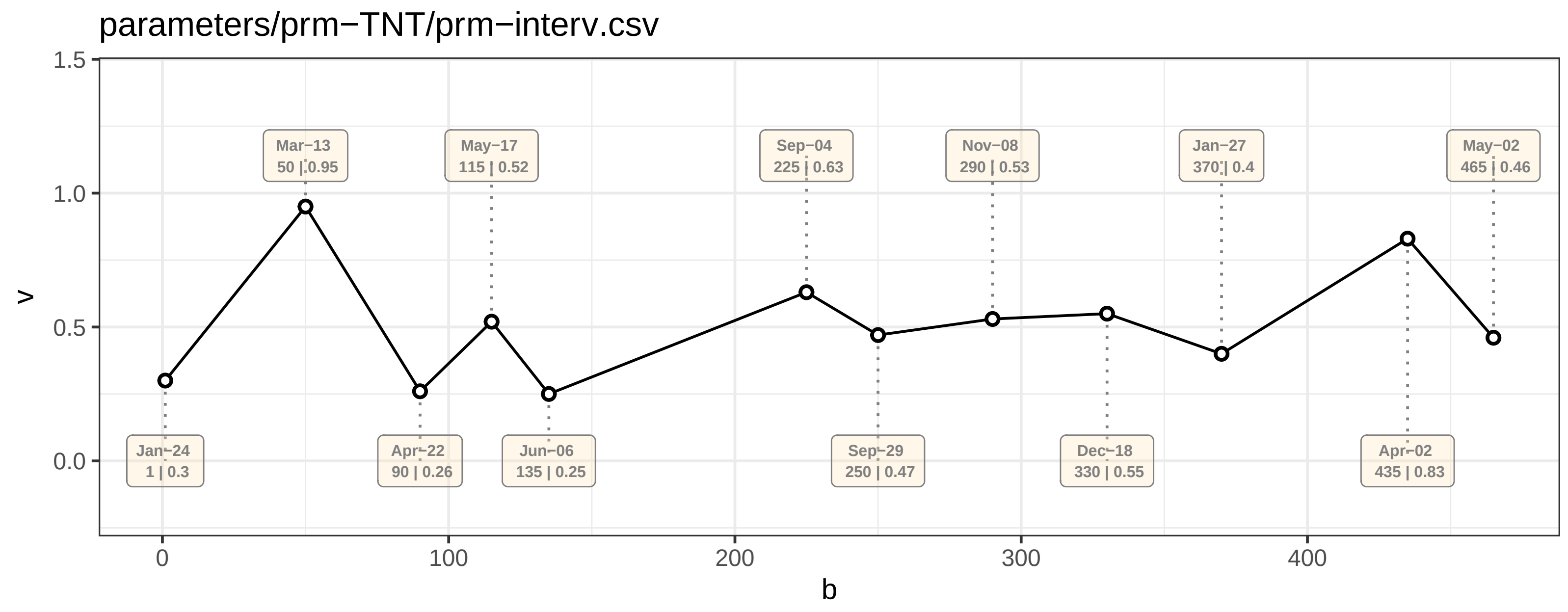

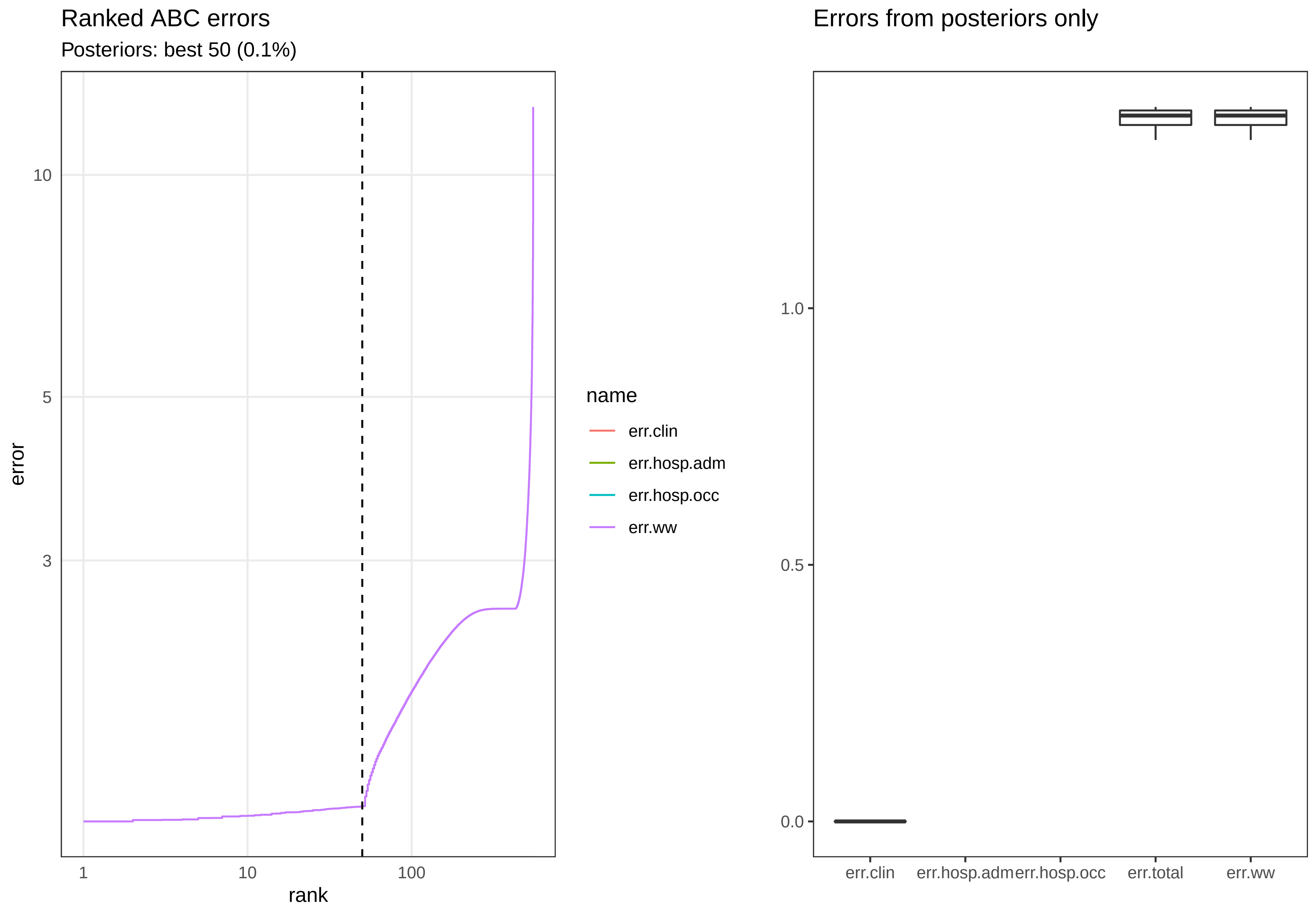

#### Posterior distributions

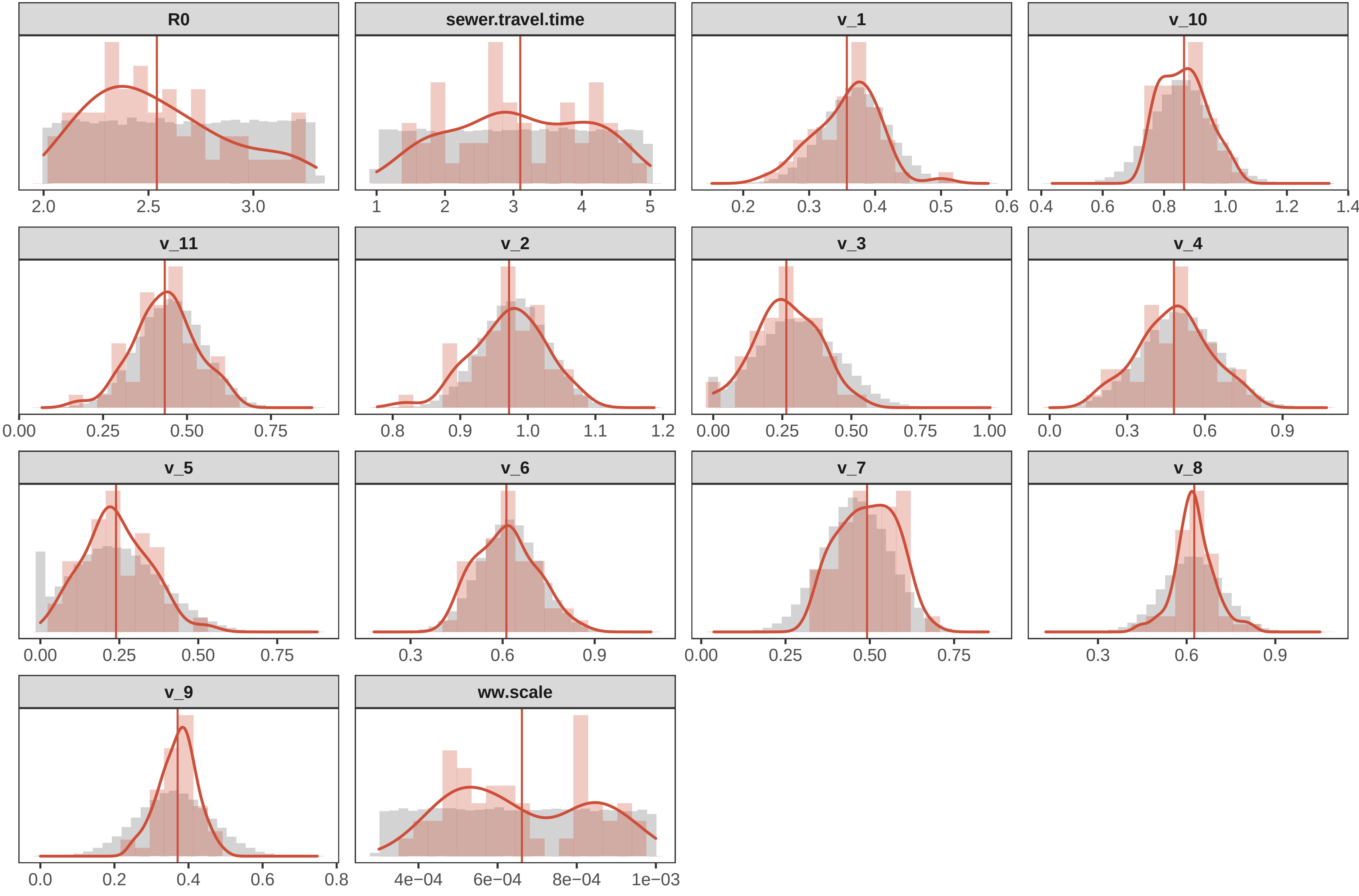

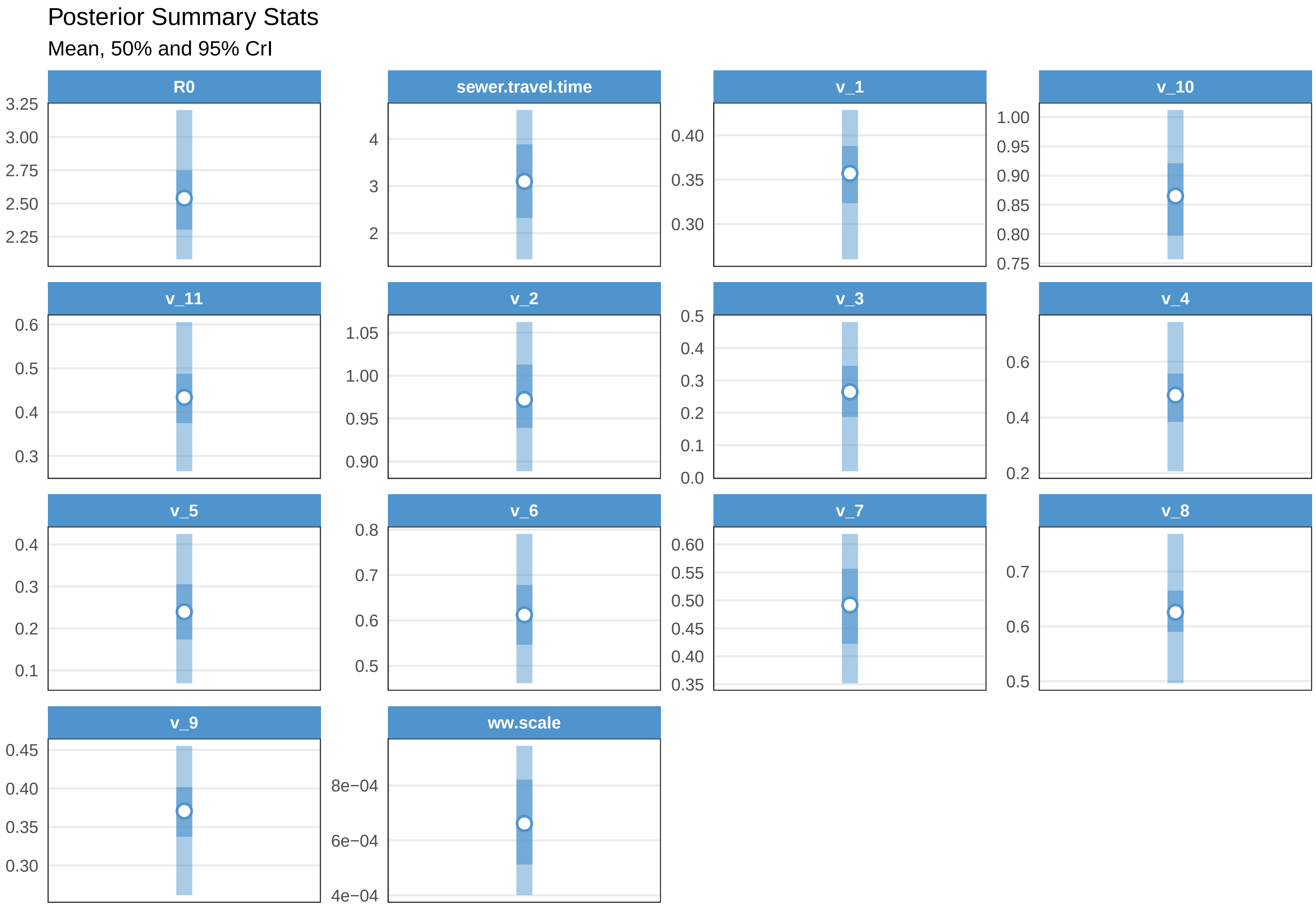

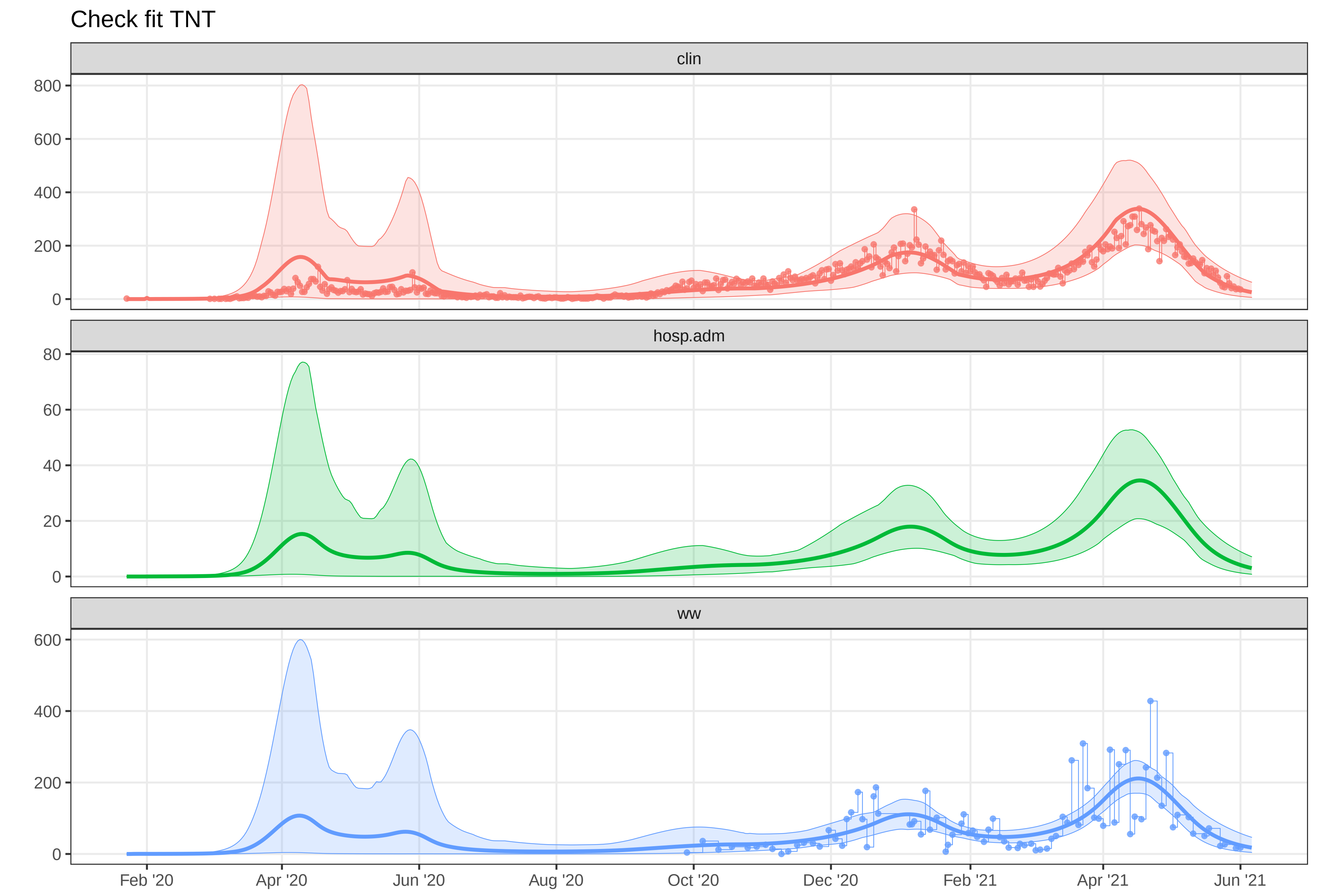
